# Selection bias due to omitting interactions from inverse probability weighting

**DOI:** 10.1101/2025.05.15.25327313

**Authors:** Liping Wen, Kate Tilling, Rosie Cornish, Rachael Hughes, Apostolos Gkatzionis

## Abstract

Inverse probability weighting (IPW) is often used to adjust for selection bias, typically using a simple logit model without interactions as a missingness model. However, the size of the selection bias depends on the interaction between exposure and outcome in their effect on selection - implying that it may be important to include interactions in the IPW model. Via a simulation and a real-data application we compare the performance of IPW with and without interaction terms to estimate a regression coefficient. The simulation study shows that IPW including interactions gives less biased estimates than IPW without interactions in all scenarios studied. Importantly, IPW using a logit model with no interactions often gives estimates close to the complete case analysis (CCA) - perhaps giving false reassurance that results are robust to selection bias. The real-data application investigates the association between unemployment and sleep duration, using data from Understanding Society. IPW including interactions suggests that unemployment is associated with a reduction in sleep duration of around 23 (9, 38) minutes, compared to 27 (14, 40) minutes for IPW without interactions, and 31 (19, 43) minutes for CCA. We strongly recommend including interactions in missingness models to adjust for selection bias.

## 1 Introduction

Epidemiologic studies often aim to estimate the causal effect of an exposure on an outcome, adjusting for confounders. However, many studies suffer from some form of non-random selection, due to participation, dropout from the study, or intermittent non-engagament with the study. If analysing only the selected participants (i.e., those with complete data on all the analysis model variables, also termed a complete case analysis, CCA), the estimate of the causal effect of the exposure on the outcome might be biased (known as selection bias).

There are two popular methods for analysing incomplete data to minimise selection bias: multiple imputation (MI) and inverse probability weighting (IPW)^1–3^. To perform IPW, a model for the selection mechanism (the missingness model) is used to predict the probability of an individual having complete data for all variables in the analysis model. The analysis model is then fitted only in complete cases, with each selected individual weighted by the inverse of their selection probability. If the missingness model is correctly specified, then IPW can adjust for selection bias.^4^ To perform MI, multiple complete datasets are created by imputing (“filling in”) the missing values of the incomplete observations, using an imputation model for each incomplete variable. The estimand of interest is estimated within each completed data set and then averaged across all imputed datasets, with standard errors calculated using Rubin’s rules.^1^ Both MI and IPW are valid when data are missing at random (MAR), i.e. the distribution of the incomplete variables does not differ between those with and without complete data, conditioning on all complete variables.^5^ MI typically attains higher precision than IPW when all models are correctly specified. However, correctly specifying a weighting model for selection may be easier than correctly specifying an imputation model in some situations, such as when multiple variables are jointly missing,^6^ for example when individuals are missing an entire wave of follow-up in a longitudinal study. This manuscript focuses on using IPW to adjust for selection bias.

The interaction between exposure and outcome in their effects on selection is informative about the magnitude of selection bias.^7–10^ For example, recent work has shown that for linear, logit and Poisson regression, the magnitude of the bias in the exposure-outcome regression coefficient is proportional to the interaction between the exposure and outcome in their effect on selection, on the log-linear scale.^10^ However, in practice, many applied researchers use a simple logit/probit model without interactions to estimate selection probabilities,^3, 11–14^ although some analyses do include interaction terms.^15,16^ Therefore, through a simulation study and a real-data application, we aim to illustrate whether including interactions in the missingness model improves the performance of IPW in reducing selection bias. In our simulation study, the participants have outcome data MAR. We then implement IPW using logit and log-additive weighting models with and without interactions and compare their performance. To further illustrate our findings, we conduct a real-data application using data from Understanding Society (US), a UK household longitudinal study. For this data analysis, we investigate the total effect of unemployment status at wave 9 on sleep duration at wave 10, adjusting for confounding and selection bias.

## 2. Simulation study

### 2.2 Notation

Throughout this manuscript, we let *X* represent the exposure and *Y* the outcome of interest. Our objective is to investigate the effect of *X* on *Y* . We simulate data according to three directed acyclic graphs (DAGs), presented in Figure 1. In all three DAGs, exposure *X* is binary: participants are exposed if *X* = 1 and non-exposed if *X* = 0. The outcome *Y* may be continuous or binary, and *D* and *C* are continuous covariates. The binary variable *S* indicates whether a participant is selected (*S* = 1 denotes a participant is selected for analysis and *S* = 0 otherwise). We assume that the exposure and covariate values are observed for all participants, but the outcome is only observed among the selected participants; therefore, *S* = 1 implies *Y* is observed, and *S* = 0 implies *Y* is missing. *PS* denotes the average probability of selecting participants, i.e., proportions of participants selected.

**Figure 1.**
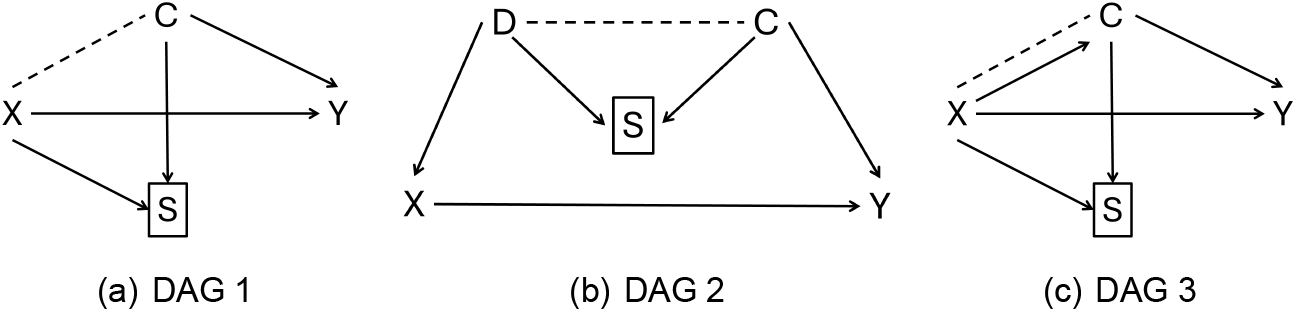
The three DAGs for simulating data. X denotes the exposure, Y denotes the outcome, C and D denote covariates, and S is the selection indicator.

### 2.2 Aim and estimand

For simplicity, we have simulated scenarios with no confounding. Under each data generating mechanism (DGM), the analysis of interest is regressing *Y* on *X* in the full sample and the estimand of interest is the coefficient of *X* in this model. Six methods, including full-data analysis, CCA and four IPW methods, (detailed in Section 2.4), are fitted to the simulated data. Note that applying the analysis model to the full sample would give unbiased results (i.e. there are no other sources of bias such as unmeasured confounding). The four IPW methods differ with respect to specification of the missingness model (i.e., using logit or log-additive models with and without a term for the interaction between the variables causing selection). This study aims to compare the performance of IPW models with and without interactions.

### 2.3 Data generating mechanism

The data are generated according to the three DAGs shown in Figure 1. Taking DAG 1 as an example, the selection indicator *S* is a common effect of variables *X* and *C*. Selection bias may arise because conditioning on this selection indicator induces an association between *X* and *C* shown by the dashed line.

Our simulation study explores 14 main DGMs in total. The DGMs vary by DAG, the type of *Y* (binary or continuous) and the model for *S* in the DGM (logit, log-additive or probit model). Under DAG 1, there are six DGMs: *Y* is continuous or binary, and for each of these, the missingness model simulating *S* is a logit, log-additive or probit regression model. For DAGs 2 and 3 we do not consider a probit model for simulating *S*; this results in four main DGMs for each of these two DAGs.

For DAG 1, the DGMs for *X, C* and *Y* are given as follows.

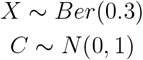

For a continuous outcome,

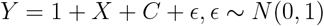

For a binary outcome, we simulate outcome probabilities as

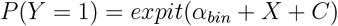

Then

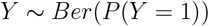

choosing the intercept *α*_*bin*_ so that the prevalence of the outcome is 50%.

For the DGM using a logit model (the DGM using a log-additive or probit model is provided in **Supplementary** 1-1) for selection, the values of the selection indicator *S* are simulated as

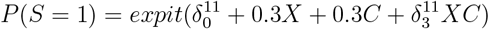

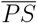 is set by specifying the intercept 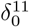 accordingly.

We then consider two simulation scenarios for each of the 14 main DGMs for DAG

1. For DAG 1, in the first simulation scenario, we fix the average selection probability to be 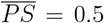 and vary the interaction parameter 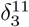, allowing it to take values *±*0.3, *±*0.2, *±*0.1, and 0. In the second simulation scenario, to examine the effect of changing the selection probability, we set 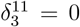 (simulating selection without an interaction term) or 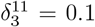 (simulating selection with an interaction term) and vary 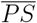, using the values 0.1, 0.3, 0.5, 0.7, 0.9. For the other two DAGs, we only consider the first simulation scenario (i.e., fixing 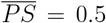 and varying the interaction parameter). Thus for DAG1 we have 28 scenarios in total (14 main DGMs, each with two simulation scenarios), and for DAGS 2 and 3 we have 8 in total (4 main DGM for each DAG, each with one simulation that varies the interaction parameters). For each simulation scenario, we generate 1000 simulated datasets, each with 100, 000 participants for the full sample. Our simulations are implemented in R version 4.3.3.^17^

More details on how to simulate our other scenarios, including using a log-additive or probit model for *S* and simulating data for DAGs 2 and 3, are provided in Supplementary 1-1.

### 2.4 Analysis methods

The analysis of interest is a regression of *Y* on *X*, and our target estimand is the regression coefficient for *X*. No adjustment for other covariates is required because there are no confounders of the exposure and outcome (DAGs 1-3, Fig 1). We estimate the exposure effect using six different methods. Method 1 is the full sample method, where the analysis model is fitted to both the selected (*S* = 1) and unselected individuals (*S* = 0). All other methods are CCA analyses, fitting the analysis model to the selected (*S* = 1) individuals only. Method 2 is an unweighted CCA. Methods 3 to 6 are IPW methods which differ only with respect to the missingness model for S. The missingness models are either a logit or log-additive model, and either include or exclude the interaction between the variables causing *S*. We refer to these four IPW models as logit model with interaction, logit model without interaction, log-additive model with interaction, and log-additive model without interaction. Method 1 should give unbiased results because it fits a correctly specified outcome model to the full sample. When performing IPW, we assume the weights are known and compute model-based standard errors using the robust standard error estimation methods from the “survey” package.^18^

### 2.5 Performance measures

The key performance measure that we consider is bias in the estimated coefficient for the effect of *X* on *Y* . Coverage, empirical standard error (EmpSE), average model-based standard error (Ave.ModSE) and mean squared error (MSE) are also reported in Supplementary 1-1 and 1-2. The performance measures are calculated based on 1000 repetitions, using the “rsimsum” package.^19^ See^20^ for details on how these measures are defined and computed, including how to compute their Monte Carlo Standard Error (MCSE).

## 3 Results

As expected the full data estimates were unbiased for all scenarios. Figure 2 shows the CCA results corresponding to data simulated under DAG 1 with a logit model for *S* and a continuous outcome *Y* . We plot the bias of the exposure-outcome effect estimates against the interaction parameter 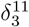, for three of the five CCA methods compared in our simulation study (IPW methods using log-additive missingness models behave similarly to the logit models and are shown in the Supplementary 1-1). Across all 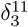 values, IPW using a logit missingness model with an interaction term has the least bias. Across all scenarios, the performance of the IPW logit model without an interaction term is very similar to that of the unweighted CCA. The log-additive model with interaction shows little bias, despite being misspecified. When 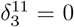, i.e., the true missingness model does not have an interaction term, there are no differences between the performances of the IPW logit model with/without interaction and the IPW log-additive model with interaction. Results from all other DGMs and simulations that vary the average selection probability 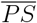 also show that including the interaction term in the missingness model is less biased than excluding this term. Detailed results are reported in Supplementary 1-1 and 1-2, including the results of the other performance measures of the methods when applied to data simulated under the DGMs for DAGs 2 and 3.

**Figure 2.**
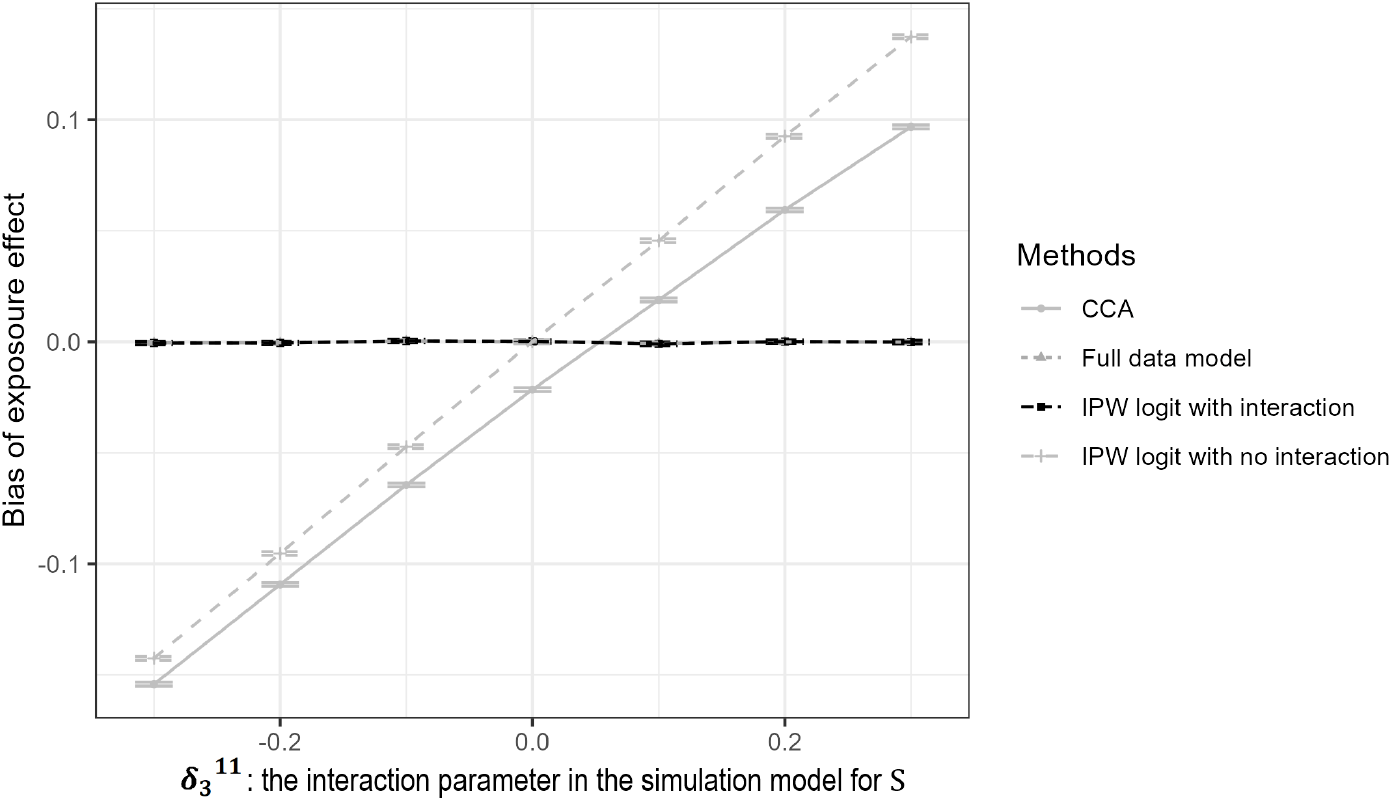
Bias in the estimation of the *X*-*Y* coefficient, across three methods for complete case analysis. Horizontal bars denote the Monte Carlo interval of the bias. The DGM is DAG 1, with continuous *Y*, logit missingness model, and proportion selected 0.5.

## 4 Real-data application

### 4.1 Data and methods

#### 4.1.1 Study description and the aim

For our real-data application, we used data from Understanding Society (US), a UK household longitudinal study. The study started in 2009 (wave 1) and every individual in a responding household is interviewed at approximately 12-month intervals. The study has a complex design with different sub-samples, detailed previously.^21^ The study provides sample design variables (for clustering and stratification) and weights that take account of unequal selection probabilities for households and differential nonresponse error.^22^

We aim to investigate the effect of unemployment status at wave 9 on sleep duration at wave 10. A DAG characterizing the assumed relationship between these variables is shown in Fig 3. The selection indicator S represents complete cases, i.e. individual participants with complete data on unemployment status, sleep duration and confounders. Apart from confounders, the structure of this DAG is the same as DAG 3 of our simulation study.

**Figure 3.**
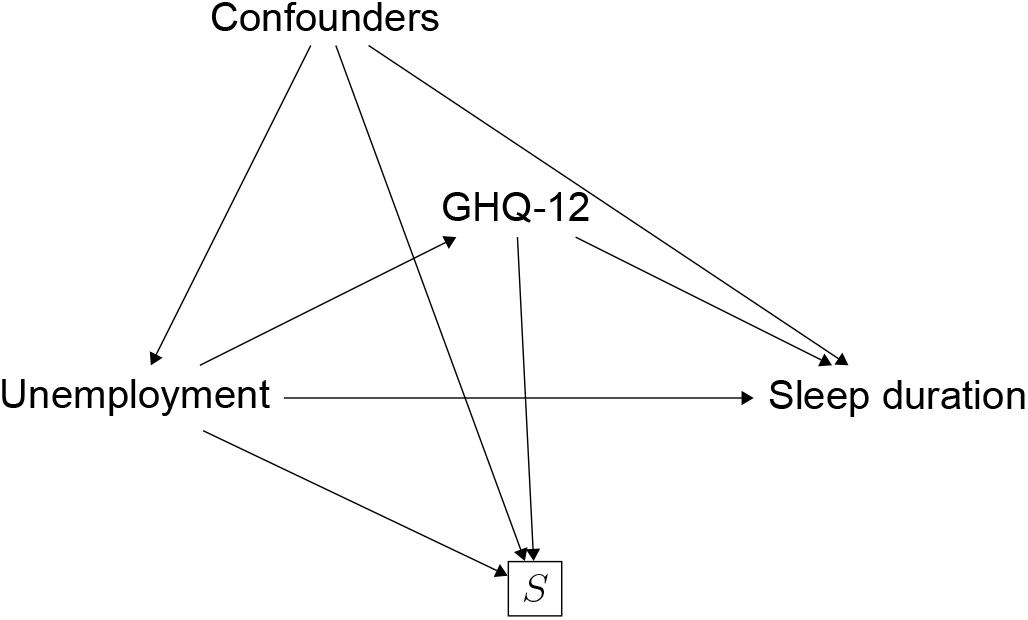
A DAG describing our assumptions about the relationships between the variables of our real-data application.

#### 4.1.2 Description of the variables

##### Outcome: sleep duration at wave 10

Sleep duration is measured in minutes of sleep that participants usually had per night during the last month.

##### Exposure: unemployment status at wave 9

Unemployment status is a binary variable: an unemployment value of 1 represents being unemployed including unemployed/long-term sick/disabled, and a value of 0 represents being self-employed/paid employment/retired/on maternity leave/family care or home/full-time student/Govt training scheme/unpaid, family business/on apprenticeship/on furlough/temporarily laid off/short term working/on shared parental leave/doing something else.

##### Confounders

We adjust for the following potential confounders: age (grouped as 24-29, 30-39, 40-49, 50-59), sex, ethnic group (broadly categorised as White British, other White, Asian, Black, Mixed/other), educational level (grouped as: Degree/higher, A level/equivalent, GCSE/equivalent, none), marital status (dichotomised as married/civil partnership/living as couple vs all other categories) and a binary variable indicating whether anyone in the household (at recruitment) was born in the UK. Missing data for educational level and marital status in wave 9 are replaced with data from wave 8, if available.

##### Mediator: GHQ-12 at wave 9 or 8

GHQ-12 is a measure that converts valid answers to 12 questions of the General Health Questionnaire (GHQ) to a single scale by recoding so that the scale for individual variables runs from 0 to 3, and then summing over the 12 variables to give a scale running from 0 (the least distressed) to 36 (the most distressed).

##### Proxy measures

The key factor affecting bias in the estimation of the exposure-outcome coefficient is the interaction between them, and mediators/confounders, in the selection model. However, for non-complete cases, we may not have observed data on exposure and/or outcome, or the confounders or mediators. Thus we cannot use the true exposure, outcome, confounders and mediators in the missingness model. We therefore use complete proxy measures, i.e. complete variables related to the incomplete analysis model variables, to model the missingness mechanism, and in particular to examine likely interactions. Proxy exposure/confounders/mediator is the unemployment status/confounders/GHQ-12 measured in wave 9, and any missing values are replaced with values from the most recent previous wave with available data.

#### 4.1.3 Inclusion criteria

The study is restricted to 49,952 individuals aged 25 to 59 at wave 10, recruited in the General Population Sample (GPS), the Ethnic Minority Boost sample (EMB) or the Immigrant and Ethnic Minority Boost sample (IEMB). We define the “first adult wave” to be the first wave at which they were (or could have been) included as an adult. Within this sample, 32,118 (64%) participants are left after excluding the following participants: 7,274 (15%) participants without a household participation weight, 9,649 (19%) participants who did not respond in the first adult wave, 911 (2%) participants without an individual participation weight and 1 (0%) participant with outcome sleep duration being zero. The household and individual participation weights are described below. Of our full-data sample with 32,117 individuals, there are 9,654 (30%) complete cases, i.e. those with complete data on the exposure, outcome and all confounders. There are three potential sources of selection: whether a household is selected for the US study, whether an individual from a selected household participates, and whether the individual has provided complete data for all variables in our analysis. To implement IPW, we used three sets of weights, one for each selection process.

##### Household participation weight

In Understanding Society, ethnic minorities are over-sampled as part of the EMB and IEMB samples. The US study provides household participation weights at the wave of recruitment, i.e., wave 1 or 6, to adjust for unequal sampling fraction and differential household response. The “Enumeration cross-sectional analysis weight” for household participation in GPS, EMB and IEMB is used here. Details of this weight can be found in the User guide.^22^

##### Individual participation weight

Among the 42,678 individuals aged 24 to 59 with a household participation weight and whose household responded at recruitment, 33,029 (77%) individuals responded (i.e., completed a full interview) at the first adult wave. Among the 9,649 (23%) individuals who did not respond at the first adult wave, the majority (6,169, 64%) did not respond at any subsequent wave (up to wave 10). Thus, we derived an individual participation weight, given by the inverse of the probability that an individual responds at the first adult wave. This response probability is modelled by logit regression, fitted using the least absolute selection and shrinkage operator (lasso) including all possible two-way interactions between covariates in this model. Lasso shrinks some coefficients to zero and retains the subset of covariates with the strongest effects, resulting in a model with minimal over-fitting.^23^ Initial covariates for in this model are the individuals’ age, sex, ethnicity, seven household characteristics (net income quintiles, whether or not there was at least one person in the household in paid employment, the highest household qualification, the number of bedrooms, housing tenure, household size, and whether anyone in the household was born in the UK), and three additional characteristics (government region of their residence, sample region, and the wave at which an individual was first eligible for the adult interview). More details about these covariates are given in Supplementary 2.

#### 4.1.4 Complete-case weight

Complete cases are individuals with all variables in the analysis model observed. The complete-case weight is the inverse probability of being a complete case, modelled using the lasso logit model with covariates including proxy exposure (unemployment status), proxy GHQ-12 and proxy confounders (age, sex, ethnic group, educational level, marital status and whether anyone in the household was born in the UK).

#### Statistical analysis

The analysis of interest is a weighted linear regression to estimate the effect of unemployment status at wave 9 on sleep duration at wave 10, adjusting for confounders. The weights account for the fact that the Understanding Society preferentially sampled minority households and the differential individual response at the first adult wave. Therefore, participants are weighted by the product of their household participation weight and individual participation weight.

We estimate the exposure effect using five weighted complete-case analyses. The first model (CCAw) here is a weighted analysis using only the product of household and individual participation weights. The four other models are IPW analyses using the product of these weights and a complete-case weight derived using a logit missingness model. We consider four missingness models that differ with respect to the possible interactions included in the model: (i) no interaction terms; (ii) interaction term between the proxy exposure and mediator (GHQ-12 score); (iii) interaction terms between proxy exposure and mediator, proxy exposure and confounders, as well as proxy mediator and confounders; (iv) all possible pairwise interactions between the proxy exposure, mediator, and confounders.

### 4.2 Results

Descriptive statistics for all variables are reported in Supplementary 2. Comparing the 9,654 complete cases with the full sample of 32,117 individuals, most characteristics are similar, except that the household participation weight is higher among the complete cases, 0.879 vs 0.797.

Table 1 shows the results of the first complete-case analysis(CCAw) (fitted to 9,654 participants) and the IPW analyses (fitted to 9,647 participants). The CCAw indicates that unemployment status reduces sleep duration by 31 (95% confidence interval (CI) 19, 43) minutes, while the lasso IPW logit model with all interaction terms yielded an estimate of 23 (95% CI 9, 38) minutes. The more interaction terms are added, the more the IPW estimates differ from those obtained using CCAw. Comparing lasso IPW logit model with all interactions and CCAw, the absolute value of the coefficient reduces by (31-23)/31 = 26%; comparing lasso IPW logit model with no interactions and CCAw, it reduces by 15.0%.

**Table 1.**
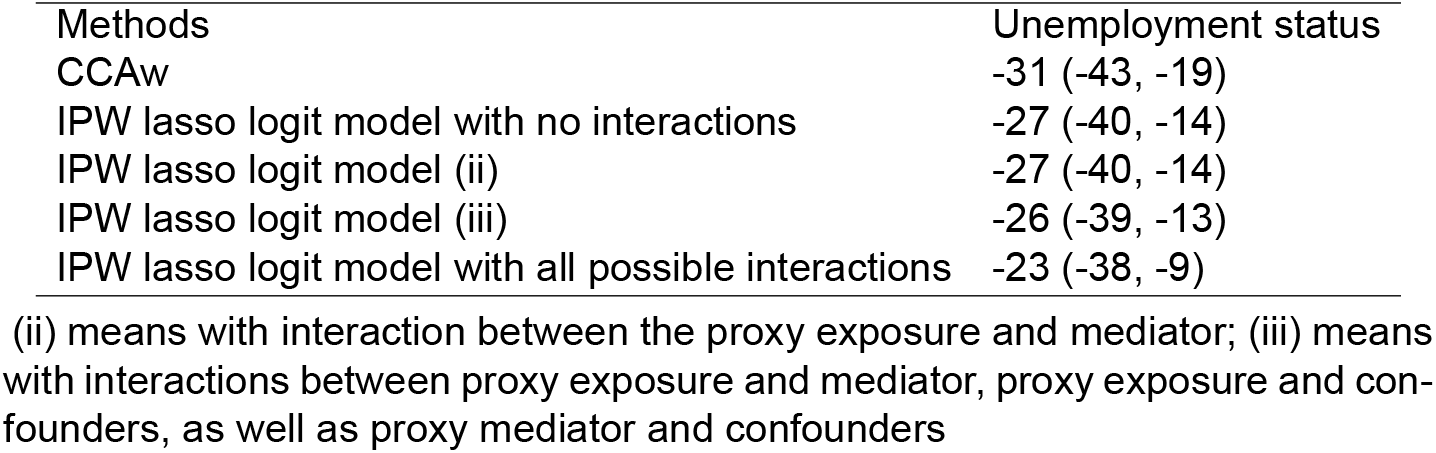
Real-data application result (coefficient and 95% confidence interval for unemployment status).

## 5 Discussion

In this paper, we show the importance of including interactions in the missingness model for IPW, through conducting a simulation study and a real-data application. In applied analyses, IPW is often used with a logit or probit missingness model without interactions. Importantly, our results show that the missingness model without interactions may not reduce bias, and can create false confidence that CCA is robust to selection bias as the results can be very close to those of CCA. Some instances of this leading to false conclusions already exist in the literature. For example, a recent paper^24^ compared the performance of IPW and CCA. The two methods had very similar performance in simulations, leading the authors to conclude that there is little benefit in using IPW over CCA. Our results suggest that this could have been due to the simulation design used by that paper, which did not consider interaction terms.

The missingness models we used in this manuscript (logit, probit and log-additive models with or without interactions) were all relatively simple. Restricting to these simple models was enough to illustrate our point about the limitations of the missingness model without interactions. In some applications, the selection mechanism may be more complex and even the inclusion of interaction terms in a simple logit or log-additive model may not correctly specify the missingness model. Several methods have been developed in the statistics and machine learning literature to construct IPW weights; these include, for example, boosted classification and regression trees (CARTs), and random forest.^25–28^ Most of these methods can deal implicitly with interactions and non-linearities in their implementations.^29^ However, implementing these methods may not be as straightforward for applied researchers as running a standard logit regression. In contrast, the inclusion of interaction terms, as suggested in this manuscript, is a very simple extension of the standard logit model that can substantially improve the performance of IPW, especially as interaction terms are known to be informative about selection bias.^7–10^ The goodness-of-fit of the missingness model should also be carefully examined before proceeding with IPW.

In our real-data application we were able to include a proxy exposure in the missingness model, making use of the exposure measured from previous waves. In practice, previous measures of the exposure and outcome may not be available, and the exposure and outcome may only be observed in the selected sample; in such cases we cannot directly model selection and have to choose alternative proxies to model this. In our application we assume MAR and do not consider a sensitivity analysis assuming data missing not at random (MNAR), i.e., the probability of being a complete case depends on the unobserved data given the observed values.^30^ If the selection mechanism is actually MNAR, then the IPW analysis would be biased, although a careful design of the weighting model might yield a less biased estimate than CCA. Alternatively, a bias analysis can be conducted using IPW incorporating the MNAR missingness mechanism (e.g.,^31^).

There are some limitations in our study. Firstly, we consider three relatively simple DAGs including only a small number of variables, with the outcome simulated using only linear or logit models. Secondly, in our simulations we only consider two variables causing selection into the study sample. A greater number of variables causing selection would require more decisions around which interactions to include in the missingness model; the lasso method used in our real-data analysis can be useful for this. Thirdly, we did not consider non-linearities in the missingness model nor examine the impact of interactions or non-linearities in the analysis model.

IPW is often used to minimise bias due to missing data, particularly if there is little information to carry out imputation. We have shown that it is crucial to carefully specify the missingness model, in particular to examine the inclusion of interactions between the variables relating to selection. Not including interactions can create a false belief that there is no selection bias, due to the similarity between complete case and weighted results. We therefore suggest that researchers who are using IPW should routinely incorporate interaction terms in their logit models for selection. We have also shown the importance of considering complete proxy variables for incomplete exposure, outcome and other analysis model variables. These proxy variables help to assess the likelihood of the exposure and outcome causing selection into the model.

## Data Availability

The R code used for the simulation study and the Stata code used for the real-data application of our paper are available at the GitHub repository https://github.com/Wen-wow/IPW_interaction. Access to Understanding Society (US) data for our real-data application was obtained under Study Number (SN) 6614; the data are available upon request to the US study on the website: https://beta.ukdataservice.ac.uk/datacatalogue/studies/study?id=6614.

https://beta.ukdataservice.ac.uk/datacatalogue/studies/study?id=6614

## Data, coding and supplementary materials

The dataset for the real-data application is Understanding Society: Waves 1-13, 2009-2022 and Harmonised BHPS: Waves 1-18, 1991-2009 SN 6614. The R coding for the simulation study, the Stata coding for the real-data application, and supplementary materials are available at [GitHub](https://github.com/Wen-wow/IPW_interaction/tree/main).

## References

1. Little RJ, Carpenter JR, Lee KJ. A comparison of three popular methods for handling missing data: complete-case analysis, inverse probability weighting, and multiple imputation. Sociological Methods & Research. 2022:00491241221113873.

2. Perkins NJ, Cole SR, Harel O, et al. Principled approaches to missing data in epidemiologic studies. American journal of epidemiology. 2018;187(3):568– 575.

3. Seaman SR, White IR. Review of inverse probability weighting for dealing with missing data. Statistical methods in medical research. 2013;22(3):278–295.

4. Hernan M, Robins J. Causal inference: What if. boca raton: Chapman & hill/crc. 2020.

5. Carpenter JR, Smuk M. Missing data: A statistical framework for practice. Biometrical Journal. 2021;63(5):915–947.

6. Seaman SR, White IR, Copas AJ, Li L. Combining multiple imputation and inverse-probability weighting. Biometrics. 2012;68(1):129–137.

7. Greenland S. Response and follow-up bias in cohort studies. American journal of epidemiology. 1977;106(3):184–187.

8. Greenland S. Basic methods for sensitivity analysis of biases. International journal of epidemiology. 1996;25(6):1107–1116.

9. Jiang Z, Ding P. The directions of selection bias. Statistics & Probability Letters. 2017;125:104–109.

10. Gkatzionis A, Seaman SR, Hughes RA, Tilling K. Relationship between collider bias and interactions on the log-additive scale. arXiv preprint 2308.00568. 2023.

11. Sun B, Perkins NJ, Cole SR, et al. Inverse-probability-weighted estimation for monotone and nonmonotone missing data. American journal of epidemiology. 2018;187(3):585–591.

12. Gyamfi-Bannerman C, Clifton RG, Tita AT, et al. Neurodevelopmental Outcomes After Late Preterm Antenatal Corticosteroids: The ALPS Follow-Up Study. JAMA. 2024.

13. Vejrup K, Magnus P, Magnus M. Lost to follow-up in the Norwegian mother, father and child cohort study. Paediatric and Perinatal Epidemiology. 2022;36(2):300–309.

14. Alonso A, Seguí-Gómez M, De Irala J, Sanchez-Villegas A, Beunza JJ, Martínez-Gonzalez MÁ. Predictors of follow-up and assessment of selection bias from dropouts using inverse probability weighting in a cohort of university graduates. European journal of epidemiology. 2006;21:351–358.

15. Härkänen T, Kaikkonen R, Virtala E, Koskinen S. Inverse probability weighting and doubly robust methods in correcting the effects of non-response in the reimbursed medication and self-reported turnout estimates in the ATH survey. BMC Public Health. 2014;14:1–10.

16. Alten S, Domingue BW, Faul J, Galama T, Marees AT. Reweighting UK Biobank corrects for pervasive selection bias due to volunteering. International Journal of Epidemiology. 2024;53(3):dyae054.

17. R Core Team . R: A Language and Environment for Statistical Computing. R Foundation for Statistical Computing Vienna, Austria 2024.

18. Lumley T. Analysis of complex survey samples. Journal of statistical software. 2004;9:1–19.

19. Gasparini A. rsimsum: Summarise results from Monte Carlo simulation studies. Journal of Open Source Software. 2018;3(26):739.

20. Morris TP, White IR, Crowther MJ. Using simulation studies to evaluate statistical methods. Statistics in medicine. 2019;38(11):2074–2102.

21. Buck N, McFall S. Understanding Society: design overview. Longitudinal and Life Course Studies. 2012;3(1):5–17.

22. Essex CU. Understanding Society: Waves 1-13, 2009-2022 and Harmonised BHPS: Waves 1-18, 1991-2009, User Guide. Institute for Social and Economic Research (2023). 2023:61.

23. Tibshirani R. Regression shrinkage and selection via the lasso. Journal of the Royal Statistical Society Series B: Statistical Methodology. 1996;58(1):267– 288.

24. Metten MA, Costet N, Multigner L, Viel JF, Chauvet G. Inverse probability weighting to handle attrition in cohort studies: some guidance and a call for caution. BMC Medical Research Methodology. 2022;22(1):45.

25. McCaffrey DF, Ridgeway G, Morral AR. Propensity score estimation with boosted regression for evaluating causal effects in observational studies.. Psychological methods. 2004;9(4):403.

26. Lee BK, Lessler J, Stuart EA. Improving propensity score weighting using machine learning. Statistics in medicine. 2010;29(3):337–346.

27. Hill JL. Bayesian nonparametric modeling for causal inference. Journal of Computational and Graphical Statistics. 2011;20(1):217–240.

28. Wager S, Athey S. Estimation and inference of heterogeneous treatment effects using random forests. Journal of the American Statistical Association. 2018;113(523):1228–1242.

29. Westreich D, Lessler J, Funk MJ. Propensity score estimation: neural networks, support vector machines, decision trees (CART), and meta-classifiers as alternatives to logistic regression. Journal of clinical epidemiology. 2010;63(8):826–833.

30. Little RJ, Rubin DB. Statistical analysis with missing data;793. John Wiley & Sons 2019.

31. White IR, Carpenter J, Horton NJ. A mean score method for sensitivity analysis to departures from the missing at random assumption in randomised trials. Statistica Sinica. 2018;28(4):1985.

