## Supplementary material for "Selection bias due to omitting interactions from inverse probability weighting": Supplementary 1-1 report of DAG 1 and simulation table.docx

Supplementary 1-1 report of DAG 1 and the simulation table

### 1 The simulation table

|  | | **DAG 1** | **DAG 2** | **DAG 3** |
| --- | --- | --- | --- | --- |
| **Variables Simulated** | | X  C  S  Y | 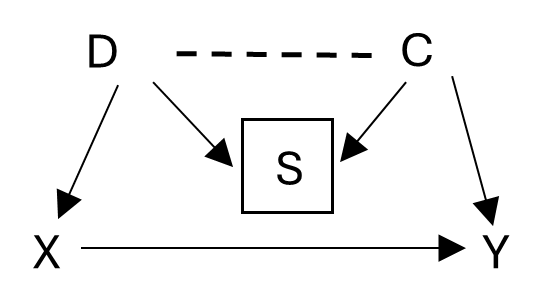 | 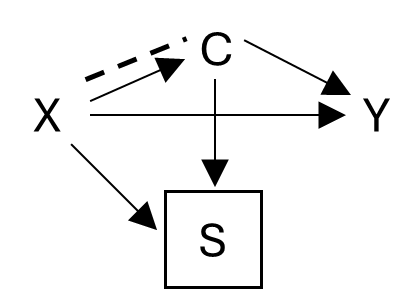 |
| Covariates (C, D) | | $C\sim N\left( 0,1 \right)$ | $C,D\sim N\left( 0,1 \right)$ | $C=1+X+\epsilon_{C}, \epsilon_{C}\sim N\left( 0,1 \right)$ |
| Exposure (X) | | $X\sim Ber\left( 0.3 \right)$ | $X\sim Ber\left( \pi_{X} \right),$  $\pi_{X}=expit(\alpha_{X}+0.3D$) | $X\sim Ber\left( 0.3 \right)$ |
| Outcome (Y) | Continuous | $Y=1+X+C+\epsilon_{Y},\epsilon_{Y}\sim N\left( 0,1 \right)$ | | |
|  | Binary | $Y\sim Ber\left( \pi_{Y} \right), \pi_{Y}=expit\left( \alpha_{bin}+X+C \right)$ | | |
| Selection (S) | Logistic | $\pi_{S}=expit\left( {\delta_{0}}^{11}+0.3X+0.3C+{\delta_{3}}^{11}XC \right)$ | $\pi_{S}=expit\left( {\delta_{0}}^{21}+0.3D+0.3C+{\delta_{3}}^{21}DC \right)$ | $\pi_{S}=expit\left( {\delta_{0}}^{31}+0.3X+0.3C+{\delta_{3}}^{31}XC \right)$ |
|  | Log-aditive | $\pi_{S}=exp\left( {\delta_{0}}^{12}-0.3X+0.1C+{\delta_{3}}^{12}XC \right)$ | $\pi_{S}=exp\left( {\delta_{0}}^{22}-0.1X+0.1C+{\delta_{3}}^{22}XC \right)$ | $\pi_{S}=exp\left( {\delta_{0}}^{32}-0.4X-0.1C+{\delta_{3}}^{32}XC \right)$ |
|  | Probit | $S=1_{S^{'}>0},$  $S^{'}\sim N\left( {\delta_{0}}^{13}+0.3X+0.3C+{\delta_{3}}^{13}XC,{1.6}^{2} \right)$ | - | - |

For binary variables, we denote $\pi_{X}=P\left( X=1 \right), \pi_{Y}=P\left( Y=1 \right), \pi_{S}=P\left( S=1 \right)$. $\alpha_{X},\alpha_{bin}$ and $\delta_{0}^{ds}\left( d=1,then s=1, 2, 3;d=2 or 3,then s=1 \right)$ are calculated by setting $\pi_{X}=0.3,\pi_{Y}=\pi_{S}=0$.

### 3 DAG 1

X

C

S

Y

#### 3.1 Strue=logit

##### 3.1.1 Y is continuous

###### Varying delta3

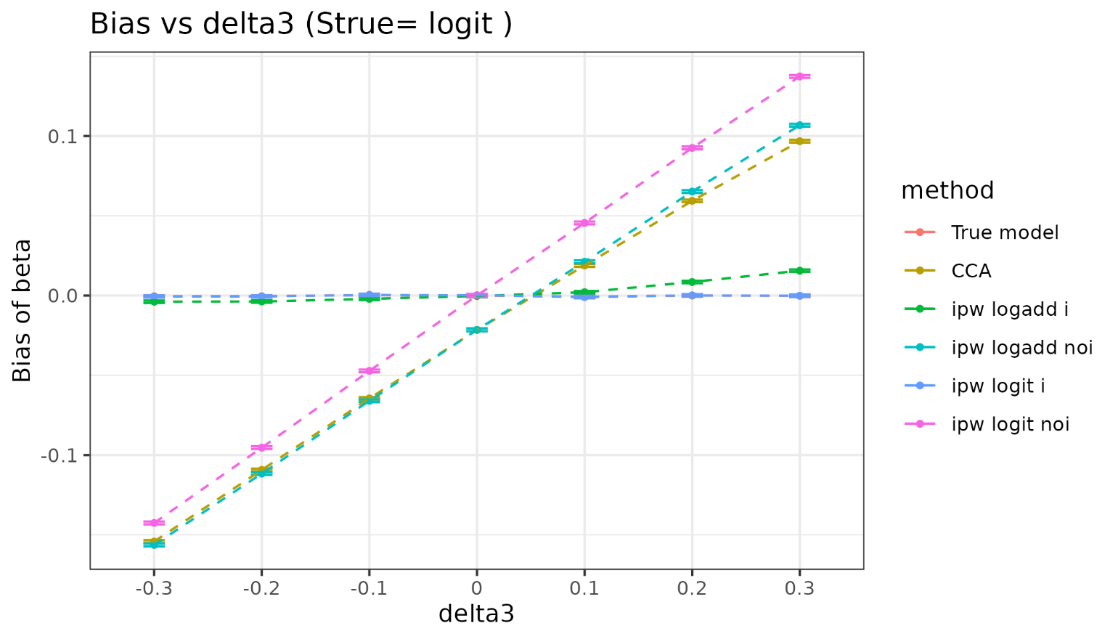

| delta3 | method | bias | coverage | EmpSE | ModSE | relative_precision | relative_error_ModSE | MSE | power |
| --- | --- | --- | --- | --- | --- | --- | --- | --- | --- |
| -0.3 | True model | -0.000457 ( 0.000307 ) | 0.949 ( 0.006957 ) | 0.009715 ( 0.000217 ) | 0.00976 ( 1e-06 ) | 0 ( 0 ) | 0.462243 ( 2.247543 ) | 9.4e-05 ( 4e-06 ) | 1 ( 0 ) |
| -0.3 | CCA | -0.154223 ( 0.000435 ) | 0 ( 0 ) | 0.013762 ( 0.000308 ) | 0.013392 ( 2e-06 ) | -50.16903 ( 2.138512 ) | -2.691679 ( 2.177001 ) | 0.023974 ( 0.000134 ) | 1 ( 0 ) |
| -0.3 | ipw logadd i | -0.003866 ( 0.000376 ) | 0.969 ( 0.005481 ) | 0.011895 ( 0.000266 ) | 0.013491 ( 2e-06 ) | -33.295466 ( 2.379658 ) | 13.423516 ( 2.537546 ) | 0.000156 ( 7e-06 ) | 1 ( 0 ) |
| -0.3 | ipw logadd noi | -0.156368 ( 0.000439 ) | 0 ( 0 ) | 0.013893 ( 0.000311 ) | 0.01352 ( 2e-06 ) | -51.105879 ( 2.123521 ) | -2.688869 ( 2.177079 ) | 0.024644 ( 0.000137 ) | 1 ( 0 ) |
| -0.3 | ipw logit i | -0.000561 ( 0.000376 ) | 0.978 ( 0.004639 ) | 0.011883 ( 0.000266 ) | 0.013549 ( 2e-06 ) | -33.165489 ( 2.389738 ) | 14.018676 ( 2.550861 ) | 0.000141 ( 6e-06 ) | 1 ( 0 ) |
| -0.3 | ipw logit noi | -0.142579 ( 0.000441 ) | 0 ( 0 ) | 0.013943 ( 0.000312 ) | 0.013587 ( 2e-06 ) | -51.456521 ( 2.112216 ) | -2.553668 ( 2.180105 ) | 0.020523 ( 0.000126 ) | 1 ( 0 ) |
| -0.2 | True model | -0.000449 ( 0.000308 ) | 0.951 ( 0.006826 ) | 0.009733 ( 0.000218 ) | 0.00976 ( 1e-06 ) | 0 ( 0 ) | 0.280413 ( 2.243476 ) | 9.5e-05 ( 4e-06 ) | 1 ( 0 ) |
| -0.2 | CCA | -0.109327 ( 0.000424 ) | 0 ( 0 ) | 0.01341 ( 3e-04 ) | 0.013389 ( 2e-06 ) | -47.326632 ( 2.273682 ) | -0.157028 ( 2.233709 ) | 0.012132 ( 9.3e-05 ) | 1 ( 0 ) |
| -0.2 | ipw logadd i | -0.003737 ( 0.000369 ) | 0.97 ( 0.005394 ) | 0.01167 ( 0.000261 ) | 0.013502 ( 2e-06 ) | -30.450544 ( 2.442652 ) | 15.689948 ( 2.588259 ) | 0.00015 ( 7e-06 ) | 1 ( 0 ) |
| -0.2 | ipw logadd noi | -0.111608 ( 0.000427 ) | 0 ( 0 ) | 0.013501 ( 0.000302 ) | 0.013549 ( 2e-06 ) | -48.029967 ( 2.282424 ) | 0.355925 ( 2.245205 ) | 0.012638 ( 9.6e-05 ) | 1 ( 0 ) |
| -0.2 | ipw logit i | -0.000531 ( 0.000369 ) | 0.979 ( 0.004534 ) | 0.011671 ( 0.000261 ) | 0.01357 ( 2e-06 ) | -30.461226 ( 2.446938 ) | 16.271656 ( 2.601275 ) | 0.000136 ( 6e-06 ) | 1 ( 0 ) |
| -0.2 | ipw logit noi | -0.095361 ( 0.000431 ) | 0 ( 0 ) | 0.013623 ( 0.000305 ) | 0.01364 ( 2e-06 ) | -48.959872 ( 2.240173 ) | 0.12491 ( 2.240039 ) | 0.009279 ( 8.3e-05 ) | 1 ( 0 ) |
| -0.1 | True model | 0.000368 ( 0.000311 ) | 0.94 ( 0.00751 ) | 0.00983 ( 0.00022 ) | 0.00976 ( 1e-06 ) | 0 ( 0 ) | -0.710937 ( 2.221297 ) | 9.7e-05 ( 4e-06 ) | 1 ( 0 ) |
| -0.1 | CCA | -0.064475 ( 0.000426 ) | 0.003 ( 0.001729 ) | 0.01346 ( 0.000301 ) | 0.013383 ( 2e-06 ) | -46.66355 ( 2.298115 ) | -0.571055 ( 2.224443 ) | 0.004338 ( 5.5e-05 ) | 1 ( 0 ) |
| -0.1 | ipw logadd i | -0.002082 ( 0.000379 ) | 0.978 ( 0.004639 ) | 0.012001 ( 0.000268 ) | 0.013526 ( 2e-06 ) | -32.902083 ( 2.40672 ) | 12.71105 ( 2.52161 ) | 0.000148 ( 6e-06 ) | 1 ( 0 ) |
| -0.1 | ipw logadd noi | -0.066008 ( 0.000431 ) | 0.002 ( 0.001413 ) | 0.013645 ( 0.000305 ) | 0.01357 ( 2e-06 ) | -48.099888 ( 2.298751 ) | -0.551665 ( 2.224896 ) | 0.004543 ( 5.7e-05 ) | 1 ( 0 ) |
| -0.1 | ipw logit i | 0.000377 ( 0.00038 ) | 0.978 ( 0.004639 ) | 0.012001 ( 0.000268 ) | 0.01363 ( 2e-06 ) | -32.908052 ( 2.410388 ) | 13.569342 ( 2.540814 ) | 0.000144 ( 6e-06 ) | 1 ( 0 ) |
| -0.1 | ipw logit noi | -0.047255 ( 0.000435 ) | 0.079 ( 0.00853 ) | 0.013753 ( 0.000308 ) | 0.013688 ( 2e-06 ) | -48.91348 ( 2.257125 ) | -0.47621 ( 2.226586 ) | 0.002422 ( 4.1e-05 ) | 1 ( 0 ) |
| 0 | True model | -5e-05 ( 0.000319 ) | 0.944 ( 0.007271 ) | 0.010073 ( 0.000225 ) | 0.009759 ( 1e-06 ) | 0 ( 0 ) | -3.115756 ( 2.167496 ) | 0.000101 ( 5e-06 ) | 1 ( 0 ) |
| 0 | CCA | -0.021547 ( 0.000447 ) | 0.628 ( 0.015285 ) | 0.014141 ( 0.000316 ) | 0.013371 ( 2e-06 ) | -49.257028 ( 2.10684 ) | -5.446826 ( 2.115365 ) | 0.000664 ( 2.1e-05 ) | 1 ( 0 ) |
| 0 | ipw logadd i | -0.000298 ( 0.000387 ) | 0.97 ( 0.005394 ) | 0.012227 ( 0.000274 ) | 0.01356 ( 2e-06 ) | -32.126096 ( 2.39966 ) | 10.903137 ( 2.481173 ) | 0.000149 ( 7e-06 ) | 1 ( 0 ) |
| 0 | ipw logadd noi | -0.02149 ( 0.00045 ) | 0.644 ( 0.015141 ) | 0.014238 ( 0.000319 ) | 0.01358 ( 2e-06 ) | -49.947373 ( 2.151118 ) | -4.626509 ( 2.133738 ) | 0.000664 ( 2.2e-05 ) | 1 ( 0 ) |
| 0 | ipw logit i | 0.000205 ( 0.000386 ) | 0.972 ( 0.005217 ) | 0.012212 ( 0.000273 ) | 0.013728 ( 2e-06 ) | -31.958138 ( 2.422582 ) | 12.413001 ( 2.514959 ) | 0.000149 ( 6e-06 ) | 1 ( 0 ) |
| 0 | ipw logit noi | 7.1e-05 ( 0.000455 ) | 0.945 ( 0.007209 ) | 0.01439 ( 0.000322 ) | 0.013728 ( 2e-06 ) | -51.000179 ( 2.090092 ) | -4.605718 ( 2.134207 ) | 0.000207 ( 9e-06 ) | 1 ( 0 ) |
| 0.1 | True model | -0.000744 ( 0.000325 ) | 0.943 ( 0.007332 ) | 0.010266 ( 0.00023 ) | 0.009759 ( 1e-06 ) | 0 ( 0 ) | -4.936156 ( 2.126772 ) | 0.000106 ( 4e-06 ) | 1 ( 0 ) |
| 0.1 | CCA | 0.018832 ( 0.00044 ) | 0.694 ( 0.014573 ) | 0.013921 ( 0.000311 ) | 0.013359 ( 2e-06 ) | -45.617216 ( 2.32188 ) | -4.039276 ( 2.146856 ) | 0.000548 ( 1.9e-05 ) | 1 ( 0 ) |
| 0.1 | ipw logadd i | 0.002099 ( 0.000398 ) | 0.969 ( 0.005481 ) | 0.012586 ( 0.000282 ) | 0.013612 ( 2e-06 ) | -33.462606 ( 2.324066 ) | 8.152639 ( 2.419646 ) | 0.000163 ( 7e-06 ) | 1 ( 0 ) |
| 0.1 | ipw logadd noi | 0.021192 ( 0.000447 ) | 0.645 ( 0.015132 ) | 0.014126 ( 0.000316 ) | 0.01359 ( 2e-06 ) | -47.183485 ( 2.288413 ) | -3.797614 ( 2.152285 ) | 0.000648 ( 2.1e-05 ) | 1 ( 0 ) |
| 0.1 | ipw logit i | -0.000943 ( 0.000399 ) | 0.963 ( 0.005969 ) | 0.012625 ( 0.000282 ) | 0.013879 ( 3e-06 ) | -33.876715 ( 2.32447 ) | 9.932902 ( 2.459494 ) | 0.00016 ( 7e-06 ) | 1 ( 0 ) |
| 0.1 | ipw logit noi | 0.045519 ( 0.000452 ) | 0.097 ( 0.009359 ) | 0.014304 ( 0.00032 ) | 0.01377 ( 2e-06 ) | -48.485481 ( 2.217718 ) | -3.728713 ( 2.153832 ) | 0.002276 ( 4.2e-05 ) | 1 ( 0 ) |
| 0.2 | True model | -3e-05 ( 0.000318 ) | 0.944 ( 0.007271 ) | 0.010041 ( 0.000225 ) | 0.00976 ( 1e-06 ) | 0 ( 0 ) | -2.79585 ( 2.174653 ) | 0.000101 ( 5e-06 ) | 1 ( 0 ) |
| 0.2 | CCA | 0.059386 ( 0.000427 ) | 0.005 ( 0.00223 ) | 0.013504 ( 0.000302 ) | 0.013343 ( 2e-06 ) | -44.711288 ( 2.336862 ) | -1.190885 ( 2.210581 ) | 0.003709 ( 5.2e-05 ) | 1 ( 0 ) |
| 0.2 | ipw logadd i | 0.008492 ( 0.00038 ) | 0.935 ( 0.007796 ) | 0.012026 ( 0.000269 ) | 0.013664 ( 2e-06 ) | -30.285663 ( 2.496603 ) | 13.622773 ( 2.542032 ) | 0.000217 ( 9e-06 ) | 1 ( 0 ) |
| 0.2 | ipw logadd noi | 0.065199 ( 0.000434 ) | 0.003 ( 0.001729 ) | 0.013737 ( 0.000307 ) | 0.013588 ( 2e-06 ) | -46.569618 ( 2.343418 ) | -1.083378 ( 2.213009 ) | 0.004439 ( 5.8e-05 ) | 1 ( 0 ) |
| 0.2 | ipw logit i | 1e-04 ( 0.000382 ) | 0.973 ( 0.005126 ) | 0.012079 ( 0.00027 ) | 0.014076 ( 3e-06 ) | -30.901075 ( 2.510836 ) | 16.5298 ( 2.607113 ) | 0.000146 ( 7e-06 ) | 1 ( 0 ) |
| 0.2 | ipw logit noi | 0.09258 ( 0.000437 ) | 0 ( 0 ) | 0.013828 ( 0.000309 ) | 0.013802 ( 2e-06 ) | -47.276424 ( 2.28833 ) | -0.194945 ( 2.232892 ) | 0.008762 ( 8.2e-05 ) | 1 ( 0 ) |
| 0.3 | True model | -0.00014 ( 0.00032 ) | 0.944 ( 0.007271 ) | 0.010127 ( 0.000227 ) | 0.009758 ( 1e-06 ) | 0 ( 0 ) | -3.645512 ( 2.155644 ) | 0.000102 ( 5e-06 ) | 1 ( 0 ) |
| 0.3 | CCA | 0.096771 ( 0.000438 ) | 0 ( 0 ) | 0.013847 ( 0.00031 ) | 0.01332 ( 2e-06 ) | -46.50846 ( 2.243083 ) | -3.802969 ( 2.152141 ) | 0.009556 ( 8.5e-05 ) | 1 ( 0 ) |
| 0.3 | ipw logadd i | 0.015583 ( 0.000393 ) | 0.828 ( 0.011934 ) | 0.012423 ( 0.000278 ) | 0.013716 ( 3e-06 ) | -33.543213 ( 2.396901 ) | 10.411891 ( 2.470204 ) | 0.000397 ( 1.4e-05 ) | 1 ( 0 ) |
| 0.3 | ipw logadd noi | 0.106798 ( 0.000443 ) | 0 ( 0 ) | 0.014024 ( 0.000314 ) | 0.013577 ( 2e-06 ) | -47.848405 ( 2.265164 ) | -3.184001 ( 2.166014 ) | 0.011602 ( 9.5e-05 ) | 1 ( 0 ) |
| 0.3 | ipw logit i | -0.00012 ( 0.000397 ) | 0.975 ( 0.004937 ) | 0.01254 ( 0.000281 ) | 0.014331 ( 4e-06 ) | -34.776882 ( 2.410342 ) | 14.283996 ( 2.556926 ) | 0.000157 ( 7e-06 ) | 1 ( 0 ) |
| 0.3 | ipw logit noi | 0.137382 ( 0.000448 ) | 0 ( 0 ) | 0.014156 ( 0.000317 ) | 0.013825 ( 2e-06 ) | -48.819206 ( 2.206803 ) | -2.337623 ( 2.184957 ) | 0.019074 ( 0.000123 ) | 1 ( 0 ) |

###### Vary selection probability

###### Delta3=0

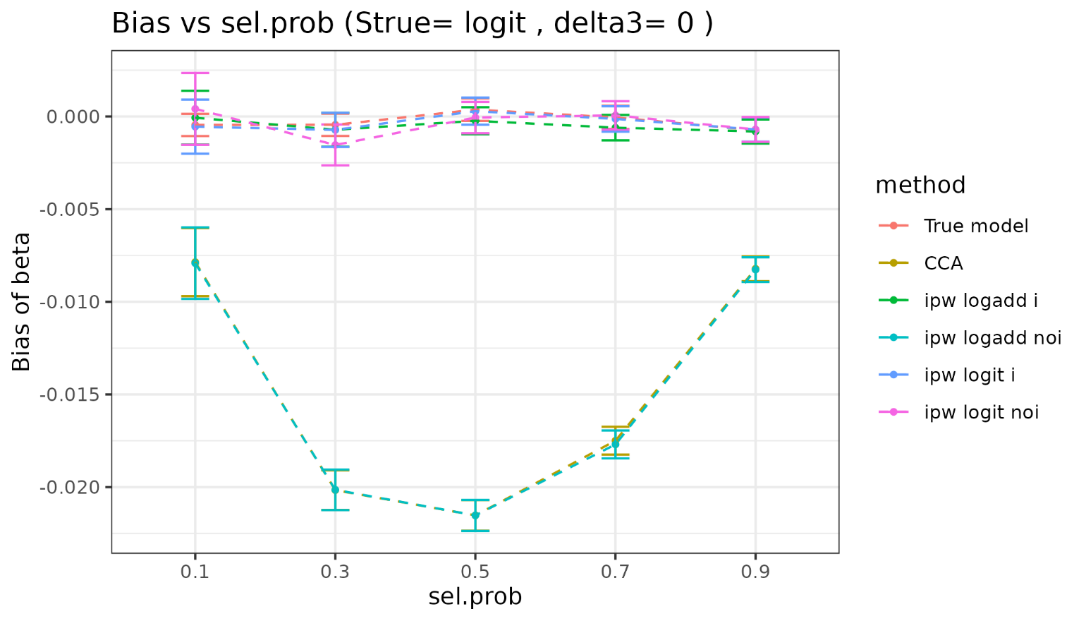

| sel.prob | method | bias | coverage | EmpSE | ModSE | relative_precision | relative_error_ModSE | MSE | power |
| --- | --- | --- | --- | --- | --- | --- | --- | --- | --- |
| 0.1 | True model | -0.000457 ( 0.000307 ) | 0.949 ( 0.006957 ) | 0.009715 ( 0.000217 ) | 0.00976 ( 1e-06 ) | 0 ( 0 ) | 0.462243 ( 2.247543 ) | 9.4e-05 ( 4e-06 ) | 1 ( 0 ) |
| 0.1 | CCA | -0.007858 ( 0.000938 ) | 0.944 ( 0.007271 ) | 0.029657 ( 0.000663 ) | 0.029431 ( 8e-06 ) | -89.26968 ( 0.642383 ) | -0.761481 ( 2.220335 ) | 0.00094 ( 4.2e-05 ) | 1 ( 0 ) |
| 0.1 | ipw logadd i | -6.6e-05 ( 0.000743 ) | 0.989 ( 0.003298 ) | 0.023507 ( 0.000526 ) | 0.031018 ( 1.3e-05 ) | -82.921166 ( 0.99073 ) | 31.952597 ( 2.952536 ) | 0.000552 ( 2.8e-05 ) | 1 ( 0 ) |
| 0.1 | ipw logadd noi | -0.007916 ( 0.000984 ) | 0.942 ( 0.007392 ) | 0.031125 ( 0.000696 ) | 0.031042 ( 1.3e-05 ) | -90.257984 ( 0.590588 ) | -0.266958 ( 2.231595 ) | 0.00103 ( 4.8e-05 ) | 1 ( 0 ) |
| 0.1 | ipw logit i | -0.000551 ( 0.000745 ) | 0.99 ( 0.003146 ) | 0.02355 ( 0.000527 ) | 0.0312 ( 1.3e-05 ) | -82.983418 ( 0.987614 ) | 32.483882 ( 2.964459 ) | 0.000554 ( 2.8e-05 ) | 1 ( 0 ) |
| 0.1 | ipw logit noi | 0.000411 ( 0.000989 ) | 0.955 ( 0.006556 ) | 0.031276 ( 7e-04 ) | 0.031192 ( 1.3e-05 ) | -90.351791 ( 0.585118 ) | -0.267423 ( 2.2316 ) | 0.000977 ( 4.6e-05 ) | 1 ( 0 ) |
| 0.3 | True model | -0.000449 ( 0.000308 ) | 0.951 ( 0.006826 ) | 0.009733 ( 0.000218 ) | 0.00976 ( 1e-06 ) | 0 ( 0 ) | 0.280413 ( 2.243476 ) | 9.5e-05 ( 4e-06 ) | 1 ( 0 ) |
| 0.3 | CCA | -0.020162 ( 0.000544 ) | 0.779 ( 0.013121 ) | 0.017213 ( 0.000385 ) | 0.017107 ( 3e-06 ) | -68.029988 ( 1.670283 ) | -0.618652 ( 2.223407 ) | 0.000703 ( 2.5e-05 ) | 1 ( 0 ) |
| 0.3 | ipw logadd i | -0.000718 ( 0.000467 ) | 0.978 ( 0.004639 ) | 0.014772 ( 0.00033 ) | 0.017618 ( 4e-06 ) | -56.591019 ( 2.079027 ) | 19.267717 ( 2.668361 ) | 0.000219 ( 1e-05 ) | 1 ( 0 ) |
| 0.3 | ipw logadd noi | -0.020145 ( 0.000557 ) | 0.78 ( 0.0131 ) | 0.017622 ( 0.000394 ) | 0.017649 ( 4e-06 ) | -69.496071 ( 1.63285 ) | 0.153067 ( 2.240715 ) | 0.000716 ( 2.6e-05 ) | 1 ( 0 ) |
| 0.3 | ipw logit i | -0.000723 ( 0.000469 ) | 0.979 ( 0.004534 ) | 0.014826 ( 0.000332 ) | 0.017824 ( 4e-06 ) | -56.904347 ( 2.070443 ) | 20.22581 ( 2.689811 ) | 0.00022 ( 1e-05 ) | 1 ( 0 ) |
| 0.3 | ipw logit noi | -0.001538 ( 0.000563 ) | 0.956 ( 0.006486 ) | 0.017803 ( 0.000398 ) | 0.017826 ( 4e-06 ) | -70.112593 ( 1.601165 ) | 0.12731 ( 2.240146 ) | 0.000319 ( 1.4e-05 ) | 1 ( 0 ) |
| 0.5 | True model | 0.000368 ( 0.000311 ) | 0.94 ( 0.00751 ) | 0.00983 ( 0.00022 ) | 0.00976 ( 1e-06 ) | 0 ( 0 ) | -0.710937 ( 2.221297 ) | 9.7e-05 ( 4e-06 ) | 1 ( 0 ) |
| 0.5 | CCA | -0.021525 ( 0.000423 ) | 0.62 ( 0.015349 ) | 0.013372 ( 0.000299 ) | 0.013373 ( 2e-06 ) | -45.961202 ( 2.371232 ) | 0.004998 ( 2.237332 ) | 0.000642 ( 1.9e-05 ) | 1 ( 0 ) |
| 0.5 | ipw logadd i | -0.000234 ( 0.000373 ) | 0.978 ( 0.004639 ) | 0.011785 ( 0.000264 ) | 0.013564 ( 2e-06 ) | -30.429827 ( 2.540719 ) | 15.091517 ( 2.574874 ) | 0.000139 ( 6e-06 ) | 1 ( 0 ) |
| 0.5 | ipw logadd noi | -0.021526 ( 0.000427 ) | 0.636 ( 0.015215 ) | 0.013492 ( 0.000302 ) | 0.013584 ( 2e-06 ) | -46.916461 ( 2.391539 ) | 0.678028 ( 2.252411 ) | 0.000645 ( 1.9e-05 ) | 1 ( 0 ) |
| 0.5 | ipw logit i | 0.000285 ( 0.000373 ) | 0.98 ( 0.004427 ) | 0.011787 ( 0.000264 ) | 0.013731 ( 2e-06 ) | -30.447348 ( 2.545692 ) | 16.493781 ( 2.606252 ) | 0.000139 ( 6e-06 ) | 1 ( 0 ) |
| 0.5 | ipw logit noi | -5.9e-05 ( 0.000431 ) | 0.956 ( 0.006486 ) | 0.013621 ( 0.000305 ) | 0.013731 ( 2e-06 ) | -47.913186 ( 2.336267 ) | 0.813417 ( 2.255443 ) | 0.000185 ( 8e-06 ) | 1 ( 0 ) |
| 0.7 | True model | -5e-05 ( 0.000319 ) | 0.944 ( 0.007271 ) | 0.010073 ( 0.000225 ) | 0.009759 ( 1e-06 ) | 0 ( 0 ) | -3.115756 ( 2.167496 ) | 0.000101 ( 5e-06 ) | 1 ( 0 ) |
| 0.7 | CCA | -0.017494 ( 0.000384 ) | 0.667 ( 0.014903 ) | 0.01215 ( 0.000272 ) | 0.011426 ( 1e-06 ) | -31.267732 ( 2.319272 ) | -5.966091 ( 2.103737 ) | 0.000454 ( 1.5e-05 ) | 1 ( 0 ) |
| 0.7 | ipw logadd i | -0.000605 ( 0.00035 ) | 0.959 ( 0.00627 ) | 0.011083 ( 0.000248 ) | 0.011484 ( 1e-06 ) | -17.389174 ( 2.071134 ) | 3.620196 ( 2.318216 ) | 0.000123 ( 6e-06 ) | 1 ( 0 ) |
| 0.7 | ipw logadd noi | -0.017696 ( 0.000385 ) | 0.657 ( 0.015012 ) | 0.012189 ( 0.000273 ) | 0.011493 ( 1e-06 ) | -31.699032 ( 2.344972 ) | -5.71193 ( 2.109436 ) | 0.000462 ( 1.5e-05 ) | 1 ( 0 ) |
| 0.7 | ipw logit i | -0.000131 ( 0.00035 ) | 0.963 ( 0.005969 ) | 0.011077 ( 0.000248 ) | 0.011592 ( 2e-06 ) | -17.307918 ( 2.06952 ) | 4.648682 ( 2.341228 ) | 0.000123 ( 6e-06 ) | 1 ( 0 ) |
| 0.7 | ipw logit noi | 6e-05 ( 0.000388 ) | 0.942 ( 0.007392 ) | 0.012271 ( 0.000275 ) | 0.011592 ( 2e-06 ) | -32.614217 ( 2.26866 ) | -5.535912 ( 2.113375 ) | 0.00015 ( 7e-06 ) | 1 ( 0 ) |
| 0.9 | True model | -0.000744 ( 0.000325 ) | 0.943 ( 0.007332 ) | 0.010266 ( 0.00023 ) | 0.009759 ( 1e-06 ) | 0 ( 0 ) | -4.936156 ( 2.126772 ) | 0.000106 ( 4e-06 ) | 1 ( 0 ) |
| 0.9 | CCA | -0.008216 ( 0.000341 ) | 0.85 ( 0.011292 ) | 0.010771 ( 0.000241 ) | 0.01021 ( 1e-06 ) | -9.158717 ( 1.721323 ) | -5.209121 ( 2.120666 ) | 0.000183 ( 7e-06 ) | 1 ( 0 ) |
| 0.9 | ipw logadd i | -0.00081 ( 0.000332 ) | 0.95 ( 0.006892 ) | 0.010512 ( 0.000235 ) | 0.010217 ( 1e-06 ) | -4.620252 ( 1.266442 ) | -2.801159 ( 2.174547 ) | 0.000111 ( 5e-06 ) | 1 ( 0 ) |
| 0.9 | ipw logadd noi | -0.008265 ( 0.000341 ) | 0.852 ( 0.011229 ) | 0.010776 ( 0.000241 ) | 0.010219 ( 1e-06 ) | -9.241793 ( 1.725853 ) | -5.17486 ( 2.121443 ) | 0.000184 ( 7e-06 ) | 1 ( 0 ) |
| 0.9 | ipw logit i | -0.000704 ( 0.000333 ) | 0.951 ( 0.006826 ) | 0.010515 ( 0.000235 ) | 0.010255 ( 1e-06 ) | -4.684249 ( 1.263745 ) | -2.472698 ( 2.181896 ) | 0.000111 ( 5e-06 ) | 1 ( 0 ) |
| 0.9 | ipw logit noi | -0.000703 ( 0.000342 ) | 0.942 ( 0.007392 ) | 0.010819 ( 0.000242 ) | 0.010255 ( 1e-06 ) | -9.963939 ( 1.674417 ) | -5.21333 ( 2.120582 ) | 0.000117 ( 5e-06 ) | 1 ( 0 ) |

###### Delta3=0.1

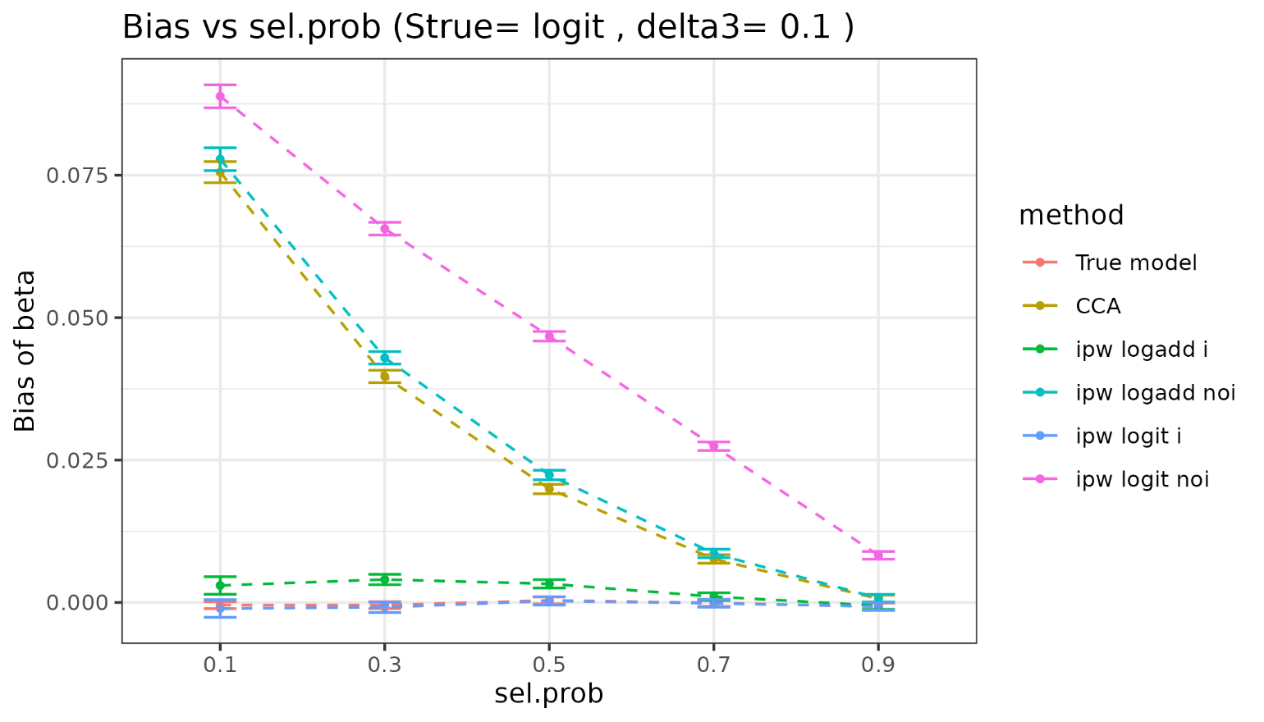

| sel.prob | method | bias | coverage | EmpSE | ModSE | relative_precision | relative_error_ModSE | MSE | power |
| --- | --- | --- | --- | --- | --- | --- | --- | --- | --- |
| 0.1 | True model | -0.000457 ( 0.000307 ) | 0.949 ( 0.006957 ) | 0.009715 ( 0.000217 ) | 0.00976 ( 1e-06 ) | 0 ( 0 ) | 0.462243 ( 2.247543 ) | 9.4e-05 ( 4e-06 ) | 1 ( 0 ) |
| 0.1 | CCA | 0.075522 ( 0.000947 ) | 0.272 ( 0.014072 ) | 0.029939 ( 0.00067 ) | 0.029317 ( 8e-06 ) | -89.470747 ( 0.630699 ) | -2.076436 ( 2.19091 ) | 0.006599 ( 0.000149 ) | 1 ( 0 ) |
| 0.1 | ipw logadd i | 0.002986 ( 0.000789 ) | 0.985 ( 0.003844 ) | 0.02494 ( 0.000558 ) | 0.031635 ( 1.5e-05 ) | -84.827568 ( 0.890956 ) | 26.842207 ( 2.838361 ) | 0.00063 ( 3.2e-05 ) | 1 ( 0 ) |
| 0.1 | ipw logadd noi | 0.077798 ( 0.001022 ) | 0.308 ( 0.014599 ) | 0.032307 ( 0.000723 ) | 0.031303 ( 1.4e-05 ) | -90.958137 ( 0.548956 ) | -3.10835 ( 2.16805 ) | 0.007095 ( 0.000166 ) | 1 ( 0 ) |
| 0.1 | ipw logit i | -0.00104 ( 0.000792 ) | 0.985 ( 0.003844 ) | 0.025055 ( 0.000561 ) | 0.031966 ( 1.7e-05 ) | -84.966141 ( 0.883753 ) | 27.581144 ( 2.854999 ) | 0.000628 ( 3.3e-05 ) | 1 ( 0 ) |
| 0.1 | ipw logit noi | 0.088836 ( 0.001028 ) | 0.205 ( 0.012766 ) | 0.032511 ( 0.000727 ) | 0.031501 ( 1.4e-05 ) | -91.070968 ( 0.542313 ) | -3.10501 ( 2.168147 ) | 0.008948 ( 0.000189 ) | 1 ( 0 ) |
| 0.3 | True model | -0.000449 ( 0.000308 ) | 0.951 ( 0.006826 ) | 0.009733 ( 0.000218 ) | 0.00976 ( 1e-06 ) | 0 ( 0 ) | 0.280413 ( 2.243476 ) | 9.5e-05 ( 4e-06 ) | 1 ( 0 ) |
| 0.3 | CCA | 0.039668 ( 0.000552 ) | 0.366 ( 0.015233 ) | 0.017442 ( 0.00039 ) | 0.017068 ( 3e-06 ) | -68.86385 ( 1.627015 ) | -2.145762 ( 2.189241 ) | 0.001877 ( 4.6e-05 ) | 1 ( 0 ) |
| 0.3 | ipw logadd i | 0.004034 ( 0.000466 ) | 0.977 ( 0.00474 ) | 0.014738 ( 0.00033 ) | 0.017779 ( 4e-06 ) | -56.392148 ( 2.082823 ) | 20.632411 ( 2.698921 ) | 0.000233 ( 1e-05 ) | 1 ( 0 ) |
| 0.3 | ipw logadd noi | 0.04294 ( 0.000564 ) | 0.336 ( 0.014937 ) | 0.017849 ( 0.000399 ) | 0.017701 ( 4e-06 ) | -70.265878 ( 1.592718 ) | -0.827512 ( 2.218783 ) | 0.002162 ( 5.1e-05 ) | 1 ( 0 ) |
| 0.3 | ipw logit i | -0.000845 ( 0.000467 ) | 0.982 ( 0.004204 ) | 0.014777 ( 0.000331 ) | 0.018125 ( 5e-06 ) | -56.619132 ( 2.081931 ) | 22.659744 ( 2.744324 ) | 0.000219 ( 1e-05 ) | 1 ( 0 ) |
| 0.3 | ipw logit noi | 0.065594 ( 0.000572 ) | 0.041 ( 0.00627 ) | 0.018097 ( 0.000405 ) | 0.017922 ( 4e-06 ) | -71.075652 ( 1.549611 ) | -0.964547 ( 2.215729 ) | 0.00463 ( 7.7e-05 ) | 1 ( 0 ) |
| 0.5 | True model | 0.000368 ( 0.000311 ) | 0.94 ( 0.00751 ) | 0.00983 ( 0.00022 ) | 0.00976 ( 1e-06 ) | 0 ( 0 ) | -0.710937 ( 2.221297 ) | 9.7e-05 ( 4e-06 ) | 1 ( 0 ) |
| 0.5 | CCA | 0.019921 ( 0.000423 ) | 0.687 ( 0.014664 ) | 0.01337 ( 0.000299 ) | 0.013359 ( 2e-06 ) | -45.939221 ( 2.364813 ) | -0.080767 ( 2.235413 ) | 0.000575 ( 2e-05 ) | 1 ( 0 ) |
| 0.5 | ipw logadd i | 0.003282 ( 0.000375 ) | 0.97 ( 0.005394 ) | 0.011851 ( 0.000265 ) | 0.01361 ( 2e-06 ) | -31.202515 ( 2.534176 ) | 14.841912 ( 2.569295 ) | 0.000151 ( 7e-06 ) | 1 ( 0 ) |
| 0.5 | ipw logadd noi | 0.022372 ( 0.000426 ) | 0.633 ( 0.015242 ) | 0.013483 ( 0.000302 ) | 0.013589 ( 2e-06 ) | -46.846292 ( 2.391429 ) | 0.781495 ( 2.254726 ) | 0.000682 ( 2.1e-05 ) | 1 ( 0 ) |
| 0.5 | ipw logit i | 0.000271 ( 0.000375 ) | 0.98 ( 0.004427 ) | 0.011857 ( 0.000265 ) | 0.013877 ( 3e-06 ) | -31.263754 ( 2.545635 ) | 17.036848 ( 2.618419 ) | 0.000141 ( 6e-06 ) | 1 ( 0 ) |
| 0.5 | ipw logit noi | 0.046717 ( 0.000432 ) | 0.071 ( 0.008122 ) | 0.013665 ( 0.000306 ) | 0.013768 ( 2e-06 ) | -48.254466 ( 2.317462 ) | 0.753983 ( 2.254115 ) | 0.002369 ( 4.2e-05 ) | 1 ( 0 ) |
| 0.7 | True model | -5e-05 ( 0.000319 ) | 0.944 ( 0.007271 ) | 0.010073 ( 0.000225 ) | 0.009759 ( 1e-06 ) | 0 ( 0 ) | -3.115756 ( 2.167496 ) | 0.000101 ( 5e-06 ) | 1 ( 0 ) |
| 0.7 | CCA | 0.007652 ( 0.000381 ) | 0.885 ( 0.010088 ) | 0.012057 ( 0.00027 ) | 0.011424 ( 1e-06 ) | -30.198077 ( 2.349499 ) | -5.253037 ( 2.119689 ) | 0.000204 ( 9e-06 ) | 1 ( 0 ) |
| 0.7 | ipw logadd i | 0.001034 ( 0.000349 ) | 0.957 ( 0.006415 ) | 0.011045 ( 0.000247 ) | 0.011492 ( 2e-06 ) | -16.824318 ( 2.091529 ) | 4.048579 ( 2.327802 ) | 0.000123 ( 5e-06 ) | 1 ( 0 ) |
| 0.7 | ipw logadd noi | 0.008602 ( 0.000382 ) | 0.868 ( 0.010704 ) | 0.012092 ( 0.000271 ) | 0.011488 ( 2e-06 ) | -30.603953 ( 2.37873 ) | -4.998864 ( 2.125389 ) | 0.00022 ( 9e-06 ) | 1 ( 0 ) |
| 0.7 | ipw logit i | -0.000153 ( 0.000349 ) | 0.969 ( 0.005481 ) | 0.011036 ( 0.000247 ) | 0.01166 ( 2e-06 ) | -16.692306 ( 2.089721 ) | 5.647362 ( 2.363576 ) | 0.000122 ( 5e-06 ) | 1 ( 0 ) |
| 0.7 | ipw logit noi | 0.027431 ( 0.000385 ) | 0.345 ( 0.015032 ) | 0.012188 ( 0.000273 ) | 0.011607 ( 2e-06 ) | -31.694901 ( 2.287391 ) | -4.772217 ( 2.130462 ) | 0.000901 ( 2.2e-05 ) | 1 ( 0 ) |
| 0.9 | True model | -0.000744 ( 0.000325 ) | 0.943 ( 0.007332 ) | 0.010266 ( 0.00023 ) | 0.009759 ( 1e-06 ) | 0 ( 0 ) | -4.936156 ( 2.126772 ) | 0.000106 ( 4e-06 ) | 1 ( 0 ) |
| 0.9 | CCA | 0.000642 ( 0.00034 ) | 0.94 ( 0.00751 ) | 0.010746 ( 0.00024 ) | 0.010211 ( 1e-06 ) | -8.733519 ( 1.77587 ) | -4.97702 ( 2.125859 ) | 0.000116 ( 5e-06 ) | 1 ( 0 ) |
| 0.9 | ipw logadd i | -0.000551 ( 0.000333 ) | 0.951 ( 0.006826 ) | 0.01052 ( 0.000235 ) | 0.010216 ( 1e-06 ) | -4.765713 ( 1.276512 ) | -2.892346 ( 2.172507 ) | 0.000111 ( 5e-06 ) | 1 ( 0 ) |
| 0.9 | ipw logadd noi | 0.000757 ( 0.00034 ) | 0.939 ( 0.007568 ) | 0.010753 ( 0.000241 ) | 0.010215 ( 1e-06 ) | -8.847093 ( 1.78027 ) | -4.998305 ( 2.125392 ) | 0.000116 ( 5e-06 ) | 1 ( 0 ) |
| 0.9 | ipw logit i | -0.00068 ( 0.000333 ) | 0.953 ( 0.006693 ) | 0.010525 ( 0.000235 ) | 0.010273 ( 1e-06 ) | -4.860249 ( 1.273392 ) | -2.39118 ( 2.18372 ) | 0.000111 ( 5e-06 ) | 1 ( 0 ) |
| 0.9 | ipw logit noi | 0.008283 ( 0.000341 ) | 0.859 ( 0.011005 ) | 0.010794 ( 0.000241 ) | 0.010259 ( 1e-06 ) | -9.539816 ( 1.713718 ) | -4.955087 ( 2.126359 ) | 0.000185 ( 7e-06 ) | 1 ( 0 ) |

##### 3.1.2 Y is binary

###### Vary delta3

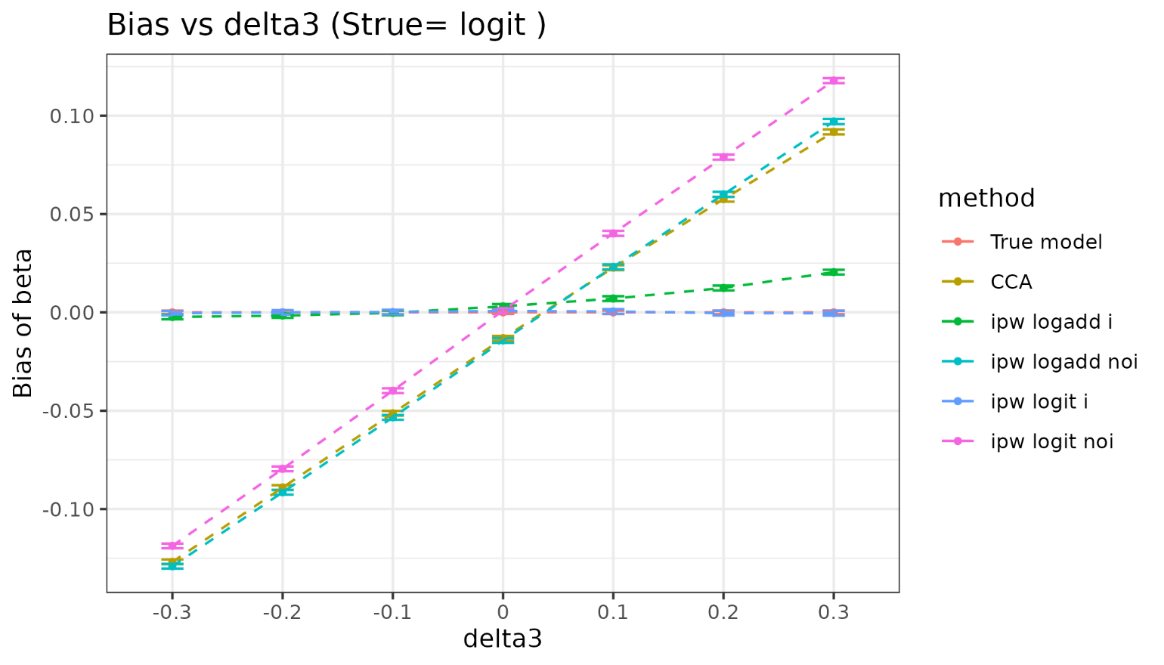

| delta3 | method | bias | coverage | EmpSE | ModSE | relative_precision | relative_error_ModSE | MSE | power |
| --- | --- | --- | --- | --- | --- | --- | --- | --- | --- |
| -0.3 | True model | 0 ( 0.00045 ) | 0.952 ( 0.00676 ) | 0.014234 ( 0.000318 ) | 0.014249 ( 1e-06 ) | 0 ( 0 ) | 0.111037 ( 2.239679 ) | 0.000202 ( 9e-06 ) | 1 ( 0 ) |
| -0.3 | CCA | -0.126873 ( 0.000605 ) | 0 ( 0 ) | 0.01912 ( 0.000428 ) | 0.019568 ( 2e-06 ) | -44.578409 ( 2.446789 ) | 2.344303 ( 2.289651 ) | 0.016462 ( 0.000154 ) | 1 ( 0 ) |
| -0.3 | ipw logadd i | -0.002347 ( 0.000579 ) | 0.96 ( 0.006197 ) | 0.018298 ( 0.000409 ) | 0.019627 ( 2e-06 ) | -39.492141 ( 2.565564 ) | 7.262578 ( 2.399683 ) | 0.00034 ( 1.5e-05 ) | 1 ( 0 ) |
| -0.3 | ipw logadd noi | -0.129068 ( 0.000604 ) | 0 ( 0 ) | 0.019114 ( 0.000428 ) | 0.019653 ( 2e-06 ) | -44.547519 ( 2.460422 ) | 2.819455 ( 2.300282 ) | 0.017024 ( 0.000157 ) | 1 ( 0 ) |
| -0.3 | ipw logit i | -0.000395 ( 0.000578 ) | 0.961 ( 0.006122 ) | 0.018287 ( 0.000409 ) | 0.019631 ( 2e-06 ) | -39.4194 ( 2.568442 ) | 7.345784 ( 2.401544 ) | 0.000334 ( 1.5e-05 ) | 1 ( 0 ) |
| -0.3 | ipw logit noi | -0.11873 ( 0.000605 ) | 0 ( 0 ) | 0.019118 ( 0.000428 ) | 0.01965 ( 2e-06 ) | -44.568454 ( 2.456087 ) | 2.786001 ( 2.299534 ) | 0.014462 ( 0.000144 ) | 1 ( 0 ) |
| -0.2 | True model | 0 ( 0.000443 ) | 0.954 ( 0.006624 ) | 0.014018 ( 0.000314 ) | 0.014249 ( 1e-06 ) | 0 ( 0 ) | 1.650235 ( 2.274113 ) | 0.000196 ( 9e-06 ) | 1 ( 0 ) |
| -0.2 | CCA | -0.089041 ( 0.000599 ) | 0.005 ( 0.00223 ) | 0.018958 ( 0.000424 ) | 0.01964 ( 2e-06 ) | -45.323829 ( 2.446676 ) | 3.597561 ( 2.31769 ) | 0.008287 ( 0.000107 ) | 1 ( 0 ) |
| -0.2 | ipw logadd i | -0.001649 ( 0.000572 ) | 0.966 ( 0.005731 ) | 0.018081 ( 0.000405 ) | 0.01971 ( 2e-06 ) | -39.894419 ( 2.589022 ) | 9.00839 ( 2.438741 ) | 0.000329 ( 1.5e-05 ) | 1 ( 0 ) |
| -0.2 | ipw logadd noi | -0.09149 ( 0.000602 ) | 0.003 ( 0.001729 ) | 0.019024 ( 0.000426 ) | 0.019752 ( 2e-06 ) | -45.705419 ( 2.437618 ) | 3.823463 ( 2.322745 ) | 0.008732 ( 0.00011 ) | 1 ( 0 ) |
| -0.2 | ipw logit i | -7e-06 ( 0.000571 ) | 0.961 ( 0.006122 ) | 0.018064 ( 0.000404 ) | 0.019714 ( 2e-06 ) | -39.778742 ( 2.59508 ) | 9.134317 ( 2.441559 ) | 0.000326 ( 1.5e-05 ) | 1 ( 0 ) |
| -0.2 | ipw logit noi | -0.079545 ( 0.000602 ) | 0.02 ( 0.004427 ) | 0.019038 ( 0.000426 ) | 0.01975 ( 2e-06 ) | -45.784369 ( 2.431085 ) | 3.737622 ( 2.320824 ) | 0.006689 ( 9.6e-05 ) | 1 ( 0 ) |
| -0.1 | True model | 0 ( 0.000453 ) | 0.947 ( 0.007085 ) | 0.014323 ( 0.00032 ) | 0.01425 ( 1e-06 ) | 0 ( 0 ) | -0.506395 ( 2.225866 ) | 0.000205 ( 9e-06 ) | 1 ( 0 ) |
| -0.1 | CCA | -0.051307 ( 0.000622 ) | 0.248 ( 0.013656 ) | 0.019681 ( 0.00044 ) | 0.019722 ( 2e-06 ) | -47.037181 ( 2.268008 ) | 0.210344 ( 2.241912 ) | 0.003019 ( 6.6e-05 ) | 1 ( 0 ) |
| -0.1 | ipw logadd i | -0.000218 ( 0.000599 ) | 0.959 ( 0.00627 ) | 0.018952 ( 0.000424 ) | 0.019825 ( 2e-06 ) | -42.882371 ( 2.383822 ) | 4.610491 ( 2.340353 ) | 0.000359 ( 1.7e-05 ) | 1 ( 0 ) |
| -0.1 | ipw logadd noi | -0.053387 ( 0.000628 ) | 0.224 ( 0.013184 ) | 0.019858 ( 0.000444 ) | 0.019864 ( 2e-06 ) | -47.976987 ( 2.247767 ) | 0.029862 ( 2.237875 ) | 0.003244 ( 6.9e-05 ) | 1 ( 0 ) |
| -0.1 | ipw logit i | 0.000166 ( 0.000599 ) | 0.96 ( 0.006197 ) | 0.018928 ( 0.000423 ) | 0.019833 ( 2e-06 ) | -42.737862 ( 2.3928 ) | 4.781767 ( 2.344185 ) | 0.000358 ( 1.7e-05 ) | 1 ( 0 ) |
| -0.1 | ipw logit noi | -0.039824 ( 0.000628 ) | 0.471 ( 0.015785 ) | 0.019872 ( 0.000445 ) | 0.019864 ( 2e-06 ) | -48.049593 ( 2.241286 ) | -0.041582 ( 2.236277 ) | 0.00198 ( 5.3e-05 ) | 1 ( 0 ) |
| 0 | True model | 0 ( 0.000442 ) | 0.953 ( 0.006693 ) | 0.013972 ( 0.000313 ) | 0.01425 ( 1e-06 ) | 0 ( 0 ) | 1.993794 ( 2.2818 ) | 0.000195 ( 9e-06 ) | 1 ( 0 ) |
| 0 | CCA | -0.013139 ( 0.000613 ) | 0.901 ( 0.009445 ) | 0.019379 ( 0.000434 ) | 0.019811 ( 2e-06 ) | -48.019637 ( 2.374298 ) | 2.231206 ( 2.287124 ) | 0.000548 ( 2.3e-05 ) | 1 ( 0 ) |
| 0 | ipw logadd i | 0.003057 ( 0.000592 ) | 0.96 ( 0.006197 ) | 0.018725 ( 0.000419 ) | 0.019968 ( 2e-06 ) | -44.327748 ( 2.445713 ) | 6.634243 ( 2.385631 ) | 0.00036 ( 1.6e-05 ) | 1 ( 0 ) |
| 0 | ipw logadd noi | -0.014279 ( 0.000622 ) | 0.891 ( 0.009855 ) | 0.019659 ( 0.00044 ) | 0.019985 ( 2e-06 ) | -49.490778 ( 2.324713 ) | 1.655763 ( 2.274251 ) | 0.00059 ( 2.5e-05 ) | 1 ( 0 ) |
| 0 | ipw logit i | 0.000767 ( 0.00059 ) | 0.962 ( 0.006046 ) | 0.018659 ( 0.000417 ) | 0.019987 ( 2e-06 ) | -43.930686 ( 2.463405 ) | 7.120119 ( 2.396502 ) | 0.000348 ( 1.6e-05 ) | 1 ( 0 ) |
| 0 | ipw logit noi | 0.000939 ( 0.000621 ) | 0.95 ( 0.006892 ) | 0.01963 ( 0.000439 ) | 0.019987 ( 2e-06 ) | -49.343474 ( 2.321451 ) | 1.816371 ( 2.277845 ) | 0.000386 ( 1.8e-05 ) | 1 ( 0 ) |
| 0.1 | True model | 0 ( 0.000454 ) | 0.952 ( 0.00676 ) | 0.014347 ( 0.000321 ) | 0.014249 ( 1e-06 ) | 0 ( 0 ) | -0.688115 ( 2.221801 ) | 0.000206 ( 9e-06 ) | 1 ( 0 ) |
| 0.1 | CCA | 0.02272 ( 0.000634 ) | 0.807 ( 0.01248 ) | 0.020045 ( 0.000448 ) | 0.019901 ( 2e-06 ) | -48.771143 ( 2.223273 ) | -0.722381 ( 2.221049 ) | 0.000918 ( 3.5e-05 ) | 1 ( 0 ) |
| 0.1 | ipw logadd i | 0.007017 ( 0.000618 ) | 0.941 ( 0.007451 ) | 0.019555 ( 0.000437 ) | 0.020127 ( 2e-06 ) | -46.17021 ( 2.288945 ) | 2.926079 ( 2.302675 ) | 0.000431 ( 2e-05 ) | 1 ( 0 ) |
| 0.1 | ipw logadd noi | 0.023145 ( 0.000643 ) | 0.809 ( 0.012431 ) | 0.020344 ( 0.000455 ) | 0.020107 ( 2e-06 ) | -50.263368 ( 2.182655 ) | -1.163341 ( 2.211185 ) | 0.000949 ( 3.6e-05 ) | 1 ( 0 ) |
| 0.1 | ipw logit i | 0.000456 ( 0.000616 ) | 0.952 ( 0.00676 ) | 0.019484 ( 0.000436 ) | 0.020176 ( 2e-06 ) | -45.77705 ( 2.314627 ) | 3.551567 ( 2.316671 ) | 0.000379 ( 1.8e-05 ) | 1 ( 0 ) |
| 0.1 | ipw logit noi | 0.040183 ( 0.000643 ) | 0.484 ( 0.015803 ) | 0.020348 ( 0.000455 ) | 0.020114 ( 2e-06 ) | -50.282489 ( 2.179518 ) | -1.151003 ( 2.211461 ) | 0.002028 ( 5.6e-05 ) | 1 ( 0 ) |
| 0.2 | True model | 0 ( 0.000448 ) | 0.951 ( 0.006826 ) | 0.014169 ( 0.000317 ) | 0.01425 ( 1e-06 ) | 0 ( 0 ) | 0.571514 ( 2.249981 ) | 0.000201 ( 9e-06 ) | 1 ( 0 ) |
| 0.2 | CCA | 0.05756 ( 0.000635 ) | 0.174 ( 0.011988 ) | 0.020066 ( 0.000449 ) | 0.019996 ( 2e-06 ) | -50.140987 ( 2.225244 ) | -0.345674 ( 2.229478 ) | 0.003715 ( 7.5e-05 ) | 1 ( 0 ) |
| 0.2 | ipw logadd i | 0.012402 ( 0.000621 ) | 0.913 ( 0.008912 ) | 0.019631 ( 0.000439 ) | 0.020307 ( 2e-06 ) | -47.905757 ( 2.25149 ) | 3.446898 ( 2.314329 ) | 0.000539 ( 2.3e-05 ) | 1 ( 0 ) |
| 0.2 | ipw logadd noi | 0.060006 ( 0.000644 ) | 0.153 ( 0.011384 ) | 0.02037 ( 0.000456 ) | 0.020236 ( 2e-06 ) | -51.619588 ( 2.184561 ) | -0.656279 ( 2.22253 ) | 0.004015 ( 7.9e-05 ) | 1 ( 0 ) |
| 0.2 | ipw logit i | -0.000353 ( 0.000618 ) | 0.961 ( 0.006122 ) | 0.019535 ( 0.000437 ) | 0.020411 ( 2e-06 ) | -47.396285 ( 2.280499 ) | 4.485225 ( 2.337564 ) | 0.000381 ( 1.6e-05 ) | 1 ( 0 ) |
| 0.2 | ipw logit noi | 0.078933 ( 0.000641 ) | 0.028 ( 0.005217 ) | 0.020281 ( 0.000454 ) | 0.020248 ( 2e-06 ) | -51.194828 ( 2.1932 ) | -0.164891 ( 2.233524 ) | 0.006641 ( 0.000103 ) | 1 ( 0 ) |
| 0.3 | True model | 0 ( 0.000442 ) | 0.946 ( 0.007147 ) | 0.013972 ( 0.000313 ) | 0.014249 ( 1e-06 ) | 0 ( 0 ) | 1.982761 ( 2.281553 ) | 0.000195 ( 9e-06 ) | 1 ( 0 ) |
| 0.3 | CCA | 0.091771 ( 0.000641 ) | 0.006 ( 0.002442 ) | 0.020281 ( 0.000454 ) | 0.020093 ( 2e-06 ) | -52.537811 ( 2.153178 ) | -0.925071 ( 2.216515 ) | 0.008833 ( 0.000119 ) | 1 ( 0 ) |
| 0.3 | ipw logadd i | 0.020456 ( 0.000629 ) | 0.836 ( 0.011709 ) | 0.019888 ( 0.000445 ) | 0.020499 ( 2e-06 ) | -50.643502 ( 2.180429 ) | 3.070201 ( 2.305903 ) | 0.000814 ( 3.1e-05 ) | 1 ( 0 ) |
| 0.3 | ipw logadd noi | 0.097029 ( 0.000653 ) | 0.002 ( 0.001413 ) | 0.020636 ( 0.000462 ) | 0.020367 ( 2e-06 ) | -54.155177 ( 2.094789 ) | -1.30367 ( 2.208046 ) | 0.00984 ( 0.000128 ) | 1 ( 0 ) |
| 0.3 | ipw logit i | -0.000405 ( 0.000629 ) | 0.953 ( 0.006693 ) | 0.019889 ( 0.000445 ) | 0.020697 ( 3e-06 ) | -50.646511 ( 2.198922 ) | 4.063096 ( 2.328124 ) | 0.000395 ( 1.8e-05 ) | 1 ( 0 ) |
| 0.3 | ipw logit noi | 0.117852 ( 0.000651 ) | 0 ( 0 ) | 0.020592 ( 0.000461 ) | 0.020384 ( 2e-06 ) | -53.958695 ( 2.094644 ) | -1.007143 ( 2.214681 ) | 0.014313 ( 0.000155 ) | 1 ( 0 ) |

###### Vary selection probability

###### Delta3=0

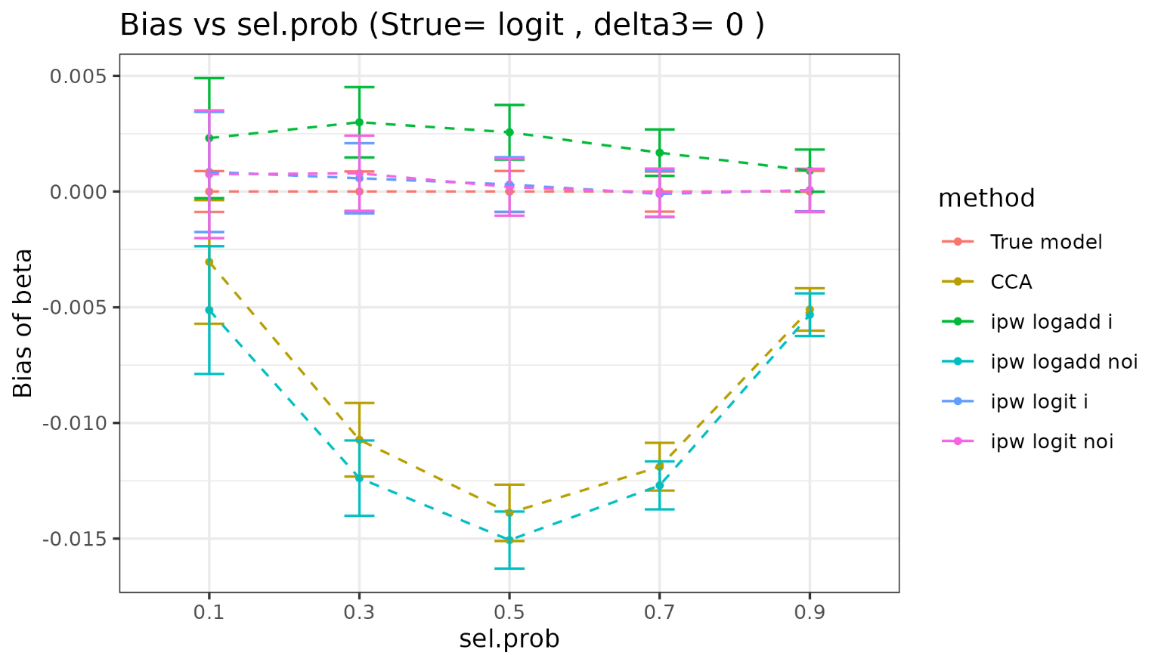

| sel.prob | method | bias | coverage | EmpSE | ModSE | relative_precision | relative_error_ModSE | MSE | power |
| --- | --- | --- | --- | --- | --- | --- | --- | --- | --- |
| 0.1 | True model | 0 ( 0.00045 ) | 0.952 ( 0.00676 ) | 0.014234 ( 0.000318 ) | 0.014249 ( 1e-06 ) | 0 ( 0 ) | 0.111037 ( 2.239679 ) | 0.000202 ( 9e-06 ) | 1 ( 0 ) |
| 0.1 | CCA | -0.003041 ( 0.001362 ) | 0.951 ( 0.006826 ) | 0.043083 ( 0.000964 ) | 0.043918 ( 1e-05 ) | -89.08486 ( 0.659311 ) | 1.938279 ( 2.280678 ) | 0.001864 ( 8.5e-05 ) | 1 ( 0 ) |
| 0.1 | ipw logadd i | 0.002309 ( 0.001324 ) | 0.962 ( 0.006046 ) | 0.041865 ( 0.000937 ) | 0.045207 ( 1.2e-05 ) | -88.440801 ( 0.690346 ) | 7.983128 ( 2.415952 ) | 0.001756 ( 8.3e-05 ) | 1 ( 0 ) |
| 0.1 | ipw logadd noi | -0.005122 ( 0.001408 ) | 0.946 ( 0.007147 ) | 0.044527 ( 0.000996 ) | 0.045232 ( 1.1e-05 ) | -89.781473 ( 0.619301 ) | 1.584532 ( 2.272782 ) | 0.002007 ( 9.3e-05 ) | 1 ( 0 ) |
| 0.1 | ipw logit i | 0.000847 ( 0.001324 ) | 0.962 ( 0.006046 ) | 0.041882 ( 0.000937 ) | 0.045257 ( 1.2e-05 ) | -88.450378 ( 0.689654 ) | 8.05634 ( 2.417594 ) | 0.001753 ( 8.3e-05 ) | 1 ( 0 ) |
| 0.1 | ipw logit noi | 0.000748 ( 0.001408 ) | 0.948 ( 0.007021 ) | 0.044523 ( 0.000996 ) | 0.045254 ( 1.2e-05 ) | -89.779568 ( 0.61941 ) | 1.642035 ( 2.274071 ) | 0.001981 ( 9.3e-05 ) | 1 ( 0 ) |
| 0.3 | True model | 0 ( 0.000443 ) | 0.954 ( 0.006624 ) | 0.014018 ( 0.000314 ) | 0.014249 ( 1e-06 ) | 0 ( 0 ) | 1.650235 ( 2.274113 ) | 0.000196 ( 9e-06 ) | 1 ( 0 ) |
| 0.3 | CCA | -0.010727 ( 0.000812 ) | 0.929 ( 0.008122 ) | 0.025679 ( 0.000574 ) | 0.025451 ( 3e-06 ) | -70.198922 ( 1.548897 ) | -0.886022 ( 2.217404 ) | 0.000774 ( 3.5e-05 ) | 1 ( 0 ) |
| 0.3 | ipw logadd i | 0.002996 ( 0.000775 ) | 0.952 ( 0.00676 ) | 0.024511 ( 0.000548 ) | 0.025868 ( 4e-06 ) | -67.291592 ( 1.653564 ) | 5.538905 ( 2.36115 ) | 0.000609 ( 2.8e-05 ) | 1 ( 0 ) |
| 0.3 | ipw logadd noi | -0.012385 ( 0.000831 ) | 0.919 ( 0.008628 ) | 0.026291 ( 0.000588 ) | 0.025895 ( 4e-06 ) | -71.571192 ( 1.485342 ) | -1.507864 ( 2.203496 ) | 0.000844 ( 3.8e-05 ) | 1 ( 0 ) |
| 0.3 | ipw logit i | 0.000575 ( 0.000774 ) | 0.961 ( 0.006122 ) | 0.024462 ( 0.000547 ) | 0.025908 ( 4e-06 ) | -67.160781 ( 1.660695 ) | 5.910877 ( 2.369474 ) | 0.000598 ( 2.8e-05 ) | 1 ( 0 ) |
| 0.3 | ipw logit noi | 0.00079 ( 0.000829 ) | 0.946 ( 0.007147 ) | 0.026228 ( 0.000587 ) | 0.025907 ( 4e-06 ) | -71.434462 ( 1.493758 ) | -1.224666 ( 2.209832 ) | 0.000688 ( 3.3e-05 ) | 1 ( 0 ) |
| 0.5 | True model | 0 ( 0.000453 ) | 0.947 ( 0.007085 ) | 0.014323 ( 0.00032 ) | 0.01425 ( 1e-06 ) | 0 ( 0 ) | -0.506395 ( 2.225866 ) | 0.000205 ( 9e-06 ) | 1 ( 0 ) |
| 0.5 | CCA | -0.013886 ( 0.000621 ) | 0.91 ( 0.00905 ) | 0.019634 ( 0.000439 ) | 0.019811 ( 2e-06 ) | -46.781399 ( 2.333793 ) | 0.901331 ( 2.257371 ) | 0.000578 ( 2.5e-05 ) | 1 ( 0 ) |
| 0.5 | ipw logadd i | 0.002563 ( 0.000602 ) | 0.957 ( 0.006415 ) | 0.019036 ( 0.000426 ) | 0.019967 ( 2e-06 ) | -43.38978 ( 2.437993 ) | 4.887061 ( 2.346542 ) | 0.000369 ( 1.7e-05 ) | 1 ( 0 ) |
| 0.5 | ipw logadd noi | -0.015061 ( 0.000628 ) | 0.907 ( 0.009184 ) | 0.019848 ( 0.000444 ) | 0.019984 ( 2e-06 ) | -47.926404 ( 2.311374 ) | 0.68353 ( 2.2525 ) | 0.00062 ( 2.7e-05 ) | 1 ( 0 ) |
| 0.5 | ipw logit i | 0.000306 ( 0.000601 ) | 0.964 ( 0.005891 ) | 0.018998 ( 0.000425 ) | 0.019986 ( 2e-06 ) | -43.158887 ( 2.452768 ) | 5.204791 ( 2.353651 ) | 0.000361 ( 1.7e-05 ) | 1 ( 0 ) |
| 0.5 | ipw logit noi | 0.000192 ( 0.000628 ) | 0.948 ( 0.007021 ) | 0.019853 ( 0.000444 ) | 0.019986 ( 2e-06 ) | -47.952175 ( 2.304773 ) | 0.670941 ( 2.252218 ) | 0.000394 ( 1.9e-05 ) | 1 ( 0 ) |
| 0.7 | True model | 0 ( 0.000442 ) | 0.953 ( 0.006693 ) | 0.013972 ( 0.000313 ) | 0.01425 ( 1e-06 ) | 0 ( 0 ) | 1.993794 ( 2.2818 ) | 0.000195 ( 9e-06 ) | 1 ( 0 ) |
| 0.7 | CCA | -0.011892 ( 0.000526 ) | 0.893 ( 0.009775 ) | 0.016629 ( 0.000372 ) | 0.016844 ( 1e-06 ) | -29.408889 ( 2.520627 ) | 1.292384 ( 2.266113 ) | 0.000418 ( 1.8e-05 ) | 1 ( 0 ) |
| 0.7 | ipw logadd i | 0.001681 ( 0.000511 ) | 0.961 ( 0.006122 ) | 0.016172 ( 0.000362 ) | 0.01689 ( 1e-06 ) | -25.357501 ( 2.478038 ) | 4.444958 ( 2.336643 ) | 0.000264 ( 1.2e-05 ) | 1 ( 0 ) |
| 0.7 | ipw logadd noi | -0.012699 ( 0.000529 ) | 0.886 ( 0.01005 ) | 0.016742 ( 0.000375 ) | 0.016898 ( 1e-06 ) | -30.354552 ( 2.504038 ) | 0.93264 ( 2.258065 ) | 0.000441 ( 1.9e-05 ) | 1 ( 0 ) |
| 0.7 | ipw logit i | -0.000104 ( 0.00051 ) | 0.965 ( 0.005812 ) | 0.016123 ( 0.000361 ) | 0.016897 ( 1e-06 ) | -24.906011 ( 2.494078 ) | 4.800412 ( 2.344596 ) | 0.00026 ( 1.2e-05 ) | 1 ( 0 ) |
| 0.7 | ipw logit noi | -4.6e-05 ( 0.00053 ) | 0.95 ( 0.006892 ) | 0.016767 ( 0.000375 ) | 0.016897 ( 1e-06 ) | -30.563358 ( 2.478886 ) | 0.774696 ( 2.254532 ) | 0.000281 ( 1.3e-05 ) | 1 ( 0 ) |
| 0.9 | True model | 0 ( 0.000454 ) | 0.952 ( 0.00676 ) | 0.014347 ( 0.000321 ) | 0.014249 ( 1e-06 ) | 0 ( 0 ) | -0.688115 ( 2.221801 ) | 0.000206 ( 9e-06 ) | 1 ( 0 ) |
| 0.9 | CCA | -0.005094 ( 0.000469 ) | 0.939 ( 0.007568 ) | 0.014829 ( 0.000332 ) | 0.014959 ( 1e-06 ) | -6.39297 ( 1.808917 ) | 0.875734 ( 2.256788 ) | 0.000246 ( 1.1e-05 ) | 1 ( 0 ) |
| 0.9 | ipw logadd i | 0.000903 ( 0.000466 ) | 0.953 ( 0.006693 ) | 0.014744 ( 0.00033 ) | 0.014964 ( 1e-06 ) | -5.305378 ( 1.698187 ) | 1.491053 ( 2.270554 ) | 0.000218 ( 1e-05 ) | 1 ( 0 ) |
| 0.9 | ipw logadd noi | -0.005322 ( 0.000469 ) | 0.936 ( 0.00774 ) | 0.014845 ( 0.000332 ) | 0.014965 ( 1e-06 ) | -6.589102 ( 1.815571 ) | 0.807401 ( 2.255259 ) | 0.000248 ( 1.1e-05 ) | 1 ( 0 ) |
| 0.9 | ipw logit i | 6.5e-05 ( 0.000466 ) | 0.951 ( 0.006826 ) | 0.01473 ( 0.00033 ) | 0.014964 ( 1e-06 ) | -5.130506 ( 1.702502 ) | 1.588761 ( 2.27274 ) | 0.000217 ( 1e-05 ) | 1 ( 0 ) |
| 0.9 | ipw logit noi | 5.3e-05 ( 0.00047 ) | 0.95 ( 0.006892 ) | 0.014861 ( 0.000332 ) | 0.014964 ( 1e-06 ) | -6.798483 ( 1.802912 ) | 0.691632 ( 2.252669 ) | 0.000221 ( 1e-05 ) | 1 ( 0 ) |

###### Delta3=0.1

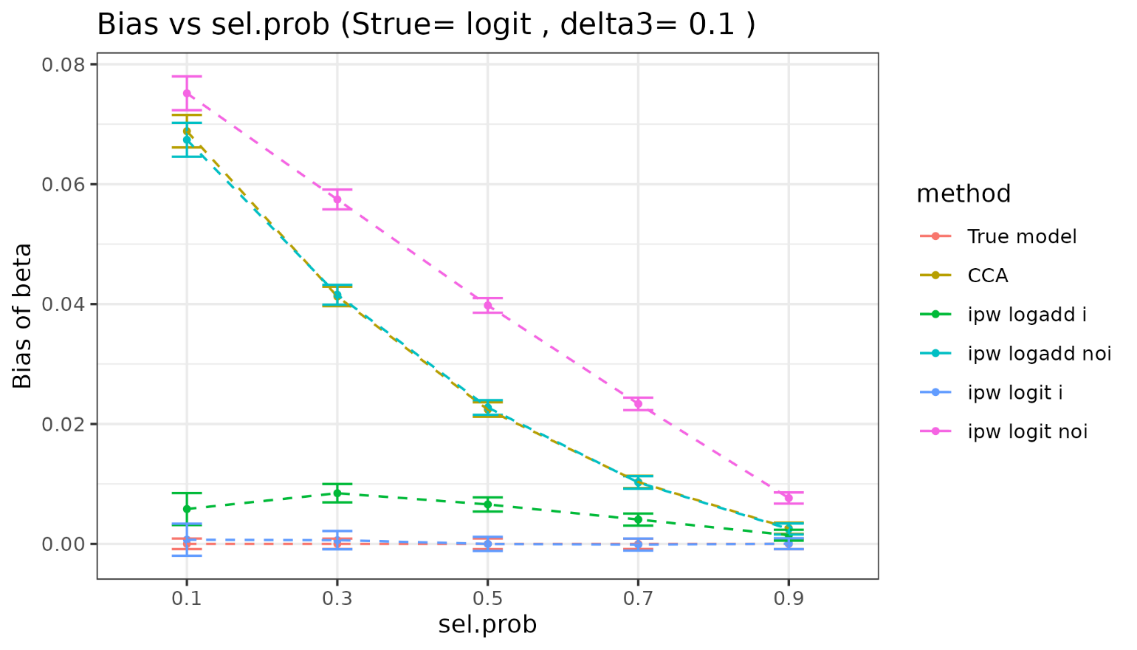

| sel.prob | method | bias | coverage | EmpSE | ModSE | relative_precision | relative_error_ModSE | MSE | power |
| --- | --- | --- | --- | --- | --- | --- | --- | --- | --- |
| 0.1 | True model | 0 ( 0.00045 ) | 0.952 ( 0.00676 ) | 0.014234 ( 0.000318 ) | 0.014249 ( 1e-06 ) | 0 ( 0 ) | 0.111037 ( 2.239679 ) | 0.000202 ( 9e-06 ) | 1 ( 0 ) |
| 0.1 | CCA | 0.068833 ( 0.001382 ) | 0.669 ( 0.014881 ) | 0.043696 ( 0.000978 ) | 0.044185 ( 1.1e-05 ) | -89.389379 ( 0.642952 ) | 1.117349 ( 2.26232 ) | 0.006645 ( 0.000211 ) | 1 ( 0 ) |
| 0.1 | ipw logadd i | 0.005812 ( 0.001364 ) | 0.956 ( 0.006486 ) | 0.043147 ( 0.000965 ) | 0.046132 ( 1.3e-05 ) | -89.117668 ( 0.654107 ) | 6.91635 ( 2.392119 ) | 0.001894 ( 8.9e-05 ) | 1 ( 0 ) |
| 0.1 | ipw logadd noi | 0.067392 ( 0.001444 ) | 0.696 ( 0.014546 ) | 0.045669 ( 0.001022 ) | 0.045829 ( 1.2e-05 ) | -90.285986 ( 0.591532 ) | 0.35197 ( 2.245219 ) | 0.006625 ( 0.000218 ) | 1 ( 0 ) |
| 0.1 | ipw logit i | 0.000701 ( 0.001366 ) | 0.956 ( 0.006486 ) | 0.043212 ( 0.000967 ) | 0.046261 ( 1.4e-05 ) | -89.149921 ( 0.652058 ) | 7.056207 ( 2.395261 ) | 0.001866 ( 8.8e-05 ) | 1 ( 0 ) |
| 0.1 | ipw logit noi | 0.075152 ( 0.001444 ) | 0.623 ( 0.015326 ) | 0.045668 ( 0.001022 ) | 0.045865 ( 1.2e-05 ) | -90.285623 ( 0.591576 ) | 0.431262 ( 2.246996 ) | 0.007731 ( 0.000238 ) | 1 ( 0 ) |
| 0.3 | True model | 0 ( 0.000443 ) | 0.954 ( 0.006624 ) | 0.014018 ( 0.000314 ) | 0.014249 ( 1e-06 ) | 0 ( 0 ) | 1.650235 ( 2.274113 ) | 0.000196 ( 9e-06 ) | 1 ( 0 ) |
| 0.3 | CCA | 0.041262 ( 0.000817 ) | 0.641 ( 0.01517 ) | 0.025826 ( 0.000578 ) | 0.025591 ( 3e-06 ) | -70.538017 ( 1.550971 ) | -0.908188 ( 2.216909 ) | 0.002369 ( 7.6e-05 ) | 1 ( 0 ) |
| 0.3 | ipw logadd i | 0.008449 ( 0.000781 ) | 0.945 ( 0.007209 ) | 0.024696 ( 0.000553 ) | 0.026201 ( 4e-06 ) | -67.781254 ( 1.65998 ) | 6.094419 ( 2.373584 ) | 0.000681 ( 3.2e-05 ) | 1 ( 0 ) |
| 0.3 | ipw logadd noi | 0.041547 ( 0.000842 ) | 0.659 ( 0.014991 ) | 0.02663 ( 0.000596 ) | 0.026128 ( 4e-06 ) | -72.290174 ( 1.468782 ) | -1.88326 ( 2.195099 ) | 0.002435 ( 8e-05 ) | 1 ( 0 ) |
| 0.3 | ipw logit i | 0.000616 ( 0.000779 ) | 0.96 ( 0.006197 ) | 0.024625 ( 0.000551 ) | 0.026299 ( 4e-06 ) | -67.595555 ( 1.671238 ) | 6.796245 ( 2.389291 ) | 0.000606 ( 2.9e-05 ) | 1 ( 0 ) |
| 0.3 | ipw logit noi | 0.05744 ( 0.000841 ) | 0.414 ( 0.015576 ) | 0.026598 ( 0.000595 ) | 0.026149 ( 4e-06 ) | -72.223882 ( 1.474103 ) | -1.687108 ( 2.199488 ) | 0.004006 ( 0.000104 ) | 1 ( 0 ) |
| 0.5 | True model | 0 ( 0.000453 ) | 0.947 ( 0.007085 ) | 0.014323 ( 0.00032 ) | 0.01425 ( 1e-06 ) | 0 ( 0 ) | -0.506395 ( 2.225866 ) | 0.000205 ( 9e-06 ) | 1 ( 0 ) |
| 0.5 | CCA | 0.022398 ( 0.000618 ) | 0.804 ( 0.012553 ) | 0.019552 ( 0.000437 ) | 0.019902 ( 2e-06 ) | -46.333649 ( 2.361842 ) | 1.794557 ( 2.277357 ) | 0.000884 ( 3.3e-05 ) | 1 ( 0 ) |
| 0.5 | ipw logadd i | 0.006582 ( 0.000602 ) | 0.945 ( 0.007209 ) | 0.01904 ( 0.000426 ) | 0.020129 ( 2e-06 ) | -43.411706 ( 2.439274 ) | 5.719462 ( 2.365168 ) | 0.000405 ( 1.9e-05 ) | 1 ( 0 ) |
| 0.5 | ipw logadd noi | 0.022746 ( 0.000626 ) | 0.806 ( 0.012505 ) | 0.019803 ( 0.000443 ) | 0.020109 ( 2e-06 ) | -47.690103 ( 2.334151 ) | 1.542612 ( 2.271721 ) | 0.000909 ( 3.4e-05 ) | 1 ( 0 ) |
| 0.5 | ipw logit i | -9e-06 ( 6e-04 ) | 0.961 ( 0.006122 ) | 0.018985 ( 0.000425 ) | 0.020178 ( 2e-06 ) | -43.08403 ( 2.45967 ) | 6.282344 ( 2.377763 ) | 0.00036 ( 1.7e-05 ) | 1 ( 0 ) |
| 0.5 | ipw logit noi | 0.03978 ( 0.000626 ) | 0.497 ( 0.015811 ) | 0.019793 ( 0.000443 ) | 0.020115 ( 2e-06 ) | -47.633676 ( 2.327228 ) | 1.629531 ( 2.273667 ) | 0.001974 ( 5.3e-05 ) | 1 ( 0 ) |
| 0.7 | True model | 0 ( 0.000442 ) | 0.953 ( 0.006693 ) | 0.013972 ( 0.000313 ) | 0.01425 ( 1e-06 ) | 0 ( 0 ) | 1.993794 ( 2.2818 ) | 0.000195 ( 9e-06 ) | 1 ( 0 ) |
| 0.7 | CCA | 0.010321 ( 0.000526 ) | 0.921 ( 0.00853 ) | 0.016639 ( 0.000372 ) | 0.0169 ( 1e-06 ) | -29.488289 ( 2.540503 ) | 1.570543 ( 2.272337 ) | 0.000383 ( 1.6e-05 ) | 1 ( 0 ) |
| 0.7 | ipw logadd i | 0.004062 ( 0.000513 ) | 0.95 ( 0.006892 ) | 0.016228 ( 0.000363 ) | 0.016967 ( 1e-06 ) | -25.874203 ( 2.490995 ) | 4.556196 ( 2.339133 ) | 0.00028 ( 1.2e-05 ) | 1 ( 0 ) |
| 0.7 | ipw logadd noi | 0.010246 ( 0.000531 ) | 0.919 ( 0.008628 ) | 0.016781 ( 0.000375 ) | 0.016963 ( 1e-06 ) | -30.677463 ( 2.517117 ) | 1.08763 ( 2.261534 ) | 0.000386 ( 1.6e-05 ) | 1 ( 0 ) |
| 0.7 | ipw logit i | -0.000124 ( 0.000511 ) | 0.962 ( 0.006046 ) | 0.016168 ( 0.000362 ) | 0.016984 ( 1e-06 ) | -25.321734 ( 2.514142 ) | 5.045624 ( 2.350083 ) | 0.000261 ( 1.2e-05 ) | 1 ( 0 ) |
| 0.7 | ipw logit noi | 0.023357 ( 0.000531 ) | 0.715 ( 0.014275 ) | 0.016788 ( 0.000376 ) | 0.016963 ( 1e-06 ) | -30.741429 ( 2.492498 ) | 1.042034 ( 2.260514 ) | 0.000827 ( 2.7e-05 ) | 1 ( 0 ) |
| 0.9 | True model | 0 ( 0.000454 ) | 0.952 ( 0.00676 ) | 0.014347 ( 0.000321 ) | 0.014249 ( 1e-06 ) | 0 ( 0 ) | -0.688115 ( 2.221801 ) | 0.000206 ( 9e-06 ) | 1 ( 0 ) |
| 0.9 | CCA | 0.002612 ( 0.000471 ) | 0.945 ( 0.007209 ) | 0.014884 ( 0.000333 ) | 0.014979 ( 1e-06 ) | -7.086868 ( 1.823843 ) | 0.633158 ( 2.251361 ) | 0.000228 ( 1e-05 ) | 1 ( 0 ) |
| 0.9 | ipw logadd i | 0.001444 ( 0.000467 ) | 0.951 ( 0.006826 ) | 0.014771 ( 0.00033 ) | 0.014985 ( 1e-06 ) | -5.651404 ( 1.701197 ) | 1.452972 ( 2.269702 ) | 0.00022 ( 1e-05 ) | 1 ( 0 ) |
| 0.9 | ipw logadd noi | 0.002484 ( 0.000471 ) | 0.944 ( 0.007271 ) | 0.014902 ( 0.000333 ) | 0.014985 ( 1e-06 ) | -7.305251 ( 1.831471 ) | 0.558381 ( 2.249688 ) | 0.000228 ( 1e-05 ) | 1 ( 0 ) |
| 0.9 | ipw logit i | 2.7e-05 ( 0.000466 ) | 0.952 ( 0.00676 ) | 0.014749 ( 0.00033 ) | 0.014987 ( 1e-06 ) | -5.377097 ( 1.7071 ) | 1.611069 ( 2.273239 ) | 0.000217 ( 1e-05 ) | 1 ( 0 ) |
| 0.9 | ipw logit noi | 0.007647 ( 0.000472 ) | 0.92 ( 0.008579 ) | 0.014921 ( 0.000334 ) | 0.014985 ( 1e-06 ) | -7.545975 ( 1.817109 ) | 0.425933 ( 2.246725 ) | 0.000281 ( 1.2e-05 ) | 1 ( 0 ) |

#### 3.2 Strue=log-additive

##### 3.2.1 Y is continuous

###### Vary delta3

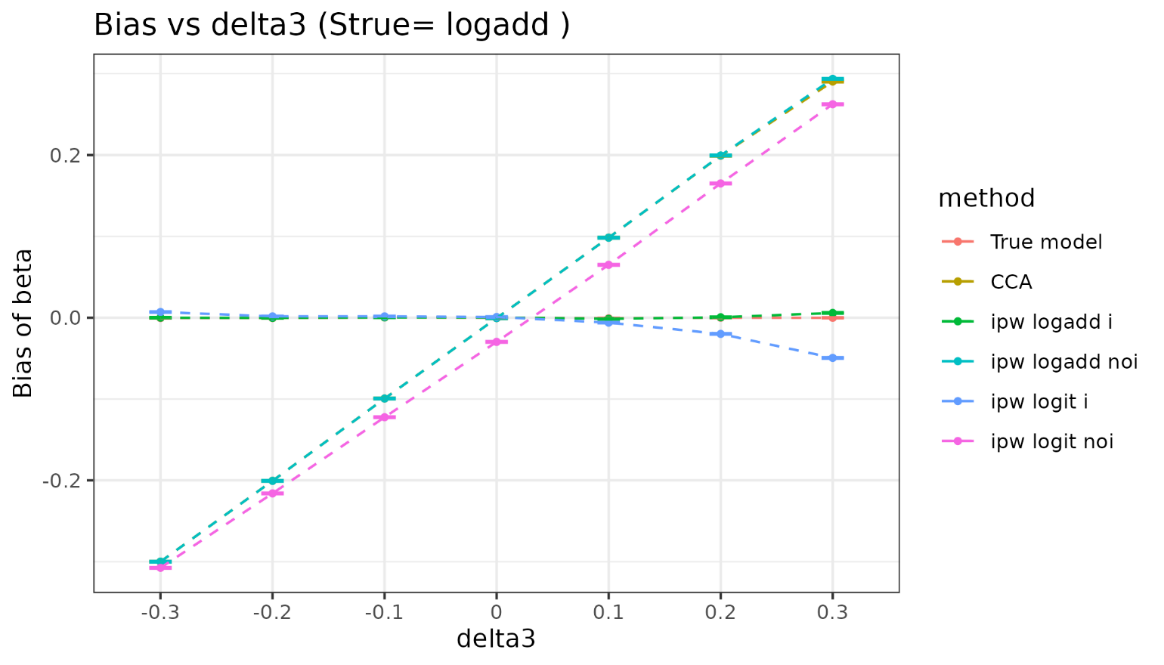

| delta3 | method | bias | coverage | EmpSE | ModSE | relative_precision | relative_error_ModSE | MSE | power |
| --- | --- | --- | --- | --- | --- | --- | --- | --- | --- |
| -0.3 | True model | -0.000457 ( 0.000307 ) | 0.949 ( 0.006957 ) | 0.009715 ( 0.000217 ) | 0.00976 ( 1e-06 ) | 0 ( 0 ) | 0.462243 ( 2.247543 ) | 9.4e-05 ( 4e-06 ) | 1 ( 0 ) |
| -0.3 | CCA | -0.299896 ( 0.000474 ) | 0 ( 0 ) | 0.014989 ( 0.000335 ) | 0.014732 ( 2e-06 ) | -57.992501 ( 1.97793 ) | -1.714813 ( 2.198868 ) | 0.090162 ( 0.000284 ) | 1 ( 0 ) |
| -0.3 | ipw logadd i | -1.3e-05 ( 0.000407 ) | 0.977 ( 0.00474 ) | 0.012865 ( 0.000288 ) | 0.0151 ( 3e-06 ) | -42.981647 ( 2.315283 ) | 17.368549 ( 2.625887 ) | 0.000165 ( 7e-06 ) | 1 ( 0 ) |
| -0.3 | ipw logadd noi | -0.299913 ( 0.000474 ) | 0 ( 0 ) | 0.014994 ( 0.000335 ) | 0.01474 ( 3e-06 ) | -58.020532 ( 1.979711 ) | -1.690843 ( 2.199441 ) | 0.090172 ( 0.000285 ) | 1 ( 0 ) |
| -0.3 | ipw logit i | 0.006998 ( 0.000408 ) | 0.961 ( 0.006122 ) | 0.012897 ( 0.000289 ) | 0.015333 ( 4e-06 ) | -43.263292 ( 2.31716 ) | 18.883576 ( 2.659814 ) | 0.000215 ( 9e-06 ) | 1 ( 0 ) |
| -0.3 | ipw logit noi | -0.30746 ( 0.000476 ) | 0 ( 0 ) | 0.015046 ( 0.000337 ) | 0.014748 ( 3e-06 ) | -58.31385 ( 1.965116 ) | -1.98153 ( 2.192938 ) | 0.094758 ( 0.000293 ) | 1 ( 0 ) |
| -0.2 | True model | -0.000449 ( 0.000308 ) | 0.951 ( 0.006826 ) | 0.009733 ( 0.000218 ) | 0.00976 ( 1e-06 ) | 0 ( 0 ) | 0.280413 ( 2.243476 ) | 9.5e-05 ( 4e-06 ) | 1 ( 0 ) |
| -0.2 | CCA | -0.200506 ( 0.000462 ) | 0 ( 0 ) | 0.014605 ( 0.000327 ) | 0.014791 ( 2e-06 ) | -55.593456 ( 2.078586 ) | 1.27135 ( 2.265675 ) | 0.040416 ( 0.000185 ) | 1 ( 0 ) |
| -0.2 | ipw logadd i | -0.000298 ( 0.000402 ) | 0.977 ( 0.00474 ) | 0.012712 ( 0.000284 ) | 0.014903 ( 3e-06 ) | -41.379087 ( 2.326538 ) | 17.236705 ( 2.622906 ) | 0.000162 ( 7e-06 ) | 1 ( 0 ) |
| -0.2 | ipw logadd noi | -0.200531 ( 0.000462 ) | 0 ( 0 ) | 0.014613 ( 0.000327 ) | 0.014821 ( 3e-06 ) | -55.639796 ( 2.079406 ) | 1.423226 ( 2.26911 ) | 0.040426 ( 0.000186 ) | 1 ( 0 ) |
| -0.2 | ipw logit i | 0.00162 ( 0.000402 ) | 0.974 ( 0.005032 ) | 0.01272 ( 0.000285 ) | 0.014968 ( 3e-06 ) | -41.451607 ( 2.326775 ) | 17.672289 ( 2.632654 ) | 0.000164 ( 7e-06 ) | 1 ( 0 ) |
| -0.2 | ipw logit noi | -0.215979 ( 0.000463 ) | 0 ( 0 ) | 0.01463 ( 0.000327 ) | 0.014852 ( 3e-06 ) | -55.744018 ( 2.070286 ) | 1.515913 ( 2.271185 ) | 0.046861 ( 2e-04 ) | 1 ( 0 ) |
| -0.1 | True model | 0.000368 ( 0.000311 ) | 0.94 ( 0.00751 ) | 0.00983 ( 0.00022 ) | 0.00976 ( 1e-06 ) | 0 ( 0 ) | -0.710937 ( 2.221297 ) | 9.7e-05 ( 4e-06 ) | 1 ( 0 ) |
| -0.1 | CCA | -0.099396 ( 0.000444 ) | 0 ( 0 ) | 0.01404 ( 0.000314 ) | 0.014807 ( 2e-06 ) | -50.981689 ( 2.342835 ) | 5.462623 ( 2.359444 ) | 0.010077 ( 8.8e-05 ) | 1 ( 0 ) |
| -0.1 | ipw logadd i | 0.000379 ( 0.000385 ) | 0.985 ( 0.003844 ) | 0.012187 ( 0.000273 ) | 0.014835 ( 3e-06 ) | -34.936006 ( 2.547166 ) | 21.732189 ( 2.723473 ) | 0.000149 ( 7e-06 ) | 1 ( 0 ) |
| -0.1 | ipw logadd noi | -0.09942 ( 0.000445 ) | 0 ( 0 ) | 0.014076 ( 0.000315 ) | 0.014873 ( 3e-06 ) | -51.227135 ( 2.342411 ) | 5.661144 ( 2.363924 ) | 0.010082 ( 8.8e-05 ) | 1 ( 0 ) |
| -0.1 | ipw logit i | 0.001607 ( 0.000385 ) | 0.981 ( 0.004317 ) | 0.012189 ( 0.000273 ) | 0.014859 ( 3e-06 ) | -34.956145 ( 2.546687 ) | 21.908987 ( 2.727427 ) | 0.000151 ( 7e-06 ) | 1 ( 0 ) |
| -0.1 | ipw logit noi | -0.122362 ( 0.000448 ) | 0 ( 0 ) | 0.014169 ( 0.000317 ) | 0.014941 ( 3e-06 ) | -51.866503 ( 2.313586 ) | 5.451596 ( 2.35924 ) | 0.015173 ( 0.000109 ) | 1 ( 0 ) |
| 0 | True model | -5e-05 ( 0.000319 ) | 0.944 ( 0.007271 ) | 0.010073 ( 0.000225 ) | 0.009759 ( 1e-06 ) | 0 ( 0 ) | -3.115756 ( 2.167496 ) | 0.000101 ( 5e-06 ) | 1 ( 0 ) |
| 0 | CCA | -0.000501 ( 0.000471 ) | 0.942 ( 0.007392 ) | 0.014881 ( 0.000333 ) | 0.014789 ( 2e-06 ) | -54.179885 ( 2.154764 ) | -0.622209 ( 2.223312 ) | 0.000221 ( 1e-05 ) | 1 ( 0 ) |
| 0 | ipw logadd i | 0.000268 ( 0.000397 ) | 0.982 ( 0.004204 ) | 0.012544 ( 0.000281 ) | 0.014898 ( 3e-06 ) | -35.510355 ( 2.556422 ) | 18.771009 ( 2.657228 ) | 0.000157 ( 7e-06 ) | 1 ( 0 ) |
| 0 | ipw logadd noi | -0.00051 ( 0.000473 ) | 0.945 ( 0.007209 ) | 0.014945 ( 0.000334 ) | 0.014899 ( 3e-06 ) | -54.571924 ( 2.171044 ) | -0.311393 ( 2.230303 ) | 0.000223 ( 1e-05 ) | 1 ( 0 ) |
| 0 | ipw logit i | 0.000824 ( 0.000397 ) | 0.98 ( 0.004427 ) | 0.012542 ( 0.000281 ) | 0.014962 ( 3e-06 ) | -35.496232 ( 2.558717 ) | 19.294982 ( 2.668953 ) | 0.000158 ( 7e-06 ) | 1 ( 0 ) |
| 0 | ipw logit noi | -0.02972 ( 0.000477 ) | 0.461 ( 0.015763 ) | 0.015083 ( 0.000337 ) | 0.015019 ( 3e-06 ) | -55.394294 ( 2.132647 ) | -0.419101 ( 2.2279 ) | 0.001111 ( 2.8e-05 ) | 1 ( 0 ) |
| 0.1 | True model | -0.000744 ( 0.000325 ) | 0.943 ( 0.007332 ) | 0.010266 ( 0.00023 ) | 0.009759 ( 1e-06 ) | 0 ( 0 ) | -4.936156 ( 2.126772 ) | 0.000106 ( 4e-06 ) | 1 ( 0 ) |
| 0.1 | CCA | 0.098445 ( 0.000469 ) | 0 ( 0 ) | 0.014842 ( 0.000332 ) | 0.014729 ( 2e-06 ) | -52.156008 ( 2.242074 ) | -0.763059 ( 2.220159 ) | 0.009912 ( 9.3e-05 ) | 1 ( 0 ) |
| 0.1 | ipw logadd i | -0.001415 ( 0.000405 ) | 0.979 ( 0.004534 ) | 0.012807 ( 0.000287 ) | 0.015099 ( 3e-06 ) | -35.741676 ( 2.586312 ) | 17.900622 ( 2.637788 ) | 0.000166 ( 7e-06 ) | 1 ( 0 ) |
| 0.1 | ipw logadd noi | 0.09838 ( 0.000472 ) | 0 ( 0 ) | 0.01492 ( 0.000334 ) | 0.014905 ( 3e-06 ) | -52.656887 ( 2.248243 ) | -0.104888 ( 2.234927 ) | 0.009901 ( 9.3e-05 ) | 1 ( 0 ) |
| 0.1 | ipw logit i | -0.006015 ( 0.000407 ) | 0.971 ( 0.005307 ) | 0.012867 ( 0.000288 ) | 0.015333 ( 4e-06 ) | -36.342366 ( 2.577118 ) | 19.159613 ( 2.665984 ) | 0.000202 ( 9e-06 ) | 1 ( 0 ) |
| 0.1 | ipw logit noi | 0.065007 ( 0.000479 ) | 0.008 ( 0.002817 ) | 0.015137 ( 0.000339 ) | 0.015093 ( 3e-06 ) | -54.004984 ( 2.186929 ) | -0.291936 ( 2.230754 ) | 0.004455 ( 6.3e-05 ) | 1 ( 0 ) |
| 0.2 | True model | -3e-05 ( 0.000318 ) | 0.944 ( 0.007271 ) | 0.010041 ( 0.000225 ) | 0.00976 ( 1e-06 ) | 0 ( 0 ) | -2.79585 ( 2.174653 ) | 0.000101 ( 5e-06 ) | 1 ( 0 ) |
| 0.2 | CCA | 0.199097 ( 0.000461 ) | 0 ( 0 ) | 0.014572 ( 0.000326 ) | 0.014639 ( 2e-06 ) | -52.519958 ( 2.170385 ) | 0.460822 ( 2.247539 ) | 0.039852 ( 0.000184 ) | 1 ( 0 ) |
| 0.2 | ipw logadd i | 0.000497 ( 0.000417 ) | 0.981 ( 0.004317 ) | 0.013192 ( 0.000295 ) | 0.01543 ( 4e-06 ) | -42.068768 ( 2.361216 ) | 16.965481 ( 2.616909 ) | 0.000174 ( 8e-06 ) | 1 ( 0 ) |
| 0.2 | ipw logadd noi | 0.199396 ( 0.000469 ) | 0 ( 0 ) | 0.014836 ( 0.000332 ) | 0.014885 ( 3e-06 ) | -54.197392 ( 2.128241 ) | 0.327765 ( 2.244603 ) | 0.039979 ( 0.000187 ) | 1 ( 0 ) |
| 0.2 | ipw logit i | -0.019946 ( 0.000424 ) | 0.808 ( 0.012455 ) | 0.013421 ( 3e-04 ) | 0.016146 ( 6e-06 ) | -44.025246 ( 2.345552 ) | 20.308755 ( 2.69189 ) | 0.000578 ( 1.9e-05 ) | 1 ( 0 ) |
| 0.2 | ipw logit noi | 0.165091 ( 0.000477 ) | 0 ( 0 ) | 0.015086 ( 0.000338 ) | 0.015156 ( 3e-06 ) | -55.701342 ( 2.044241 ) | 0.45991 ( 2.247575 ) | 0.027482 ( 0.000158 ) | 1 ( 0 ) |
| 0.3 | True model | -0.00014 ( 0.00032 ) | 0.944 ( 0.007271 ) | 0.010127 ( 0.000227 ) | 0.009758 ( 1e-06 ) | 0 ( 0 ) | -3.645512 ( 2.155644 ) | 0.000102 ( 5e-06 ) | 1 ( 0 ) |
| 0.3 | CCA | 0.290365 ( 0.000471 ) | 0 ( 0 ) | 0.014894 ( 0.000333 ) | 0.014509 ( 2e-06 ) | -53.767294 ( 2.096448 ) | -2.586906 ( 2.179354 ) | 0.084534 ( 0.000273 ) | 1 ( 0 ) |
| 0.3 | ipw logadd i | 0.005824 ( 0.000441 ) | 0.956 ( 0.006486 ) | 0.013958 ( 0.000312 ) | 0.015838 ( 5e-06 ) | -47.356244 ( 2.202566 ) | 13.471656 ( 2.538842 ) | 0.000229 ( 1e-05 ) | 1 ( 0 ) |
| 0.3 | ipw logadd noi | 0.293635 ( 0.000482 ) | 0 ( 0 ) | 0.01524 ( 0.000341 ) | 0.014825 ( 3e-06 ) | -55.840462 ( 2.031134 ) | -2.719741 ( 2.176428 ) | 0.086453 ( 0.000283 ) | 1 ( 0 ) |
| 0.3 | ipw logit i | -0.049405 ( 0.000474 ) | 0.16 ( 0.011593 ) | 0.014982 ( 0.000335 ) | 0.017778 ( 1.4e-05 ) | -54.308301 ( 2.07266 ) | 18.660102 ( 2.656323 ) | 0.002665 ( 4.7e-05 ) | 1 ( 0 ) |
| 0.3 | ipw logit noi | 0.26244 ( 0.000492 ) | 0 ( 0 ) | 0.015574 ( 0.000348 ) | 0.015184 ( 3e-06 ) | -57.714071 ( 1.927252 ) | -2.499745 ( 2.181373 ) | 0.069117 ( 0.000258 ) | 1 ( 0 ) |

###### Vary selection probability

###### Delta3=0

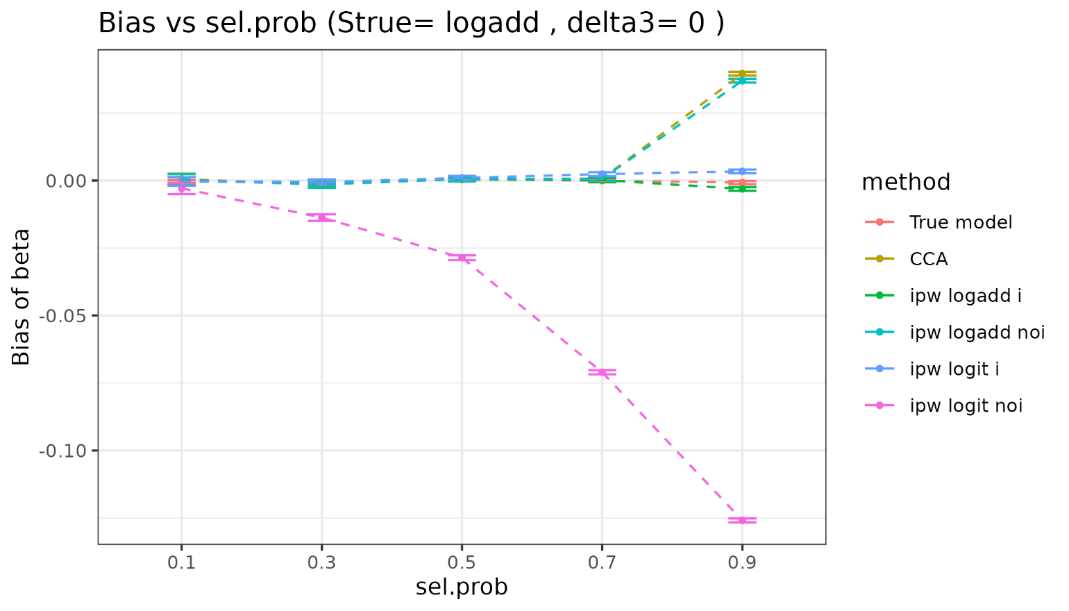

| sel.prob | method | bias | coverage | EmpSE | ModSE | relative_precision | relative_error_ModSE | MSE | power |
| --- | --- | --- | --- | --- | --- | --- | --- | --- | --- |
| 0.1 | True model | -0.000457 ( 0.000307 ) | 0.949 ( 0.006957 ) | 0.009715 ( 0.000217 ) | 0.00976 ( 1e-06 ) | 0 ( 0 ) | 0.462243 ( 2.247543 ) | 9.4e-05 ( 4e-06 ) | 1 ( 0 ) |
| 0.1 | CCA | 0.000476 ( 0.001075 ) | 0.94 ( 0.00751 ) | 0.034002 ( 0.000761 ) | 0.033076 ( 1.1e-05 ) | -91.836927 ( 0.496415 ) | -2.723639 ( 2.176487 ) | 0.001155 ( 5.2e-05 ) | 1 ( 0 ) |
| 0.1 | ipw logadd i | -0.000327 ( 0.000796 ) | 0.993 ( 0.002636 ) | 0.025178 ( 0.000563 ) | 0.033346 ( 1.5e-05 ) | -85.112654 ( 0.871243 ) | 32.441516 ( 2.963554 ) | 0.000633 ( 2.9e-05 ) | 1 ( 0 ) |
| 0.1 | ipw logadd noi | 0.000239 ( 0.001083 ) | 0.944 ( 0.007271 ) | 0.034239 ( 0.000766 ) | 0.033338 ( 1.5e-05 ) | -91.949346 ( 0.490859 ) | -2.629275 ( 2.178788 ) | 0.001171 ( 5.3e-05 ) | 1 ( 0 ) |
| 0.1 | ipw logit i | -0.000301 ( 0.000796 ) | 0.993 ( 0.002636 ) | 0.025179 ( 0.000563 ) | 0.033364 ( 1.5e-05 ) | -85.113398 ( 0.871221 ) | 32.507688 ( 2.965038 ) | 0.000633 ( 2.9e-05 ) | 1 ( 0 ) |
| 0.1 | ipw logit noi | -0.002901 ( 0.001083 ) | 0.941 ( 0.007451 ) | 0.034243 ( 0.000766 ) | 0.033364 ( 1.5e-05 ) | -91.951441 ( 0.490804 ) | -2.566411 ( 2.180198 ) | 0.00118 ( 5.3e-05 ) | 1 ( 0 ) |
| 0.3 | True model | -0.000449 ( 0.000308 ) | 0.951 ( 0.006826 ) | 0.009733 ( 0.000218 ) | 0.00976 ( 1e-06 ) | 0 ( 0 ) | 0.280413 ( 2.243476 ) | 9.5e-05 ( 4e-06 ) | 1 ( 0 ) |
| 0.3 | CCA | -0.001322 ( 0.000609 ) | 0.944 ( 0.007271 ) | 0.019252 ( 0.000431 ) | 0.019092 ( 4e-06 ) | -74.443771 ( 1.392762 ) | -0.831414 ( 2.218667 ) | 0.000372 ( 1.7e-05 ) | 1 ( 0 ) |
| 0.3 | ipw logadd i | -0.000743 ( 0.000507 ) | 0.981 ( 0.004317 ) | 0.016046 ( 0.000359 ) | 0.019242 ( 5e-06 ) | -63.211893 ( 1.801114 ) | 19.914835 ( 2.682889 ) | 0.000258 ( 1.2e-05 ) | 1 ( 0 ) |
| 0.3 | ipw logadd noi | -0.001498 ( 0.000613 ) | 0.947 ( 0.007085 ) | 0.0194 ( 0.000434 ) | 0.019242 ( 5e-06 ) | -74.832357 ( 1.377163 ) | -0.815583 ( 2.21908 ) | 0.000378 ( 1.7e-05 ) | 1 ( 0 ) |
| 0.3 | ipw logit i | -0.000574 ( 0.000508 ) | 0.981 ( 0.004317 ) | 0.016054 ( 0.000359 ) | 0.019279 ( 5e-06 ) | -63.246876 ( 1.799782 ) | 20.086748 ( 2.686737 ) | 0.000258 ( 1.2e-05 ) | 1 ( 0 ) |
| 0.3 | ipw logit noi | -0.013732 ( 0.000613 ) | 0.903 ( 0.009359 ) | 0.019389 ( 0.000434 ) | 0.019303 ( 5e-06 ) | -74.803624 ( 1.379877 ) | -0.446491 ( 2.227341 ) | 0.000564 ( 2.3e-05 ) | 1 ( 0 ) |
| 0.5 | True model | 0.000368 ( 0.000311 ) | 0.94 ( 0.00751 ) | 0.00983 ( 0.00022 ) | 0.00976 ( 1e-06 ) | 0 ( 0 ) | -0.710937 ( 2.221297 ) | 9.7e-05 ( 4e-06 ) | 1 ( 0 ) |
| 0.5 | CCA | 0.000618 ( 0.000452 ) | 0.961 ( 0.006122 ) | 0.014307 ( 0.00032 ) | 0.014788 ( 2e-06 ) | -52.792797 ( 2.24494 ) | 3.360425 ( 2.312413 ) | 0.000205 ( 9e-06 ) | 1 ( 0 ) |
| 0.5 | ipw logadd i | 0.000443 ( 0.000392 ) | 0.983 ( 0.004088 ) | 0.012385 ( 0.000277 ) | 0.014901 ( 3e-06 ) | -37.003316 ( 2.481647 ) | 20.313679 ( 2.691745 ) | 0.000153 ( 7e-06 ) | 1 ( 0 ) |
| 0.5 | ipw logadd noi | 0.000585 ( 0.000452 ) | 0.963 ( 0.005969 ) | 0.014303 ( 0.00032 ) | 0.0149 ( 3e-06 ) | -52.765217 ( 2.258715 ) | 4.17691 ( 2.33072 ) | 0.000205 ( 9e-06 ) | 1 ( 0 ) |
| 0.5 | ipw logit i | 0.000972 ( 0.000392 ) | 0.982 ( 0.004204 ) | 0.012394 ( 0.000277 ) | 0.014966 ( 3e-06 ) | -37.093069 ( 2.479782 ) | 20.750686 ( 2.701525 ) | 0.000154 ( 7e-06 ) | 1 ( 0 ) |
| 0.5 | ipw logit noi | -0.028586 ( 0.000456 ) | 0.521 ( 0.015797 ) | 0.014418 ( 0.000323 ) | 0.015021 ( 3e-06 ) | -53.513856 ( 2.222087 ) | 4.185792 ( 2.330926 ) | 0.001025 ( 2.7e-05 ) | 1 ( 0 ) |
| 0.7 | True model | -5e-05 ( 0.000319 ) | 0.944 ( 0.007271 ) | 0.010073 ( 0.000225 ) | 0.009759 ( 1e-06 ) | 0 ( 0 ) | -3.115756 ( 2.167496 ) | 0.000101 ( 5e-06 ) | 1 ( 0 ) |
| 0.7 | CCA | 0.000698 ( 0.000403 ) | 0.931 ( 0.008015 ) | 0.012759 ( 0.000285 ) | 0.012495 ( 1e-06 ) | -37.666562 ( 2.425331 ) | -2.064045 ( 2.191041 ) | 0.000163 ( 8e-06 ) | 1 ( 0 ) |
| 0.7 | ipw logadd i | 0.000179 ( 0.000369 ) | 0.966 ( 0.005731 ) | 0.011678 ( 0.000261 ) | 0.01259 ( 2e-06 ) | -25.598701 ( 2.285823 ) | 7.8059 ( 2.411883 ) | 0.000136 ( 6e-06 ) | 1 ( 0 ) |
| 0.7 | ipw logadd noi | 0.00065 ( 0.000404 ) | 0.932 ( 0.007961 ) | 0.012769 ( 0.000286 ) | 0.012589 ( 2e-06 ) | -37.764715 ( 2.455996 ) | -1.406365 ( 2.205781 ) | 0.000163 ( 8e-06 ) | 1 ( 0 ) |
| 0.7 | ipw logit i | 0.002375 ( 0.00037 ) | 0.965 ( 0.005812 ) | 0.011704 ( 0.000262 ) | 0.012715 ( 2e-06 ) | -25.920686 ( 2.279058 ) | 8.64505 ( 2.43066 ) | 0.000142 ( 7e-06 ) | 1 ( 0 ) |
| 0.7 | ipw logit noi | -0.07101 ( 0.000404 ) | 0.001 ( 0.000999 ) | 0.01277 ( 0.000286 ) | 0.012892 ( 2e-06 ) | -37.77522 ( 2.453217 ) | 0.959195 ( 2.258721 ) | 0.005205 ( 5.7e-05 ) | 1 ( 0 ) |
| 0.9 | True model | -0.000744 ( 0.000325 ) | 0.943 ( 0.007332 ) | 0.010266 ( 0.00023 ) | 0.009759 ( 1e-06 ) | 0 ( 0 ) | -4.936156 ( 2.126772 ) | 0.000106 ( 4e-06 ) | 1 ( 0 ) |
| 0.9 | CCA | 0.039579 ( 0.000361 ) | 0.058 ( 0.007392 ) | 0.011403 ( 0.000255 ) | 0.010983 ( 1e-06 ) | -18.938445 ( 2.388412 ) | -3.680819 ( 2.154865 ) | 0.001696 ( 2.9e-05 ) | 1 ( 0 ) |
| 0.9 | ipw logadd i | -0.00312 ( 0.000349 ) | 0.952 ( 0.00676 ) | 0.011028 ( 0.000247 ) | 0.011108 ( 2e-06 ) | -13.333137 ( 1.886032 ) | 0.724208 ( 2.253442 ) | 0.000131 ( 6e-06 ) | 1 ( 0 ) |
| 0.9 | ipw logadd noi | 0.036973 ( 0.000362 ) | 0.098 ( 0.009402 ) | 0.011455 ( 0.000256 ) | 0.011077 ( 2e-06 ) | -19.679369 ( 2.388006 ) | -3.296334 ( 2.16349 ) | 0.001498 ( 2.7e-05 ) | 1 ( 0 ) |
| 0.9 | ipw logit i | 0.003397 ( 0.00035 ) | 0.948 ( 0.007021 ) | 0.011074 ( 0.000248 ) | 0.011394 ( 2e-06 ) | -14.049927 ( 1.900844 ) | 2.892589 ( 2.301965 ) | 0.000134 ( 6e-06 ) | 1 ( 0 ) |
| 0.9 | ipw logit noi | -0.125883 ( 0.000372 ) | 0 ( 0 ) | 0.011759 ( 0.000263 ) | 0.01189 ( 3e-06 ) | -23.778822 ( 2.219337 ) | 1.110139 ( 2.262172 ) | 0.015985 ( 9.4e-05 ) | 1 ( 0 ) |

###### Delta3=0.1

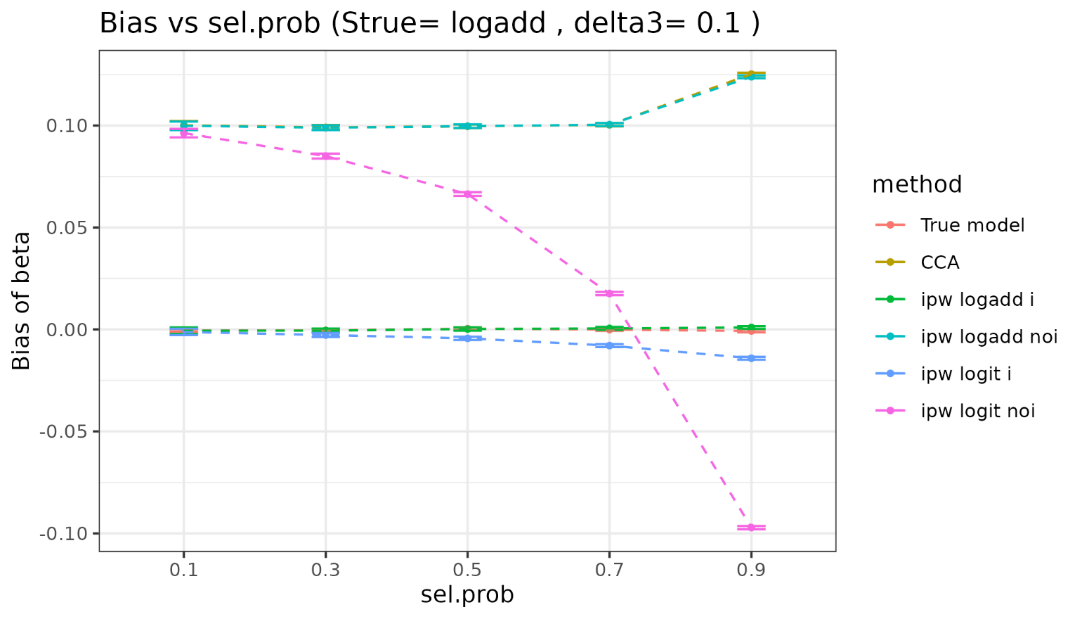

| sel.prob | method | bias | coverage | EmpSE | ModSE | relative_precision | relative_error_ModSE | MSE | power |
| --- | --- | --- | --- | --- | --- | --- | --- | --- | --- |
| 0.1 | True model | -0.000457 ( 0.000307 ) | 0.949 ( 0.006957 ) | 0.009715 ( 0.000217 ) | 0.00976 ( 1e-06 ) | 0 ( 0 ) | 0.462243 ( 2.247543 ) | 9.4e-05 ( 4e-06 ) | 1 ( 0 ) |
| 0.1 | CCA | 0.100119 ( 0.001059 ) | 0.139 ( 0.01094 ) | 0.033478 ( 0.000749 ) | 0.032949 ( 1.1e-05 ) | -91.579285 ( 0.508944 ) | -1.579512 ( 2.202083 ) | 0.011144 ( 0.000218 ) | 1 ( 0 ) |
| 0.1 | ipw logadd i | -0.000601 ( 0.000808 ) | 0.995 ( 0.00223 ) | 0.025549 ( 0.000572 ) | 0.033797 ( 1.7e-05 ) | -85.541629 ( 0.853136 ) | 32.283104 ( 2.960164 ) | 0.000652 ( 3.1e-05 ) | 1 ( 0 ) |
| 0.1 | ipw logadd noi | 0.099852 ( 0.001075 ) | 0.154 ( 0.011414 ) | 0.033984 ( 0.00076 ) | 0.033351 ( 1.5e-05 ) | -91.828397 ( 0.495619 ) | -1.863815 ( 2.195923 ) | 0.011124 ( 0.00022 ) | 1 ( 0 ) |
| 0.1 | ipw logit i | -0.001225 ( 0.000808 ) | 0.993 ( 0.002636 ) | 0.025555 ( 0.000572 ) | 0.033858 ( 1.7e-05 ) | -85.548547 ( 0.852877 ) | 32.490663 ( 2.964826 ) | 0.000654 ( 3.1e-05 ) | 1 ( 0 ) |
| 0.1 | ipw logit noi | 0.096281 ( 0.001075 ) | 0.181 ( 0.012175 ) | 0.034007 ( 0.000761 ) | 0.033391 ( 1.5e-05 ) | -91.839327 ( 0.49508 ) | -1.812376 ( 2.197079 ) | 0.010425 ( 0.000212 ) | 1 ( 0 ) |
| 0.3 | True model | -0.000449 ( 0.000308 ) | 0.951 ( 0.006826 ) | 0.009733 ( 0.000218 ) | 0.00976 ( 1e-06 ) | 0 ( 0 ) | 0.280413 ( 2.243476 ) | 9.5e-05 ( 4e-06 ) | 1 ( 0 ) |
| 0.3 | CCA | 0.0991 ( 0.000603 ) | 0.002 ( 0.001413 ) | 0.019059 ( 0.000426 ) | 0.019019 ( 4e-06 ) | -73.923099 ( 1.424618 ) | -0.213561 ( 2.232489 ) | 0.010184 ( 0.000122 ) | 1 ( 0 ) |
| 0.3 | ipw logadd i | -0.000549 ( 0.000509 ) | 0.981 ( 0.004317 ) | 0.016082 ( 0.00036 ) | 0.019497 ( 6e-06 ) | -63.371928 ( 1.816966 ) | 21.236559 ( 2.712508 ) | 0.000259 ( 1.2e-05 ) | 1 ( 0 ) |
| 0.3 | ipw logadd noi | 0.098939 ( 0.000609 ) | 0.002 ( 0.001413 ) | 0.019264 ( 0.000431 ) | 0.019245 ( 5e-06 ) | -74.474793 ( 1.402657 ) | -0.096782 ( 2.235166 ) | 0.01016 ( 0.000124 ) | 1 ( 0 ) |
| 0.3 | ipw logit i | -0.00273 ( 0.00051 ) | 0.981 ( 0.004317 ) | 0.016114 ( 0.000361 ) | 0.019629 ( 6e-06 ) | -63.519665 ( 1.812463 ) | 21.81056 ( 2.725369 ) | 0.000267 ( 1.2e-05 ) | 1 ( 0 ) |
| 0.3 | ipw logit noi | 0.085001 ( 0.00061 ) | 0.005 ( 0.00223 ) | 0.019297 ( 0.000432 ) | 0.019339 ( 5e-06 ) | -74.561933 ( 1.399358 ) | 0.217635 ( 2.242207 ) | 0.007597 ( 0.000108 ) | 1 ( 0 ) |
| 0.5 | True model | 0.000368 ( 0.000311 ) | 0.94 ( 0.00751 ) | 0.00983 ( 0.00022 ) | 0.00976 ( 1e-06 ) | 0 ( 0 ) | -0.710937 ( 2.221297 ) | 9.7e-05 ( 4e-06 ) | 1 ( 0 ) |
| 0.5 | CCA | 0.099627 ( 0.00046 ) | 0 ( 0 ) | 0.014531 ( 0.000325 ) | 0.014731 ( 2e-06 ) | -54.234234 ( 2.166068 ) | 1.378371 ( 2.268066 ) | 0.010136 ( 9.2e-05 ) | 1 ( 0 ) |
| 0.5 | ipw logadd i | 0.000196 ( 0.000399 ) | 0.98 ( 0.004427 ) | 0.012623 ( 0.000282 ) | 0.015096 ( 3e-06 ) | -39.351948 ( 2.435509 ) | 19.595736 ( 2.675702 ) | 0.000159 ( 8e-06 ) | 1 ( 0 ) |
| 0.5 | ipw logadd noi | 0.099661 ( 0.000461 ) | 0 ( 0 ) | 0.014581 ( 0.000326 ) | 0.014903 ( 3e-06 ) | -54.548065 ( 2.177905 ) | 2.20788 ( 2.286662 ) | 0.010145 ( 9.3e-05 ) | 1 ( 0 ) |
| 0.5 | ipw logit i | -0.004355 ( 0.000401 ) | 0.973 ( 0.005126 ) | 0.012671 ( 0.000283 ) | 0.015328 ( 4e-06 ) | -39.816279 ( 2.423371 ) | 20.966229 ( 2.70639 ) | 0.000179 ( 8e-06 ) | 1 ( 0 ) |
| 0.5 | ipw logit noi | 0.066373 ( 0.000468 ) | 0.004 ( 0.001996 ) | 0.014784 ( 0.000331 ) | 0.015091 ( 3e-06 ) | -55.787852 ( 2.118679 ) | 2.074308 ( 2.283685 ) | 0.004624 ( 6.3e-05 ) | 1 ( 0 ) |
| 0.7 | True model | -5e-05 ( 0.000319 ) | 0.944 ( 0.007271 ) | 0.010073 ( 0.000225 ) | 0.009759 ( 1e-06 ) | 0 ( 0 ) | -3.115756 ( 2.167496 ) | 0.000101 ( 5e-06 ) | 1 ( 0 ) |
| 0.7 | CCA | 0.100338 ( 0.000396 ) | 0 ( 0 ) | 0.012529 ( 0.00028 ) | 0.012449 ( 1e-06 ) | -35.35646 ( 2.483703 ) | -0.640149 ( 2.222894 ) | 0.010225 ( 8e-05 ) | 1 ( 0 ) |
| 0.7 | ipw logadd i | 0.000538 ( 0.000366 ) | 0.972 ( 0.005217 ) | 0.011582 ( 0.000259 ) | 0.012758 ( 2e-06 ) | -24.353766 ( 2.389707 ) | 10.153533 ( 2.464419 ) | 0.000134 ( 6e-06 ) | 1 ( 0 ) |
| 0.7 | ipw logadd noi | 0.10043 ( 0.000398 ) | 0 ( 0 ) | 0.012582 ( 0.000281 ) | 0.012593 ( 2e-06 ) | -35.898706 ( 2.529352 ) | 0.091848 ( 2.239298 ) | 0.010244 ( 8e-05 ) | 1 ( 0 ) |
| 0.7 | ipw logit i | -0.007913 ( 0.000369 ) | 0.932 ( 0.007961 ) | 0.011664 ( 0.000261 ) | 0.013196 ( 3e-06 ) | -25.416231 ( 2.395131 ) | 13.133165 ( 2.531126 ) | 0.000199 ( 8e-06 ) | 1 ( 0 ) |
| 0.7 | ipw logit noi | 0.01762 ( 0.000408 ) | 0.723 ( 0.014152 ) | 0.012891 ( 0.000288 ) | 0.013078 ( 3e-06 ) | -38.935315 ( 2.430699 ) | 1.453064 ( 2.269785 ) | 0.000476 ( 1.6e-05 ) | 1 ( 0 ) |
| 0.9 | True model | -0.000744 ( 0.000325 ) | 0.943 ( 0.007332 ) | 0.010266 ( 0.00023 ) | 0.009759 ( 1e-06 ) | 0 ( 0 ) | -4.936156 ( 2.126772 ) | 0.000106 ( 4e-06 ) | 1 ( 0 ) |
| 0.9 | CCA | 0.125204 ( 0.000359 ) | 0 ( 0 ) | 0.011359 ( 0.000254 ) | 0.010948 ( 1e-06 ) | -18.316932 ( 2.447317 ) | -3.617093 ( 2.156288 ) | 0.015805 ( 9e-05 ) | 1 ( 0 ) |
| 0.9 | ipw logadd i | 0.000954 ( 0.00035 ) | 0.959 ( 0.00627 ) | 0.011055 ( 0.000247 ) | 0.011215 ( 2e-06 ) | -13.765229 ( 2.038439 ) | 1.441523 ( 2.269496 ) | 0.000123 ( 5e-06 ) | 1 ( 0 ) |
| 0.9 | ipw logadd noi | 0.123899 ( 0.000362 ) | 0 ( 0 ) | 0.011451 ( 0.000256 ) | 0.01105 ( 2e-06 ) | -19.619642 ( 2.452008 ) | -3.501363 ( 2.1589 ) | 0.015482 ( 9e-05 ) | 1 ( 0 ) |
| 0.9 | ipw logit i | -0.014116 ( 0.000358 ) | 0.79 ( 0.01288 ) | 0.01131 ( 0.000253 ) | 0.012112 ( 4e-06 ) | -17.60103 ( 2.118895 ) | 7.092044 ( 2.396118 ) | 0.000327 ( 1.1e-05 ) | 1 ( 0 ) |
| 0.9 | ipw logit noi | -0.097163 ( 0.000402 ) | 0 ( 0 ) | 0.012697 ( 0.000284 ) | 0.012984 ( 1.6e-05 ) | -34.623324 ( 2.139352 ) | 2.264307 ( 2.291397 ) | 0.009602 ( 7.9e-05 ) | 1 ( 0 ) |

##### 3.2.2 Y is binary

###### Vary delta3

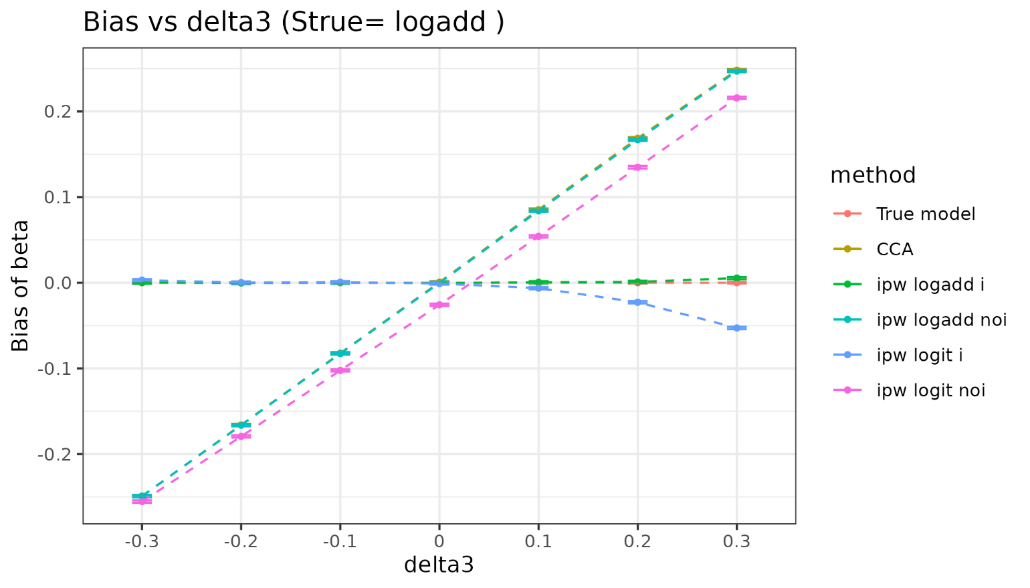

| delta3 | method | bias | coverage | EmpSE | ModSE | relative_precision | relative_error_ModSE | MSE | power |
| --- | --- | --- | --- | --- | --- | --- | --- | --- | --- |
| -0.3 | True model | 0 ( 0.00045 ) | 0.952 ( 0.00676 ) | 0.014234 ( 0.000318 ) | 0.014249 ( 1e-06 ) | 0 ( 0 ) | 0.111037 ( 2.239679 ) | 0.000202 ( 9e-06 ) | 1 ( 0 ) |
| -0.3 | CCA | -0.249059 ( 0.000681 ) | 0 ( 0 ) | 0.021523 ( 0.000482 ) | 0.021192 ( 2e-06 ) | -56.265916 ( 2.029729 ) | -1.537239 ( 2.202823 ) | 0.062493 ( 0.00034 ) | 1 ( 0 ) |
| -0.3 | ipw logadd i | 0.000135 ( 0.000657 ) | 0.96 ( 0.006197 ) | 0.020766 ( 0.000465 ) | 0.021486 ( 2e-06 ) | -53.018848 ( 2.093767 ) | 3.466622 ( 2.314773 ) | 0.000431 ( 2e-05 ) | 1 ( 0 ) |
| -0.3 | ipw logadd noi | -0.249139 ( 0.000681 ) | 0 ( 0 ) | 0.021541 ( 0.000482 ) | 0.021199 ( 2e-06 ) | -56.337901 ( 2.027996 ) | -1.587904 ( 2.20169 ) | 0.062534 ( 0.00034 ) | 1 ( 0 ) |
| -0.3 | ipw logit i | 0.003016 ( 0.000654 ) | 0.959 ( 0.00627 ) | 0.020685 ( 0.000463 ) | 0.021498 ( 2e-06 ) | -52.651269 ( 2.111201 ) | 3.928893 ( 2.325115 ) | 0.000437 ( 2e-05 ) | 1 ( 0 ) |
| -0.3 | ipw logit noi | -0.255451 ( 0.000681 ) | 0 ( 0 ) | 0.021544 ( 0.000482 ) | 0.021201 ( 2e-06 ) | -56.348665 ( 2.030101 ) | -1.589948 ( 2.201645 ) | 0.065719 ( 0.000349 ) | 1 ( 0 ) |
| -0.2 | True model | 0 ( 0.000443 ) | 0.954 ( 0.006624 ) | 0.014018 ( 0.000314 ) | 0.014249 ( 1e-06 ) | 0 ( 0 ) | 1.650235 ( 2.274113 ) | 0.000196 ( 9e-06 ) | 1 ( 0 ) |
| -0.2 | CCA | -0.166019 ( 0.000681 ) | 0 ( 0 ) | 0.021544 ( 0.000482 ) | 0.021433 ( 3e-06 ) | -57.664615 ( 2.001152 ) | -0.517208 ( 2.225646 ) | 0.028026 ( 0.000226 ) | 1 ( 0 ) |
| -0.2 | ipw logadd i | -0.000323 ( 0.000659 ) | 0.962 ( 0.006046 ) | 0.020841 ( 0.000466 ) | 0.021522 ( 3e-06 ) | -54.759875 ( 2.042381 ) | 3.267293 ( 2.310314 ) | 0.000434 ( 2e-05 ) | 1 ( 0 ) |
| -0.2 | ipw logadd noi | -0.166274 ( 0.000683 ) | 0 ( 0 ) | 0.021599 ( 0.000483 ) | 0.021457 ( 3e-06 ) | -57.876464 ( 1.993329 ) | -0.654171 ( 2.222583 ) | 0.028113 ( 0.000227 ) | 1 ( 0 ) |
| -0.2 | ipw logit i | 0.000214 ( 0.000658 ) | 0.96 ( 0.006197 ) | 0.020818 ( 0.000466 ) | 0.021522 ( 3e-06 ) | -54.658348 ( 2.046574 ) | 3.383814 ( 2.31292 ) | 0.000433 ( 2e-05 ) | 1 ( 0 ) |
| -0.2 | ipw logit noi | -0.179393 ( 0.000681 ) | 0 ( 0 ) | 0.021534 ( 0.000482 ) | 0.021466 ( 3e-06 ) | -57.624567 ( 2.00572 ) | -0.314581 ( 2.23018 ) | 0.032645 ( 0.000244 ) | 1 ( 0 ) |
| -0.1 | True model | 0 ( 0.000453 ) | 0.947 ( 0.007085 ) | 0.014323 ( 0.00032 ) | 0.01425 ( 1e-06 ) | 0 ( 0 ) | -0.506395 ( 2.225866 ) | 0.000205 ( 9e-06 ) | 1 ( 0 ) |
| -0.1 | CCA | -0.082196 ( 0.000675 ) | 0.028 ( 0.005217 ) | 0.021347 ( 0.000478 ) | 0.021646 ( 3e-06 ) | -54.981398 ( 2.120465 ) | 1.400163 ( 2.268546 ) | 0.007211 ( 0.000113 ) | 1 ( 0 ) |
| -0.1 | ipw logadd i | 0.000124 ( 0.000645 ) | 0.968 ( 0.005566 ) | 0.020387 ( 0.000456 ) | 0.021667 ( 3e-06 ) | -50.643772 ( 2.206762 ) | 6.274862 ( 2.377604 ) | 0.000415 ( 1.9e-05 ) | 1 ( 0 ) |
| -0.1 | ipw logadd noi | -0.082653 ( 0.000675 ) | 0.027 ( 0.005126 ) | 0.021341 ( 0.000477 ) | 0.021698 ( 3e-06 ) | -54.955282 ( 2.118953 ) | 1.673844 ( 2.27467 ) | 0.007286 ( 0.000113 ) | 1 ( 0 ) |
| -0.1 | ipw logit i | 0.000668 ( 0.000644 ) | 0.968 ( 0.005566 ) | 0.02038 ( 0.000456 ) | 0.021667 ( 3e-06 ) | -50.608958 ( 2.208541 ) | 6.314878 ( 2.378499 ) | 0.000415 ( 2e-05 ) | 1 ( 0 ) |
| -0.1 | ipw logit noi | -0.102327 ( 0.000672 ) | 0.005 ( 0.00223 ) | 0.021254 ( 0.000475 ) | 0.02172 ( 3e-06 ) | -54.584899 ( 2.132764 ) | 2.193125 ( 2.286288 ) | 0.010922 ( 0.000139 ) | 1 ( 0 ) |
| 0 | True model | 0 ( 0.000442 ) | 0.953 ( 0.006693 ) | 0.013972 ( 0.000313 ) | 0.01425 ( 1e-06 ) | 0 ( 0 ) | 1.993794 ( 2.2818 ) | 0.000195 ( 9e-06 ) | 1 ( 0 ) |
| 0 | CCA | 0.000582 ( 0.000677 ) | 0.952 ( 0.00676 ) | 0.021419 ( 0.000479 ) | 0.021831 ( 3e-06 ) | -57.451337 ( 2.105138 ) | 1.923825 ( 2.280267 ) | 0.000459 ( 2.2e-05 ) | 1 ( 0 ) |
| 0 | ipw logadd i | -0.000307 ( 0.000648 ) | 0.958 ( 0.006343 ) | 0.020488 ( 0.000458 ) | 0.021923 ( 3e-06 ) | -53.497614 ( 2.190528 ) | 7.001937 ( 2.393878 ) | 0.000419 ( 2e-05 ) | 1 ( 0 ) |
| 0 | ipw logadd noi | -0.000202 ( 0.00068 ) | 0.947 ( 0.007085 ) | 0.0215 ( 0.000481 ) | 0.021923 ( 3e-06 ) | -57.771442 ( 2.093217 ) | 1.964387 ( 2.281175 ) | 0.000462 ( 2.2e-05 ) | 1 ( 0 ) |
| 0 | ipw logit i | -0.000941 ( 0.000647 ) | 0.961 ( 0.006122 ) | 0.020469 ( 0.000458 ) | 0.021928 ( 3e-06 ) | -53.407763 ( 2.193945 ) | 7.127359 ( 2.396685 ) | 0.000419 ( 2e-05 ) | 1 ( 0 ) |
| 0 | ipw logit noi | -0.025733 ( 0.000679 ) | 0.794 ( 0.012789 ) | 0.021468 ( 0.00048 ) | 0.021963 ( 3e-06 ) | -57.644768 ( 2.09826 ) | 2.303574 ( 2.288765 ) | 0.001123 ( 3.9e-05 ) | 1 ( 0 ) |
| 0.1 | True model | 0 ( 0.000454 ) | 0.952 ( 0.00676 ) | 0.014347 ( 0.000321 ) | 0.014249 ( 1e-06 ) | 0 ( 0 ) | -0.688115 ( 2.221801 ) | 0.000206 ( 9e-06 ) | 1 ( 0 ) |
| 0.1 | CCA | 0.085279 ( 0.000718 ) | 0.033 ( 0.005649 ) | 0.022693 ( 0.000508 ) | 0.021984 ( 3e-06 ) | -60.028244 ( 1.83668 ) | -3.12335 ( 2.167352 ) | 0.007787 ( 0.000123 ) | 1 ( 0 ) |
| 0.1 | ipw logadd i | 0.000566 ( 0.000679 ) | 0.958 ( 0.006343 ) | 0.021464 ( 0.00048 ) | 0.022294 ( 3e-06 ) | -55.319998 ( 1.98736 ) | 3.863445 ( 2.32367 ) | 0.000461 ( 2.1e-05 ) | 1 ( 0 ) |
| 0.1 | ipw logadd noi | 0.084037 ( 0.000717 ) | 0.038 ( 0.006046 ) | 0.02268 ( 0.000507 ) | 0.022127 ( 3e-06 ) | -59.981798 ( 1.842381 ) | -2.439712 ( 2.182648 ) | 0.007576 ( 0.000121 ) | 1 ( 0 ) |
| 0.1 | ipw logit i | -0.006303 ( 0.000676 ) | 0.944 ( 0.007271 ) | 0.021387 ( 0.000478 ) | 0.022345 ( 3e-06 ) | -54.996916 ( 2.006915 ) | 4.47925 ( 2.337451 ) | 0.000497 ( 2.3e-05 ) | 1 ( 0 ) |
| 0.1 | ipw logit noi | 0.054114 ( 0.000717 ) | 0.309 ( 0.014612 ) | 0.022682 ( 0.000507 ) | 0.02219 ( 3e-06 ) | -59.990091 ( 1.848275 ) | -2.169051 ( 2.188705 ) | 0.003442 ( 8e-05 ) | 1 ( 0 ) |
| 0.2 | True model | 0 ( 0.000448 ) | 0.951 ( 0.006826 ) | 0.014169 ( 0.000317 ) | 0.01425 ( 1e-06 ) | 0 ( 0 ) | 0.571514 ( 2.249981 ) | 0.000201 ( 9e-06 ) | 1 ( 0 ) |
| 0.2 | CCA | 0.168713 ( 0.000715 ) | 0 ( 0 ) | 0.02262 ( 0.000506 ) | 0.022109 ( 3e-06 ) | -60.766139 ( 1.826981 ) | -2.259249 ( 2.186688 ) | 0.028975 ( 0.000241 ) | 1 ( 0 ) |
| 0.2 | ipw logadd i | 0.000995 ( 0.000701 ) | 0.961 ( 0.006122 ) | 0.022167 ( 0.000496 ) | 0.022787 ( 4e-06 ) | -59.146664 ( 1.852595 ) | 2.79641 ( 2.299811 ) | 0.000492 ( 2.2e-05 ) | 1 ( 0 ) |
| 0.2 | ipw logadd noi | 0.166988 ( 0.000725 ) | 0 ( 0 ) | 0.02293 ( 0.000513 ) | 0.022316 ( 3e-06 ) | -61.81776 ( 1.791976 ) | -2.674624 ( 2.177397 ) | 0.02841 ( 0.000242 ) | 1 ( 0 ) |
| 0.2 | ipw logit i | -0.022774 ( 0.000702 ) | 0.84 ( 0.011593 ) | 0.022207 ( 0.000497 ) | 0.023059 ( 4e-06 ) | -59.291365 ( 1.85837 ) | 3.835941 ( 2.323086 ) | 0.001011 ( 4e-05 ) | 1 ( 0 ) |
| 0.2 | ipw logit noi | 0.134873 ( 0.000726 ) | 0 ( 0 ) | 0.022949 ( 0.000513 ) | 0.022409 ( 3e-06 ) | -61.880568 ( 1.792582 ) | -2.350847 ( 2.184643 ) | 0.018717 ( 0.000196 ) | 1 ( 0 ) |
| 0.3 | True model | 0 ( 0.000442 ) | 0.946 ( 0.007147 ) | 0.013972 ( 0.000313 ) | 0.014249 ( 1e-06 ) | 0 ( 0 ) | 1.982761 ( 2.281553 ) | 0.000195 ( 9e-06 ) | 1 ( 0 ) |
| 0.3 | CCA | 0.248117 ( 0.00069 ) | 0 ( 0 ) | 0.021826 ( 0.000488 ) | 0.022221 ( 3e-06 ) | -59.021191 ( 2.003277 ) | 1.805864 ( 2.277633 ) | 0.062038 ( 0.000343 ) | 1 ( 0 ) |
| 0.3 | ipw logadd i | 0.005427 ( 0.000697 ) | 0.956 ( 0.006486 ) | 0.022051 ( 0.000493 ) | 0.023383 ( 4e-06 ) | -59.853022 ( 1.953486 ) | 6.038161 ( 2.372344 ) | 0.000515 ( 2.3e-05 ) | 1 ( 0 ) |
| 0.3 | ipw logadd noi | 0.247104 ( 0.000696 ) | 0 ( 0 ) | 0.022008 ( 0.000492 ) | 0.022501 ( 3e-06 ) | -59.693616 ( 1.985116 ) | 2.241868 ( 2.287389 ) | 0.061544 ( 0.000345 ) | 1 ( 0 ) |
| 0.3 | ipw logit i | -0.052795 ( 0.000718 ) | 0.415 ( 0.015581 ) | 0.0227 ( 0.000508 ) | 0.024354 ( 7e-06 ) | -62.113951 ( 1.894162 ) | 7.288598 ( 2.400427 ) | 0.003302 ( 8e-05 ) | 1 ( 0 ) |
| 0.3 | ipw logit noi | 0.215823 ( 0.000699 ) | 0 ( 0 ) | 0.022111 ( 0.000495 ) | 0.022624 ( 3e-06 ) | -60.067537 ( 1.966415 ) | 2.324326 ( 2.289237 ) | 0.047068 ( 0.000303 ) | 1 ( 0 ) |

###### Vary selection probability

###### Delta3=0

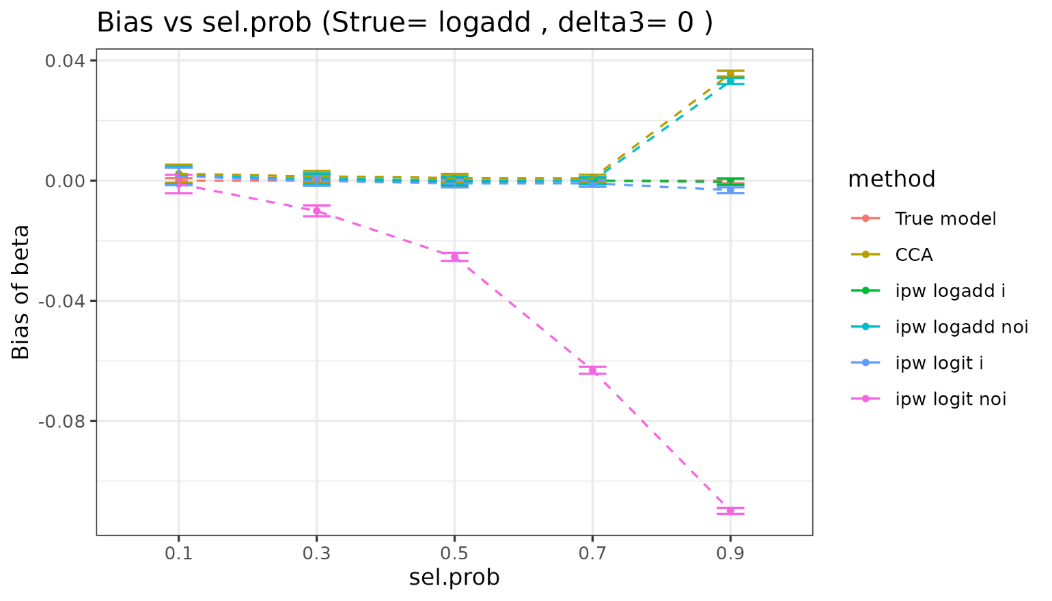

| sel.prob | method | bias | coverage | EmpSE | ModSE | relative_precision | relative_error_ModSE | MSE | power |
| --- | --- | --- | --- | --- | --- | --- | --- | --- | --- |
| 0.1 | True model | 0 ( 0.00045 ) | 0.952 ( 0.00676 ) | 0.014234 ( 0.000318 ) | 0.014249 ( 1e-06 ) | 0 ( 0 ) | 0.111037 ( 2.239679 ) | 0.000202 ( 9e-06 ) | 1 ( 0 ) |
| 0.1 | CCA | 0.002309 ( 0.001543 ) | 0.947 ( 0.007085 ) | 0.048779 ( 0.001091 ) | 0.048799 ( 1.5e-05 ) | -91.485265 ( 0.520991 ) | 0.040825 ( 2.238309 ) | 0.002382 ( 0.000113 ) | 1 ( 0 ) |
| 0.1 | ipw logadd i | 0.001608 ( 0.00144 ) | 0.963 ( 0.005969 ) | 0.045523 ( 0.001018 ) | 0.04901 ( 1.6e-05 ) | -90.223663 ( 0.589055 ) | 7.660831 ( 2.408816 ) | 0.002073 ( 9.3e-05 ) | 1 ( 0 ) |
| 0.1 | ipw logadd noi | 0.001608 ( 0.001552 ) | 0.947 ( 0.007085 ) | 0.049073 ( 0.001098 ) | 0.049004 ( 1.5e-05 ) | -91.587088 ( 0.515057 ) | -0.140364 ( 2.234259 ) | 0.002408 ( 0.000113 ) | 1 ( 0 ) |
| 0.1 | ipw logit i | 0.001507 ( 0.001439 ) | 0.963 ( 0.005969 ) | 0.045518 ( 0.001018 ) | 0.049011 ( 1.6e-05 ) | -90.221621 ( 0.58915 ) | 7.675012 ( 2.409134 ) | 0.002072 ( 9.3e-05 ) | 1 ( 0 ) |
| 0.1 | ipw logit noi | -0.001106 ( 0.001552 ) | 0.951 ( 0.006826 ) | 0.049073 ( 0.001098 ) | 0.049012 ( 1.5e-05 ) | -91.587154 ( 0.515008 ) | -0.123868 ( 2.234629 ) | 0.002407 ( 0.000113 ) | 1 ( 0 ) |
| 0.3 | True model | 0 ( 0.000443 ) | 0.954 ( 0.006624 ) | 0.014018 ( 0.000314 ) | 0.014249 ( 1e-06 ) | 0 ( 0 ) | 1.650235 ( 2.274113 ) | 0.000196 ( 9e-06 ) | 1 ( 0 ) |
| 0.3 | CCA | 0.001418 ( 0.000916 ) | 0.946 ( 0.007147 ) | 0.02897 ( 0.000648 ) | 0.028182 ( 5e-06 ) | -76.585905 ( 1.283686 ) | -2.719038 ( 2.176421 ) | 0.00084 ( 4.2e-05 ) | 1 ( 0 ) |
| 0.3 | ipw logadd i | 0.000286 ( 0.000858 ) | 0.958 ( 0.006343 ) | 0.027138 ( 0.000607 ) | 0.028302 ( 5e-06 ) | -73.318864 ( 1.43343 ) | 4.286906 ( 2.333167 ) | 0.000736 ( 3.5e-05 ) | 1 ( 0 ) |
| 0.3 | ipw logadd noi | 0.000604 ( 0.000922 ) | 0.947 ( 0.007085 ) | 0.029152 ( 0.000652 ) | 0.0283 ( 5e-06 ) | -76.876768 ( 1.268704 ) | -2.919752 ( 2.171932 ) | 0.000849 ( 4.2e-05 ) | 1 ( 0 ) |
| 0.3 | ipw logit i | -4.9e-05 ( 0.000858 ) | 0.959 ( 0.00627 ) | 0.027122 ( 0.000607 ) | 0.028304 ( 5e-06 ) | -73.285863 ( 1.435303 ) | 4.361097 ( 2.334827 ) | 0.000735 ( 3.5e-05 ) | 1 ( 0 ) |
| 0.3 | ipw logit noi | -0.010076 ( 0.000922 ) | 0.928 ( 0.008174 ) | 0.029162 ( 0.000652 ) | 0.02832 ( 5e-06 ) | -76.892922 ( 1.267659 ) | -2.886229 ( 2.172683 ) | 0.000951 ( 4.5e-05 ) | 1 ( 0 ) |
| 0.5 | True model | 0 ( 0.000453 ) | 0.947 ( 0.007085 ) | 0.014323 ( 0.00032 ) | 0.01425 ( 1e-06 ) | 0 ( 0 ) | -0.506395 ( 2.225866 ) | 0.000205 ( 9e-06 ) | 1 ( 0 ) |
| 0.5 | CCA | 0.000922 ( 0.000692 ) | 0.955 ( 0.006556 ) | 0.021895 ( 0.00049 ) | 0.021829 ( 3e-06 ) | -57.208433 ( 2.032592 ) | -0.304133 ( 2.230421 ) | 0.00048 ( 2.3e-05 ) | 1 ( 0 ) |
| 0.5 | ipw logadd i | -0.000266 ( 0.000661 ) | 0.964 ( 0.005891 ) | 0.020891 ( 0.000467 ) | 0.021921 ( 3e-06 ) | -52.993052 ( 2.119381 ) | 4.930908 ( 2.347543 ) | 0.000436 ( 2e-05 ) | 1 ( 0 ) |
| 0.5 | ipw logadd noi | 0.00014 ( 0.000693 ) | 0.953 ( 0.006693 ) | 0.021927 ( 0.000491 ) | 0.02192 ( 3e-06 ) | -57.330784 ( 2.023832 ) | -0.031272 ( 2.236526 ) | 0.00048 ( 2.3e-05 ) | 1 ( 0 ) |
| 0.5 | ipw logit i | -0.000913 ( 0.00066 ) | 0.964 ( 0.005891 ) | 0.020865 ( 0.000467 ) | 0.021925 ( 3e-06 ) | -52.879995 ( 2.123379 ) | 5.078802 ( 2.350852 ) | 0.000436 ( 2e-05 ) | 1 ( 0 ) |
| 0.5 | ipw logit noi | -0.025378 ( 0.000691 ) | 0.801 ( 0.012625 ) | 0.021841 ( 0.000489 ) | 0.02196 ( 3e-06 ) | -56.996165 ( 2.036238 ) | 0.54272 ( 2.249368 ) | 0.001121 ( 4.2e-05 ) | 1 ( 0 ) |
| 0.7 | True model | 0 ( 0.000442 ) | 0.953 ( 0.006693 ) | 0.013972 ( 0.000313 ) | 0.01425 ( 1e-06 ) | 0 ( 0 ) | 1.993794 ( 2.2818 ) | 0.000195 ( 9e-06 ) | 1 ( 0 ) |
| 0.7 | CCA | 0.000839 ( 0.00058 ) | 0.947 ( 0.007085 ) | 0.01833 ( 0.00041 ) | 0.018451 ( 2e-06 ) | -41.900121 ( 2.40153 ) | 0.662569 ( 2.252034 ) | 0.000336 ( 1.5e-05 ) | 1 ( 0 ) |
| 0.7 | ipw logadd i | -3.3e-05 ( 0.000561 ) | 0.957 ( 0.006415 ) | 0.017755 ( 0.000397 ) | 0.018528 ( 2e-06 ) | -38.074718 ( 2.4422 ) | 4.355847 ( 2.334662 ) | 0.000315 ( 1.4e-05 ) | 1 ( 0 ) |
| 0.7 | ipw logadd noi | -1.8e-05 ( 0.000581 ) | 0.951 ( 0.006826 ) | 0.018363 ( 0.000411 ) | 0.018528 ( 2e-06 ) | -42.112694 ( 2.402579 ) | 0.895291 ( 2.257241 ) | 0.000337 ( 1.5e-05 ) | 1 ( 0 ) |
| 0.7 | ipw logit i | -0.000882 ( 0.00056 ) | 0.957 ( 0.006415 ) | 0.01771 ( 0.000396 ) | 0.018537 ( 2e-06 ) | -37.761474 ( 2.457211 ) | 4.669827 ( 2.341687 ) | 0.000314 ( 1.4e-05 ) | 1 ( 0 ) |
| 0.7 | ipw logit noi | -0.063111 ( 0.000581 ) | 0.069 ( 0.008015 ) | 0.018377 ( 0.000411 ) | 0.018634 ( 2e-06 ) | -42.194761 ( 2.402552 ) | 1.399266 ( 2.268518 ) | 0.00432 ( 7.5e-05 ) | 1 ( 0 ) |
| 0.9 | True model | 0 ( 0.000454 ) | 0.952 ( 0.00676 ) | 0.014347 ( 0.000321 ) | 0.014249 ( 1e-06 ) | 0 ( 0 ) | -0.688115 ( 2.221801 ) | 0.000206 ( 9e-06 ) | 1 ( 0 ) |
| 0.9 | CCA | 0.03566 ( 0.000505 ) | 0.415 ( 0.015581 ) | 0.015954 ( 0.000357 ) | 0.01633 ( 2e-06 ) | -19.126811 ( 2.464585 ) | 2.358007 ( 2.289959 ) | 0.001526 ( 3.8e-05 ) | 1 ( 0 ) |
| 0.9 | ipw logadd i | -0.000376 ( 0.000498 ) | 0.958 ( 0.006343 ) | 0.015755 ( 0.000352 ) | 0.016388 ( 2e-06 ) | -17.070878 ( 2.354223 ) | 4.019215 ( 2.327125 ) | 0.000248 ( 1.1e-05 ) | 1 ( 0 ) |
| 0.9 | ipw logadd noi | 0.033155 ( 0.000507 ) | 0.476 ( 0.015793 ) | 0.016025 ( 0.000359 ) | 0.016363 ( 2e-06 ) | -19.84322 ( 2.462632 ) | 2.10726 ( 2.28435 ) | 0.001356 ( 3.6e-05 ) | 1 ( 0 ) |
| 0.9 | ipw logit i | -0.003135 ( 0.000496 ) | 0.952 ( 0.00676 ) | 0.01568 ( 0.000351 ) | 0.016414 ( 2e-06 ) | -16.27138 ( 2.3824 ) | 4.681726 ( 2.341947 ) | 0.000255 ( 1.1e-05 ) | 1 ( 0 ) |
| 0.9 | ipw logit noi | -0.109894 ( 0.000506 ) | 0 ( 0 ) | 0.016006 ( 0.000358 ) | 0.016656 ( 2e-06 ) | -19.650356 ( 2.474851 ) | 4.062242 ( 2.328094 ) | 0.012333 ( 0.000112 ) | 1 ( 0 ) |

###### Delta3=0.1

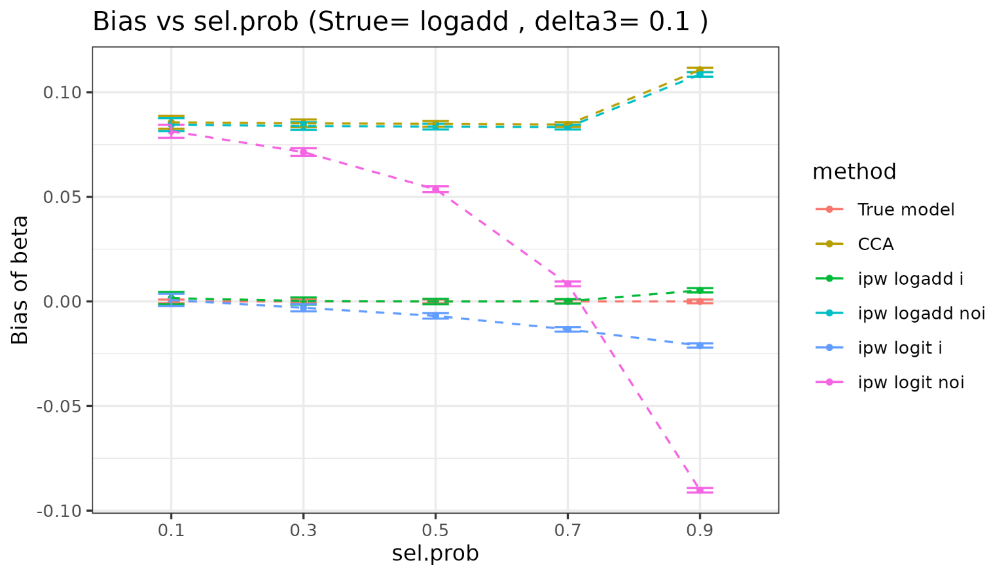

| sel.prob | method | bias | coverage | EmpSE | ModSE | relative_precision | relative_error_ModSE | MSE | power |
| --- | --- | --- | --- | --- | --- | --- | --- | --- | --- |
| 0.1 | True model | 0 ( 0.00045 ) | 0.952 ( 0.00676 ) | 0.014234 ( 0.000318 ) | 0.014249 ( 1e-06 ) | 0 ( 0 ) | 0.111037 ( 2.239679 ) | 0.000202 ( 9e-06 ) | 1 ( 0 ) |
| 0.1 | CCA | 0.085617 ( 0.001562 ) | 0.602 ( 0.015479 ) | 0.049401 ( 0.001105 ) | 0.049145 ( 1.5e-05 ) | -91.698567 ( 0.508286 ) | -0.519791 ( 2.225778 ) | 0.009768 ( 0.000298 ) | 1 ( 0 ) |
| 0.1 | ipw logadd i | 0.001582 ( 0.001478 ) | 0.954 ( 0.006624 ) | 0.046738 ( 0.001046 ) | 0.04984 ( 1.7e-05 ) | -90.725436 ( 0.558899 ) | 6.637164 ( 2.38596 ) | 0.002185 ( 9.8e-05 ) | 1 ( 0 ) |
| 0.1 | ipw logadd noi | 0.084519 ( 0.001579 ) | 0.611 ( 0.015417 ) | 0.049925 ( 0.001117 ) | 0.049464 ( 1.6e-05 ) | -91.871801 ( 0.497759 ) | -0.92299 ( 2.216762 ) | 0.009634 ( 0.000299 ) | 1 ( 0 ) |
| 0.1 | ipw logit i | 0.000692 ( 0.001478 ) | 0.953 ( 0.006693 ) | 0.046733 ( 0.001046 ) | 0.049853 ( 1.7e-05 ) | -90.723538 ( 0.558927 ) | 6.676246 ( 2.386837 ) | 0.002182 ( 9.8e-05 ) | 1 ( 0 ) |
| 0.1 | ipw logit noi | 0.081358 ( 0.001579 ) | 0.63 ( 0.015268 ) | 0.049933 ( 0.001117 ) | 0.049477 ( 1.6e-05 ) | -91.87421 ( 0.497569 ) | -0.9117 ( 2.217016 ) | 0.00911 ( 0.00029 ) | 1 ( 0 ) |
| 0.3 | True model | 0 ( 0.000443 ) | 0.954 ( 0.006624 ) | 0.014018 ( 0.000314 ) | 0.014249 ( 1e-06 ) | 0 ( 0 ) | 1.650235 ( 2.274113 ) | 0.000196 ( 9e-06 ) | 1 ( 0 ) |
| 0.3 | CCA | 0.08517 ( 0.000923 ) | 0.156 ( 0.011474 ) | 0.029193 ( 0.000653 ) | 0.028382 ( 5e-06 ) | -76.942135 ( 1.26701 ) | -2.776813 ( 2.175135 ) | 0.008105 ( 0.000164 ) | 1 ( 0 ) |
| 0.3 | ipw logadd i | 0.000208 ( 0.00088 ) | 0.958 ( 0.006343 ) | 0.027831 ( 0.000623 ) | 0.028783 ( 6e-06 ) | -74.630108 ( 1.371838 ) | 3.421222 ( 2.313815 ) | 0.000774 ( 3.7e-05 ) | 1 ( 0 ) |
| 0.3 | ipw logadd noi | 0.083866 ( 0.000933 ) | 0.16 ( 0.011593 ) | 0.029494 ( 0.00066 ) | 0.028566 ( 5e-06 ) | -77.410184 ( 1.244421 ) | -3.144137 ( 2.166918 ) | 0.007902 ( 0.000164 ) | 1 ( 0 ) |
| 0.3 | ipw logit i | -0.003026 ( 0.00088 ) | 0.96 ( 0.006197 ) | 0.027815 ( 0.000622 ) | 0.028812 ( 6e-06 ) | -74.601134 ( 1.373679 ) | 3.583484 ( 2.317448 ) | 0.000782 ( 3.7e-05 ) | 1 ( 0 ) |
| 0.3 | ipw logit noi | 0.071392 ( 0.000933 ) | 0.296 ( 0.014436 ) | 0.029512 ( 0.00066 ) | 0.028597 ( 5e-06 ) | -77.437921 ( 1.242943 ) | -3.098597 ( 2.167938 ) | 0.005967 ( 0.000141 ) | 1 ( 0 ) |
| 0.5 | True model | 0 ( 0.000453 ) | 0.947 ( 0.007085 ) | 0.014323 ( 0.00032 ) | 0.01425 ( 1e-06 ) | 0 ( 0 ) | -0.506395 ( 2.225866 ) | 0.000205 ( 9e-06 ) | 1 ( 0 ) |
| 0.5 | CCA | 0.084913 ( 0.000704 ) | 0.025 ( 0.004937 ) | 0.022272 ( 0.000498 ) | 0.021985 ( 3e-06 ) | -58.641717 ( 1.925112 ) | -1.285426 ( 2.208471 ) | 0.007706 ( 0.000121 ) | 1 ( 0 ) |
| 0.5 | ipw logadd i | -1.9e-05 ( 0.000669 ) | 0.971 ( 0.005307 ) | 0.021161 ( 0.000473 ) | 0.022294 ( 3e-06 ) | -54.185423 ( 2.0824 ) | 5.357021 ( 2.357087 ) | 0.000447 ( 2.1e-05 ) | 1 ( 0 ) |
| 0.5 | ipw logadd noi | 0.083588 ( 0.000705 ) | 0.031 ( 0.005481 ) | 0.022303 ( 0.000499 ) | 0.022127 ( 3e-06 ) | -58.758988 ( 1.923909 ) | -0.788589 ( 2.219588 ) | 0.007484 ( 0.000119 ) | 1 ( 0 ) |
| 0.5 | ipw logit i | -0.006909 ( 0.000667 ) | 0.95 ( 0.006892 ) | 0.02109 ( 0.000472 ) | 0.022346 ( 3e-06 ) | -53.879768 ( 2.100298 ) | 5.952556 ( 2.370414 ) | 0.000492 ( 2.3e-05 ) | 1 ( 0 ) |
| 0.5 | ipw logit noi | 0.053644 ( 0.000703 ) | 0.319 ( 0.014739 ) | 0.022227 ( 0.000497 ) | 0.022191 ( 3e-06 ) | -58.474164 ( 1.934949 ) | -0.160257 ( 2.233647 ) | 0.003371 ( 7.8e-05 ) | 1 ( 0 ) |
| 0.7 | True model | 0 ( 0.000442 ) | 0.953 ( 0.006693 ) | 0.013972 ( 0.000313 ) | 0.01425 ( 1e-06 ) | 0 ( 0 ) | 1.993794 ( 2.2818 ) | 0.000195 ( 9e-06 ) | 1 ( 0 ) |
| 0.7 | CCA | 0.084562 ( 0.000575 ) | 0.003 ( 0.001729 ) | 0.018173 ( 0.000407 ) | 0.018584 ( 2e-06 ) | -40.891039 ( 2.410464 ) | 2.260701 ( 2.28779 ) | 0.007481 ( 9.9e-05 ) | 1 ( 0 ) |
| 0.7 | ipw logadd i | 7.6e-05 ( 0.000565 ) | 0.966 ( 0.005731 ) | 0.017851 ( 0.000399 ) | 0.018843 ( 2e-06 ) | -38.743705 ( 2.410015 ) | 5.556367 ( 2.361526 ) | 0.000318 ( 1.3e-05 ) | 1 ( 0 ) |
| 0.7 | ipw logadd noi | 0.083333 ( 0.000579 ) | 0.004 ( 0.001996 ) | 0.018319 ( 0.00041 ) | 0.018703 ( 2e-06 ) | -41.833565 ( 2.396318 ) | 2.094364 ( 2.28407 ) | 0.00728 ( 9.8e-05 ) | 1 ( 0 ) |
| 0.7 | ipw logit i | -0.013354 ( 0.000563 ) | 0.907 ( 0.009184 ) | 0.017792 ( 0.000398 ) | 0.018945 ( 2e-06 ) | -38.332395 ( 2.446696 ) | 6.483075 ( 2.382264 ) | 0.000495 ( 2e-05 ) | 1 ( 0 ) |
| 0.7 | ipw logit noi | 0.008392 ( 0.000583 ) | 0.939 ( 0.007568 ) | 0.018423 ( 0.000412 ) | 0.018879 ( 2e-06 ) | -42.488859 ( 2.391214 ) | 2.475071 ( 2.292591 ) | 0.00041 ( 1.8e-05 ) | 1 ( 0 ) |
| 0.9 | True model | 0 ( 0.000454 ) | 0.952 ( 0.00676 ) | 0.014347 ( 0.000321 ) | 0.014249 ( 1e-06 ) | 0 ( 0 ) | -0.688115 ( 2.221801 ) | 0.000206 ( 9e-06 ) | 1 ( 0 ) |
| 0.9 | CCA | 0.110743 ( 0.000529 ) | 0 ( 0 ) | 0.016743 ( 0.000375 ) | 0.016464 ( 2e-06 ) | -26.571097 ( 2.261314 ) | -1.669098 ( 2.199866 ) | 0.012544 ( 0.000118 ) | 1 ( 0 ) |
| 0.9 | ipw logadd i | 0.005326 ( 0.000528 ) | 0.933 ( 0.007906 ) | 0.016692 ( 0.000373 ) | 0.016655 ( 2e-06 ) | -26.117116 ( 2.168921 ) | -0.222586 ( 2.23223 ) | 0.000307 ( 1.3e-05 ) | 1 ( 0 ) |
| 0.9 | ipw logadd noi | 0.108502 ( 0.000532 ) | 0 ( 0 ) | 0.01682 ( 0.000376 ) | 0.016521 ( 2e-06 ) | -27.243414 ( 2.251887 ) | -1.779349 ( 2.1974 ) | 0.012055 ( 0.000116 ) | 1 ( 0 ) |
| 0.9 | ipw logit i | -0.021064 ( 0.000526 ) | 0.761 ( 0.013486 ) | 0.016647 ( 0.000372 ) | 0.01688 ( 2e-06 ) | -25.719818 ( 2.247549 ) | 1.401663 ( 2.268578 ) | 0.000721 ( 2.6e-05 ) | 1 ( 0 ) |
| 0.9 | ipw logit noi | -0.090253 ( 0.000546 ) | 0 ( 0 ) | 0.017264 ( 0.000386 ) | 0.017308 ( 3e-06 ) | -30.934472 ( 2.306058 ) | 0.25583 ( 2.242971 ) | 0.008443 ( 1e-04 ) | 1 ( 0 ) |

#### 3.3 Strue=probit

##### 3.3.1 Y is continuous

###### Vary delta3

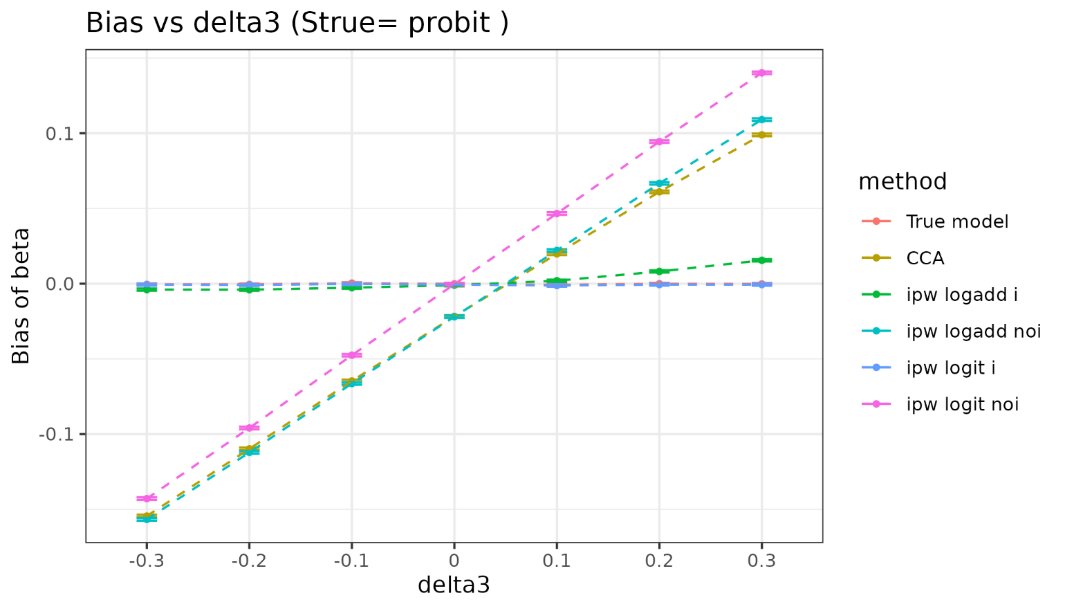

| delta3 | method | bias | coverage | EmpSE | ModSE | relative_precision | relative_error_ModSE | MSE | power |
| --- | --- | --- | --- | --- | --- | --- | --- | --- | --- |
| -0.3 | True model | -0.000457 ( 0.000307 ) | 0.949 ( 0.006957 ) | 0.009715 ( 0.000217 ) | 0.00976 ( 1e-06 ) | 0 ( 0 ) | 0.462243 ( 2.247543 ) | 9.4e-05 ( 4e-06 ) | 1 ( 0 ) |
| -0.3 | CCA | -0.154422 ( 0.000419 ) | 0 ( 0 ) | 0.013236 ( 0.000296 ) | 0.013389 ( 2e-06 ) | -46.128004 ( 2.395646 ) | 1.154522 ( 2.263053 ) | 0.024021 ( 0.00013 ) | 1 ( 0 ) |
| -0.3 | ipw logadd i | -0.003927 ( 0.000373 ) | 0.962 ( 0.006046 ) | 0.01181 ( 0.000264 ) | 0.01349 ( 2e-06 ) | -32.332842 ( 2.511165 ) | 14.227747 ( 2.555545 ) | 0.000155 ( 7e-06 ) | 1 ( 0 ) |
| -0.3 | ipw logadd noi | -0.156694 ( 0.000421 ) | 0 ( 0 ) | 0.01332 ( 0.000298 ) | 0.013519 ( 2e-06 ) | -46.806473 ( 2.387943 ) | 1.492183 ( 2.270625 ) | 0.02473 ( 0.000132 ) | 1 ( 0 ) |
| -0.3 | ipw logit i | -0.000614 ( 0.000374 ) | 0.975 ( 0.004937 ) | 0.011813 ( 0.000264 ) | 0.013548 ( 2e-06 ) | -32.366588 ( 2.514757 ) | 14.689216 ( 2.56587 ) | 0.00014 ( 6e-06 ) | 1 ( 0 ) |
| -0.3 | ipw logit noi | -0.142859 ( 0.000424 ) | 0 ( 0 ) | 0.013406 ( 3e-04 ) | 0.013587 ( 2e-06 ) | -47.489067 ( 2.362358 ) | 1.347008 ( 2.267378 ) | 0.020588 ( 0.000122 ) | 1 ( 0 ) |
| -0.2 | True model | -0.000449 ( 0.000308 ) | 0.951 ( 0.006826 ) | 0.009733 ( 0.000218 ) | 0.00976 ( 1e-06 ) | 0 ( 0 ) | 0.280413 ( 2.243476 ) | 9.5e-05 ( 4e-06 ) | 1 ( 0 ) |
| -0.2 | CCA | -0.109769 ( 0.000426 ) | 0 ( 0 ) | 0.013481 ( 0.000302 ) | 0.013386 ( 2e-06 ) | -47.880546 ( 2.301515 ) | -0.704909 ( 2.221456 ) | 0.012231 ( 9.4e-05 ) | 1 ( 0 ) |
| -0.2 | ipw logadd i | -0.003986 ( 0.000371 ) | 0.97 ( 0.005394 ) | 0.011722 ( 0.000262 ) | 0.013499 ( 2e-06 ) | -31.058175 ( 2.527994 ) | 15.164222 ( 2.576501 ) | 0.000153 ( 7e-06 ) | 1 ( 0 ) |
| -0.2 | ipw logadd noi | -0.112149 ( 0.000429 ) | 0 ( 0 ) | 0.013564 ( 0.000303 ) | 0.013547 ( 2e-06 ) | -48.51407 ( 2.315408 ) | -0.127457 ( 2.234395 ) | 0.012761 ( 9.7e-05 ) | 1 ( 0 ) |
| -0.2 | ipw logit i | -0.000786 ( 0.000371 ) | 0.979 ( 0.004534 ) | 0.011721 ( 0.000262 ) | 0.013568 ( 2e-06 ) | -31.046597 ( 2.533373 ) | 15.762725 ( 2.589892 ) | 0.000138 ( 6e-06 ) | 1 ( 0 ) |
| -0.2 | ipw logit noi | -0.095894 ( 0.00043 ) | 0 ( 0 ) | 0.01361 ( 0.000304 ) | 0.013638 ( 2e-06 ) | -48.864566 ( 2.300654 ) | 0.205918 ( 2.241856 ) | 0.009381 ( 8.3e-05 ) | 1 ( 0 ) |
| -0.1 | True model | 0.000368 ( 0.000311 ) | 0.94 ( 0.00751 ) | 0.00983 ( 0.00022 ) | 0.00976 ( 1e-06 ) | 0 ( 0 ) | -0.710937 ( 2.221297 ) | 9.7e-05 ( 4e-06 ) | 1 ( 0 ) |
| -0.1 | CCA | -0.064634 ( 0.000424 ) | 0.002 ( 0.001413 ) | 0.013395 ( 3e-04 ) | 0.013381 ( 2e-06 ) | -46.142133 ( 2.360422 ) | -0.104857 ( 2.234879 ) | 0.004357 ( 5.5e-05 ) | 1 ( 0 ) |
| -0.1 | ipw logadd i | -0.002642 ( 0.00037 ) | 0.974 ( 0.005032 ) | 0.011706 ( 0.000262 ) | 0.013526 ( 2e-06 ) | -29.478262 ( 2.528582 ) | 15.548823 ( 2.585107 ) | 0.000144 ( 6e-06 ) | 1 ( 0 ) |
| -0.1 | ipw logadd noi | -0.066365 ( 0.000427 ) | 0.002 ( 0.001413 ) | 0.013507 ( 0.000302 ) | 0.01357 ( 2e-06 ) | -47.032538 ( 2.360022 ) | 0.465632 ( 2.247663 ) | 0.004587 ( 5.7e-05 ) | 1 ( 0 ) |
| -0.1 | ipw logit i | -0.000206 ( 0.00037 ) | 0.977 ( 0.00474 ) | 0.011716 ( 0.000262 ) | 0.01363 ( 2e-06 ) | -29.600872 ( 2.529322 ) | 16.338886 ( 2.602785 ) | 0.000137 ( 6e-06 ) | 1 ( 0 ) |
| -0.1 | ipw logit noi | -0.047541 ( 0.00043 ) | 0.066 ( 0.007851 ) | 0.013593 ( 0.000304 ) | 0.013688 ( 2e-06 ) | -47.700623 ( 2.327676 ) | 0.703349 ( 2.252985 ) | 0.002445 ( 4.2e-05 ) | 1 ( 0 ) |
| 0 | True model | -5e-05 ( 0.000319 ) | 0.944 ( 0.007271 ) | 0.010073 ( 0.000225 ) | 0.009759 ( 1e-06 ) | 0 ( 0 ) | -3.115756 ( 2.167496 ) | 0.000101 ( 5e-06 ) | 1 ( 0 ) |
| 0 | CCA | -0.021729 ( 0.000433 ) | 0.626 ( 0.015301 ) | 0.013699 ( 0.000306 ) | 0.013369 ( 2e-06 ) | -45.925383 ( 2.305929 ) | -2.408631 ( 2.183339 ) | 0.00066 ( 2.1e-05 ) | 1 ( 0 ) |
| 0 | ipw logadd i | -0.001001 ( 0.000384 ) | 0.971 ( 0.005307 ) | 0.01213 ( 0.000271 ) | 0.01356 ( 2e-06 ) | -31.031877 ( 2.454386 ) | 11.794074 ( 2.501106 ) | 0.000148 ( 6e-06 ) | 1 ( 0 ) |
| 0 | ipw logadd noi | -0.021942 ( 0.000438 ) | 0.631 ( 0.015259 ) | 0.013855 ( 0.00031 ) | 0.013579 ( 2e-06 ) | -47.139805 ( 2.309381 ) | -1.989254 ( 2.19274 ) | 0.000673 ( 2.1e-05 ) | 1 ( 0 ) |
| 0 | ipw logit i | -0.000542 ( 0.000384 ) | 0.973 ( 0.005126 ) | 0.012144 ( 0.000272 ) | 0.013729 ( 2e-06 ) | -31.192373 ( 2.457884 ) | 13.050013 ( 2.529212 ) | 0.000148 ( 6e-06 ) | 1 ( 0 ) |
| 0 | ipw logit noi | -0.000335 ( 0.000444 ) | 0.941 ( 0.007451 ) | 0.014026 ( 0.000314 ) | 0.013728 ( 2e-06 ) | -48.420165 ( 2.248185 ) | -2.125765 ( 2.18969 ) | 0.000197 ( 9e-06 ) | 1 ( 0 ) |
| 0.1 | True model | -0.000744 ( 0.000325 ) | 0.943 ( 0.007332 ) | 0.010266 ( 0.00023 ) | 0.009759 ( 1e-06 ) | 0 ( 0 ) | -4.936156 ( 2.126772 ) | 0.000106 ( 4e-06 ) | 1 ( 0 ) |
| 0.1 | CCA | 0.019842 ( 0.000434 ) | 0.679 ( 0.014763 ) | 0.01372 ( 0.000307 ) | 0.013354 ( 2e-06 ) | -44.009429 ( 2.33887 ) | -2.663853 ( 2.17763 ) | 0.000582 ( 1.9e-05 ) | 1 ( 0 ) |
| 0.1 | ipw logadd i | 0.001968 ( 0.000396 ) | 0.96 ( 0.006197 ) | 0.012531 ( 0.00028 ) | 0.013609 ( 2e-06 ) | -32.875589 ( 2.380042 ) | 8.609319 ( 2.429865 ) | 0.000161 ( 7e-06 ) | 1 ( 0 ) |
| 0.1 | ipw logadd noi | 0.022077 ( 0.000442 ) | 0.616 ( 0.01538 ) | 0.013969 ( 0.000313 ) | 0.013586 ( 2e-06 ) | -45.990689 ( 2.323597 ) | -2.742931 ( 2.175884 ) | 0.000682 ( 2.1e-05 ) | 1 ( 0 ) |
| 0.1 | ipw logit i | -0.001196 ( 0.000398 ) | 0.972 ( 0.005217 ) | 0.01257 ( 0.000281 ) | 0.01388 ( 3e-06 ) | -33.297995 ( 2.384785 ) | 10.419013 ( 2.47037 ) | 0.000159 ( 7e-06 ) | 1 ( 0 ) |
| 0.1 | ipw logit noi | 0.046644 ( 0.000447 ) | 0.085 ( 0.008819 ) | 0.01414 ( 0.000316 ) | 0.013767 ( 2e-06 ) | -47.283838 ( 2.257024 ) | -2.633513 ( 2.178336 ) | 0.002375 ( 4.3e-05 ) | 1 ( 0 ) |
| 0.2 | True model | -3e-05 ( 0.000318 ) | 0.944 ( 0.007271 ) | 0.010041 ( 0.000225 ) | 0.00976 ( 1e-06 ) | 0 ( 0 ) | -2.79585 ( 2.174653 ) | 0.000101 ( 5e-06 ) | 1 ( 0 ) |
| 0.2 | CCA | 0.061068 ( 0.000424 ) | 0.004 ( 0.001996 ) | 0.013397 ( 3e-04 ) | 0.01334 ( 2e-06 ) | -43.824173 ( 2.444875 ) | -0.424523 ( 2.227728 ) | 0.003909 ( 5.2e-05 ) | 1 ( 0 ) |
| 0.2 | ipw logadd i | 0.008148 ( 0.00038 ) | 0.94 ( 0.00751 ) | 0.01202 ( 0.000269 ) | 0.013665 ( 2e-06 ) | -30.220265 ( 2.651652 ) | 13.679977 ( 2.54331 ) | 0.000211 ( 9e-06 ) | 1 ( 0 ) |
| 0.2 | ipw logadd noi | 0.066688 ( 0.000433 ) | 0.002 ( 0.001413 ) | 0.013696 ( 0.000306 ) | 0.013586 ( 2e-06 ) | -46.254081 ( 2.427469 ) | -0.806268 ( 2.219208 ) | 0.004635 ( 5.8e-05 ) | 1 ( 0 ) |
| 0.2 | ipw logit i | -0.000558 ( 0.000382 ) | 0.976 ( 0.00484 ) | 0.01209 ( 0.00027 ) | 0.014085 ( 3e-06 ) | -31.018703 ( 2.662935 ) | 16.506148 ( 2.60658 ) | 0.000146 ( 7e-06 ) | 1 ( 0 ) |
| 0.2 | ipw logit noi | 0.094467 ( 0.000438 ) | 0 ( 0 ) | 0.013839 ( 0.00031 ) | 0.013801 ( 2e-06 ) | -47.353245 ( 2.35071 ) | -0.269346 ( 2.231227 ) | 0.009115 ( 8.3e-05 ) | 1 ( 0 ) |
| 0.3 | True model | -0.00014 ( 0.00032 ) | 0.944 ( 0.007271 ) | 0.010127 ( 0.000227 ) | 0.009758 ( 1e-06 ) | 0 ( 0 ) | -3.645512 ( 2.155644 ) | 0.000102 ( 5e-06 ) | 1 ( 0 ) |
| 0.3 | CCA | 0.098927 ( 0.000435 ) | 0 ( 0 ) | 0.013749 ( 0.000308 ) | 0.013316 ( 2e-06 ) | -45.748242 ( 2.357691 ) | -3.14955 ( 2.166762 ) | 0.009975 ( 8.6e-05 ) | 1 ( 0 ) |
| 0.3 | ipw logadd i | 0.015536 ( 0.000389 ) | 0.841 ( 0.011564 ) | 0.01229 ( 0.000275 ) | 0.01371 ( 3e-06 ) | -32.095337 ( 2.443189 ) | 11.560366 ( 2.495899 ) | 0.000392 ( 1.4e-05 ) | 1 ( 0 ) |
| 0.3 | ipw logadd noi | 0.109073 ( 0.000438 ) | 0 ( 0 ) | 0.013865 ( 0.00031 ) | 0.013569 ( 2e-06 ) | -46.649962 ( 2.361598 ) | -2.13705 ( 2.189438 ) | 0.012089 ( 9.6e-05 ) | 1 ( 0 ) |
| 0.3 | ipw logit i | -0.000586 ( 0.000393 ) | 0.972 ( 0.005217 ) | 0.012419 ( 0.000278 ) | 0.014339 ( 4e-06 ) | -33.505483 ( 2.429486 ) | 15.4568 ( 2.583165 ) | 0.000154 ( 7e-06 ) | 1 ( 0 ) |
| 0.3 | ipw logit noi | 0.140181 ( 0.000442 ) | 0 ( 0 ) | 0.013989 ( 0.000313 ) | 0.013819 ( 2e-06 ) | -47.590999 ( 2.293617 ) | -1.217344 ( 2.210023 ) | 0.019846 ( 0.000124 ) | 1 ( 0 ) |

###### Vary selection probability

###### Delta3=0

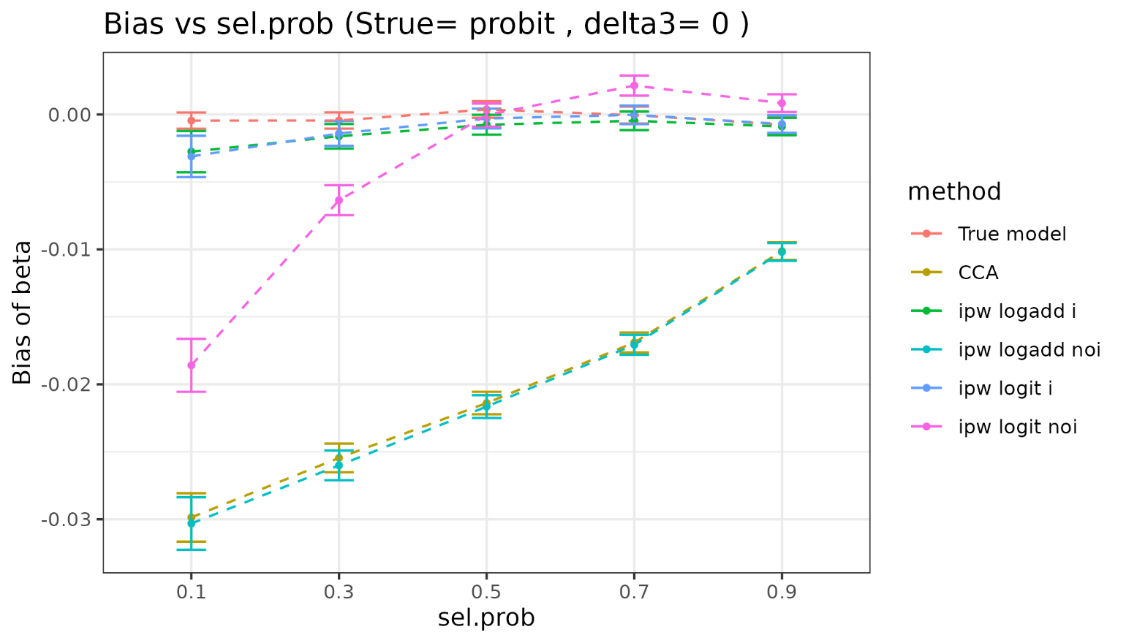

| sel.prob | method | bias | coverage | EmpSE | ModSE | relative_precision | relative_error_ModSE | MSE | power |
| --- | --- | --- | --- | --- | --- | --- | --- | --- | --- |
| 0.1 | True model | -0.000457 ( 0.000307 ) | 0.949 ( 0.006957 ) | 0.009715 ( 0.000217 ) | 0.00976 ( 1e-06 ) | 0 ( 0 ) | 0.462243 ( 2.247543 ) | 9.4e-05 ( 4e-06 ) | 1 ( 0 ) |
| 0.1 | CCA | -0.02986 ( 0.000915 ) | 0.825 ( 0.012016 ) | 0.02895 ( 0.000648 ) | 0.029078 ( 9e-06 ) | -88.739233 ( 0.676834 ) | 0.443171 ( 2.247301 ) | 0.001729 ( 6.7e-05 ) | 1 ( 0 ) |
| 0.1 | ipw logadd i | -0.002758 ( 0.000778 ) | 0.988 ( 0.003443 ) | 0.024616 ( 0.000551 ) | 0.031251 ( 1.5e-05 ) | -84.424936 ( 0.909524 ) | 26.954398 ( 2.840834 ) | 0.000613 ( 2.7e-05 ) | 1 ( 0 ) |
| 0.1 | ipw logadd noi | -0.03031 ( 0.000992 ) | 0.84 ( 0.011593 ) | 0.031384 ( 0.000702 ) | 0.031349 ( 1.5e-05 ) | -90.418516 ( 0.580499 ) | -0.113753 ( 2.235159 ) | 0.001903 ( 7.9e-05 ) | 1 ( 0 ) |
| 0.1 | ipw logit i | -0.003113 ( 0.000783 ) | 0.987 ( 0.003582 ) | 0.024768 ( 0.000554 ) | 0.031532 ( 1.6e-05 ) | -84.616103 ( 0.899216 ) | 27.307673 ( 2.848812 ) | 0.000623 ( 2.8e-05 ) | 1 ( 0 ) |
| 0.1 | ipw logit noi | -0.018588 ( 0.001001 ) | 0.91 ( 0.00905 ) | 0.03166 ( 0.000708 ) | 0.031593 ( 1.6e-05 ) | -90.584593 ( 0.570743 ) | -0.212387 ( 2.232994 ) | 0.001347 ( 6.3e-05 ) | 1 ( 0 ) |
| 0.3 | True model | -0.000449 ( 0.000308 ) | 0.951 ( 0.006826 ) | 0.009733 ( 0.000218 ) | 0.00976 ( 1e-06 ) | 0 ( 0 ) | 0.280413 ( 2.243476 ) | 9.5e-05 ( 4e-06 ) | 1 ( 0 ) |
| 0.3 | CCA | -0.025449 ( 0.000543 ) | 0.67 ( 0.014869 ) | 0.017179 ( 0.000384 ) | 0.017061 ( 3e-06 ) | -67.90278 ( 1.689813 ) | -0.686252 ( 2.221908 ) | 0.000942 ( 3.1e-05 ) | 1 ( 0 ) |
| 0.3 | ipw logadd i | -0.001616 ( 0.000464 ) | 0.98 ( 0.004427 ) | 0.014682 ( 0.000328 ) | 0.017594 ( 4e-06 ) | -56.056579 ( 2.15391 ) | 19.833957 ( 2.681047 ) | 0.000218 ( 1e-05 ) | 1 ( 0 ) |
| 0.3 | ipw logadd noi | -0.026005 ( 0.000562 ) | 0.684 ( 0.014702 ) | 0.01776 ( 0.000397 ) | 0.017633 ( 4e-06 ) | -69.967555 ( 1.62779 ) | -0.713822 ( 2.221335 ) | 0.000991 ( 3.2e-05 ) | 1 ( 0 ) |
| 0.3 | ipw logit i | -0.001411 ( 0.000465 ) | 0.981 ( 0.004317 ) | 0.014716 ( 0.000329 ) | 0.017812 ( 4e-06 ) | -56.259932 ( 2.150407 ) | 21.039789 ( 2.70804 ) | 0.000218 ( 1e-05 ) | 1 ( 0 ) |
| 0.3 | ipw logit noi | -0.006356 ( 0.000566 ) | 0.932 ( 0.007961 ) | 0.0179 ( 4e-04 ) | 0.017823 ( 4e-06 ) | -70.436133 ( 1.604557 ) | -0.428995 ( 2.227716 ) | 0.00036 ( 1.6e-05 ) | 1 ( 0 ) |
| 0.5 | True model | 0.000368 ( 0.000311 ) | 0.94 ( 0.00751 ) | 0.00983 ( 0.00022 ) | 0.00976 ( 1e-06 ) | 0 ( 0 ) | -0.710937 ( 2.221297 ) | 9.7e-05 ( 4e-06 ) | 1 ( 0 ) |
| 0.5 | CCA | -0.021384 ( 0.000426 ) | 0.639 ( 0.015188 ) | 0.013467 ( 0.000301 ) | 0.01337 ( 2e-06 ) | -46.717318 ( 2.36286 ) | -0.717199 ( 2.22118 ) | 0.000638 ( 2e-05 ) | 1 ( 0 ) |
| 0.5 | ipw logadd i | -0.000762 ( 0.000374 ) | 0.971 ( 0.005307 ) | 0.011813 ( 0.000264 ) | 0.013564 ( 2e-06 ) | -30.757481 ( 2.528761 ) | 14.822052 ( 2.56885 ) | 0.00014 ( 7e-06 ) | 1 ( 0 ) |
| 0.5 | ipw logadd noi | -0.021646 ( 0.000432 ) | 0.646 ( 0.015122 ) | 0.013648 ( 0.000305 ) | 0.013584 ( 2e-06 ) | -48.119439 ( 2.352356 ) | -0.469337 ( 2.226746 ) | 0.000655 ( 2e-05 ) | 1 ( 0 ) |
| 0.5 | ipw logit i | -3e-04 ( 0.000374 ) | 0.975 ( 0.004937 ) | 0.011834 ( 0.000265 ) | 0.013733 ( 2e-06 ) | -30.998734 ( 2.527054 ) | 16.048025 ( 2.596285 ) | 0.00014 ( 7e-06 ) | 1 ( 0 ) |
| 0.5 | ipw logit noi | -4.5e-05 ( 0.000435 ) | 0.952 ( 0.00676 ) | 0.013742 ( 0.000307 ) | 0.013732 ( 2e-06 ) | -48.828041 ( 2.311859 ) | -0.068881 ( 2.23571 ) | 0.000189 ( 9e-06 ) | 1 ( 0 ) |
| 0.7 | True model | -5e-05 ( 0.000319 ) | 0.944 ( 0.007271 ) | 0.010073 ( 0.000225 ) | 0.009759 ( 1e-06 ) | 0 ( 0 ) | -3.115756 ( 2.167496 ) | 0.000101 ( 5e-06 ) | 1 ( 0 ) |
| 0.7 | CCA | -0.016899 ( 0.000377 ) | 0.676 ( 0.014799 ) | 0.011907 ( 0.000266 ) | 0.011422 ( 1e-06 ) | -28.431993 ( 2.372399 ) | -4.074926 ( 2.146048 ) | 0.000427 ( 1.4e-05 ) | 1 ( 0 ) |
| 0.7 | ipw logadd i | -0.00047 ( 0.00035 ) | 0.959 ( 0.00627 ) | 0.011084 ( 0.000248 ) | 0.011485 ( 1e-06 ) | -17.401575 ( 2.076138 ) | 3.61599 ( 2.318122 ) | 0.000123 ( 6e-06 ) | 1 ( 0 ) |
| 0.7 | ipw logadd noi | -0.017067 ( 0.000377 ) | 0.675 ( 0.014811 ) | 0.011907 ( 0.000266 ) | 0.011493 ( 1e-06 ) | -28.429042 ( 2.41789 ) | -3.477258 ( 2.159431 ) | 0.000433 ( 1.4e-05 ) | 1 ( 0 ) |
| 0.7 | ipw logit i | -2.8e-05 ( 0.000351 ) | 0.958 ( 0.006343 ) | 0.011085 ( 0.000248 ) | 0.011604 ( 2e-06 ) | -17.422583 ( 2.070774 ) | 4.677477 ( 2.341873 ) | 0.000123 ( 6e-06 ) | 1 ( 0 ) |
| 0.7 | ipw logit noi | 0.002141 ( 0.00038 ) | 0.944 ( 0.007271 ) | 0.012019 ( 0.000269 ) | 0.0116 ( 2e-06 ) | -29.751385 ( 2.354998 ) | -3.482235 ( 2.159321 ) | 0.000149 ( 7e-06 ) | 1 ( 0 ) |
| 0.9 | True model | -0.000744 ( 0.000325 ) | 0.943 ( 0.007332 ) | 0.010266 ( 0.00023 ) | 0.009759 ( 1e-06 ) | 0 ( 0 ) | -4.936156 ( 2.126772 ) | 0.000106 ( 4e-06 ) | 1 ( 0 ) |
| 0.9 | CCA | -0.010128 ( 0.000335 ) | 0.82 ( 0.012149 ) | 0.010591 ( 0.000237 ) | 0.010194 ( 1e-06 ) | -6.032259 ( 1.726366 ) | -3.740676 ( 2.153519 ) | 0.000215 ( 8e-06 ) | 1 ( 0 ) |
| 0.9 | ipw logadd i | -0.000893 ( 0.00033 ) | 0.956 ( 0.006486 ) | 0.010451 ( 0.000234 ) | 0.010204 ( 1e-06 ) | -3.509757 ( 1.237928 ) | -2.364171 ( 2.184325 ) | 0.00011 ( 5e-06 ) | 1 ( 0 ) |
| 0.9 | ipw logadd noi | -0.010186 ( 0.000335 ) | 0.818 ( 0.012201 ) | 0.010589 ( 0.000237 ) | 0.010206 ( 1e-06 ) | -6.010325 ( 1.740025 ) | -3.621873 ( 2.156187 ) | 0.000216 ( 8e-06 ) | 1 ( 0 ) |
| 0.9 | ipw logit i | -0.000713 ( 0.000331 ) | 0.958 ( 0.006343 ) | 0.010459 ( 0.000234 ) | 0.010261 ( 1e-06 ) | -3.649817 ( 1.236725 ) | -1.890228 ( 2.194929 ) | 0.00011 ( 5e-06 ) | 1 ( 0 ) |
| 0.9 | ipw logit noi | 0.000839 ( 0.000335 ) | 0.952 ( 0.00676 ) | 0.0106 ( 0.000237 ) | 0.010259 ( 1e-06 ) | -6.198777 ( 1.698607 ) | -3.220684 ( 2.165163 ) | 0.000113 ( 5e-06 ) | 1 ( 0 ) |

###### Delta3=0.1

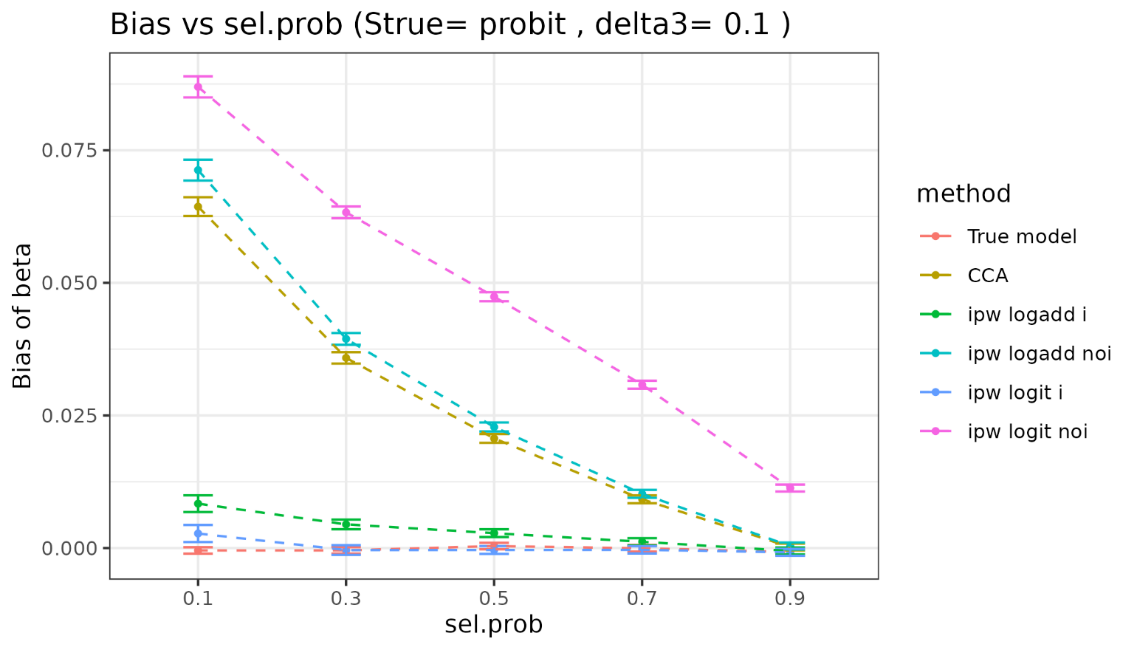

| sel.prob | method | bias | coverage | EmpSE | ModSE | relative_precision | relative_error_ModSE | MSE | power |
| --- | --- | --- | --- | --- | --- | --- | --- | --- | --- |
| 0.1 | True model | -0.000457 ( 0.000307 ) | 0.949 ( 0.006957 ) | 0.009715 ( 0.000217 ) | 0.00976 ( 1e-06 ) | 0 ( 0 ) | 0.462243 ( 2.247543 ) | 9.4e-05 ( 4e-06 ) | 1 ( 0 ) |
| 0.1 | CCA | 0.064371 ( 0.000906 ) | 0.395 ( 0.015459 ) | 0.028642 ( 0.000641 ) | 0.028925 ( 9e-06 ) | -88.495572 ( 0.692194 ) | 0.989713 ( 2.259532 ) | 0.004963 ( 0.000121 ) | 1 ( 0 ) |
| 0.1 | ipw logadd i | 0.008365 ( 0.000809 ) | 0.986 ( 0.003715 ) | 0.02559 ( 0.000572 ) | 0.031932 ( 1.8e-05 ) | -85.587634 ( 0.843712 ) | 24.786747 ( 2.792583 ) | 0.000724 ( 3e-05 ) | 1 ( 0 ) |
| 0.1 | ipw logadd noi | 0.071266 ( 0.001002 ) | 0.37 ( 0.015268 ) | 0.031697 ( 0.000709 ) | 0.031642 ( 1.6e-05 ) | -90.606708 ( 0.569392 ) | -0.17341 ( 2.233867 ) | 0.006083 ( 0.000147 ) | 1 ( 0 ) |
| 0.1 | ipw logit i | 0.002739 ( 0.000819 ) | 0.989 ( 0.003298 ) | 0.025906 ( 0.00058 ) | 0.032435 ( 2e-05 ) | -85.937292 ( 0.824379 ) | 25.20207 ( 2.802117 ) | 0.000678 ( 2.8e-05 ) | 1 ( 0 ) |
| 0.1 | ipw logit noi | 0.08696 ( 0.001014 ) | 0.217 ( 0.013035 ) | 0.032067 ( 0.000717 ) | 0.031965 ( 1.7e-05 ) | -90.821785 ( 0.556789 ) | -0.315588 ( 2.230745 ) | 0.008589 ( 0.00018 ) | 1 ( 0 ) |
| 0.3 | True model | -0.000449 ( 0.000308 ) | 0.951 ( 0.006826 ) | 0.009733 ( 0.000218 ) | 0.00976 ( 1e-06 ) | 0 ( 0 ) | 0.280413 ( 2.243476 ) | 9.5e-05 ( 4e-06 ) | 1 ( 0 ) |
| 0.3 | CCA | 0.035853 ( 0.000546 ) | 0.43 ( 0.015656 ) | 0.017253 ( 0.000386 ) | 0.017019 ( 3e-06 ) | -68.177869 ( 1.668853 ) | -1.355323 ( 2.206939 ) | 0.001583 ( 4.1e-05 ) | 1 ( 0 ) |
| 0.3 | ipw logadd i | 0.00446 ( 0.000467 ) | 0.98 ( 0.004427 ) | 0.01477 ( 0.00033 ) | 0.017746 ( 4e-06 ) | -56.577936 ( 2.154489 ) | 20.148211 ( 2.688108 ) | 0.000238 ( 1e-05 ) | 1 ( 0 ) |
| 0.3 | ipw logadd noi | 0.039443 ( 0.000564 ) | 0.383 ( 0.015372 ) | 0.017834 ( 0.000399 ) | 0.017675 ( 4e-06 ) | -70.215485 ( 1.615356 ) | -0.886998 ( 2.217468 ) | 0.001873 ( 4.6e-05 ) | 1 ( 0 ) |
| 0.3 | ipw logit i | -0.000388 ( 0.000469 ) | 0.981 ( 0.004317 ) | 0.014827 ( 0.000332 ) | 0.018109 ( 5e-06 ) | -56.914208 ( 2.150711 ) | 22.130106 ( 2.732496 ) | 0.00022 ( 1e-05 ) | 1 ( 0 ) |
| 0.3 | ipw logit noi | 0.063303 ( 0.000569 ) | 0.062 ( 0.007626 ) | 0.017994 ( 0.000403 ) | 0.017913 ( 4e-06 ) | -70.745425 ( 1.587191 ) | -0.453535 ( 2.227178 ) | 0.004331 ( 7.3e-05 ) | 1 ( 0 ) |
| 0.5 | True model | 0.000368 ( 0.000311 ) | 0.94 ( 0.00751 ) | 0.00983 ( 0.00022 ) | 0.00976 ( 1e-06 ) | 0 ( 0 ) | -0.710937 ( 2.221297 ) | 9.7e-05 ( 4e-06 ) | 1 ( 0 ) |
| 0.5 | CCA | 0.020671 ( 0.000426 ) | 0.663 ( 0.014948 ) | 0.013462 ( 0.000301 ) | 0.013357 ( 2e-06 ) | -46.679499 ( 2.357836 ) | -0.782276 ( 2.219724 ) | 0.000608 ( 2e-05 ) | 1 ( 0 ) |
| 0.5 | ipw logadd i | 0.002804 ( 0.000375 ) | 0.975 ( 0.004937 ) | 0.011872 ( 0.000266 ) | 0.013613 ( 2e-06 ) | -31.442307 ( 2.540377 ) | 14.664649 ( 2.565335 ) | 0.000149 ( 7e-06 ) | 1 ( 0 ) |
| 0.5 | ipw logadd noi | 0.022826 ( 0.000433 ) | 0.615 ( 0.015387 ) | 0.013701 ( 0.000307 ) | 0.01359 ( 2e-06 ) | -48.522863 ( 2.341997 ) | -0.809158 ( 2.219146 ) | 0.000709 ( 2.2e-05 ) | 1 ( 0 ) |
| 0.5 | ipw logit i | -0.000361 ( 0.000377 ) | 0.977 ( 0.00474 ) | 0.01191 ( 0.000266 ) | 0.013884 ( 3e-06 ) | -31.877582 ( 2.541062 ) | 16.574563 ( 2.608082 ) | 0.000142 ( 7e-06 ) | 1 ( 0 ) |
| 0.5 | ipw logit noi | 0.04739 ( 0.000437 ) | 0.068 ( 0.007961 ) | 0.013829 ( 0.000309 ) | 0.013772 ( 2e-06 ) | -49.467942 ( 2.28513 ) | -0.411815 ( 2.228041 ) | 0.002437 ( 4.3e-05 ) | 1 ( 0 ) |
| 0.7 | True model | -5e-05 ( 0.000319 ) | 0.944 ( 0.007271 ) | 0.010073 ( 0.000225 ) | 0.009759 ( 1e-06 ) | 0 ( 0 ) | -3.115756 ( 2.167496 ) | 0.000101 ( 5e-06 ) | 1 ( 0 ) |
| 0.7 | CCA | 0.009198 ( 0.000379 ) | 0.86 ( 0.010973 ) | 0.011996 ( 0.000268 ) | 0.011419 ( 1e-06 ) | -29.48285 ( 2.351212 ) | -4.804616 ( 2.129723 ) | 0.000228 ( 9e-06 ) | 1 ( 0 ) |
| 0.7 | ipw logadd i | 0.001173 ( 0.000352 ) | 0.956 ( 0.006486 ) | 0.01112 ( 0.000249 ) | 0.011494 ( 1e-06 ) | -17.934805 ( 2.079667 ) | 3.370575 ( 2.312632 ) | 0.000125 ( 6e-06 ) | 1 ( 0 ) |
| 0.7 | ipw logadd noi | 0.010211 ( 0.00038 ) | 0.845 ( 0.011444 ) | 0.012005 ( 0.000269 ) | 0.011489 ( 1e-06 ) | -29.58892 ( 2.396506 ) | -4.298339 ( 2.14106 ) | 0.000248 ( 1e-05 ) | 1 ( 0 ) |
| 0.7 | ipw logit i | -0.000338 ( 0.000352 ) | 0.961 ( 0.006122 ) | 0.011124 ( 0.000249 ) | 0.011681 ( 2e-06 ) | -17.991444 ( 2.077834 ) | 5.008311 ( 2.349279 ) | 0.000124 ( 6e-06 ) | 1 ( 0 ) |
| 0.7 | ipw logit noi | 0.030789 ( 0.000384 ) | 0.25 ( 0.013693 ) | 0.012139 ( 0.000272 ) | 0.011617 ( 2e-06 ) | -31.137337 ( 2.315496 ) | -4.296946 ( 2.141094 ) | 0.001095 ( 2.4e-05 ) | 1 ( 0 ) |
| 0.9 | True model | -0.000744 ( 0.000325 ) | 0.943 ( 0.007332 ) | 0.010266 ( 0.00023 ) | 0.009759 ( 1e-06 ) | 0 ( 0 ) | -4.936156 ( 2.126772 ) | 0.000106 ( 4e-06 ) | 1 ( 0 ) |
| 0.9 | CCA | 0.000191 ( 0.000334 ) | 0.952 ( 0.00676 ) | 0.010551 ( 0.000236 ) | 0.010196 ( 1e-06 ) | -5.328453 ( 1.761939 ) | -3.368724 ( 2.161841 ) | 0.000111 ( 5e-06 ) | 1 ( 0 ) |
| 0.9 | ipw logadd i | -0.000548 ( 0.00033 ) | 0.957 ( 0.006415 ) | 0.010431 ( 0.000233 ) | 0.010202 ( 1e-06 ) | -3.135969 ( 1.26315 ) | -2.194066 ( 2.18813 ) | 0.000109 ( 4e-06 ) | 1 ( 0 ) |
| 0.9 | ipw logadd noi | 0.000378 ( 0.000334 ) | 0.95 ( 0.006892 ) | 0.010552 ( 0.000236 ) | 0.010202 ( 1e-06 ) | -5.341213 ( 1.777929 ) | -3.315869 ( 2.163033 ) | 0.000111 ( 5e-06 ) | 1 ( 0 ) |
| 0.9 | ipw logit i | -0.000805 ( 0.00033 ) | 0.959 ( 0.00627 ) | 0.010443 ( 0.000234 ) | 0.010289 ( 1e-06 ) | -3.356304 ( 1.262273 ) | -1.474702 ( 2.204226 ) | 0.00011 ( 5e-06 ) | 1 ( 0 ) |
| 0.9 | ipw logit noi | 0.011309 ( 0.000334 ) | 0.786 ( 0.012969 ) | 0.010567 ( 0.000236 ) | 0.010265 ( 1e-06 ) | -5.618805 ( 1.722398 ) | -2.863847 ( 2.173147 ) | 0.000239 ( 9e-06 ) | 1 ( 0 ) |

##### 3.3.2 Y is binary

###### Vary delta3

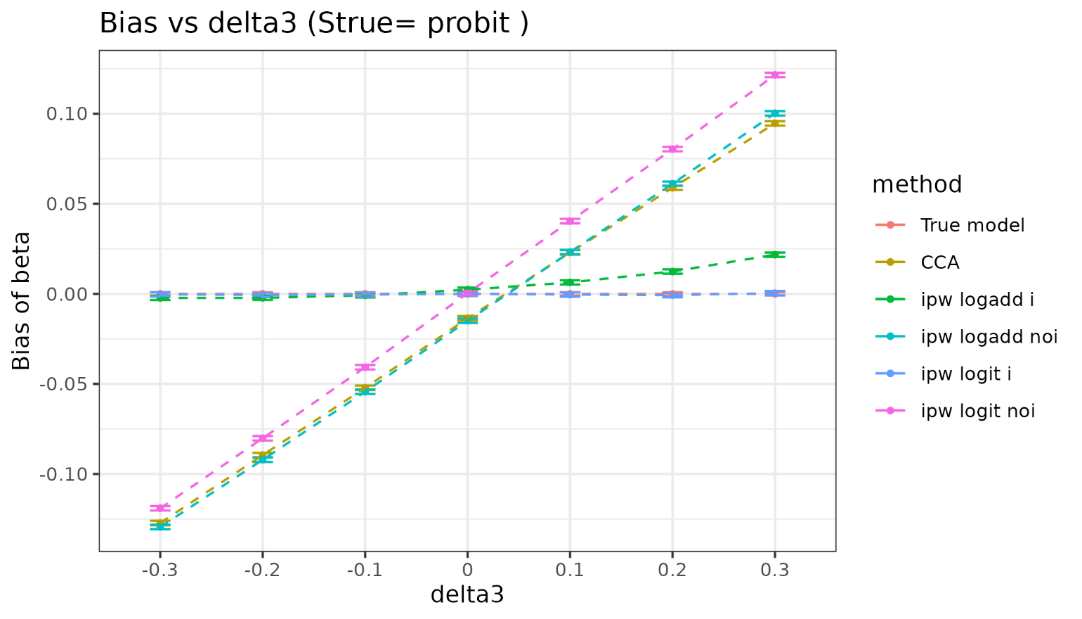

| delta3 | method | bias | coverage | EmpSE | ModSE | relative_precision | relative_error_ModSE | MSE | power |
| --- | --- | --- | --- | --- | --- | --- | --- | --- | --- |
| -0.3 | True model | 0 ( 0.00045 ) | 0.952 ( 0.00676 ) | 0.014234 ( 0.000318 ) | 0.014249 ( 1e-06 ) | 0 ( 0 ) | 0.111037 ( 2.239679 ) | 0.000202 ( 9e-06 ) | 1 ( 0 ) |
| -0.3 | CCA | -0.12721 ( 0.000632 ) | 0 ( 0 ) | 0.019992 ( 0.000447 ) | 0.019566 ( 2e-06 ) | -49.311843 ( 2.230507 ) | -2.134676 ( 2.189451 ) | 0.016582 ( 0.000161 ) | 1 ( 0 ) |
| -0.3 | ipw logadd i | -0.002272 ( 0.000614 ) | 0.958 ( 0.006343 ) | 0.019414 ( 0.000434 ) | 0.019625 ( 2e-06 ) | -46.247827 ( 2.231623 ) | 1.087676 ( 2.261542 ) | 0.000382 ( 1.8e-05 ) | 1 ( 0 ) |
| -0.3 | ipw logadd noi | -0.129401 ( 0.000638 ) | 0 ( 0 ) | 0.020167 ( 0.000451 ) | 0.019651 ( 2e-06 ) | -50.186433 ( 2.20573 ) | -2.557797 ( 2.179986 ) | 0.017151 ( 0.000165 ) | 1 ( 0 ) |
| -0.3 | ipw logit i | -0.00032 ( 0.000614 ) | 0.959 ( 0.00627 ) | 0.019404 ( 0.000434 ) | 0.019629 ( 2e-06 ) | -46.194017 ( 2.233368 ) | 1.156082 ( 2.263072 ) | 0.000376 ( 1.8e-05 ) | 1 ( 0 ) |
| -0.3 | ipw logit noi | -0.119025 ( 0.000637 ) | 0 ( 0 ) | 0.020154 ( 0.000451 ) | 0.019649 ( 2e-06 ) | -50.122758 ( 2.210678 ) | -2.508902 ( 2.18108 ) | 0.014573 ( 0.000152 ) | 1 ( 0 ) |
| -0.2 | True model | 0 ( 0.000443 ) | 0.954 ( 0.006624 ) | 0.014018 ( 0.000314 ) | 0.014249 ( 1e-06 ) | 0 ( 0 ) | 1.650235 ( 2.274113 ) | 0.000196 ( 9e-06 ) | 1 ( 0 ) |
| -0.2 | CCA | -0.089509 ( 0.000617 ) | 0.006 ( 0.002442 ) | 0.019522 ( 0.000437 ) | 0.019641 ( 2e-06 ) | -48.436301 ( 2.231645 ) | 0.609946 ( 2.250855 ) | 0.008393 ( 0.000111 ) | 1 ( 0 ) |
| -0.2 | ipw logadd i | -0.002214 ( 0.000594 ) | 0.959 ( 0.00627 ) | 0.018787 ( 0.00042 ) | 0.019711 ( 2e-06 ) | -44.326664 ( 2.306058 ) | 4.919512 ( 2.347269 ) | 0.000358 ( 1.7e-05 ) | 1 ( 0 ) |
| -0.2 | ipw logadd noi | -0.09208 ( 0.000625 ) | 0.006 ( 0.002442 ) | 0.019759 ( 0.000442 ) | 0.019753 ( 2e-06 ) | -49.670492 ( 2.203347 ) | -0.030748 ( 2.236522 ) | 0.008869 ( 0.000116 ) | 1 ( 0 ) |
| -0.2 | ipw logit i | -0.000574 ( 0.000593 ) | 0.966 ( 0.005731 ) | 0.018762 ( 0.00042 ) | 0.019715 ( 2e-06 ) | -44.17595 ( 2.312483 ) | 5.082006 ( 2.350904 ) | 0.000352 ( 1.7e-05 ) | 1 ( 0 ) |
| -0.2 | ipw logit noi | -0.080159 ( 0.000624 ) | 0.018 ( 0.004204 ) | 0.019734 ( 0.000441 ) | 0.019751 ( 2e-06 ) | -49.542896 ( 2.20889 ) | 0.086299 ( 2.239141 ) | 0.006815 ( 0.000101 ) | 1 ( 0 ) |
| -0.1 | True model | 0 ( 0.000453 ) | 0.947 ( 0.007085 ) | 0.014323 ( 0.00032 ) | 0.01425 ( 1e-06 ) | 0 ( 0 ) | -0.506395 ( 2.225866 ) | 0.000205 ( 9e-06 ) | 1 ( 0 ) |
| -0.1 | CCA | -0.052145 ( 0.000637 ) | 0.248 ( 0.013656 ) | 0.020152 ( 0.000451 ) | 0.019724 ( 2e-06 ) | -49.483331 ( 2.251748 ) | -2.125233 ( 2.189665 ) | 0.003125 ( 6.9e-05 ) | 1 ( 0 ) |
| -0.1 | ipw logadd i | -0.000812 ( 0.000606 ) | 0.951 ( 0.006826 ) | 0.019169 ( 0.000429 ) | 0.019827 ( 2e-06 ) | -44.16799 ( 2.394223 ) | 3.436672 ( 2.314098 ) | 0.000368 ( 1.7e-05 ) | 1 ( 0 ) |
| -0.1 | ipw logadd noi | -0.054312 ( 0.000642 ) | 0.219 ( 0.013078 ) | 0.020301 ( 0.000454 ) | 0.019866 ( 2e-06 ) | -50.223536 ( 2.241793 ) | -2.142258 ( 2.189285 ) | 0.003362 ( 7.3e-05 ) | 1 ( 0 ) |
| -0.1 | ipw logit i | -0.000423 ( 0.000605 ) | 0.953 ( 0.006693 ) | 0.019119 ( 0.000428 ) | 0.019835 ( 2e-06 ) | -43.880205 ( 2.40708 ) | 3.742033 ( 2.320929 ) | 0.000365 ( 1.7e-05 ) | 1 ( 0 ) |
| -0.1 | ipw logit noi | -0.040715 ( 0.000642 ) | 0.456 ( 0.01575 ) | 0.020287 ( 0.000454 ) | 0.019866 ( 2e-06 ) | -50.153136 ( 2.240203 ) | -2.074294 ( 2.190806 ) | 0.002069 ( 5.6e-05 ) | 1 ( 0 ) |
| 0 | True model | 0 ( 0.000442 ) | 0.953 ( 0.006693 ) | 0.013972 ( 0.000313 ) | 0.01425 ( 1e-06 ) | 0 ( 0 ) | 1.993794 ( 2.2818 ) | 0.000195 ( 9e-06 ) | 1 ( 0 ) |
| 0 | CCA | -0.013562 ( 0.000625 ) | 0.89 ( 0.009894 ) | 0.019775 ( 0.000442 ) | 0.019811 ( 2e-06 ) | -50.081408 ( 2.207346 ) | 0.181417 ( 2.241271 ) | 0.000575 ( 2.3e-05 ) | 1 ( 0 ) |
| 0 | ipw logadd i | 0.002298 ( 0.000608 ) | 0.961 ( 0.006122 ) | 0.019223 ( 0.00043 ) | 0.019969 ( 2e-06 ) | -47.171861 ( 2.283153 ) | 3.88003 ( 2.324019 ) | 0.000374 ( 1.6e-05 ) | 1 ( 0 ) |
| 0 | ipw logadd noi | -0.014901 ( 0.000638 ) | 0.876 ( 0.010422 ) | 0.020188 ( 0.000452 ) | 0.019985 ( 2e-06 ) | -52.101615 ( 2.153581 ) | -1.001716 ( 2.214803 ) | 0.000629 ( 2.5e-05 ) | 1 ( 0 ) |
| 0 | ipw logit i | -4.8e-05 ( 0.000606 ) | 0.966 ( 0.005731 ) | 0.019163 ( 0.000429 ) | 0.019989 ( 2e-06 ) | -46.840706 ( 2.301266 ) | 4.31056 ( 2.333652 ) | 0.000367 ( 1.6e-05 ) | 1 ( 0 ) |
| 0 | ipw logit noi | 0.000399 ( 0.000636 ) | 0.957 ( 0.006415 ) | 0.020123 ( 0.00045 ) | 0.019988 ( 2e-06 ) | -51.793125 ( 2.161359 ) | -0.670728 ( 2.222209 ) | 0.000405 ( 1.7e-05 ) | 1 ( 0 ) |
| 0.1 | True model | 0 ( 0.000454 ) | 0.952 ( 0.00676 ) | 0.014347 ( 0.000321 ) | 0.014249 ( 1e-06 ) | 0 ( 0 ) | -0.688115 ( 2.221801 ) | 0.000206 ( 9e-06 ) | 1 ( 0 ) |
| 0.1 | CCA | 0.023031 ( 0.000629 ) | 0.788 ( 0.012925 ) | 0.0199 ( 0.000445 ) | 0.0199 ( 2e-06 ) | -48.017298 ( 2.249853 ) | 0.004064 ( 2.237307 ) | 0.000926 ( 3.6e-05 ) | 1 ( 0 ) |
| 0.1 | ipw logadd i | 0.00635 ( 0.000611 ) | 0.946 ( 0.007147 ) | 0.019319 ( 0.000432 ) | 0.020129 ( 2e-06 ) | -44.845543 ( 2.286714 ) | 4.195054 ( 2.331072 ) | 0.000413 ( 1.9e-05 ) | 1 ( 0 ) |
| 0.1 | ipw logadd noi | 0.023203 ( 0.000634 ) | 0.795 ( 0.012766 ) | 0.020059 ( 0.000449 ) | 0.020108 ( 2e-06 ) | -48.840859 ( 2.235399 ) | 0.245051 ( 2.2427 ) | 0.00094 ( 3.6e-05 ) | 1 ( 0 ) |
| 0.1 | ipw logit i | -4e-04 ( 0.000609 ) | 0.963 ( 0.005969 ) | 0.019259 ( 0.000431 ) | 0.020179 ( 2e-06 ) | -44.500544 ( 2.304964 ) | 4.779488 ( 2.344149 ) | 0.000371 ( 1.7e-05 ) | 1 ( 0 ) |
| 0.1 | ipw logit noi | 0.040376 ( 0.000635 ) | 0.488 ( 0.015807 ) | 0.02009 ( 0.000449 ) | 0.020115 ( 2e-06 ) | -48.99681 ( 2.217992 ) | 0.12422 ( 2.239997 ) | 0.002033 ( 5.6e-05 ) | 1 ( 0 ) |
| 0.2 | True model | 0 ( 0.000448 ) | 0.951 ( 0.006826 ) | 0.014169 ( 0.000317 ) | 0.01425 ( 1e-06 ) | 0 ( 0 ) | 0.571514 ( 2.249981 ) | 0.000201 ( 9e-06 ) | 1 ( 0 ) |
| 0.2 | CCA | 0.058904 ( 0.000624 ) | 0.161 ( 0.011622 ) | 0.019748 ( 0.000442 ) | 0.019997 ( 2e-06 ) | -48.524808 ( 2.271139 ) | 1.261981 ( 2.265448 ) | 0.003859 ( 7.5e-05 ) | 1 ( 0 ) |
| 0.2 | ipw logadd i | 0.012401 ( 0.000612 ) | 0.917 ( 0.008724 ) | 0.019346 ( 0.000433 ) | 0.020313 ( 2e-06 ) | -46.361219 ( 2.287768 ) | 4.999303 ( 2.349064 ) | 0.000528 ( 2.2e-05 ) | 1 ( 0 ) |
| 0.2 | ipw logadd noi | 0.061131 ( 0.000635 ) | 0.146 ( 0.011166 ) | 0.020075 ( 0.000449 ) | 0.02024 ( 2e-06 ) | -50.187219 ( 2.223728 ) | 0.822004 ( 2.255606 ) | 0.00414 ( 7.9e-05 ) | 1 ( 0 ) |
| 0.2 | ipw logit i | -0.000708 ( 0.000611 ) | 0.968 ( 0.005566 ) | 0.019329 ( 0.000432 ) | 0.020421 ( 3e-06 ) | -46.2669 ( 2.305776 ) | 5.64819 ( 2.363586 ) | 0.000374 ( 1.7e-05 ) | 1 ( 0 ) |
| 0.2 | ipw logit noi | 0.08036 ( 0.000635 ) | 0.024 ( 0.00484 ) | 0.020073 ( 0.000449 ) | 0.020252 ( 2e-06 ) | -50.176647 ( 2.219173 ) | 0.890254 ( 2.257134 ) | 0.00686 ( 0.000103 ) | 1 ( 0 ) |
| 0.3 | True model | 0 ( 0.000442 ) | 0.946 ( 0.007147 ) | 0.013972 ( 0.000313 ) | 0.014249 ( 1e-06 ) | 0 ( 0 ) | 1.982761 ( 2.281553 ) | 0.000195 ( 9e-06 ) | 1 ( 0 ) |
| 0.3 | CCA | 0.094655 ( 0.000626 ) | 0.005 ( 0.00223 ) | 0.0198 ( 0.000443 ) | 0.0201 ( 2e-06 ) | -50.204863 ( 2.201664 ) | 1.516534 ( 2.271145 ) | 0.009351 ( 0.00012 ) | 1 ( 0 ) |
| 0.3 | ipw logadd i | 0.021744 ( 0.000612 ) | 0.84 ( 0.011593 ) | 0.01935 ( 0.000433 ) | 0.020513 ( 3e-06 ) | -47.861453 ( 2.258449 ) | 6.012302 ( 2.371731 ) | 0.000847 ( 3.1e-05 ) | 1 ( 0 ) |
| 0.3 | ipw logadd noi | 0.100225 ( 0.000633 ) | 0.002 ( 0.001413 ) | 0.020011 ( 0.000448 ) | 0.020377 ( 2e-06 ) | -51.249476 ( 2.181297 ) | 1.828831 ( 2.278134 ) | 0.010445 ( 0.000128 ) | 1 ( 0 ) |
| 0.3 | ipw logit i | 0.000368 ( 0.000613 ) | 0.967 ( 0.005649 ) | 0.019386 ( 0.000434 ) | 0.020719 ( 3e-06 ) | -48.05664 ( 2.267898 ) | 6.872325 ( 2.390981 ) | 0.000376 ( 1.8e-05 ) | 1 ( 0 ) |
| 0.3 | ipw logit noi | 0.121535 ( 0.000634 ) | 0 ( 0 ) | 0.020034 ( 0.000448 ) | 0.020395 ( 2e-06 ) | -51.358868 ( 2.170489 ) | 1.802822 ( 2.277552 ) | 0.015172 ( 0.000155 ) | 1 ( 0 ) |

###### Vary selection probability

###### Delta3=0

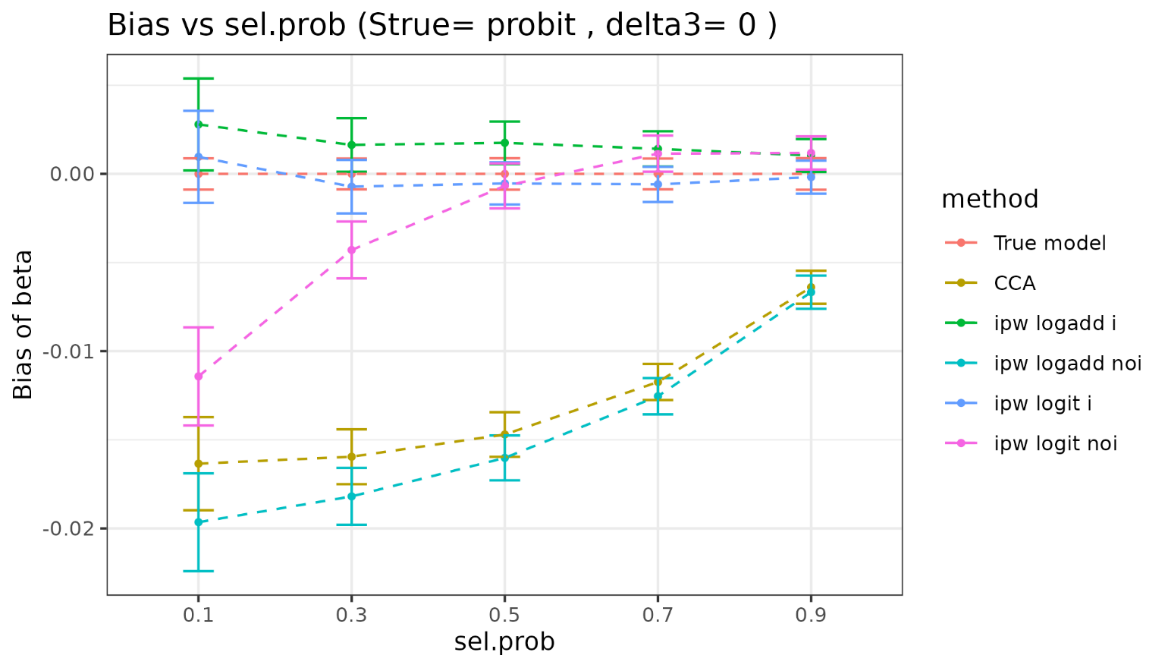

| sel.prob | method | bias | coverage | EmpSE | ModSE | relative_precision | relative_error_ModSE | MSE | power |
| --- | --- | --- | --- | --- | --- | --- | --- | --- | --- |
| 0.1 | True model | 0 ( 0.00045 ) | 0.952 ( 0.00676 ) | 0.014234 ( 0.000318 ) | 0.014249 ( 1e-06 ) | 0 ( 0 ) | 0.111037 ( 2.239679 ) | 0.000202 ( 9e-06 ) | 1 ( 0 ) |
| 0.1 | CCA | -0.016343 ( 0.001341 ) | 0.945 ( 0.007209 ) | 0.042414 ( 0.000949 ) | 0.043796 ( 1.3e-05 ) | -88.738074 ( 0.677543 ) | 3.257385 ( 2.310249 ) | 0.002064 ( 8.2e-05 ) | 1 ( 0 ) |
| 0.1 | ipw logadd i | 0.00279 ( 0.001323 ) | 0.964 ( 0.005891 ) | 0.041836 ( 0.000936 ) | 0.045586 ( 1.5e-05 ) | -88.424883 ( 0.692351 ) | 8.963861 ( 2.437981 ) | 0.001756 ( 8e-05 ) | 1 ( 0 ) |
| 0.1 | ipw logadd noi | -0.019644 ( 0.001408 ) | 0.935 ( 0.007796 ) | 0.04454 ( 0.000996 ) | 0.045677 ( 1.4e-05 ) | -89.787625 ( 0.61556 ) | 2.55102 ( 2.294488 ) | 0.002368 ( 9.6e-05 ) | 1 ( 0 ) |
| 0.1 | ipw logit i | 0.000959 ( 0.001325 ) | 0.965 ( 0.005812 ) | 0.041898 ( 0.000937 ) | 0.045679 ( 1.5e-05 ) | -88.459118 ( 0.690359 ) | 9.023472 ( 2.439323 ) | 0.001755 ( 8e-05 ) | 1 ( 0 ) |
| 0.1 | ipw logit noi | -0.01142 ( 0.001411 ) | 0.949 ( 0.006957 ) | 0.044614 ( 0.000998 ) | 0.04573 ( 1.5e-05 ) | -89.821117 ( 0.613583 ) | 2.50354 ( 2.29343 ) | 0.002119 ( 8.8e-05 ) | 1 ( 0 ) |
| 0.3 | True model | 0 ( 0.000443 ) | 0.954 ( 0.006624 ) | 0.014018 ( 0.000314 ) | 0.014249 ( 1e-06 ) | 0 ( 0 ) | 1.650235 ( 2.274113 ) | 0.000196 ( 9e-06 ) | 1 ( 0 ) |
| 0.3 | CCA | -0.015952 ( 0.00079 ) | 0.905 ( 0.009272 ) | 0.024992 ( 0.000559 ) | 0.025434 ( 4e-06 ) | -68.540031 ( 1.592771 ) | 1.766205 ( 2.276748 ) | 0.000878 ( 3.7e-05 ) | 1 ( 0 ) |
| 0.3 | ipw logadd i | 0.001632 ( 0.000772 ) | 0.966 ( 0.005731 ) | 0.024418 ( 0.000546 ) | 0.025874 ( 4e-06 ) | -67.041901 ( 1.659575 ) | 5.963534 ( 2.370659 ) | 0.000598 ( 2.7e-05 ) | 1 ( 0 ) |
| 0.3 | ipw logadd noi | -0.01819 ( 0.000818 ) | 0.898 ( 0.009571 ) | 0.025876 ( 0.000579 ) | 0.025908 ( 4e-06 ) | -70.650919 ( 1.505676 ) | 0.12721 ( 2.240085 ) | 0.001 ( 4.2e-05 ) | 1 ( 0 ) |
| 0.3 | ipw logit i | -0.000722 ( 0.000771 ) | 0.965 ( 0.005812 ) | 0.024395 ( 0.000546 ) | 0.025917 ( 4e-06 ) | -66.979541 ( 1.665222 ) | 6.240655 ( 2.376861 ) | 0.000595 ( 2.7e-05 ) | 1 ( 0 ) |
| 0.3 | ipw logit noi | -0.004292 ( 0.000818 ) | 0.946 ( 0.007147 ) | 0.025861 ( 0.000579 ) | 0.025924 ( 4e-06 ) | -70.617927 ( 1.506728 ) | 0.241904 ( 2.242652 ) | 0.000687 ( 3e-05 ) | 1 ( 0 ) |
| 0.5 | True model | 0 ( 0.000453 ) | 0.947 ( 0.007085 ) | 0.014323 ( 0.00032 ) | 0.01425 ( 1e-06 ) | 0 ( 0 ) | -0.506395 ( 2.225866 ) | 0.000205 ( 9e-06 ) | 1 ( 0 ) |
| 0.5 | CCA | -0.014698 ( 0.000641 ) | 0.878 ( 0.01035 ) | 0.020277 ( 0.000454 ) | 0.019811 ( 2e-06 ) | -50.104406 ( 2.238498 ) | -2.299608 ( 2.185765 ) | 0.000627 ( 2.7e-05 ) | 1 ( 0 ) |
| 0.5 | ipw logadd i | 0.001753 ( 0.000611 ) | 0.951 ( 0.006826 ) | 0.019309 ( 0.000432 ) | 0.019968 ( 2e-06 ) | -44.975483 ( 2.37529 ) | 3.414204 ( 2.313597 ) | 0.000376 ( 1.8e-05 ) | 1 ( 0 ) |
| 0.5 | ipw logadd noi | -0.016018 ( 0.000647 ) | 0.868 ( 0.010704 ) | 0.020459 ( 0.000458 ) | 0.019985 ( 2e-06 ) | -50.988067 ( 2.2303 ) | -2.313962 ( 2.185445 ) | 0.000675 ( 2.9e-05 ) | 1 ( 0 ) |
| 0.5 | ipw logit i | -0.000536 ( 0.000608 ) | 0.953 ( 0.006693 ) | 0.019236 ( 0.00043 ) | 0.019988 ( 2e-06 ) | -44.556173 ( 2.393987 ) | 3.911658 ( 2.324727 ) | 0.00037 ( 1.7e-05 ) | 1 ( 0 ) |
| 0.5 | ipw logit noi | -0.000678 ( 0.000646 ) | 0.945 ( 0.007209 ) | 0.020443 ( 0.000457 ) | 0.019988 ( 2e-06 ) | -50.912048 ( 2.227784 ) | -2.225814 ( 2.187417 ) | 0.000418 ( 2e-05 ) | 1 ( 0 ) |
| 0.7 | True model | 0 ( 0.000442 ) | 0.953 ( 0.006693 ) | 0.013972 ( 0.000313 ) | 0.01425 ( 1e-06 ) | 0 ( 0 ) | 1.993794 ( 2.2818 ) | 0.000195 ( 9e-06 ) | 1 ( 0 ) |
| 0.7 | CCA | -0.011737 ( 0.000518 ) | 0.903 ( 0.009359 ) | 0.016387 ( 0.000367 ) | 0.016837 ( 1e-06 ) | -27.302507 ( 2.450481 ) | 2.750123 ( 2.298727 ) | 0.000406 ( 1.7e-05 ) | 1 ( 0 ) |
| 0.7 | ipw logadd i | 0.001413 ( 0.000507 ) | 0.959 ( 0.00627 ) | 0.016039 ( 0.000359 ) | 0.016887 ( 1e-06 ) | -24.122069 ( 2.433561 ) | 5.286168 ( 2.355464 ) | 0.000259 ( 1.2e-05 ) | 1 ( 0 ) |
| 0.7 | ipw logadd noi | -0.012541 ( 0.000522 ) | 0.89 ( 0.009894 ) | 0.016513 ( 0.000369 ) | 0.016895 ( 1e-06 ) | -28.409355 ( 2.447474 ) | 2.313813 ( 2.288967 ) | 0.00043 ( 1.8e-05 ) | 1 ( 0 ) |
| 0.7 | ipw logit i | -0.000591 ( 0.000506 ) | 0.96 ( 0.006197 ) | 0.015989 ( 0.000358 ) | 0.016895 ( 1e-06 ) | -23.646702 ( 2.453166 ) | 5.663363 ( 2.363903 ) | 0.000256 ( 1.2e-05 ) | 1 ( 0 ) |
| 0.7 | ipw logit noi | 0.001143 ( 0.000521 ) | 0.959 ( 0.00627 ) | 0.016474 ( 0.000369 ) | 0.016894 ( 1e-06 ) | -28.075032 ( 2.442491 ) | 2.546157 ( 2.294165 ) | 0.000272 ( 1.2e-05 ) | 1 ( 0 ) |
| 0.9 | True model | 0 ( 0.000454 ) | 0.952 ( 0.00676 ) | 0.014347 ( 0.000321 ) | 0.014249 ( 1e-06 ) | 0 ( 0 ) | -0.688115 ( 2.221801 ) | 0.000206 ( 9e-06 ) | 1 ( 0 ) |
| 0.9 | CCA | -0.006392 ( 0.000476 ) | 0.926 ( 0.008278 ) | 0.015065 ( 0.000337 ) | 0.014947 ( 1e-06 ) | -9.302311 ( 1.618683 ) | -0.78407 ( 2.219656 ) | 0.000268 ( 1.2e-05 ) | 1 ( 0 ) |
| 0.9 | ipw logadd i | 0.001036 ( 0.000475 ) | 0.948 ( 0.007021 ) | 0.015031 ( 0.000336 ) | 0.014954 ( 1e-06 ) | -8.883982 ( 1.525346 ) | -0.512141 ( 2.22574 ) | 0.000227 ( 1e-05 ) | 1 ( 0 ) |
| 0.9 | ipw logadd noi | -0.006672 ( 0.000477 ) | 0.918 ( 0.008676 ) | 0.015093 ( 0.000338 ) | 0.014955 ( 1e-06 ) | -9.640083 ( 1.62396 ) | -0.916143 ( 2.216701 ) | 0.000272 ( 1.2e-05 ) | 1 ( 0 ) |
| 0.9 | ipw logit i | -0.000182 ( 0.000475 ) | 0.944 ( 0.007271 ) | 0.01501 ( 0.000336 ) | 0.014955 ( 1e-06 ) | -8.637795 ( 1.530691 ) | -0.36965 ( 2.228927 ) | 0.000225 ( 1e-05 ) | 1 ( 0 ) |
| 0.9 | ipw logit noi | 0.001176 ( 0.000478 ) | 0.95 ( 0.006892 ) | 0.015108 ( 0.000338 ) | 0.014954 ( 1e-06 ) | -9.819223 ( 1.607497 ) | -1.018255 ( 2.214417 ) | 0.000229 ( 1e-05 ) | 1 ( 0 ) |

###### Delta3=0.1

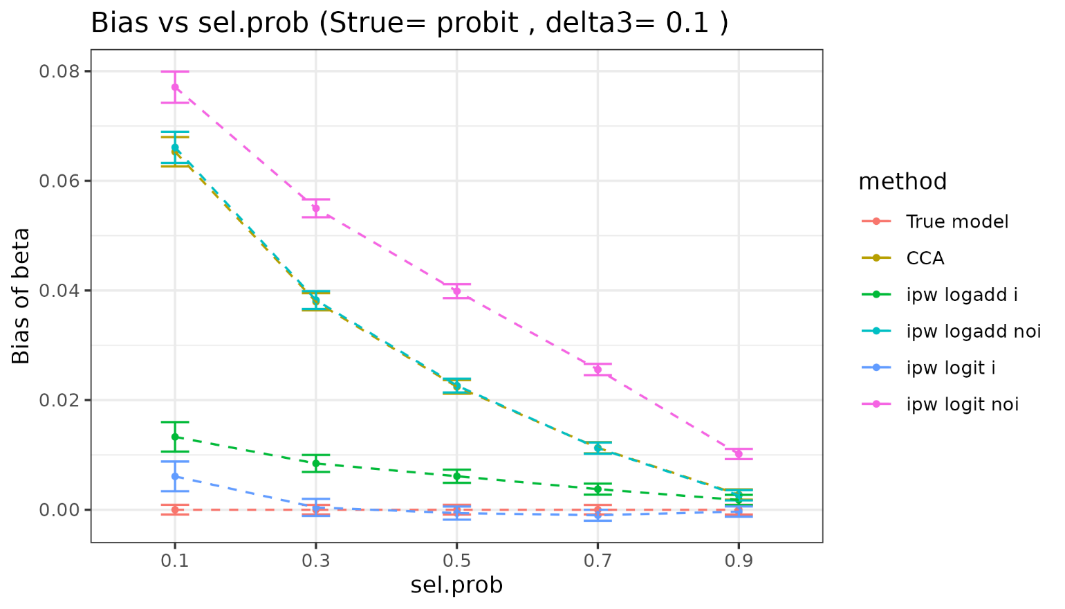

| sel.prob | method | bias | coverage | EmpSE | ModSE | relative_precision | relative_error_ModSE | MSE | power |
| --- | --- | --- | --- | --- | --- | --- | --- | --- | --- |
| 0.1 | True model | 0 ( 0.00045 ) | 0.952 ( 0.00676 ) | 0.014234 ( 0.000318 ) | 0.014249 ( 1e-06 ) | 0 ( 0 ) | 0.111037 ( 2.239679 ) | 0.000202 ( 9e-06 ) | 1 ( 0 ) |
| 0.1 | CCA | 0.065305 ( 0.001365 ) | 0.682 ( 0.014727 ) | 0.043165 ( 0.000966 ) | 0.044131 ( 1.3e-05 ) | -89.126644 ( 0.653842 ) | 2.236526 ( 2.28742 ) | 0.006126 ( 0.000203 ) | 1 ( 0 ) |
| 0.1 | ipw logadd i | 0.01329 ( 0.001377 ) | 0.953 ( 0.006693 ) | 0.043538 ( 0.000974 ) | 0.046739 ( 1.6e-05 ) | -89.311901 ( 0.639732 ) | 7.353174 ( 2.401985 ) | 0.00207 ( 9.4e-05 ) | 1 ( 0 ) |
| 0.1 | ipw logadd noi | 0.066109 ( 0.001447 ) | 0.714 ( 0.01429 ) | 0.045772 ( 0.001024 ) | 0.046455 ( 1.5e-05 ) | -90.329816 ( 0.582653 ) | 1.492836 ( 2.270829 ) | 0.006463 ( 0.000217 ) | 1 ( 0 ) |
| 0.1 | ipw logit i | 0.006082 ( 0.001383 ) | 0.958 ( 0.006343 ) | 0.043724 ( 0.000978 ) | 0.046966 ( 1.7e-05 ) | -89.402567 ( 0.634409 ) | 7.416179 ( 2.403417 ) | 0.001947 ( 8.8e-05 ) | 1 ( 0 ) |
| 0.1 | ipw logit noi | 0.077102 ( 0.001451 ) | 0.637 ( 0.015206 ) | 0.04589 ( 0.001027 ) | 0.04654 ( 1.5e-05 ) | -90.379642 ( 0.579666 ) | 1.416166 ( 2.269119 ) | 0.008049 ( 0.000247 ) | 1 ( 0 ) |
| 0.3 | True model | 0 ( 0.000443 ) | 0.954 ( 0.006624 ) | 0.014018 ( 0.000314 ) | 0.014249 ( 1e-06 ) | 0 ( 0 ) | 1.650235 ( 2.274113 ) | 0.000196 ( 9e-06 ) | 1 ( 0 ) |
| 0.3 | CCA | 0.037941 ( 0.000799 ) | 0.696 ( 0.014546 ) | 0.02526 ( 0.000565 ) | 0.025585 ( 4e-06 ) | -69.204051 ( 1.571368 ) | 1.283998 ( 2.265962 ) | 0.002077 ( 6.9e-05 ) | 1 ( 0 ) |
| 0.3 | ipw logadd i | 0.008443 ( 0.000792 ) | 0.951 ( 0.006826 ) | 0.025057 ( 0.000561 ) | 0.026225 ( 4e-06 ) | -68.701692 ( 1.589364 ) | 4.661309 ( 2.341531 ) | 0.000699 ( 3.1e-05 ) | 1 ( 0 ) |
| 0.3 | ipw logadd noi | 0.038244 ( 0.000835 ) | 0.696 ( 0.014546 ) | 0.026414 ( 0.000591 ) | 0.026158 ( 4e-06 ) | -71.835531 ( 1.457893 ) | -0.970409 ( 2.215531 ) | 0.00216 ( 7.2e-05 ) | 1 ( 0 ) |
| 0.3 | ipw logit i | 0.000413 ( 0.000793 ) | 0.963 ( 0.005969 ) | 0.025066 ( 0.000561 ) | 0.026331 ( 4e-06 ) | -68.725121 ( 1.591567 ) | 5.043954 ( 2.350097 ) | 0.000628 ( 2.8e-05 ) | 1 ( 0 ) |
| 0.3 | ipw logit noi | 0.054989 ( 0.000836 ) | 0.451 ( 0.015735 ) | 0.026438 ( 0.000591 ) | 0.026183 ( 4e-06 ) | -71.885634 ( 1.455447 ) | -0.963056 ( 2.215697 ) | 0.003722 ( 9.8e-05 ) | 1 ( 0 ) |
| 0.5 | True model | 0 ( 0.000453 ) | 0.947 ( 0.007085 ) | 0.014323 ( 0.00032 ) | 0.01425 ( 1e-06 ) | 0 ( 0 ) | -0.506395 ( 2.225866 ) | 0.000205 ( 9e-06 ) | 1 ( 0 ) |
| 0.5 | CCA | 0.022418 ( 0.000636 ) | 0.79 ( 0.01288 ) | 0.020119 ( 0.00045 ) | 0.019902 ( 2e-06 ) | -49.316873 ( 2.265245 ) | -1.075781 ( 2.213146 ) | 0.000907 ( 3.3e-05 ) | 1 ( 0 ) |
| 0.5 | ipw logadd i | 0.006113 ( 0.000613 ) | 0.947 ( 0.007085 ) | 0.019395 ( 0.000434 ) | 0.020132 ( 2e-06 ) | -45.46658 ( 2.358948 ) | 3.795684 ( 2.322134 ) | 0.000413 ( 1.9e-05 ) | 1 ( 0 ) |
| 0.5 | ipw logadd noi | 0.022645 ( 0.000645 ) | 0.793 ( 0.012812 ) | 0.020388 ( 0.000456 ) | 0.020111 ( 2e-06 ) | -50.645019 ( 2.243012 ) | -1.357099 ( 2.206853 ) | 0.000928 ( 3.4e-05 ) | 1 ( 0 ) |
| 0.5 | ipw logit i | -0.000618 ( 0.000611 ) | 0.953 ( 0.006693 ) | 0.019326 ( 0.000432 ) | 0.020182 ( 2e-06 ) | -45.072893 ( 2.379852 ) | 4.428126 ( 2.336285 ) | 0.000373 ( 1.8e-05 ) | 1 ( 0 ) |
| 0.5 | ipw logit noi | 0.039871 ( 0.000644 ) | 0.481 ( 0.0158 ) | 0.020377 ( 0.000456 ) | 0.020117 ( 2e-06 ) | -50.593211 ( 2.238437 ) | -1.273497 ( 2.208724 ) | 0.002005 ( 5.4e-05 ) | 1 ( 0 ) |
| 0.7 | True model | 0 ( 0.000442 ) | 0.953 ( 0.006693 ) | 0.013972 ( 0.000313 ) | 0.01425 ( 1e-06 ) | 0 ( 0 ) | 1.993794 ( 2.2818 ) | 0.000195 ( 9e-06 ) | 1 ( 0 ) |
| 0.7 | CCA | 0.011305 ( 0.00052 ) | 0.9 ( 0.009487 ) | 0.016444 ( 0.000368 ) | 0.016894 ( 1e-06 ) | -27.813335 ( 2.459172 ) | 2.73135 ( 2.298308 ) | 0.000398 ( 1.6e-05 ) | 1 ( 0 ) |
| 0.7 | ipw logadd i | 0.003755 ( 0.000511 ) | 0.954 ( 0.006624 ) | 0.016167 ( 0.000362 ) | 0.016967 ( 1e-06 ) | -25.314595 ( 2.414333 ) | 4.946547 ( 2.347868 ) | 0.000275 ( 1.2e-05 ) | 1 ( 0 ) |
| 0.7 | ipw logadd noi | 0.011233 ( 0.000524 ) | 0.902 ( 0.009402 ) | 0.016576 ( 0.000371 ) | 0.016962 ( 1e-06 ) | -28.952311 ( 2.457564 ) | 2.328087 ( 2.289287 ) | 0.000401 ( 1.7e-05 ) | 1 ( 0 ) |
| 0.7 | ipw logit i | -0.001 ( 0.000509 ) | 0.956 ( 0.006486 ) | 0.016096 ( 0.00036 ) | 0.016986 ( 1e-06 ) | -24.655501 ( 2.442315 ) | 5.529883 ( 2.360918 ) | 0.00026 ( 1.2e-05 ) | 1 ( 0 ) |
| 0.7 | ipw logit noi | 0.025569 ( 0.000523 ) | 0.672 ( 0.014846 ) | 0.016526 ( 0.00037 ) | 0.016962 ( 1e-06 ) | -28.523825 ( 2.450633 ) | 2.638678 ( 2.296236 ) | 0.000927 ( 2.9e-05 ) | 1 ( 0 ) |
| 0.9 | True model | 0 ( 0.000454 ) | 0.952 ( 0.00676 ) | 0.014347 ( 0.000321 ) | 0.014249 ( 1e-06 ) | 0 ( 0 ) | -0.688115 ( 2.221801 ) | 0.000206 ( 9e-06 ) | 1 ( 0 ) |
| 0.9 | CCA | 0.002768 ( 0.000478 ) | 0.947 ( 0.007085 ) | 0.01512 ( 0.000338 ) | 0.014971 ( 1e-06 ) | -9.961393 ( 1.646899 ) | -0.988741 ( 2.215077 ) | 0.000236 ( 1.1e-05 ) | 1 ( 0 ) |
| 0.9 | ipw logadd i | 0.001778 ( 0.000477 ) | 0.948 ( 0.007021 ) | 0.015092 ( 0.000338 ) | 0.01498 ( 1e-06 ) | -9.619245 ( 1.533776 ) | -0.73776 ( 2.220692 ) | 0.000231 ( 1e-05 ) | 1 ( 0 ) |
| 0.9 | ipw logadd noi | 0.002643 ( 0.000479 ) | 0.946 ( 0.007147 ) | 0.015156 ( 0.000339 ) | 0.01498 ( 1e-06 ) | -10.384969 ( 1.652997 ) | -1.160552 ( 2.211234 ) | 0.000236 ( 1.1e-05 ) | 1 ( 0 ) |
| 0.9 | ipw logit i | -0.000339 ( 0.000476 ) | 0.946 ( 0.007147 ) | 0.015063 ( 0.000337 ) | 0.014984 ( 1e-06 ) | -9.279868 ( 1.539527 ) | -0.529919 ( 2.225342 ) | 0.000227 ( 1e-05 ) | 1 ( 0 ) |
| 0.9 | ipw logit noi | 0.010165 ( 0.000479 ) | 0.891 ( 0.009855 ) | 0.015157 ( 0.000339 ) | 0.01498 ( 1e-06 ) | -10.402928 ( 1.630892 ) | -1.172387 ( 2.210969 ) | 0.000333 ( 1.4e-05 ) | 1 ( 0 ) |
