## Supplementary material for "Selection bias due to omitting interactions from inverse probability weighting": Supplementary 1-2 report of DAGs 2 and 3.docx

Supplementary 1-2 report for DAGs 2 and 3

### 1 DAG2 (only vary delta3)

X

S

Y

D1

C2

#### 1.1 Strue=logit

##### 1.1.1 Y is continuous

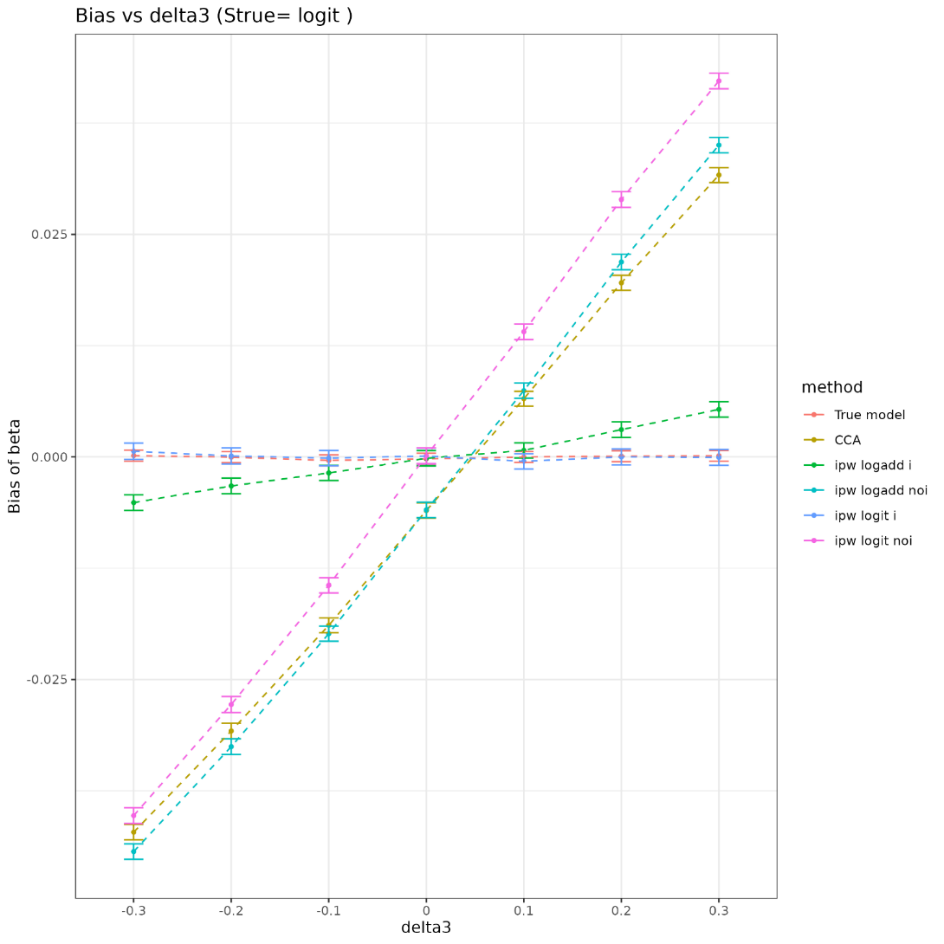

| delta3 | method | bias | coverage | EmpSE | ModSE | relative_precision | relative_error_ModSE | MSE | power |
| --- | --- | --- | --- | --- | --- | --- | --- | --- | --- |
| -0.3 | True model | 0.000146 ( 0.000318 ) | 0.941 ( 0.007451 ) | 0.010068 ( 0.000225 ) | 0.009758 ( 1e-06 ) | 0 ( 0 ) | -3.077896 ( 2.168344 ) | 0.000101 ( 5e-06 ) | 1 ( 0 ) |
| -0.3 | CCA | -0.042151 ( 0.000438 ) | 0.14 ( 0.010973 ) | 0.013851 ( 0.00031 ) | 0.013653 ( 2e-06 ) | -47.163211 ( 2.26679 ) | -1.427943 ( 2.205277 ) | 0.001968 ( 3.8e-05 ) | 1 ( 0 ) |
| -0.3 | ipw logadd i | -0.005133 ( 0.00045 ) | 0.938 ( 0.007626 ) | 0.014229 ( 0.000318 ) | 0.014084 ( 3e-06 ) | -49.93036 ( 2.26558 ) | -1.019376 ( 2.214474 ) | 0.000229 ( 1e-05 ) | 1 ( 0 ) |
| -0.3 | ipw logadd noi | -0.044332 ( 0.000445 ) | 0.114 ( 0.01005 ) | 0.014067 ( 0.000315 ) | 0.013916 ( 2e-06 ) | -48.775609 ( 2.248144 ) | -1.072811 ( 2.213247 ) | 0.002163 ( 4.1e-05 ) | 1 ( 0 ) |
| -0.3 | ipw logit i | 0.000639 ( 0.000468 ) | 0.954 ( 0.006624 ) | 0.014809 ( 0.000331 ) | 0.014756 ( 1e-05 ) | -53.776299 ( 2.158823 ) | -0.35596 ( 2.230277 ) | 0.000219 ( 1e-05 ) | 1 ( 0 ) |
| -0.3 | ipw logit noi | -0.040291 ( 0.000449 ) | 0.188 ( 0.012355 ) | 0.014193 ( 0.000318 ) | 0.014043 ( 2e-06 ) | -49.676864 ( 2.215484 ) | -1.053654 ( 2.213682 ) | 0.001825 ( 3.7e-05 ) | 1 ( 0 ) |
| -0.2 | True model | -3e-05 ( 0.000317 ) | 0.948 ( 0.007021 ) | 0.010028 ( 0.000224 ) | 0.009758 ( 1e-06 ) | 0 ( 0 ) | -2.69312 ( 2.176952 ) | 1e-04 ( 4e-06 ) | 1 ( 0 ) |
| -0.2 | CCA | -0.030788 ( 0.000444 ) | 0.367 ( 0.015242 ) | 0.014052 ( 0.000314 ) | 0.01364 ( 2e-06 ) | -49.071603 ( 2.143129 ) | -2.925649 ( 2.171769 ) | 0.001145 ( 2.8e-05 ) | 1 ( 0 ) |
| -0.2 | ipw logadd i | -0.00326 ( 0.000453 ) | 0.935 ( 0.007796 ) | 0.014311 ( 0.00032 ) | 0.014057 ( 3e-06 ) | -50.900646 ( 2.169589 ) | -1.777101 ( 2.197513 ) | 0.000215 ( 1e-05 ) | 1 ( 0 ) |
| -0.2 | ipw logadd noi | -0.032544 ( 0.000451 ) | 0.343 ( 0.015012 ) | 0.014266 ( 0.000319 ) | 0.013932 ( 2e-06 ) | -50.594117 ( 2.136376 ) | -2.346509 ( 2.184754 ) | 0.001262 ( 3e-05 ) | 1 ( 0 ) |
| -0.2 | ipw logit i | 0.000105 ( 0.000464 ) | 0.946 ( 0.007147 ) | 0.014682 ( 0.000328 ) | 0.014451 ( 5e-06 ) | -53.354029 ( 2.095934 ) | -1.574343 ( 2.202225 ) | 0.000215 ( 1e-05 ) | 1 ( 0 ) |
| -0.2 | ipw logit noi | -0.027811 ( 0.000456 ) | 0.501 ( 0.015811 ) | 0.014419 ( 0.000323 ) | 0.014082 ( 3e-06 ) | -51.632094 ( 2.100818 ) | -2.332656 ( 2.185071 ) | 0.000981 ( 2.6e-05 ) | 1 ( 0 ) |
| -0.1 | True model | -0.000392 ( 0.000311 ) | 0.946 ( 0.007147 ) | 0.009844 ( 0.00022 ) | 0.00976 ( 1e-06 ) | 0 ( 0 ) | -0.856261 ( 2.218046 ) | 9.7e-05 ( 4e-06 ) | 1 ( 0 ) |
| -0.1 | CCA | -0.018916 ( 0.000425 ) | 0.707 ( 0.014393 ) | 0.013448 ( 0.000301 ) | 0.013633 ( 2e-06 ) | -46.415364 ( 2.308539 ) | 1.380431 ( 2.268105 ) | 0.000538 ( 1.7e-05 ) | 1 ( 0 ) |
| -0.1 | ipw logadd i | -0.0018 ( 0.000433 ) | 0.959 ( 0.00627 ) | 0.013701 ( 0.000307 ) | 0.014017 ( 3e-06 ) | -48.376709 ( 2.304335 ) | 2.307653 ( 2.28889 ) | 0.000191 ( 8e-06 ) | 1 ( 0 ) |
| -0.1 | ipw logadd noi | -0.019856 ( 0.000432 ) | 0.694 ( 0.014573 ) | 0.01365 ( 0.000305 ) | 0.013945 ( 2e-06 ) | -47.992802 ( 2.304072 ) | 2.160418 ( 2.285584 ) | 0.00058 ( 1.8e-05 ) | 1 ( 0 ) |
| -0.1 | ipw logit i | -0.000131 ( 0.000441 ) | 0.96 ( 0.006197 ) | 0.013934 ( 0.000312 ) | 0.014256 ( 3e-06 ) | -50.088912 ( 2.245086 ) | 2.313359 ( 2.289051 ) | 0.000194 ( 9e-06 ) | 1 ( 0 ) |
| -0.1 | ipw logit noi | -0.014419 ( 0.000437 ) | 0.819 ( 0.012175 ) | 0.013808 ( 0.000309 ) | 0.014121 ( 3e-06 ) | -49.172936 ( 2.261342 ) | 2.267803 ( 2.287996 ) | 0.000398 ( 1.4e-05 ) | 1 ( 0 ) |
| 0 | True model | -0.000222 ( 0.000321 ) | 0.944 ( 0.007271 ) | 0.010154 ( 0.000227 ) | 0.009759 ( 1e-06 ) | 0 ( 0 ) | -3.889194 ( 2.150194 ) | 0.000103 ( 5e-06 ) | 1 ( 0 ) |
| 0 | CCA | -0.006024 ( 0.000439 ) | 0.918 ( 0.008676 ) | 0.013871 ( 0.00031 ) | 0.013619 ( 2e-06 ) | -46.419647 ( 2.399452 ) | -1.819569 ( 2.196517 ) | 0.000229 ( 1e-05 ) | 1 ( 0 ) |
| 0 | ipw logadd i | -0.000153 ( 0.000446 ) | 0.944 ( 0.007271 ) | 0.014115 ( 0.000316 ) | 0.013978 ( 2e-06 ) | -48.25101 ( 2.387686 ) | -0.968791 ( 2.215583 ) | 0.000199 ( 9e-06 ) | 1 ( 0 ) |
| 0 | ipw logadd noi | -0.005957 ( 0.000445 ) | 0.919 ( 0.008628 ) | 0.014076 ( 0.000315 ) | 0.013957 ( 2e-06 ) | -47.96698 ( 2.400398 ) | -0.846281 ( 2.218321 ) | 0.000233 ( 1e-05 ) | 1 ( 0 ) |
| 0 | ipw logit i | 9.7e-05 ( 0.000452 ) | 0.944 ( 0.007271 ) | 0.014308 ( 0.00032 ) | 0.014158 ( 3e-06 ) | -49.639694 ( 2.330906 ) | -1.049593 ( 2.213783 ) | 0.000205 ( 9e-06 ) | 1 ( 0 ) |
| 0 | ipw logit noi | 0.000129 ( 0.000452 ) | 0.938 ( 0.007626 ) | 0.014296 ( 0.00032 ) | 0.014158 ( 3e-06 ) | -49.556 ( 2.343294 ) | -0.967496 ( 2.21562 ) | 0.000204 ( 9e-06 ) | 1 ( 0 ) |
| 0.1 | True model | -1.1e-05 ( 0.00031 ) | 0.955 ( 0.006556 ) | 0.009787 ( 0.000219 ) | 0.00976 ( 1e-06 ) | 0 ( 0 ) | -0.277763 ( 2.23099 ) | 9.6e-05 ( 4e-06 ) | 1 ( 0 ) |
| 0.1 | CCA | 0.00654 ( 0.000427 ) | 0.922 ( 0.00848 ) | 0.013518 ( 0.000302 ) | 0.013608 ( 2e-06 ) | -47.579062 ( 2.424722 ) | 0.667029 ( 2.252149 ) | 0.000225 ( 1e-05 ) | 1 ( 0 ) |
| 0.1 | ipw logadd i | 0.000733 ( 0.000433 ) | 0.954 ( 0.006624 ) | 0.013699 ( 0.000306 ) | 0.013951 ( 2e-06 ) | -48.957199 ( 2.395234 ) | 1.834602 ( 2.278298 ) | 0.000188 ( 8e-06 ) | 1 ( 0 ) |
| 0.1 | ipw logadd noi | 0.007442 ( 0.000435 ) | 0.924 ( 0.00838 ) | 0.01374 ( 0.000307 ) | 0.013972 ( 2e-06 ) | -49.261413 ( 2.391331 ) | 1.688197 ( 2.275025 ) | 0.000244 ( 1.1e-05 ) | 1 ( 0 ) |
| 0.1 | ipw logit i | -0.000494 ( 0.000439 ) | 0.956 ( 0.006486 ) | 0.013874 ( 0.00031 ) | 0.014149 ( 3e-06 ) | -50.233686 ( 2.338334 ) | 1.981128 ( 2.281584 ) | 0.000193 ( 9e-06 ) | 1 ( 0 ) |
| 0.1 | ipw logit noi | 0.014061 ( 0.000441 ) | 0.833 ( 0.011795 ) | 0.013948 ( 0.000312 ) | 0.014196 ( 3e-06 ) | -50.756869 ( 2.331446 ) | 1.779111 ( 2.277071 ) | 0.000392 ( 1.5e-05 ) | 1 ( 0 ) |
| 0.2 | True model | 8.1e-05 ( 0.00031 ) | 0.949 ( 0.006957 ) | 0.009805 ( 0.000219 ) | 0.009759 ( 1e-06 ) | 0 ( 0 ) | -0.469642 ( 2.226696 ) | 9.6e-05 ( 4e-06 ) | 1 ( 0 ) |
| 0.2 | CCA | 0.019555 ( 0.000432 ) | 0.692 ( 0.014599 ) | 0.013647 ( 0.000305 ) | 0.013602 ( 2e-06 ) | -48.377128 ( 2.366225 ) | -0.33363 ( 2.22976 ) | 0.000568 ( 1.9e-05 ) | 1 ( 0 ) |
| 0.2 | ipw logadd i | 0.003068 ( 0.000443 ) | 0.949 ( 0.006957 ) | 0.014 ( 0.000313 ) | 0.013929 ( 2e-06 ) | -50.949104 ( 2.289852 ) | -0.511961 ( 2.225797 ) | 0.000205 ( 9e-06 ) | 1 ( 0 ) |
| 0.2 | ipw logadd noi | 0.021899 ( 0.000446 ) | 0.642 ( 0.01516 ) | 0.014092 ( 0.000315 ) | 0.013981 ( 2e-06 ) | -51.586074 ( 2.281032 ) | -0.790103 ( 2.219578 ) | 0.000678 ( 2.2e-05 ) | 1 ( 0 ) |
| 0.2 | ipw logit i | 0 ( 0.000452 ) | 0.955 ( 0.006556 ) | 0.01429 ( 0.00032 ) | 0.014222 ( 3e-06 ) | -52.918884 ( 2.206534 ) | -0.479853 ( 2.226533 ) | 0.000204 ( 9e-06 ) | 1 ( 0 ) |
| 0.2 | ipw logit noi | 0.028917 ( 0.000453 ) | 0.473 ( 0.015788 ) | 0.014334 ( 0.000321 ) | 0.01422 ( 3e-06 ) | -53.204395 ( 2.215764 ) | -0.792365 ( 2.219541 ) | 0.001041 ( 2.8e-05 ) | 1 ( 0 ) |
| 0.3 | True model | 0.00013 ( 0.00031 ) | 0.952 ( 0.00676 ) | 0.009813 ( 0.00022 ) | 0.009758 ( 1e-06 ) | 0 ( 0 ) | -0.565252 ( 2.224558 ) | 9.6e-05 ( 4e-06 ) | 1 ( 0 ) |
| 0.3 | CCA | 0.031668 ( 0.000426 ) | 0.347 ( 0.015053 ) | 0.013456 ( 0.000301 ) | 0.013597 ( 2e-06 ) | -46.811365 ( 2.405836 ) | 1.048011 ( 2.260671 ) | 0.001184 ( 2.8e-05 ) | 1 ( 0 ) |
| 0.3 | ipw logadd i | 0.005339 ( 0.000439 ) | 0.925 ( 0.008329 ) | 0.013872 ( 0.00031 ) | 0.013916 ( 2e-06 ) | -49.954922 ( 2.311137 ) | 0.314085 ( 2.244282 ) | 0.000221 ( 1e-05 ) | 1 ( 0 ) |
| 0.3 | ipw logadd noi | 0.035027 ( 0.000441 ) | 0.294 ( 0.014407 ) | 0.01396 ( 0.000312 ) | 0.013988 ( 2e-06 ) | -50.583949 ( 2.276659 ) | 0.197414 ( 2.241674 ) | 0.001422 ( 3.2e-05 ) | 1 ( 0 ) |
| 0.3 | ipw logit i | -5.3e-05 ( 0.000453 ) | 0.948 ( 0.007021 ) | 0.014322 ( 0.00032 ) | 0.014392 ( 4e-06 ) | -53.051622 ( 2.192706 ) | 0.49071 ( 2.248349 ) | 0.000205 ( 9e-06 ) | 1 ( 0 ) |
| 0.3 | ipw logit noi | 0.042236 ( 0.000449 ) | 0.145 ( 0.011134 ) | 0.01421 ( 0.000318 ) | 0.014234 ( 3e-06 ) | -52.306268 ( 2.200697 ) | 0.168534 ( 2.241044 ) | 0.001986 ( 3.9e-05 ) | 1 ( 0 ) |

##### 1.1.2 Y is binary

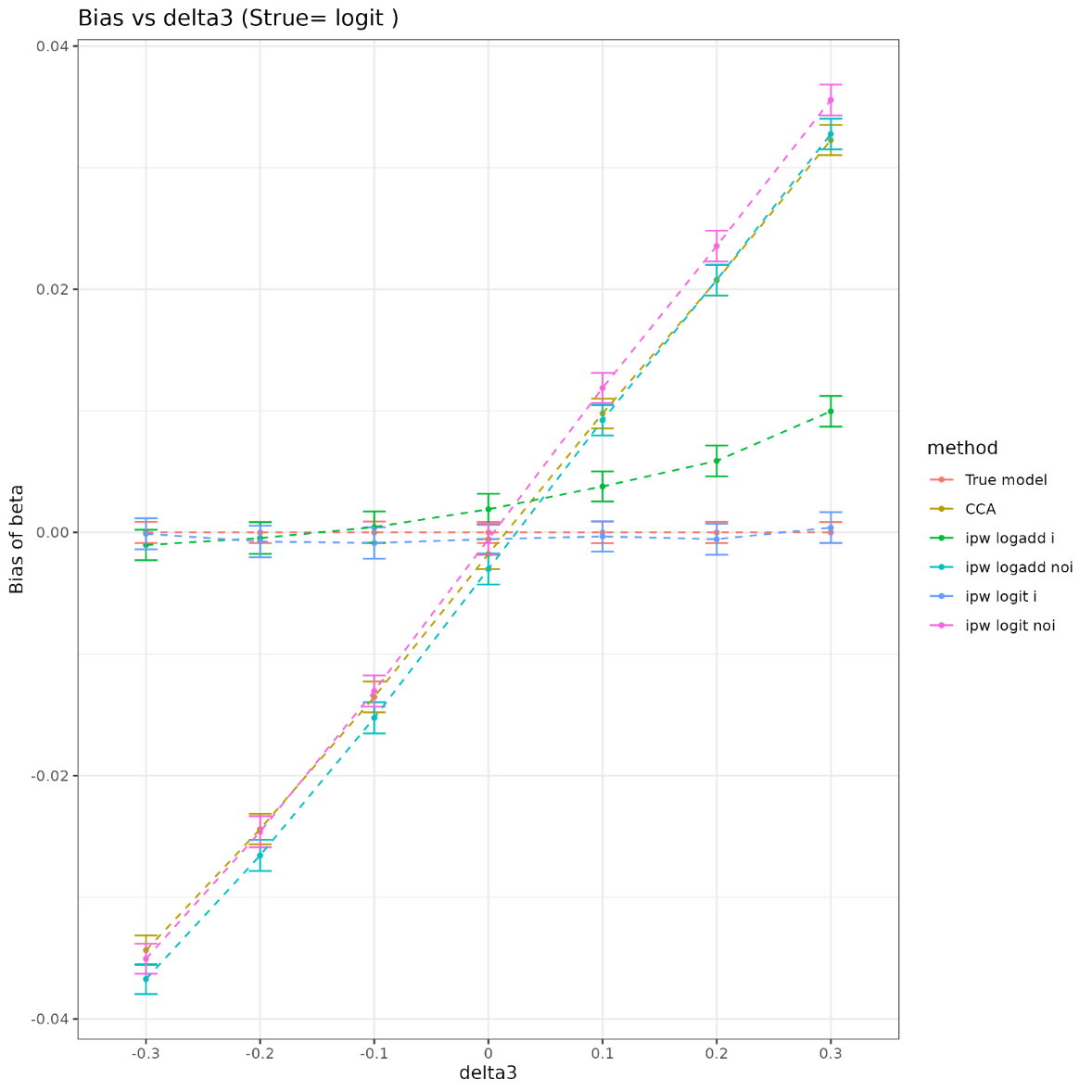

| delta3 | method | bias | coverage | EmpSE | ModSE | relative_precision | relative_error_ModSE | MSE | power |
| --- | --- | --- | --- | --- | --- | --- | --- | --- | --- |
| -0.3 | True model | 0 ( 0.000445 ) | 0.95 ( 0.006892 ) | 0.014063 ( 0.000315 ) | 0.01425 ( 1e-06 ) | 0 ( 0 ) | 1.326802 ( 2.266878 ) | 0.000198 ( 9e-06 ) | 1 ( 0 ) |
| -0.3 | CCA | -0.034351 ( 0.000618 ) | 0.612 ( 0.01541 ) | 0.019528 ( 0.000437 ) | 0.020153 ( 2e-06 ) | -48.138042 ( 2.397826 ) | 3.198481 ( 2.308768 ) | 0.001561 ( 4.6e-05 ) | 1 ( 0 ) |
| -0.3 | ipw logadd i | -0.00103 ( 0.000639 ) | 0.958 ( 0.006343 ) | 0.020214 ( 0.000452 ) | 0.020785 ( 2e-06 ) | -51.597254 ( 2.308383 ) | 2.826249 ( 2.300445 ) | 0.000409 ( 1.9e-05 ) | 1 ( 0 ) |
| -0.3 | ipw logadd noi | -0.036724 ( 0.000628 ) | 0.574 ( 0.015637 ) | 0.019848 ( 0.000444 ) | 0.020447 ( 2e-06 ) | -49.794887 ( 2.365465 ) | 3.017303 ( 2.304715 ) | 0.001742 ( 4.9e-05 ) | 1 ( 0 ) |
| -0.3 | ipw logit i | -0.00011 ( 0.000651 ) | 0.96 ( 0.006197 ) | 0.0206 ( 0.000461 ) | 0.021128 ( 5e-06 ) | -53.396379 ( 2.256881 ) | 2.560697 ( 2.294592 ) | 0.000424 ( 1.9e-05 ) | 1 ( 0 ) |
| -0.3 | ipw logit noi | -0.035056 ( 0.000629 ) | 0.605 ( 0.015459 ) | 0.019882 ( 0.000445 ) | 0.020482 ( 2e-06 ) | -49.968967 ( 2.361663 ) | 3.016704 ( 2.304703 ) | 0.001624 ( 4.7e-05 ) | 1 ( 0 ) |
| -0.2 | True model | 0 ( 0.000447 ) | 0.947 ( 0.007085 ) | 0.014145 ( 0.000316 ) | 0.014249 ( 1e-06 ) | 0 ( 0 ) | 0.731612 ( 2.253563 ) | 2e-04 ( 9e-06 ) | 1 ( 0 ) |
| -0.2 | CCA | -0.024392 ( 0.000642 ) | 0.749 ( 0.013711 ) | 0.020292 ( 0.000454 ) | 0.02017 ( 2e-06 ) | -51.405985 ( 2.208512 ) | -0.600615 ( 2.223774 ) | 0.001006 ( 3.4e-05 ) | 1 ( 0 ) |
| -0.2 | ipw logadd i | -0.000467 ( 0.000659 ) | 0.949 ( 0.006957 ) | 0.020825 ( 0.000466 ) | 0.020718 ( 2e-06 ) | -53.861432 ( 2.150789 ) | -0.515031 ( 2.225693 ) | 0.000433 ( 1.9e-05 ) | 1 ( 0 ) |
| -0.2 | ipw logadd noi | -0.026554 ( 0.000655 ) | 0.734 ( 0.013973 ) | 0.020719 ( 0.000464 ) | 0.020492 ( 2e-06 ) | -53.389938 ( 2.153417 ) | -1.095823 ( 2.212696 ) | 0.001134 ( 3.8e-05 ) | 1 ( 0 ) |
| -0.2 | ipw logit i | -0.000744 ( 0.000663 ) | 0.95 ( 0.006892 ) | 0.020955 ( 0.000469 ) | 0.020882 ( 3e-06 ) | -54.433953 ( 2.138757 ) | -0.348409 ( 2.229431 ) | 0.000439 ( 1.9e-05 ) | 1 ( 0 ) |
| -0.2 | ipw logit noi | -0.024616 ( 0.000656 ) | 0.754 ( 0.013619 ) | 0.020733 ( 0.000464 ) | 0.020537 ( 2e-06 ) | -53.453029 ( 2.154817 ) | -0.948658 ( 2.215989 ) | 0.001035 ( 3.5e-05 ) | 1 ( 0 ) |
| -0.1 | True model | 0 ( 0.000456 ) | 0.942 ( 0.007392 ) | 0.014407 ( 0.000322 ) | 0.014249 ( 1e-06 ) | 0 ( 0 ) | -1.093843 ( 2.212724 ) | 0.000207 ( 1e-05 ) | 1 ( 0 ) |
| -0.1 | CCA | -0.013525 ( 0.000645 ) | 0.896 ( 0.009653 ) | 0.020407 ( 0.000457 ) | 0.020192 ( 2e-06 ) | -50.158688 ( 2.261483 ) | -1.054118 ( 2.213629 ) | 0.000599 ( 2.6e-05 ) | 1 ( 0 ) |
| -0.1 | ipw logadd i | 0.000438 ( 0.000654 ) | 0.95 ( 0.006892 ) | 0.020679 ( 0.000463 ) | 0.020658 ( 2e-06 ) | -51.464485 ( 2.247353 ) | -0.10512 ( 2.234863 ) | 0.000427 ( 1.9e-05 ) | 1 ( 0 ) |
| -0.1 | ipw logadd noi | -0.015246 ( 0.000654 ) | 0.884 ( 0.010126 ) | 0.020676 ( 0.000463 ) | 0.02054 ( 2e-06 ) | -51.447003 ( 2.236738 ) | -0.658513 ( 2.222481 ) | 0.00066 ( 2.8e-05 ) | 1 ( 0 ) |
| -0.1 | ipw logit i | -0.000869 ( 0.000656 ) | 0.949 ( 0.006957 ) | 0.020749 ( 0.000464 ) | 0.020729 ( 2e-06 ) | -51.787533 ( 2.236202 ) | -0.096178 ( 2.235065 ) | 0.000431 ( 1.9e-05 ) | 1 ( 0 ) |
| -0.1 | ipw logit noi | -0.013038 ( 0.000655 ) | 0.899 ( 0.009529 ) | 0.020703 ( 0.000463 ) | 0.020594 ( 2e-06 ) | -51.575747 ( 2.233867 ) | -0.528494 ( 2.22539 ) | 0.000598 ( 2.6e-05 ) | 1 ( 0 ) |
| 0 | True model | 0 ( 0.000444 ) | 0.953 ( 0.006693 ) | 0.014039 ( 0.000314 ) | 0.01425 ( 1e-06 ) | 0 ( 0 ) | 1.50209 ( 2.2708 ) | 0.000197 ( 9e-06 ) | 1 ( 0 ) |
| 0 | CCA | -0.001773 ( 0.000635 ) | 0.952 ( 0.00676 ) | 0.02009 ( 0.000449 ) | 0.020215 ( 2e-06 ) | -51.165638 ( 2.260709 ) | 0.619005 ( 2.25106 ) | 0.000406 ( 1.8e-05 ) | 1 ( 0 ) |
| 0 | ipw logadd i | 0.001909 ( 0.000647 ) | 0.95 ( 0.006892 ) | 0.020468 ( 0.000458 ) | 0.020613 ( 2e-06 ) | -52.952166 ( 2.209677 ) | 0.708838 ( 2.253072 ) | 0.000422 ( 2e-05 ) | 1 ( 0 ) |
| 0 | ipw logadd noi | -0.003006 ( 0.000649 ) | 0.945 ( 0.007209 ) | 0.020508 ( 0.000459 ) | 0.020583 ( 2e-06 ) | -53.135512 ( 2.202261 ) | 0.363762 ( 2.245351 ) | 0.000429 ( 2e-05 ) | 1 ( 0 ) |
| 0 | ipw logit i | -0.000551 ( 0.000649 ) | 0.951 ( 0.006826 ) | 0.020508 ( 0.000459 ) | 0.020647 ( 2e-06 ) | -53.132703 ( 2.201279 ) | 0.677583 ( 2.252373 ) | 0.00042 ( 1.9e-05 ) | 1 ( 0 ) |
| 0 | ipw logit noi | -0.000558 ( 0.000649 ) | 0.949 ( 0.006957 ) | 0.020537 ( 0.000459 ) | 0.020646 ( 2e-06 ) | -53.26801 ( 2.200158 ) | 0.531154 ( 2.249097 ) | 0.000422 ( 1.9e-05 ) | 1 ( 0 ) |
| 0.1 | True model | 0 ( 0.000451 ) | 0.943 ( 0.007332 ) | 0.014254 ( 0.000319 ) | 0.014249 ( 1e-06 ) | 0 ( 0 ) | -0.037078 ( 2.236365 ) | 0.000203 ( 1e-05 ) | 1 ( 0 ) |
| 0.1 | CCA | 0.009781 ( 0.000624 ) | 0.93 ( 0.008068 ) | 0.019732 ( 0.000441 ) | 0.020233 ( 2e-06 ) | -47.814705 ( 2.421448 ) | 2.535294 ( 2.293932 ) | 0.000485 ( 2.2e-05 ) | 1 ( 0 ) |
| 0.1 | ipw logadd i | 0.003782 ( 0.000633 ) | 0.952 ( 0.00676 ) | 0.020024 ( 0.000448 ) | 0.020589 ( 2e-06 ) | -49.324357 ( 2.390395 ) | 2.822766 ( 2.300364 ) | 0.000415 ( 1.9e-05 ) | 1 ( 0 ) |
| 0.1 | ipw logadd noi | 0.009227 ( 0.000634 ) | 0.926 ( 0.008278 ) | 0.020052 ( 0.000449 ) | 0.020614 ( 2e-06 ) | -49.466015 ( 2.38715 ) | 2.802538 ( 2.299912 ) | 0.000487 ( 2.2e-05 ) | 1 ( 0 ) |
| 0.1 | ipw logit i | -0.000327 ( 0.000634 ) | 0.962 ( 0.006046 ) | 0.020044 ( 0.000448 ) | 0.020627 ( 2e-06 ) | -49.426937 ( 2.390144 ) | 2.905857 ( 2.302224 ) | 0.000401 ( 1.8e-05 ) | 1 ( 0 ) |
| 0.1 | ipw logit noi | 0.011877 ( 0.000635 ) | 0.915 ( 0.008819 ) | 0.020089 ( 0.000449 ) | 0.020685 ( 2e-06 ) | -49.64939 ( 2.388294 ) | 2.971284 ( 2.303688 ) | 0.000544 ( 2.4e-05 ) | 1 ( 0 ) |
| 0.2 | True model | 0 ( 0.000447 ) | 0.948 ( 0.007021 ) | 0.01415 ( 0.000317 ) | 0.014249 ( 1e-06 ) | 0 ( 0 ) | 0.702921 ( 2.252921 ) | 2e-04 ( 9e-06 ) | 1 ( 0 ) |
| 0.2 | CCA | 0.020726 ( 0.000639 ) | 0.817 ( 0.012227 ) | 0.02021 ( 0.000452 ) | 0.02025 ( 2e-06 ) | -50.98158 ( 2.213976 ) | 0.194503 ( 2.241563 ) | 0.000838 ( 3.3e-05 ) | 1 ( 0 ) |
| 0.2 | ipw logadd i | 0.005886 ( 0.000647 ) | 0.939 ( 0.007568 ) | 0.020473 ( 0.000458 ) | 0.020595 ( 2e-06 ) | -52.229487 ( 2.195076 ) | 0.598919 ( 2.250612 ) | 0.000453 ( 2e-05 ) | 1 ( 0 ) |
| 0.2 | ipw logadd noi | 0.020758 ( 0.000645 ) | 0.828 ( 0.011934 ) | 0.020396 ( 0.000456 ) | 0.020636 ( 2e-06 ) | -51.869254 ( 2.208285 ) | 1.178398 ( 2.263577 ) | 0.000846 ( 3.3e-05 ) | 1 ( 0 ) |
| 0.2 | ipw logit i | -0.000553 ( 0.00065 ) | 0.947 ( 0.007085 ) | 0.020554 ( 0.00046 ) | 0.020677 ( 2e-06 ) | -52.608637 ( 2.188809 ) | 0.595813 ( 2.250543 ) | 0.000422 ( 1.8e-05 ) | 1 ( 0 ) |
| 0.2 | ipw logit noi | 0.023553 ( 0.000646 ) | 0.8 ( 0.012649 ) | 0.020421 ( 0.000457 ) | 0.020712 ( 2e-06 ) | -51.986703 ( 2.207848 ) | 1.425963 ( 2.269117 ) | 0.000971 ( 3.7e-05 ) | 1 ( 0 ) |
| 0.3 | True model | 0 ( 0.000437 ) | 0.951 ( 0.006826 ) | 0.013822 ( 0.000309 ) | 0.01425 ( 1e-06 ) | 0 ( 0 ) | 3.090203 ( 2.306328 ) | 0.000191 ( 9e-06 ) | 1 ( 0 ) |
| 0.3 | CCA | 0.032266 ( 0.000631 ) | 0.637 ( 0.015206 ) | 0.019966 ( 0.000447 ) | 0.02027 ( 2e-06 ) | -52.069648 ( 2.148318 ) | 1.522712 ( 2.271279 ) | 0.001439 ( 4.4e-05 ) | 1 ( 0 ) |
| 0.3 | ipw logadd i | 0.00996 ( 0.000644 ) | 0.932 ( 0.007961 ) | 0.020351 ( 0.000455 ) | 0.020632 ( 2e-06 ) | -53.866868 ( 2.10267 ) | 1.381982 ( 2.268131 ) | 0.000513 ( 2.2e-05 ) | 1 ( 0 ) |
| 0.3 | ipw logadd noi | 0.032765 ( 0.000643 ) | 0.635 ( 0.015224 ) | 0.020331 ( 0.000455 ) | 0.020652 ( 2e-06 ) | -53.779507 ( 2.109386 ) | 1.578129 ( 2.27252 ) | 0.001487 ( 4.5e-05 ) | 1 ( 0 ) |
| 0.3 | ipw logit i | 0.000396 ( 0.000649 ) | 0.954 ( 0.006624 ) | 0.020535 ( 0.000459 ) | 0.020805 ( 2e-06 ) | -54.69008 ( 2.077083 ) | 1.316621 ( 2.266673 ) | 0.000421 ( 1.9e-05 ) | 1 ( 0 ) |
| 0.3 | ipw logit noi | 0.035562 ( 0.000646 ) | 0.592 ( 0.015541 ) | 0.020414 ( 0.000457 ) | 0.020729 ( 2e-06 ) | -54.15232 ( 2.102539 ) | 1.541568 ( 2.271703 ) | 0.001681 ( 4.9e-05 ) | 1 ( 0 ) |

#### 1.2 Strue=log-additive

##### 1.2.1 Y is continuous

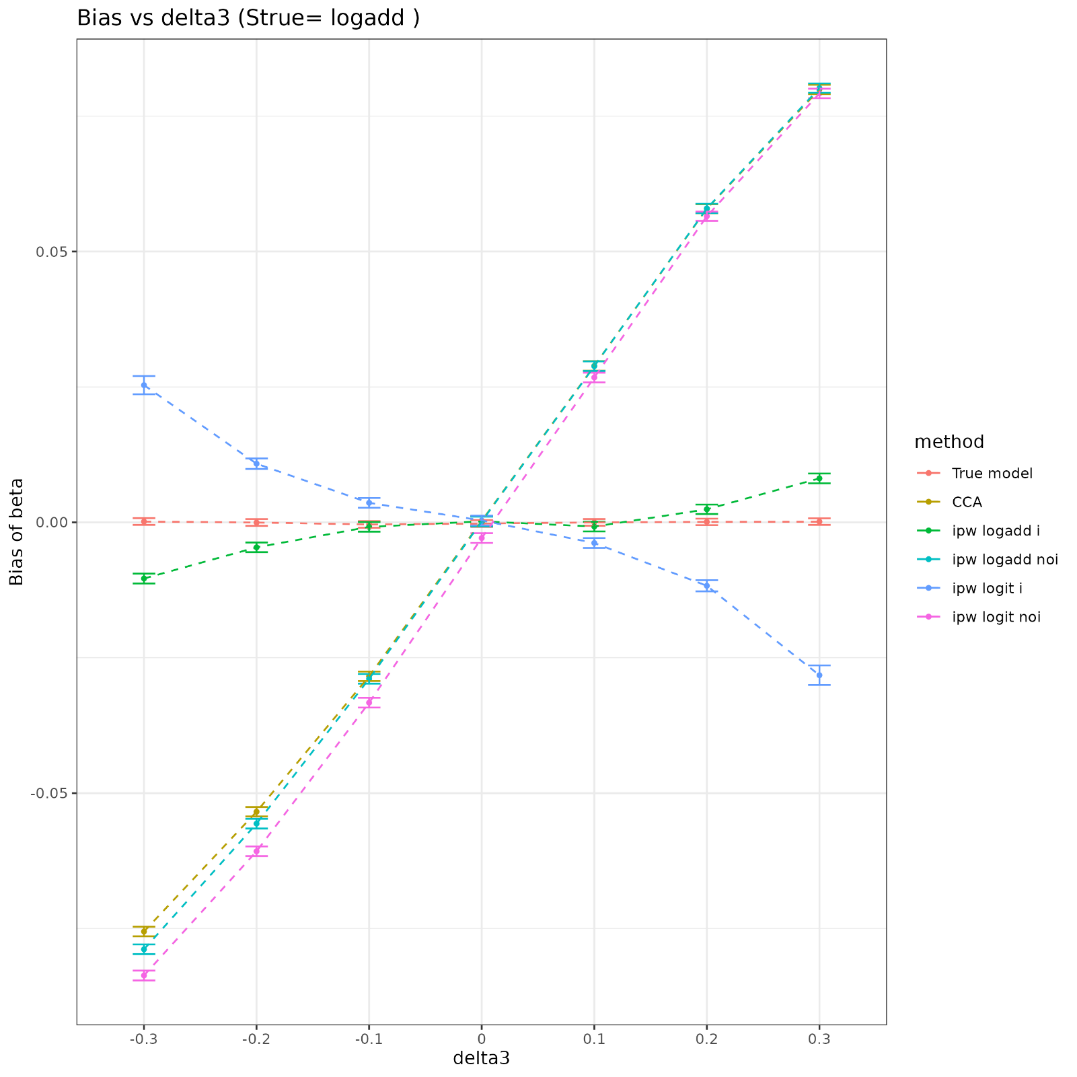

| delta3 | method | bias | coverage | EmpSE | ModSE | relative_precision | relative_error_ModSE | MSE | power |
| --- | --- | --- | --- | --- | --- | --- | --- | --- | --- |
| -0.3 | True model | 0.000146 ( 0.000318 ) | 0.941 ( 0.007451 ) | 0.010068 ( 0.000225 ) | 0.009758 ( 1e-06 ) | 0 ( 0 ) | -3.077896 ( 2.168344 ) | 0.000101 ( 5e-06 ) | 1 ( 0 ) |
| -0.3 | CCA | -0.075551 ( 0.000451 ) | 0 ( 0 ) | 0.014269 ( 0.000319 ) | 0.014149 ( 2e-06 ) | -50.214921 ( 2.175281 ) | -0.844242 ( 2.218338 ) | 0.005911 ( 6.9e-05 ) | 1 ( 0 ) |
| -0.3 | ipw logadd i | -0.010371 ( 0.000464 ) | 0.894 ( 0.009735 ) | 0.014671 ( 0.000328 ) | 0.014595 ( 3e-06 ) | -52.903918 ( 2.179916 ) | -0.515658 ( 2.225778 ) | 0.000323 ( 1.4e-05 ) | 1 ( 0 ) |
| -0.3 | ipw logadd noi | -0.078837 ( 0.000458 ) | 0 ( 0 ) | 0.014477 ( 0.000324 ) | 0.014485 ( 2e-06 ) | -51.637037 ( 2.163168 ) | 0.049584 ( 2.23836 ) | 0.006425 ( 7.4e-05 ) | 1 ( 0 ) |
| -0.3 | ipw logit i | 0.025331 ( 0.000855 ) | 0.786 ( 0.012969 ) | 0.027024 ( 0.000605 ) | 0.026664 ( 0.003598 ) | -86.119354 ( 0.815428 ) | -1.331999 ( 13.495404 ) | 0.001371 ( 0.00021 ) | 0.999 ( 0.000999 ) |
| -0.3 | ipw logit noi | -0.083665 ( 0.000463 ) | 0 ( 0 ) | 0.014629 ( 0.000327 ) | 0.014646 ( 3e-06 ) | -52.632874 ( 2.122953 ) | 0.117662 ( 2.23989 ) | 0.007214 ( 7.9e-05 ) | 1 ( 0 ) |
| -0.2 | True model | -3e-05 ( 0.000317 ) | 0.948 ( 0.007021 ) | 0.010028 ( 0.000224 ) | 0.009758 ( 1e-06 ) | 0 ( 0 ) | -2.69312 ( 2.176952 ) | 1e-04 ( 4e-06 ) | 1 ( 0 ) |
| -0.2 | CCA | -0.053429 ( 0.000441 ) | 0.032 ( 0.005566 ) | 0.013946 ( 0.000312 ) | 0.013999 ( 2e-06 ) | -48.296039 ( 2.331571 ) | 0.381958 ( 2.245772 ) | 0.003049 ( 4.7e-05 ) | 1 ( 0 ) |
| -0.2 | ipw logadd i | -0.004618 ( 0.000448 ) | 0.947 ( 0.007085 ) | 0.014171 ( 0.000317 ) | 0.014331 ( 3e-06 ) | -49.925928 ( 2.325371 ) | 1.129114 ( 2.262537 ) | 0.000222 ( 1e-05 ) | 1 ( 0 ) |
| -0.2 | ipw logadd noi | -0.055628 ( 0.000451 ) | 0.028 ( 0.005217 ) | 0.01425 ( 0.000319 ) | 0.014313 ( 3e-06 ) | -50.478853 ( 2.26765 ) | 0.443845 ( 2.247189 ) | 0.003297 ( 5e-05 ) | 1 ( 0 ) |
| -0.2 | ipw logit i | 0.010823 ( 0.000494 ) | 0.902 ( 0.009402 ) | 0.015622 ( 0.00035 ) | 0.015901 ( 3.6e-05 ) | -58.798397 ( 2.040003 ) | 1.7819 ( 2.288706 ) | 0.000361 ( 1.7e-05 ) | 1 ( 0 ) |
| -0.2 | ipw logit noi | -0.060738 ( 0.000456 ) | 0.012 ( 0.003443 ) | 0.014421 ( 0.000323 ) | 0.014486 ( 3e-06 ) | -51.650234 ( 2.218207 ) | 0.445944 ( 2.247245 ) | 0.003897 ( 5.5e-05 ) | 1 ( 0 ) |
| -0.1 | True model | -0.000392 ( 0.000311 ) | 0.946 ( 0.007147 ) | 0.009844 ( 0.00022 ) | 0.00976 ( 1e-06 ) | 0 ( 0 ) | -0.856261 ( 2.218046 ) | 9.7e-05 ( 4e-06 ) | 1 ( 0 ) |
| -0.1 | CCA | -0.028423 ( 0.000448 ) | 0.459 ( 0.015758 ) | 0.014154 ( 0.000317 ) | 0.013918 ( 2e-06 ) | -51.625935 ( 2.140509 ) | -1.663438 ( 2.20001 ) | 0.001008 ( 2.7e-05 ) | 1 ( 0 ) |
| -0.1 | ipw logadd i | -0.000836 ( 0.000453 ) | 0.952 ( 0.00676 ) | 0.014318 ( 0.00032 ) | 0.01413 ( 2e-06 ) | -52.73287 ( 2.129849 ) | -1.315956 ( 2.207813 ) | 0.000206 ( 9e-06 ) | 1 ( 0 ) |
| -0.1 | ipw logadd noi | -0.028896 ( 0.000455 ) | 0.47 ( 0.015783 ) | 0.014388 ( 0.000322 ) | 0.014159 ( 2e-06 ) | -53.187519 ( 2.103771 ) | -1.587855 ( 2.201731 ) | 0.001042 ( 2.8e-05 ) | 1 ( 0 ) |
| -0.1 | ipw logit i | 0.0036 ( 0.000462 ) | 0.935 ( 0.007796 ) | 0.014596 ( 0.000327 ) | 0.01441 ( 3e-06 ) | -54.513374 ( 2.05567 ) | -1.272334 ( 2.208815 ) | 0.000226 ( 1e-05 ) | 1 ( 0 ) |
| -0.1 | ipw logit noi | -0.033296 ( 0.00046 ) | 0.357 ( 0.015151 ) | 0.014534 ( 0.000325 ) | 0.014302 ( 3e-06 ) | -54.126949 ( 2.062954 ) | -1.599462 ( 2.201479 ) | 0.00132 ( 3.2e-05 ) | 1 ( 0 ) |
| 0 | True model | -0.000222 ( 0.000321 ) | 0.944 ( 0.007271 ) | 0.010154 ( 0.000227 ) | 0.009759 ( 1e-06 ) | 0 ( 0 ) | -3.889194 ( 2.150194 ) | 0.000103 ( 5e-06 ) | 1 ( 0 ) |
| 0 | CCA | 0.000275 ( 0.00045 ) | 0.941 ( 0.007451 ) | 0.014237 ( 0.000319 ) | 0.013882 ( 2e-06 ) | -49.138033 ( 2.258817 ) | -2.493889 ( 2.181431 ) | 0.000203 ( 1e-05 ) | 1 ( 0 ) |
| 0 | ipw logadd i | 0.000189 ( 0.000455 ) | 0.944 ( 0.007271 ) | 0.014385 ( 0.000322 ) | 0.014054 ( 2e-06 ) | -50.181366 ( 2.226712 ) | -2.301476 ( 2.185762 ) | 0.000207 ( 1e-05 ) | 1 ( 0 ) |
| 0 | ipw logadd noi | 0.000161 ( 0.000456 ) | 0.941 ( 0.007451 ) | 0.01442 ( 0.000323 ) | 0.014054 ( 2e-06 ) | -50.423571 ( 2.227196 ) | -2.538877 ( 2.180451 ) | 0.000208 ( 1e-05 ) | 1 ( 0 ) |
| 0 | ipw logit i | 0.000347 ( 0.000458 ) | 0.942 ( 0.007392 ) | 0.014478 ( 0.000324 ) | 0.014133 ( 2e-06 ) | -50.813644 ( 2.199202 ) | -2.379321 ( 2.184022 ) | 0.00021 ( 1e-05 ) | 1 ( 0 ) |
| 0 | ipw logit noi | -0.002899 ( 0.000459 ) | 0.939 ( 0.007568 ) | 0.014519 ( 0.000325 ) | 0.014148 ( 2e-06 ) | -51.094663 ( 2.198214 ) | -2.556499 ( 2.18006 ) | 0.000219 ( 1e-05 ) | 1 ( 0 ) |
| 0.1 | True model | -1.1e-05 ( 0.00031 ) | 0.955 ( 0.006556 ) | 0.009787 ( 0.000219 ) | 0.00976 ( 1e-06 ) | 0 ( 0 ) | -0.277763 ( 2.23099 ) | 9.6e-05 ( 4e-06 ) | 1 ( 0 ) |
| 0.1 | CCA | 0.028892 ( 0.000447 ) | 0.468 ( 0.015779 ) | 0.014136 ( 0.000316 ) | 0.013912 ( 2e-06 ) | -52.058871 ( 2.082439 ) | -1.580086 ( 2.201875 ) | 0.001034 ( 2.8e-05 ) | 1 ( 0 ) |
| 0.1 | ipw logadd i | -0.000777 ( 0.000456 ) | 0.962 ( 0.006046 ) | 0.014408 ( 0.000322 ) | 0.014195 ( 3e-06 ) | -53.854098 ( 2.0631 ) | -1.48134 ( 2.204123 ) | 0.000208 ( 9e-06 ) | 1 ( 0 ) |
| 0.1 | ipw logadd noi | 0.028832 ( 0.000451 ) | 0.478 ( 0.015796 ) | 0.014265 ( 0.000319 ) | 0.014041 ( 2e-06 ) | -52.923913 ( 2.066007 ) | -1.570728 ( 2.202103 ) | 0.001035 ( 2.8e-05 ) | 1 ( 0 ) |
| 0.1 | ipw logit i | -0.003811 ( 0.000465 ) | 0.953 ( 0.006693 ) | 0.014706 ( 0.000329 ) | 0.01451 ( 4e-06 ) | -55.70431 ( 2.007571 ) | -1.329412 ( 2.207627 ) | 0.000231 ( 9e-06 ) | 1 ( 0 ) |
| 0.1 | ipw logit noi | 0.026745 ( 0.000453 ) | 0.542 ( 0.015756 ) | 0.014333 ( 0.000321 ) | 0.014102 ( 2e-06 ) | -53.371827 ( 2.048033 ) | -1.614563 ( 2.201125 ) | 0.000921 ( 2.7e-05 ) | 1 ( 0 ) |
| 0.2 | True model | 8.1e-05 ( 0.00031 ) | 0.949 ( 0.006957 ) | 0.009805 ( 0.000219 ) | 0.009759 ( 1e-06 ) | 0 ( 0 ) | -0.469642 ( 2.226696 ) | 9.6e-05 ( 4e-06 ) | 1 ( 0 ) |
| 0.2 | CCA | 0.057906 ( 0.000432 ) | 0.005 ( 0.00223 ) | 0.013665 ( 0.000306 ) | 0.013984 ( 2e-06 ) | -48.512473 ( 2.330894 ) | 2.331776 ( 2.289393 ) | 0.00354 ( 5.2e-05 ) | 1 ( 0 ) |
| 0.2 | ipw logadd i | 0.002405 ( 0.000445 ) | 0.955 ( 0.006556 ) | 0.014063 ( 0.000315 ) | 0.014489 ( 4e-06 ) | -51.383517 ( 2.290536 ) | 3.028534 ( 2.305112 ) | 0.000203 ( 1e-05 ) | 1 ( 0 ) |
| 0.2 | ipw logadd noi | 0.057957 ( 0.000437 ) | 0.006 ( 0.002442 ) | 0.013808 ( 0.000309 ) | 0.01408 ( 2e-06 ) | -49.57304 ( 2.301781 ) | 1.971538 ( 2.281352 ) | 0.003549 ( 5.3e-05 ) | 1 ( 0 ) |
| 0.2 | ipw logit i | -0.011709 ( 0.000526 ) | 0.905 ( 0.009272 ) | 0.016644 ( 0.000372 ) | 0.016979 ( 0.000109 ) | -65.292853 ( 1.757226 ) | 2.014633 ( 2.374174 ) | 0.000414 ( 1.7e-05 ) | 1 ( 0 ) |
| 0.2 | ipw logit noi | 0.056517 ( 0.000438 ) | 0.011 ( 0.003298 ) | 0.013851 ( 0.00031 ) | 0.014121 ( 2e-06 ) | -49.886944 ( 2.287886 ) | 1.950811 ( 2.28089 ) | 0.003386 ( 5.2e-05 ) | 1 ( 0 ) |
| 0.3 | True model | 0.00013 ( 0.00031 ) | 0.952 ( 0.00676 ) | 0.009813 ( 0.00022 ) | 0.009758 ( 1e-06 ) | 0 ( 0 ) | -0.565252 ( 2.224558 ) | 9.6e-05 ( 4e-06 ) | 1 ( 0 ) |
| 0.3 | CCA | 0.07993 ( 0.000442 ) | 0 ( 0 ) | 0.013993 ( 0.000313 ) | 0.014103 ( 2e-06 ) | -50.813997 ( 2.241748 ) | 0.789375 ( 2.254888 ) | 0.006584 ( 7.2e-05 ) | 1 ( 0 ) |
| 0.3 | ipw logadd i | 0.008117 ( 0.000461 ) | 0.932 ( 0.007961 ) | 0.014585 ( 0.000326 ) | 0.014773 ( 5e-06 ) | -54.731241 ( 2.198359 ) | 1.283335 ( 2.266173 ) | 0.000278 ( 1.2e-05 ) | 1 ( 0 ) |
| 0.3 | ipw logadd noi | 0.08017 ( 0.000446 ) | 0 ( 0 ) | 0.01409 ( 0.000315 ) | 0.014178 ( 2e-06 ) | -51.489833 ( 2.221752 ) | 0.624248 ( 2.251212 ) | 0.006626 ( 7.2e-05 ) | 1 ( 0 ) |
| 0.3 | ipw logit i | -0.028223 ( 0.000924 ) | 0.794 ( 0.012789 ) | 0.029229 ( 0.000654 ) | 0.028895 ( 0.001426 ) | -88.727679 ( 0.678378 ) | -1.140563 ( 5.356592 ) | 0.00165 ( 0.000125 ) | 1 ( 0 ) |
| 0.3 | ipw logit noi | 0.079194 ( 0.000446 ) | 0 ( 0 ) | 0.014119 ( 0.000316 ) | 0.014206 ( 2e-06 ) | -51.693942 ( 2.213498 ) | 0.609916 ( 2.250893 ) | 0.006471 ( 7.2e-05 ) | 1 ( 0 ) |

##### 1.2.2 Y is binary

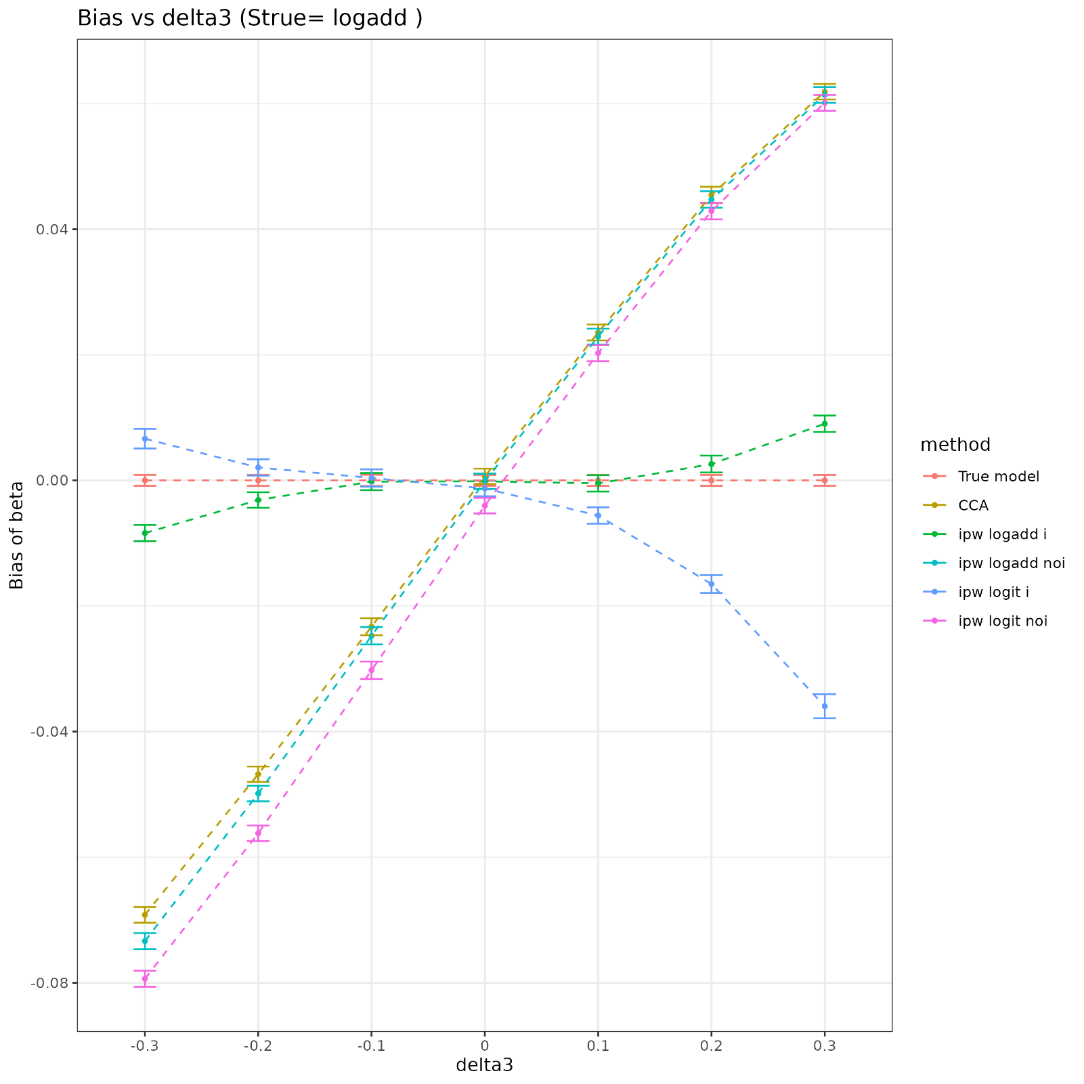

| delta3 | method | bias | coverage | EmpSE | ModSE | relative_precision | relative_error_ModSE | MSE | power |
| --- | --- | --- | --- | --- | --- | --- | --- | --- | --- |
| -0.3 | True model | 0 ( 0.000445 ) | 0.95 ( 0.006892 ) | 0.014063 ( 0.000315 ) | 0.01425 ( 1e-06 ) | 0 ( 0 ) | 1.326802 ( 2.266878 ) | 0.000198 ( 9e-06 ) | 1 ( 0 ) |
| -0.3 | CCA | -0.069181 ( 0.000643 ) | 0.088 ( 0.008959 ) | 0.02033 ( 0.000455 ) | 0.020582 ( 2e-06 ) | -52.149555 ( 2.09845 ) | 1.238448 ( 2.26492 ) | 0.005199 ( 9.1e-05 ) | 1 ( 0 ) |
| -0.3 | ipw logadd i | -0.008393 ( 0.000664 ) | 0.931 ( 0.008015 ) | 0.021001 ( 0.00047 ) | 0.021188 ( 2e-06 ) | -55.157824 ( 2.027117 ) | 0.891231 ( 2.257156 ) | 0.000511 ( 2.2e-05 ) | 1 ( 0 ) |
| -0.3 | ipw logadd noi | -0.073363 ( 0.000651 ) | 0.067 ( 0.007906 ) | 0.020584 ( 0.000461 ) | 0.02084 ( 2e-06 ) | -53.324031 ( 2.094031 ) | 1.243556 ( 2.265037 ) | 0.005805 ( 9.8e-05 ) | 1 ( 0 ) |
| -0.3 | ipw logit i | 0.006635 ( 0.000795 ) | 0.941 ( 0.007451 ) | 0.025149 ( 0.000563 ) | 0.025332 ( 0.000427 ) | -68.728431 ( 1.599333 ) | 0.728233 ( 2.822447 ) | 0.000676 ( 3.8e-05 ) | 1 ( 0 ) |
| -0.3 | ipw logit noi | -0.07936 ( 0.000652 ) | 0.029 ( 0.005307 ) | 0.02062 ( 0.000461 ) | 0.020887 ( 2e-06 ) | -53.483339 ( 2.091606 ) | 1.296085 ( 2.266214 ) | 0.006723 ( 0.000105 ) | 1 ( 0 ) |
| -0.2 | True model | 0 ( 0.000447 ) | 0.947 ( 0.007085 ) | 0.014145 ( 0.000316 ) | 0.014249 ( 1e-06 ) | 0 ( 0 ) | 0.731612 ( 2.253563 ) | 2e-04 ( 9e-06 ) | 1 ( 0 ) |
| -0.2 | CCA | -0.046792 ( 0.000627 ) | 0.364 ( 0.015215 ) | 0.019833 ( 0.000444 ) | 0.020466 ( 2e-06 ) | -49.129266 ( 2.362052 ) | 3.194649 ( 2.308683 ) | 0.002582 ( 6.1e-05 ) | 1 ( 0 ) |
| -0.2 | ipw logadd i | -0.00313 ( 0.000635 ) | 0.955 ( 0.006556 ) | 0.020089 ( 0.000449 ) | 0.020856 ( 2e-06 ) | -50.416797 ( 2.346478 ) | 3.820991 ( 2.322696 ) | 0.000413 ( 1.8e-05 ) | 1 ( 0 ) |
| -0.2 | ipw logadd noi | -0.049863 ( 0.000632 ) | 0.324 ( 0.014799 ) | 0.019974 ( 0.000447 ) | 0.020733 ( 2e-06 ) | -49.846468 ( 2.365406 ) | 3.798446 ( 2.322193 ) | 0.002885 ( 6.6e-05 ) | 1 ( 0 ) |
| -0.2 | ipw logit i | 0.002057 ( 0.000658 ) | 0.959 ( 0.00627 ) | 0.020803 ( 0.000465 ) | 0.021492 ( 3.6e-05 ) | -53.765663 ( 2.23417 ) | 3.308601 ( 2.317639 ) | 0.000437 ( 1.9e-05 ) | 1 ( 0 ) |
| -0.2 | ipw logit noi | -0.056196 ( 0.000633 ) | 0.233 ( 0.013368 ) | 0.020029 ( 0.000448 ) | 0.020785 ( 2e-06 ) | -50.122177 ( 2.357952 ) | 3.772234 ( 2.321608 ) | 0.003559 ( 7.4e-05 ) | 1 ( 0 ) |
| -0.1 | True model | 0 ( 0.000456 ) | 0.942 ( 0.007392 ) | 0.014407 ( 0.000322 ) | 0.014249 ( 1e-06 ) | 0 ( 0 ) | -1.093843 ( 2.212724 ) | 0.000207 ( 1e-05 ) | 1 ( 0 ) |
| -0.1 | CCA | -0.023305 ( 0.00069 ) | 0.766 ( 0.013388 ) | 0.021814 ( 0.000488 ) | 0.020438 ( 2e-06 ) | -56.383985 ( 1.941184 ) | -6.309333 ( 2.09606 ) | 0.001019 ( 3.9e-05 ) | 1 ( 0 ) |
| -0.1 | ipw logadd i | -0.000193 ( 0.000698 ) | 0.94 ( 0.00751 ) | 0.022086 ( 0.000494 ) | 0.020666 ( 2e-06 ) | -57.45128 ( 1.895514 ) | -6.429639 ( 2.09337 ) | 0.000487 ( 2.1e-05 ) | 1 ( 0 ) |
| -0.1 | ipw logadd noi | -0.024744 ( 0.000701 ) | 0.752 ( 0.013656 ) | 0.022161 ( 0.000496 ) | 0.02068 ( 2e-06 ) | -57.735883 ( 1.887276 ) | -6.681896 ( 2.087727 ) | 0.001103 ( 4.2e-05 ) | 1 ( 0 ) |
| -0.1 | ipw logit i | 0.000381 ( 0.000701 ) | 0.941 ( 0.007451 ) | 0.022165 ( 0.000496 ) | 0.020751 ( 2e-06 ) | -57.752477 ( 1.884909 ) | -6.3779 ( 2.094528 ) | 0.000491 ( 2.2e-05 ) | 1 ( 0 ) |
| -0.1 | ipw logit noi | -0.030239 ( 0.000703 ) | 0.674 ( 0.014823 ) | 0.022216 ( 0.000497 ) | 0.020722 ( 2e-06 ) | -57.946372 ( 1.878846 ) | -6.722801 ( 2.086813 ) | 0.001407 ( 4.9e-05 ) | 1 ( 0 ) |
| 0 | True model | 0 ( 0.000444 ) | 0.953 ( 0.006693 ) | 0.014039 ( 0.000314 ) | 0.01425 ( 1e-06 ) | 0 ( 0 ) | 1.50209 ( 2.2708 ) | 0.000197 ( 9e-06 ) | 1 ( 0 ) |
| 0 | CCA | 0.000644 ( 0.000629 ) | 0.949 ( 0.006957 ) | 0.019894 ( 0.000445 ) | 0.020441 ( 2e-06 ) | -50.197343 ( 2.298621 ) | 2.749959 ( 2.298737 ) | 0.000396 ( 1.7e-05 ) | 1 ( 0 ) |
| 0 | ipw logadd i | -0.000148 ( 0.000627 ) | 0.953 ( 0.006693 ) | 0.019836 ( 0.000444 ) | 0.020628 ( 2e-06 ) | -49.907039 ( 2.339207 ) | 3.988952 ( 2.326457 ) | 0.000393 ( 1.7e-05 ) | 1 ( 0 ) |
| 0 | ipw logadd noi | -0.000162 ( 0.000629 ) | 0.954 ( 0.006624 ) | 0.019903 ( 0.000445 ) | 0.020628 ( 2e-06 ) | -50.244001 ( 2.321516 ) | 3.63832 ( 2.318613 ) | 0.000396 ( 1.8e-05 ) | 1 ( 0 ) |
| 0 | ipw logit i | -0.001291 ( 0.000628 ) | 0.956 ( 0.006486 ) | 0.019844 ( 0.000444 ) | 0.020639 ( 2e-06 ) | -49.946115 ( 2.338176 ) | 4.005916 ( 2.326837 ) | 0.000395 ( 1.8e-05 ) | 1 ( 0 ) |
| 0 | ipw logit noi | -0.004025 ( 0.00063 ) | 0.947 ( 0.007085 ) | 0.019926 ( 0.000446 ) | 0.020653 ( 2e-06 ) | -50.356679 ( 2.31852 ) | 3.649609 ( 2.318866 ) | 0.000413 ( 1.9e-05 ) | 1 ( 0 ) |
| 0.1 | True model | 0 ( 0.000451 ) | 0.943 ( 0.007332 ) | 0.014254 ( 0.000319 ) | 0.014249 ( 1e-06 ) | 0 ( 0 ) | -0.037078 ( 2.236365 ) | 0.000203 ( 1e-05 ) | 1 ( 0 ) |
| 0.1 | CCA | 0.023556 ( 0.000651 ) | 0.785 ( 0.012991 ) | 0.020575 ( 0.00046 ) | 0.020454 ( 2e-06 ) | -52.001887 ( 2.10732 ) | -0.590242 ( 2.224009 ) | 0.000978 ( 3.6e-05 ) | 1 ( 0 ) |
| 0.1 | ipw logadd i | -0.000478 ( 0.000664 ) | 0.946 ( 0.007147 ) | 0.021008 ( 0.00047 ) | 0.020805 ( 2e-06 ) | -53.961622 ( 2.05232 ) | -0.969758 ( 2.215523 ) | 0.000441 ( 2e-05 ) | 1 ( 0 ) |
| 0.1 | ipw logadd noi | 0.022897 ( 0.000655 ) | 0.799 ( 0.012673 ) | 0.020704 ( 0.000463 ) | 0.020595 ( 2e-06 ) | -52.598417 ( 2.09725 ) | -0.52686 ( 2.225429 ) | 0.000953 ( 3.6e-05 ) | 1 ( 0 ) |
| 0.1 | ipw logit i | -0.00559 ( 0.000669 ) | 0.932 ( 0.007961 ) | 0.021166 ( 0.000474 ) | 0.020941 ( 3e-06 ) | -54.646214 ( 2.030266 ) | -1.063978 ( 2.213425 ) | 0.000479 ( 2.1e-05 ) | 1 ( 0 ) |
| 0.1 | ipw logit noi | 0.020252 ( 0.000655 ) | 0.829 ( 0.011906 ) | 0.020715 ( 0.000463 ) | 0.02061 ( 2e-06 ) | -52.650914 ( 2.095314 ) | -0.50916 ( 2.225825 ) | 0.000839 ( 3.3e-05 ) | 1 ( 0 ) |
| 0.2 | True model | 0 ( 0.000447 ) | 0.948 ( 0.007021 ) | 0.01415 ( 0.000317 ) | 0.014249 ( 1e-06 ) | 0 ( 0 ) | 0.702921 ( 2.252921 ) | 2e-04 ( 9e-06 ) | 1 ( 0 ) |
| 0.2 | CCA | 0.045452 ( 0.000666 ) | 0.399 ( 0.015485 ) | 0.021071 ( 0.000471 ) | 0.02049 ( 2e-06 ) | -54.90298 ( 2.060206 ) | -2.755007 ( 2.175578 ) | 0.002509 ( 6.4e-05 ) | 1 ( 0 ) |
| 0.2 | ipw logadd i | 0.002598 ( 0.000683 ) | 0.942 ( 0.007392 ) | 0.021614 ( 0.000484 ) | 0.021162 ( 3e-06 ) | -57.141725 ( 2.023645 ) | -2.090443 ( 2.190457 ) | 0.000473 ( 2.2e-05 ) | 1 ( 0 ) |
| 0.2 | ipw logadd noi | 0.044722 ( 0.000669 ) | 0.415 ( 0.015581 ) | 0.021144 ( 0.000473 ) | 0.020599 ( 2e-06 ) | -55.213966 ( 2.062679 ) | -2.577475 ( 2.17955 ) | 0.002447 ( 6.3e-05 ) | 1 ( 0 ) |
| 0.2 | ipw logit i | -0.016502 ( 0.000723 ) | 0.889 ( 0.009934 ) | 0.022879 ( 0.000512 ) | 0.022718 ( 0.00017 ) | -61.74934 ( 1.900272 ) | -0.70462 ( 2.342511 ) | 0.000795 ( 3.3e-05 ) | 1 ( 0 ) |
| 0.2 | ipw logit noi | 0.042888 ( 0.000669 ) | 0.458 ( 0.015756 ) | 0.021151 ( 0.000473 ) | 0.020608 ( 2e-06 ) | -55.246707 ( 2.062512 ) | -2.570079 ( 2.179716 ) | 0.002286 ( 6.1e-05 ) | 1 ( 0 ) |
| 0.3 | True model | 0 ( 0.000437 ) | 0.951 ( 0.006826 ) | 0.013822 ( 0.000309 ) | 0.01425 ( 1e-06 ) | 0 ( 0 ) | 3.090203 ( 2.306328 ) | 0.000191 ( 9e-06 ) | 1 ( 0 ) |
| 0.3 | CCA | 0.061887 ( 0.000633 ) | 0.157 ( 0.011504 ) | 0.020031 ( 0.000448 ) | 0.020611 ( 2e-06 ) | -52.383943 ( 2.20954 ) | 2.8959 ( 2.301999 ) | 0.004231 ( 8.1e-05 ) | 1 ( 0 ) |
| 0.3 | ipw logadd i | 0.00902 ( 0.000668 ) | 0.944 ( 0.007271 ) | 0.021126 ( 0.000473 ) | 0.021582 ( 3e-06 ) | -57.190001 ( 2.05411 ) | 2.16108 ( 2.28558 ) | 0.000527 ( 2.3e-05 ) | 1 ( 0 ) |
| 0.3 | ipw logadd noi | 0.061381 ( 0.000638 ) | 0.161 ( 0.011622 ) | 0.020162 ( 0.000451 ) | 0.020696 ( 2e-06 ) | -52.998019 ( 2.182322 ) | 2.650494 ( 2.296509 ) | 0.004174 ( 8.1e-05 ) | 1 ( 0 ) |
| 0.3 | ipw logit i | -0.035963 ( 0.000976 ) | 0.787 ( 0.012947 ) | 0.030859 ( 0.00069 ) | 0.030711 ( 0.000688 ) | -79.936589 ( 1.133813 ) | -0.480405 ( 3.150316 ) | 0.002245 ( 0.000113 ) | 1 ( 0 ) |
| 0.3 | ipw logit noi | 0.060117 ( 0.000638 ) | 0.18 ( 0.012149 ) | 0.02017 ( 0.000451 ) | 0.020702 ( 2e-06 ) | -53.037936 ( 2.180842 ) | 2.63433 ( 2.296147 ) | 0.004021 ( 7.9e-05 ) | 1 ( 0 ) |

### 2 DAG3 (only vary delta3)

X

C

S

Y

#### 2.1 Strue=logit

##### 2.1.1 Y is continuous

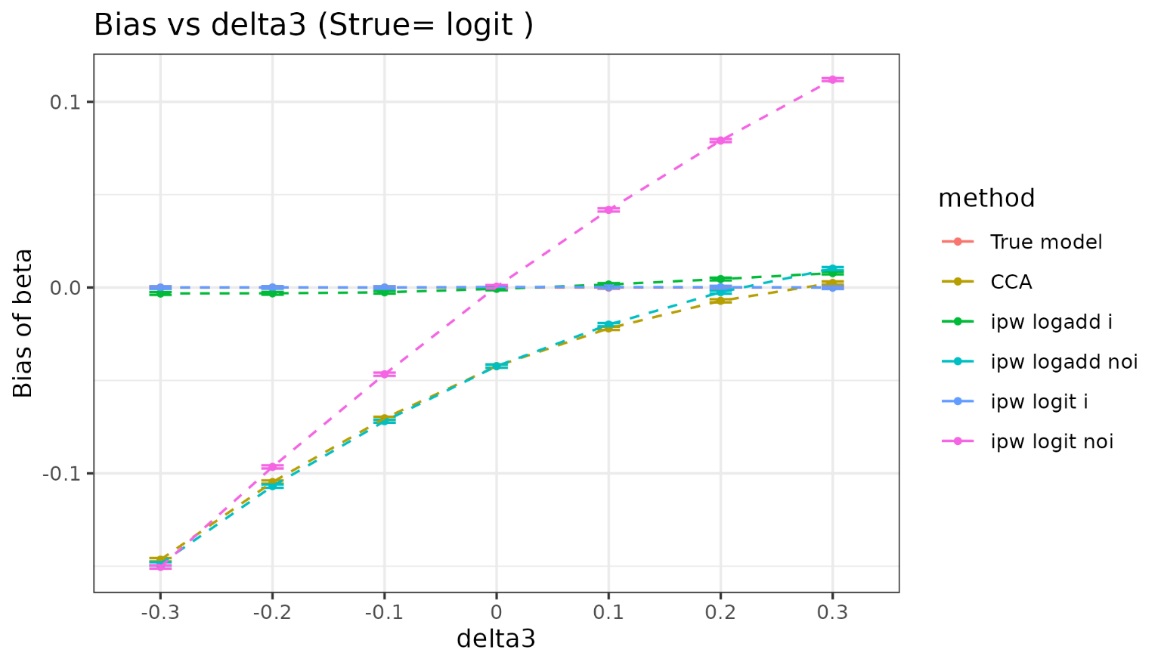

| delta3 | method | bias | coverage | EmpSE | ModSE | relative_precision | relative_error_ModSE | MSE | power |
| --- | --- | --- | --- | --- | --- | --- | --- | --- | --- |
| -0.3 | True model | 0 ( 0.000307 ) | 0.952 ( 0.00676 ) | 0.009715 ( 0.000217 ) | 0.00976 ( 1e-06 ) | 0 ( 0 ) | 0.462243 ( 2.247543 ) | 9.4e-05 ( 4e-06 ) | 1 ( 0 ) |
| -0.3 | CCA | -0.146504 ( 0.000452 ) | 0 ( 0 ) | 0.014285 ( 0.00032 ) | 0.013753 ( 2e-06 ) | -53.752218 ( 2.071993 ) | -3.725402 ( 2.153877 ) | 0.021667 ( 0.000133 ) | 1 ( 0 ) |
| -0.3 | ipw logadd i | -0.003194 ( 0.000386 ) | 0.967 ( 0.005649 ) | 0.012196 ( 0.000273 ) | 0.013848 ( 2e-06 ) | -36.553349 ( 2.346521 ) | 13.54134 ( 2.540192 ) | 0.000159 ( 7e-06 ) | 1 ( 0 ) |
| -0.3 | ipw logadd noi | -0.148739 ( 0.000455 ) | 0 ( 0 ) | 0.014396 ( 0.000322 ) | 0.01389 ( 2e-06 ) | -54.463436 ( 2.051229 ) | -3.513991 ( 2.158628 ) | 0.02233 ( 0.000136 ) | 1 ( 0 ) |
| -0.3 | ipw logit i | -3.2e-05 ( 0.000385 ) | 0.979 ( 0.004534 ) | 0.012183 ( 0.000273 ) | 0.013901 ( 2e-06 ) | -36.417158 ( 2.355904 ) | 14.09921 ( 2.552673 ) | 0.000148 ( 6e-06 ) | 1 ( 0 ) |
| -0.3 | ipw logit noi | -0.15039 ( 0.000458 ) | 0 ( 0 ) | 0.014494 ( 0.000324 ) | 0.013971 ( 2e-06 ) | -55.078223 ( 2.026812 ) | -3.608351 ( 2.15652 ) | 0.022827 ( 0.000138 ) | 1 ( 0 ) |
| -0.2 | True model | 0 ( 0.000308 ) | 0.951 ( 0.006826 ) | 0.009733 ( 0.000218 ) | 0.00976 ( 1e-06 ) | 0 ( 0 ) | 0.280413 ( 2.243476 ) | 9.5e-05 ( 4e-06 ) | 1 ( 0 ) |
| -0.2 | CCA | -0.1047 ( 0.000432 ) | 0 ( 0 ) | 0.013664 ( 0.000306 ) | 0.013499 ( 2e-06 ) | -49.262403 ( 2.217571 ) | -1.206653 ( 2.210228 ) | 0.011149 ( 9.1e-05 ) | 1 ( 0 ) |
| -0.2 | ipw logadd i | -0.003105 ( 0.000373 ) | 0.975 ( 0.004937 ) | 0.011808 ( 0.000264 ) | 0.013609 ( 2e-06 ) | -32.05819 ( 2.415835 ) | 15.258494 ( 2.57861 ) | 0.000149 ( 7e-06 ) | 1 ( 0 ) |
| -0.2 | ipw logadd noi | -0.106918 ( 0.000436 ) | 0 ( 0 ) | 0.013791 ( 0.000309 ) | 0.013661 ( 2e-06 ) | -50.19139 ( 2.217607 ) | -0.940007 ( 2.216215 ) | 0.011621 ( 9.4e-05 ) | 1 ( 0 ) |
| -0.2 | ipw logit i | 5e-05 ( 0.000373 ) | 0.98 ( 0.004427 ) | 0.011809 ( 0.000264 ) | 0.013677 ( 2e-06 ) | -32.071426 ( 2.419795 ) | 15.822275 ( 2.591224 ) | 0.000139 ( 6e-06 ) | 1 ( 0 ) |
| -0.2 | ipw logit noi | -0.096562 ( 0.00044 ) | 0 ( 0 ) | 0.01391 ( 0.000311 ) | 0.013757 ( 2e-06 ) | -51.041626 ( 2.178013 ) | -1.095484 ( 2.21274 ) | 0.009518 ( 8.5e-05 ) | 1 ( 0 ) |
| -0.1 | True model | 0 ( 0.000311 ) | 0.942 ( 0.007392 ) | 0.00983 ( 0.00022 ) | 0.00976 ( 1e-06 ) | 0 ( 0 ) | -0.710937 ( 2.221297 ) | 9.7e-05 ( 4e-06 ) | 1 ( 0 ) |
| -0.1 | CCA | -0.070396 ( 0.000425 ) | 0 ( 0 ) | 0.013437 ( 0.000301 ) | 0.013283 ( 2e-06 ) | -46.478445 ( 2.258948 ) | -1.142619 ( 2.211656 ) | 0.005136 ( 6e-05 ) | 1 ( 0 ) |
| -0.1 | ipw logadd i | -0.002551 ( 0.000375 ) | 0.975 ( 0.004937 ) | 0.011864 ( 0.000265 ) | 0.013425 ( 2e-06 ) | -31.348173 ( 2.368012 ) | 13.153041 ( 2.531497 ) | 0.000147 ( 6e-06 ) | 1 ( 0 ) |
| -0.1 | ipw logadd noi | -0.071933 ( 0.00043 ) | 0 ( 0 ) | 0.013602 ( 0.000304 ) | 0.013467 ( 2e-06 ) | -47.767174 ( 2.268233 ) | -0.989526 ( 2.215098 ) | 0.005359 ( 6.2e-05 ) | 1 ( 0 ) |
| -0.1 | ipw logit i | -1e-05 ( 0.000375 ) | 0.98 ( 0.004427 ) | 0.011856 ( 0.000265 ) | 0.013528 ( 2e-06 ) | -31.252045 ( 2.371441 ) | 14.105614 ( 2.552811 ) | 0.00014 ( 6e-06 ) | 1 ( 0 ) |
| -0.1 | ipw logit noi | -0.046658 ( 0.000433 ) | 0.086 ( 0.008866 ) | 0.013678 ( 0.000306 ) | 0.01358 ( 2e-06 ) | -48.347168 ( 2.236292 ) | -0.716234 ( 2.221215 ) | 0.002364 ( 4e-05 ) | 1 ( 0 ) |
| 0 | True model | 0 ( 0.000319 ) | 0.944 ( 0.007271 ) | 0.010073 ( 0.000225 ) | 0.009759 ( 1e-06 ) | 0 ( 0 ) | -3.115756 ( 2.167496 ) | 0.000101 ( 5e-06 ) | 1 ( 0 ) |
| 0 | CCA | -0.042273 ( 0.000433 ) | 0.109 ( 0.009855 ) | 0.013696 ( 0.000306 ) | 0.0131 ( 2e-06 ) | -45.905523 ( 2.190305 ) | -4.353022 ( 2.139835 ) | 0.001974 ( 3.7e-05 ) | 1 ( 0 ) |
| 0 | ipw logadd i | -0.000679 ( 0.000383 ) | 0.963 ( 0.005969 ) | 0.012108 ( 0.000271 ) | 0.013273 ( 2e-06 ) | -30.783085 ( 2.283148 ) | 9.623781 ( 2.452543 ) | 0.000147 ( 7e-06 ) | 1 ( 0 ) |
| 0 | ipw logadd noi | -0.04231 ( 0.000435 ) | 0.113 ( 0.010012 ) | 0.013758 ( 0.000308 ) | 0.0133 ( 2e-06 ) | -46.389518 ( 2.244896 ) | -3.324782 ( 2.162855 ) | 0.001979 ( 3.8e-05 ) | 1 ( 0 ) |
| 0 | ipw logit i | 0.000321 ( 0.000383 ) | 0.97 ( 0.005394 ) | 0.012104 ( 0.000271 ) | 0.013433 ( 2e-06 ) | -30.740248 ( 2.292027 ) | 10.979161 ( 2.482872 ) | 0.000146 ( 7e-06 ) | 1 ( 0 ) |
| 0 | ipw logit noi | 0.000469 ( 0.000439 ) | 0.942 ( 0.007392 ) | 0.013879 ( 0.00031 ) | 0.013432 ( 2e-06 ) | -47.320583 ( 2.178802 ) | -3.215614 ( 2.1653 ) | 0.000193 ( 9e-06 ) | 1 ( 0 ) |
| 0.1 | True model | 0 ( 0.000325 ) | 0.946 ( 0.007147 ) | 0.010266 ( 0.00023 ) | 0.009759 ( 1e-06 ) | 0 ( 0 ) | -4.936156 ( 2.126772 ) | 0.000105 ( 4e-06 ) | 1 ( 0 ) |
| 0.1 | CCA | -0.021978 ( 0.000417 ) | 0.608 ( 0.015438 ) | 0.013201 ( 0.000295 ) | 0.012945 ( 2e-06 ) | -39.522299 ( 2.487803 ) | -1.93703 ( 2.193883 ) | 0.000657 ( 2e-05 ) | 1 ( 0 ) |
| 0.1 | ipw logadd i | 0.001624 ( 0.00038 ) | 0.97 ( 0.005394 ) | 0.012023 ( 0.000269 ) | 0.013147 ( 2e-06 ) | -27.085845 ( 2.370364 ) | 9.35216 ( 2.446463 ) | 0.000147 ( 7e-06 ) | 1 ( 0 ) |
| 0.1 | ipw logadd noi | -0.019942 ( 0.000426 ) | 0.676 ( 0.014799 ) | 0.013471 ( 0.000301 ) | 0.013161 ( 2e-06 ) | -41.921613 ( 2.457157 ) | -2.303578 ( 2.185698 ) | 0.000579 ( 1.9e-05 ) | 1 ( 0 ) |
| 0.1 | ipw logit i | 0.000273 ( 0.000381 ) | 0.973 ( 0.005126 ) | 0.01205 ( 0.00027 ) | 0.013379 ( 2e-06 ) | -27.411476 ( 2.369179 ) | 11.031072 ( 2.484034 ) | 0.000145 ( 6e-06 ) | 1 ( 0 ) |
| 0.1 | ipw logit noi | 0.041796 ( 0.000429 ) | 0.121 ( 0.010313 ) | 0.013581 ( 0.000304 ) | 0.013316 ( 2e-06 ) | -42.855706 ( 2.386709 ) | -1.951871 ( 2.19357 ) | 0.001931 ( 3.7e-05 ) | 1 ( 0 ) |
| 0.2 | True model | 0 ( 0.000318 ) | 0.945 ( 0.007209 ) | 0.010041 ( 0.000225 ) | 0.00976 ( 1e-06 ) | 0 ( 0 ) | -2.79585 ( 2.174653 ) | 0.000101 ( 5e-06 ) | 1 ( 0 ) |
| 0.2 | CCA | -0.007128 ( 0.000425 ) | 0.899 ( 0.009529 ) | 0.013442 ( 0.000301 ) | 0.01282 ( 1e-06 ) | -44.200212 ( 2.279419 ) | -4.628297 ( 2.133672 ) | 0.000231 ( 1e-05 ) | 1 ( 0 ) |
| 0.2 | ipw logadd i | 0.004589 ( 0.000383 ) | 0.952 ( 0.00676 ) | 0.01211 ( 0.000271 ) | 0.013044 ( 2e-06 ) | -31.247862 ( 2.278597 ) | 7.711918 ( 2.409762 ) | 0.000168 ( 8e-06 ) | 1 ( 0 ) |
| 0.2 | ipw logadd noi | -0.002474 ( 0.000432 ) | 0.932 ( 0.007961 ) | 0.013662 ( 0.000306 ) | 0.013048 ( 2e-06 ) | -45.980683 ( 2.285404 ) | -4.494956 ( 2.136667 ) | 0.000193 ( 9e-06 ) | 1 ( 0 ) |
| 0.2 | ipw logit i | 0.000177 ( 0.000384 ) | 0.967 ( 0.005649 ) | 0.012129 ( 0.000271 ) | 0.013362 ( 2e-06 ) | -31.468413 ( 2.29064 ) | 10.166932 ( 2.464702 ) | 0.000147 ( 7e-06 ) | 1 ( 0 ) |
| 0.2 | ipw logit noi | 0.079148 ( 0.000429 ) | 0 ( 0 ) | 0.013568 ( 0.000304 ) | 0.013231 ( 2e-06 ) | -45.232036 ( 2.257614 ) | -2.485649 ( 2.181622 ) | 0.006448 ( 6.9e-05 ) | 1 ( 0 ) |
| 0.3 | True model | 0 ( 0.00032 ) | 0.944 ( 0.007271 ) | 0.010127 ( 0.000227 ) | 0.009758 ( 1e-06 ) | 0 ( 0 ) | -3.645512 ( 2.155644 ) | 0.000102 ( 5e-06 ) | 1 ( 0 ) |
| 0.3 | CCA | 0.002481 ( 0.00042 ) | 0.934 ( 0.007851 ) | 0.013276 ( 0.000297 ) | 0.012712 ( 1e-06 ) | -41.813508 ( 2.269529 ) | -4.252798 ( 2.142072 ) | 0.000182 ( 8e-06 ) | 1 ( 0 ) |
| 0.3 | ipw logadd i | 0.007804 ( 0.00038 ) | 0.936 ( 0.00774 ) | 0.012026 ( 0.000269 ) | 0.012949 ( 2e-06 ) | -29.089127 ( 2.232136 ) | 7.675578 ( 2.408948 ) | 0.000205 ( 8e-06 ) | 1 ( 0 ) |
| 0.3 | ipw logadd noi | 0.01021 ( 0.000423 ) | 0.877 ( 0.010386 ) | 0.013364 ( 0.000299 ) | 0.012948 ( 2e-06 ) | -42.572751 ( 2.316725 ) | -3.109913 ( 2.167652 ) | 0.000283 ( 1.1e-05 ) | 1 ( 0 ) |
| 0.3 | ipw logit i | -2.2e-05 ( 0.000382 ) | 0.971 ( 0.005307 ) | 0.012064 ( 0.00027 ) | 0.013364 ( 2e-06 ) | -29.535264 ( 2.232301 ) | 10.774894 ( 2.478313 ) | 0.000145 ( 6e-06 ) | 1 ( 0 ) |
| 0.3 | ipw logit noi | 0.111955 ( 0.000422 ) | 0 ( 0 ) | 0.01333 ( 0.000298 ) | 0.013164 ( 2e-06 ) | -42.28467 ( 2.299047 ) | -1.248518 ( 2.2093 ) | 0.012711 ( 9.4e-05 ) | 1 ( 0 ) |

##### 2.1.2 Y is binary

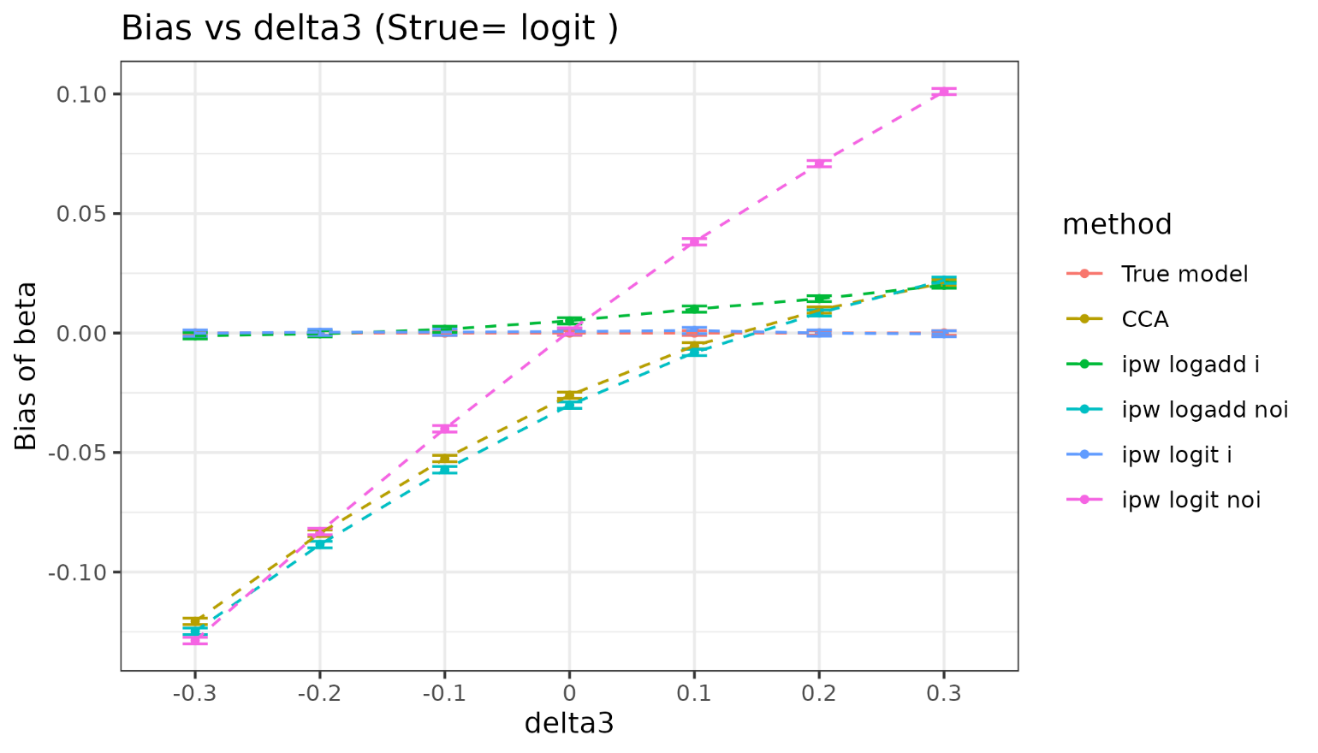

| delta3 | method | bias | coverage | EmpSE | ModSE | relative_precision | relative_error_ModSE | MSE | power |
| --- | --- | --- | --- | --- | --- | --- | --- | --- | --- |
| -0.3 | True model | 0 ( 0.000507 ) | 0.95 ( 0.006892 ) | 0.016037 ( 0.000359 ) | 0.015739 ( 2e-06 ) | 0 ( 0 ) | -1.858385 ( 2.195635 ) | 0.000257 ( 1.1e-05 ) | 1 ( 0 ) |
| -0.3 | CCA | -0.1206 ( 0.000694 ) | 0 ( 0 ) | 0.021947 ( 0.000491 ) | 0.022194 ( 4e-06 ) | -46.606108 ( 2.402323 ) | 1.123326 ( 2.262375 ) | 0.015025 ( 0.000167 ) | 1 ( 0 ) |
| -0.3 | ipw logadd i | -0.001179 ( 0.000672 ) | 0.962 ( 0.006046 ) | 0.021237 ( 0.000475 ) | 0.022241 ( 4e-06 ) | -42.973263 ( 2.476918 ) | 4.730501 ( 2.343076 ) | 0.000452 ( 2e-05 ) | 1 ( 0 ) |
| -0.3 | ipw logadd noi | -0.124765 ( 0.000693 ) | 0 ( 0 ) | 0.021927 ( 0.000491 ) | 0.022293 ( 4e-06 ) | -46.508828 ( 2.425436 ) | 1.665837 ( 2.274514 ) | 0.016047 ( 0.000173 ) | 1 ( 0 ) |
| -0.3 | ipw logit i | -8.4e-05 ( 0.000671 ) | 0.961 ( 0.006122 ) | 0.021221 ( 0.000475 ) | 0.022243 ( 4e-06 ) | -42.888065 ( 2.480813 ) | 4.815643 ( 2.344981 ) | 0.00045 ( 2e-05 ) | 1 ( 0 ) |
| -0.3 | ipw logit noi | -0.128635 ( 0.000693 ) | 0 ( 0 ) | 0.021904 ( 0.00049 ) | 0.022304 ( 4e-06 ) | -46.396168 ( 2.434338 ) | 1.82536 ( 2.278084 ) | 0.017026 ( 0.000178 ) | 1 ( 0 ) |
| -0.2 | True model | 0 ( 0.000489 ) | 0.948 ( 0.007021 ) | 0.015453 ( 0.000346 ) | 0.015739 ( 2e-06 ) | 0 ( 0 ) | 1.85193 ( 2.278641 ) | 0.000239 ( 1e-05 ) | 1 ( 0 ) |
| -0.2 | CCA | -0.083767 ( 0.000693 ) | 0.032 ( 0.005566 ) | 0.021918 ( 0.00049 ) | 0.0219 ( 3e-06 ) | -50.295042 ( 2.210763 ) | -0.080977 ( 2.235424 ) | 0.007497 ( 0.000118 ) | 1 ( 0 ) |
| -0.2 | ipw logadd i | -0.000343 ( 0.000666 ) | 0.963 ( 0.005969 ) | 0.021073 ( 0.000471 ) | 0.021968 ( 3e-06 ) | -46.231297 ( 2.309862 ) | 4.242668 ( 2.332154 ) | 0.000444 ( 2e-05 ) | 1 ( 0 ) |
| -0.2 | ipw logadd noi | -0.088485 ( 0.000695 ) | 0.019 ( 0.004317 ) | 0.021974 ( 0.000492 ) | 0.022027 ( 3e-06 ) | -50.548606 ( 2.214761 ) | 0.242399 ( 2.24266 ) | 0.008312 ( 0.000125 ) | 1 ( 0 ) |
| -0.2 | ipw logit i | 0.000303 ( 0.000666 ) | 0.959 ( 0.00627 ) | 0.021045 ( 0.000471 ) | 0.02197 ( 3e-06 ) | -46.085942 ( 2.316393 ) | 4.395596 ( 2.335576 ) | 0.000443 ( 2e-05 ) | 1 ( 0 ) |
| -0.2 | ipw logit noi | -0.08306 ( 0.000695 ) | 0.037 ( 0.005969 ) | 0.021992 ( 0.000492 ) | 0.022033 ( 3e-06 ) | -50.630107 ( 2.215091 ) | 0.184151 ( 2.241357 ) | 0.007382 ( 0.000118 ) | 1 ( 0 ) |
| -0.1 | True model | 0 ( 0.000497 ) | 0.944 ( 0.007271 ) | 0.01573 ( 0.000352 ) | 0.015743 ( 2e-06 ) | 0 ( 0 ) | 0.083381 ( 2.239077 ) | 0.000247 ( 1.1e-05 ) | 1 ( 0 ) |
| -0.1 | CCA | -0.05252 ( 0.000692 ) | 0.32 ( 0.014751 ) | 0.021883 ( 0.00049 ) | 0.021648 ( 3e-06 ) | -48.330266 ( 2.200819 ) | -1.075417 ( 2.213175 ) | 0.003237 ( 7.4e-05 ) | 1 ( 0 ) |
| -0.1 | ipw logadd i | 0.001513 ( 0.000677 ) | 0.952 ( 0.00676 ) | 0.0214 ( 0.000479 ) | 0.021754 ( 3e-06 ) | -45.971306 ( 2.230395 ) | 1.653869 ( 2.274236 ) | 0.00046 ( 2.1e-05 ) | 1 ( 0 ) |
| -0.1 | ipw logadd noi | -0.057221 ( 0.000696 ) | 0.252 ( 0.013729 ) | 0.022021 ( 0.000493 ) | 0.021805 ( 3e-06 ) | -48.974304 ( 2.200935 ) | -0.982735 ( 2.215249 ) | 0.003759 ( 8.1e-05 ) | 1 ( 0 ) |
| -0.1 | ipw logit i | 0.000366 ( 0.000676 ) | 0.956 ( 0.006486 ) | 0.021381 ( 0.000478 ) | 0.021763 ( 3e-06 ) | -45.871816 ( 2.235804 ) | 1.788798 ( 2.277255 ) | 0.000457 ( 2.1e-05 ) | 1 ( 0 ) |
| -0.1 | ipw logit noi | -0.04005 ( 0.000696 ) | 0.532 ( 0.015779 ) | 0.022 ( 0.000492 ) | 0.021803 ( 3e-06 ) | -48.875806 ( 2.206234 ) | -0.896338 ( 2.217182 ) | 0.002088 ( 5.8e-05 ) | 1 ( 0 ) |
| 0 | True model | 0 ( 0.000493 ) | 0.945 ( 0.007209 ) | 0.015598 ( 0.000349 ) | 0.015743 ( 2e-06 ) | 0 ( 0 ) | 0.927701 ( 2.257967 ) | 0.000243 ( 1.1e-05 ) | 1 ( 0 ) |
| 0 | CCA | -0.026045 ( 0.000679 ) | 0.781 ( 0.013078 ) | 0.021474 ( 0.00048 ) | 0.021428 ( 3e-06 ) | -47.236724 ( 2.276163 ) | -0.213848 ( 2.232448 ) | 0.001139 ( 4.2e-05 ) | 1 ( 0 ) |
| 0 | ipw logadd i | 0.005086 ( 0.000663 ) | 0.947 ( 0.007085 ) | 0.020954 ( 0.000469 ) | 0.021579 ( 3e-06 ) | -44.583213 ( 2.304151 ) | 2.986429 ( 2.304047 ) | 0.000464 ( 2.1e-05 ) | 1 ( 0 ) |
| 0 | ipw logadd noi | -0.030161 ( 0.000688 ) | 0.731 ( 0.014023 ) | 0.02176 ( 0.000487 ) | 0.021613 ( 3e-06 ) | -48.613902 ( 2.229189 ) | -0.673796 ( 2.222159 ) | 0.001383 ( 4.8e-05 ) | 1 ( 0 ) |
| 0 | ipw logit i | 0.000635 ( 0.000661 ) | 0.954 ( 0.006624 ) | 0.020909 ( 0.000468 ) | 0.021604 ( 3e-06 ) | -44.344501 ( 2.314997 ) | 3.328686 ( 2.311705 ) | 0.000437 ( 2e-05 ) | 1 ( 0 ) |
| 0 | ipw logit noi | 0.000804 ( 0.000686 ) | 0.953 ( 0.006693 ) | 0.021692 ( 0.000485 ) | 0.021604 ( 3e-06 ) | -48.293589 ( 2.240751 ) | -0.406576 ( 2.228137 ) | 0.000471 ( 2.1e-05 ) | 1 ( 0 ) |
| 0.1 | True model | 0 ( 0.000504 ) | 0.947 ( 0.007085 ) | 0.015946 ( 0.000357 ) | 0.01574 ( 2e-06 ) | 0 ( 0 ) | -1.292767 ( 2.208289 ) | 0.000254 ( 1.1e-05 ) | 1 ( 0 ) |
| 0.1 | CCA | -0.005322 ( 0.000668 ) | 0.938 ( 0.007626 ) | 0.021129 ( 0.000473 ) | 0.021235 ( 3e-06 ) | -43.042944 ( 2.422224 ) | 0.499917 ( 2.248414 ) | 0.000474 ( 2.1e-05 ) | 1 ( 0 ) |
| 0.1 | ipw logadd i | 0.010005 ( 0.000657 ) | 0.935 ( 0.007796 ) | 0.020785 ( 0.000465 ) | 0.021428 ( 3e-06 ) | -41.14522 ( 2.379284 ) | 3.089186 ( 2.306344 ) | 0.000532 ( 2.4e-05 ) | 1 ( 0 ) |
| 0.1 | ipw logadd noi | -0.008135 ( 0.000682 ) | 0.927 ( 0.008226 ) | 0.021557 ( 0.000482 ) | 0.021446 ( 3e-06 ) | -45.285121 ( 2.351528 ) | -0.518706 ( 2.225626 ) | 0.00053 ( 2.4e-05 ) | 1 ( 0 ) |
| 0.1 | ipw logit i | 0.001054 ( 0.000656 ) | 0.96 ( 0.006197 ) | 0.020754 ( 0.000464 ) | 0.021482 ( 3e-06 ) | -40.964506 ( 2.385329 ) | 3.507311 ( 2.315701 ) | 0.000431 ( 1.9e-05 ) | 1 ( 0 ) |
| 0.1 | ipw logit noi | 0.038145 ( 0.00068 ) | 0.576 ( 0.015628 ) | 0.021501 ( 0.000481 ) | 0.021431 ( 3e-06 ) | -44.994783 ( 2.354105 ) | -0.321516 ( 2.230038 ) | 0.001917 ( 5.6e-05 ) | 1 ( 0 ) |
| 0.2 | True model | 0 ( 0.000485 ) | 0.95 ( 0.006892 ) | 0.015333 ( 0.000343 ) | 0.015743 ( 2e-06 ) | 0 ( 0 ) | 2.671548 ( 2.296979 ) | 0.000235 ( 1e-05 ) | 1 ( 0 ) |
| 0.2 | CCA | 0.009584 ( 0.000655 ) | 0.927 ( 0.008226 ) | 0.020707 ( 0.000463 ) | 0.021069 ( 3e-06 ) | -45.171644 ( 2.295821 ) | 1.746315 ( 2.276295 ) | 0.00052 ( 2.3e-05 ) | 1 ( 0 ) |
| 0.2 | ipw logadd i | 0.014363 ( 0.000642 ) | 0.912 ( 0.008959 ) | 0.020295 ( 0.000454 ) | 0.021296 ( 3e-06 ) | -42.91934 ( 2.30408 ) | 4.931705 ( 2.347562 ) | 0.000618 ( 2.6e-05 ) | 1 ( 0 ) |
| 0.2 | ipw logadd noi | 0.008378 ( 0.000661 ) | 0.936 ( 0.00774 ) | 0.02089 ( 0.000467 ) | 0.021302 ( 3e-06 ) | -46.12502 ( 2.292229 ) | 1.970497 ( 2.281312 ) | 0.000506 ( 2.2e-05 ) | 1 ( 0 ) |
| 0.2 | ipw logit i | -3.3e-05 ( 0.000642 ) | 0.964 ( 0.005891 ) | 0.020294 ( 0.000454 ) | 0.021392 ( 3e-06 ) | -42.916611 ( 2.312825 ) | 5.4062 ( 2.358181 ) | 0.000411 ( 1.8e-05 ) | 1 ( 0 ) |
| 0.2 | ipw logit noi | 0.070836 ( 0.000657 ) | 0.08 ( 0.008579 ) | 0.020787 ( 0.000465 ) | 0.021285 ( 3e-06 ) | -45.590341 ( 2.305831 ) | 2.397662 ( 2.290868 ) | 0.005449 ( 9.6e-05 ) | 1 ( 0 ) |
| 0.3 | True model | 0 ( 0.000501 ) | 0.952 ( 0.00676 ) | 0.015855 ( 0.000355 ) | 0.015741 ( 2e-06 ) | 0 ( 0 ) | -0.72214 ( 2.221055 ) | 0.000251 ( 1.1e-05 ) | 1 ( 0 ) |
| 0.3 | CCA | 0.021121 ( 0.000665 ) | 0.823 ( 0.012069 ) | 0.02103 ( 0.00047 ) | 0.020927 ( 3e-06 ) | -43.155833 ( 2.326344 ) | -0.488084 ( 2.226304 ) | 0.000888 ( 3.3e-05 ) | 1 ( 0 ) |
| 0.3 | ipw logadd i | 0.020005 ( 0.000647 ) | 0.842 ( 0.011534 ) | 0.020469 ( 0.000458 ) | 0.021179 ( 3e-06 ) | -40.000179 ( 2.340001 ) | 3.465823 ( 2.314765 ) | 0.000819 ( 3.1e-05 ) | 1 ( 0 ) |
| 0.3 | ipw logadd noi | 0.022103 ( 0.000672 ) | 0.821 ( 0.012123 ) | 0.02124 ( 0.000475 ) | 0.021177 ( 3e-06 ) | -44.273138 ( 2.324051 ) | -0.296078 ( 2.230602 ) | 0.000939 ( 3.5e-05 ) | 1 ( 0 ) |
| 0.3 | ipw logit i | -0.000344 ( 0.000646 ) | 0.957 ( 0.006415 ) | 0.020421 ( 0.000457 ) | 0.021329 ( 3e-06 ) | -39.715544 ( 2.359567 ) | 4.44683 ( 2.336718 ) | 0.000417 ( 1.8e-05 ) | 1 ( 0 ) |
| 0.3 | ipw logit noi | 0.101 ( 0.000668 ) | 0.005 ( 0.00223 ) | 0.021124 ( 0.000473 ) | 0.021163 ( 3e-06 ) | -43.66204 ( 2.327747 ) | 0.184163 ( 2.241345 ) | 0.010647 ( 0.000135 ) | 1 ( 0 ) |

#### 2.2 Strue=log-additive

##### 2.2.1 Y is continuous

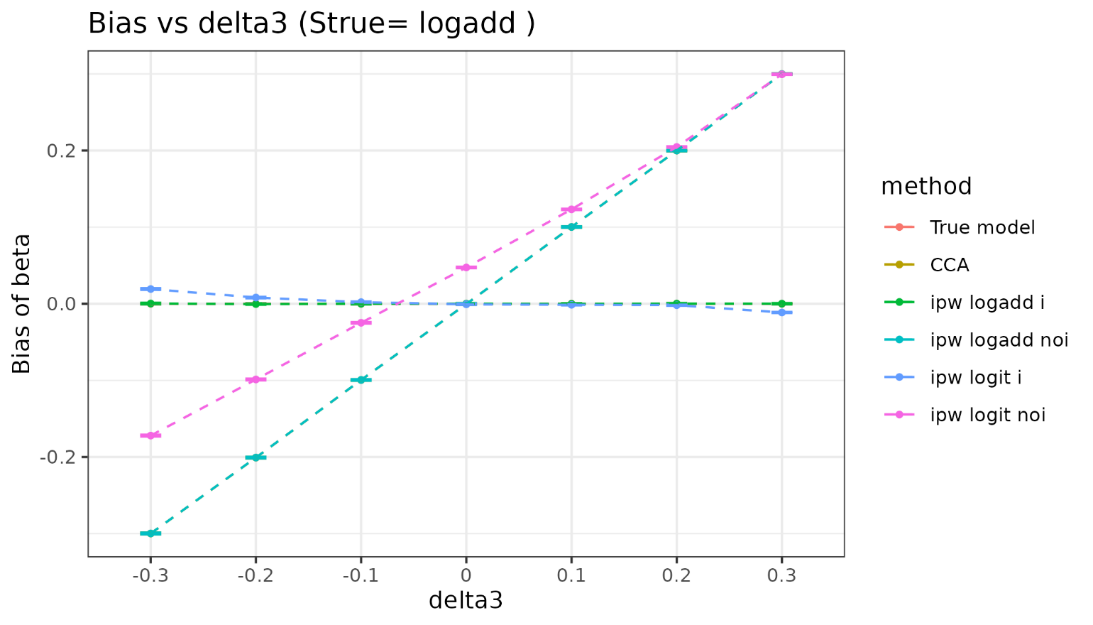

| delta3 | method | bias | coverage | EmpSE | ModSE | relative_precision | relative_error_ModSE | MSE | power |
| --- | --- | --- | --- | --- | --- | --- | --- | --- | --- |
| -0.3 | True model | 0 ( 0.000307 ) | 0.952 ( 0.00676 ) | 0.009715 ( 0.000217 ) | 0.00976 ( 1e-06 ) | 0 ( 0 ) | 0.462243 ( 2.247543 ) | 9.4e-05 ( 4e-06 ) | 1 ( 0 ) |
| -0.3 | CCA | -0.299817 ( 0.000586 ) | 0 ( 0 ) | 0.018523 ( 0.000414 ) | 0.018612 ( 4e-06 ) | -72.49465 ( 1.489045 ) | 0.475364 ( 2.247907 ) | 0.090233 ( 0.000351 ) | 1 ( 0 ) |
| -0.3 | ipw logadd i | 0.000306 ( 0.000526 ) | 0.989 ( 0.003298 ) | 0.016646 ( 0.000372 ) | 0.020682 ( 1.2e-05 ) | -65.941242 ( 1.753934 ) | 24.244638 ( 2.78054 ) | 0.000277 ( 1.2e-05 ) | 1 ( 0 ) |
| -0.3 | ipw logadd noi | -0.299765 ( 0.000594 ) | 0 ( 0 ) | 0.018787 ( 0.00042 ) | 0.018892 ( 6e-06 ) | -73.261918 ( 1.452141 ) | 0.55867 ( 2.249914 ) | 0.090212 ( 0.000356 ) | 1 ( 0 ) |
| -0.3 | ipw logit i | 0.01924 ( 0.000545 ) | 0.919 ( 0.008628 ) | 0.017244 ( 0.000386 ) | 0.021568 ( 1.7e-05 ) | -68.260973 ( 1.66633 ) | 25.077471 ( 2.7999 ) | 0.000667 ( 2.5e-05 ) | 1 ( 0 ) |
| -0.3 | ipw logit noi | -0.172084 ( 0.000612 ) | 0 ( 0 ) | 0.01936 ( 0.000433 ) | 0.019667 ( 8e-06 ) | -74.819426 ( 1.375241 ) | 1.586086 ( 2.273038 ) | 0.029987 ( 0.000211 ) | 1 ( 0 ) |
| -0.2 | True model | 0 ( 0.000308 ) | 0.951 ( 0.006826 ) | 0.009733 ( 0.000218 ) | 0.00976 ( 1e-06 ) | 0 ( 0 ) | 0.280413 ( 2.243476 ) | 9.5e-05 ( 4e-06 ) | 1 ( 0 ) |
| -0.2 | CCA | -0.200812 ( 0.000553 ) | 0 ( 0 ) | 0.017479 ( 0.000391 ) | 0.017546 ( 3e-06 ) | -68.995641 ( 1.637854 ) | 0.380646 ( 2.245772 ) | 0.040631 ( 0.000222 ) | 1 ( 0 ) |
| -0.2 | ipw logadd i | -0.000372 ( 0.000491 ) | 0.982 ( 0.004204 ) | 0.015532 ( 0.000347 ) | 0.018606 ( 7e-06 ) | -60.734551 ( 1.894823 ) | 19.788916 ( 2.680327 ) | 0.000241 ( 1.1e-05 ) | 1 ( 0 ) |
| -0.2 | ipw logadd noi | -0.200723 ( 0.000562 ) | 0 ( 0 ) | 0.017767 ( 0.000397 ) | 0.017773 ( 5e-06 ) | -69.990624 ( 1.588735 ) | 0.034819 ( 2.238143 ) | 0.040605 ( 0.000225 ) | 1 ( 0 ) |
| -0.2 | ipw logit i | 0.008122 ( 0.000497 ) | 0.965 ( 0.005812 ) | 0.015731 ( 0.000352 ) | 0.019027 ( 9e-06 ) | -61.723863 ( 1.860259 ) | 20.946821 ( 2.706393 ) | 0.000313 ( 1.4e-05 ) | 1 ( 0 ) |
| -0.2 | ipw logit noi | -0.098765 ( 0.000575 ) | 0.001 ( 0.000999 ) | 0.018194 ( 0.000407 ) | 0.018298 ( 6e-06 ) | -71.384271 ( 1.50796 ) | 0.573463 ( 2.25027 ) | 0.010085 ( 0.000114 ) | 1 ( 0 ) |
| -0.1 | True model | 0 ( 0.000311 ) | 0.942 ( 0.007392 ) | 0.00983 ( 0.00022 ) | 0.00976 ( 1e-06 ) | 0 ( 0 ) | -0.710937 ( 2.221297 ) | 9.7e-05 ( 4e-06 ) | 1 ( 0 ) |
| -0.1 | CCA | -0.0995 ( 0.000496 ) | 0 ( 0 ) | 0.015678 ( 0.000351 ) | 0.016548 ( 3e-06 ) | -60.684557 ( 2.042314 ) | 5.553677 ( 2.361496 ) | 0.010146 ( 9.9e-05 ) | 1 ( 0 ) |
| -0.1 | ipw logadd i | 5.8e-05 ( 0.000426 ) | 0.99 ( 0.003146 ) | 0.013469 ( 0.000301 ) | 0.016985 ( 5e-06 ) | -46.733049 ( 2.433238 ) | 26.102669 ( 2.821377 ) | 0.000181 ( 8e-06 ) | 1 ( 0 ) |
| -0.1 | ipw logadd noi | -0.099459 ( 0.000501 ) | 0 ( 0 ) | 0.015838 ( 0.000354 ) | 0.016722 ( 4e-06 ) | -61.4779 ( 2.016517 ) | 5.582747 ( 2.362225 ) | 0.010143 ( 1e-04 ) | 1 ( 0 ) |
| -0.1 | ipw logit i | 0.002067 ( 0.000428 ) | 0.988 ( 0.003443 ) | 0.013524 ( 0.000303 ) | 0.017157 ( 5e-06 ) | -47.164848 ( 2.417582 ) | 26.866664 ( 2.838498 ) | 0.000187 ( 8e-06 ) | 1 ( 0 ) |
| -0.1 | ipw logit noi | -0.024802 ( 0.000507 ) | 0.706 ( 0.014407 ) | 0.016037 ( 0.000359 ) | 0.017042 ( 5e-06 ) | -62.425731 ( 1.963573 ) | 6.26929 ( 2.377623 ) | 0.000872 ( 2.8e-05 ) | 1 ( 0 ) |
| 0 | True model | 0 ( 0.000319 ) | 0.944 ( 0.007271 ) | 0.010073 ( 0.000225 ) | 0.009759 ( 1e-06 ) | 0 ( 0 ) | -3.115756 ( 2.167496 ) | 0.000101 ( 5e-06 ) | 1 ( 0 ) |
| 0 | CCA | -0.000358 ( 0.000486 ) | 0.957 ( 0.006415 ) | 0.015375 ( 0.000344 ) | 0.01563 ( 2e-06 ) | -57.072725 ( 2.202252 ) | 1.658626 ( 2.274343 ) | 0.000236 ( 1.1e-05 ) | 1 ( 0 ) |
| 0 | ipw logadd i | 6.8e-05 ( 0.000411 ) | 0.984 ( 0.003968 ) | 0.012992 ( 0.000291 ) | 0.015751 ( 3e-06 ) | -39.88198 ( 2.574273 ) | 21.234986 ( 2.712384 ) | 0.000169 ( 8e-06 ) | 1 ( 0 ) |
| 0 | ipw logadd noi | -0.000316 ( 0.000492 ) | 0.955 ( 0.006556 ) | 0.015557 ( 0.000348 ) | 0.01575 ( 3e-06 ) | -58.073214 ( 2.160321 ) | 1.237886 ( 2.264989 ) | 0.000242 ( 1.1e-05 ) | 1 ( 0 ) |
| 0 | ipw logit i | -0.000733 ( 0.000411 ) | 0.981 ( 0.004317 ) | 0.012999 ( 0.000291 ) | 0.01581 ( 4e-06 ) | -39.950808 ( 2.571073 ) | 21.622577 ( 2.721058 ) | 0.000169 ( 8e-06 ) | 1 ( 0 ) |
| 0 | ipw logit noi | 0.047354 ( 0.000499 ) | 0.143 ( 0.01107 ) | 0.015774 ( 0.000353 ) | 0.015919 ( 4e-06 ) | -59.218994 ( 2.097078 ) | 0.91957 ( 2.257881 ) | 0.002491 ( 4.9e-05 ) | 1 ( 0 ) |
| 0.1 | True model | 0 ( 0.000325 ) | 0.946 ( 0.007147 ) | 0.010266 ( 0.00023 ) | 0.009759 ( 1e-06 ) | 0 ( 0 ) | -4.936156 ( 2.126772 ) | 0.000105 ( 4e-06 ) | 1 ( 0 ) |
| 0.1 | CCA | 0.100259 ( 0.000454 ) | 0 ( 0 ) | 0.014354 ( 0.000321 ) | 0.014808 ( 2e-06 ) | -48.845617 ( 2.40615 ) | 3.165951 ( 2.308061 ) | 0.010258 ( 9.2e-05 ) | 1 ( 0 ) |
| 0.1 | ipw logadd i | -0.000113 ( 0.000395 ) | 0.979 ( 0.004534 ) | 0.012486 ( 0.000279 ) | 0.014837 ( 3e-06 ) | -32.393078 ( 2.633761 ) | 18.830994 ( 2.658569 ) | 0.000156 ( 7e-06 ) | 1 ( 0 ) |
| 0.1 | ipw logadd noi | 0.100273 ( 0.000458 ) | 0 ( 0 ) | 0.014475 ( 0.000324 ) | 0.014874 ( 3e-06 ) | -49.699762 ( 2.376029 ) | 2.756597 ( 2.298946 ) | 0.010264 ( 9.3e-05 ) | 1 ( 0 ) |
| 0.1 | ipw logit i | -0.001341 ( 0.000395 ) | 0.98 ( 0.004427 ) | 0.012492 ( 0.000279 ) | 0.014861 ( 3e-06 ) | -32.46185 ( 2.633204 ) | 18.961556 ( 2.661489 ) | 0.000158 ( 7e-06 ) | 1 ( 0 ) |
| 0.1 | ipw logit noi | 0.123223 ( 0.00046 ) | 0 ( 0 ) | 0.014535 ( 0.000325 ) | 0.014943 ( 3e-06 ) | -50.111903 ( 2.353677 ) | 2.807227 ( 2.300083 ) | 0.015395 ( 0.000114 ) | 1 ( 0 ) |
| 0.2 | True model | 0 ( 0.000318 ) | 0.945 ( 0.007209 ) | 0.010041 ( 0.000225 ) | 0.00976 ( 1e-06 ) | 0 ( 0 ) | -2.79585 ( 2.174653 ) | 0.000101 ( 5e-06 ) | 1 ( 0 ) |
| 0.2 | CCA | 0.19995 ( 0.000441 ) | 0 ( 0 ) | 0.013957 ( 0.000312 ) | 0.014095 ( 2e-06 ) | -48.241626 ( 2.33297 ) | 0.991289 ( 2.259401 ) | 0.040175 ( 0.000177 ) | 1 ( 0 ) |
| 0.2 | ipw logadd i | 0.000165 ( 0.000386 ) | 0.981 ( 0.004317 ) | 0.012211 ( 0.000273 ) | 0.014198 ( 2e-06 ) | -32.381183 ( 2.53133 ) | 16.276616 ( 2.601398 ) | 0.000149 ( 7e-06 ) | 1 ( 0 ) |
| 0.2 | ipw logadd noi | 0.199921 ( 0.000444 ) | 0 ( 0 ) | 0.01403 ( 0.000314 ) | 0.014113 ( 2e-06 ) | -48.780286 ( 2.313386 ) | 0.593035 ( 2.250512 ) | 0.040165 ( 0.000178 ) | 1 ( 0 ) |
| 0.2 | ipw logit i | -0.001811 ( 0.000387 ) | 0.976 ( 0.00484 ) | 0.012223 ( 0.000273 ) | 0.01427 ( 2e-06 ) | -32.518307 ( 2.527732 ) | 16.742787 ( 2.611831 ) | 0.000153 ( 7e-06 ) | 1 ( 0 ) |
| 0.2 | ipw logit noi | 0.204678 ( 0.000444 ) | 0 ( 0 ) | 0.014035 ( 0.000314 ) | 0.014129 ( 2e-06 ) | -48.818625 ( 2.31835 ) | 0.670696 ( 2.25225 ) | 0.04209 ( 0.000182 ) | 1 ( 0 ) |
| 0.3 | True model | 0 ( 0.00032 ) | 0.944 ( 0.007271 ) | 0.010127 ( 0.000227 ) | 0.009758 ( 1e-06 ) | 0 ( 0 ) | -3.645512 ( 2.155644 ) | 0.000102 ( 5e-06 ) | 1 ( 0 ) |
| 0.3 | CCA | 0.299824 ( 0.000437 ) | 0 ( 0 ) | 0.013825 ( 0.000309 ) | 0.013506 ( 2e-06 ) | -46.335957 ( 2.34355 ) | -2.302867 ( 2.185701 ) | 0.090085 ( 0.000263 ) | 1 ( 0 ) |
| 0.3 | ipw logadd i | 9.6e-05 ( 0.000391 ) | 0.975 ( 0.004937 ) | 0.012374 ( 0.000277 ) | 0.013816 ( 2e-06 ) | -33.016724 ( 2.434349 ) | 11.652639 ( 2.497952 ) | 0.000153 ( 7e-06 ) | 1 ( 0 ) |
| 0.3 | ipw logadd noi | 0.29982 ( 0.000437 ) | 0 ( 0 ) | 0.013828 ( 0.000309 ) | 0.013504 ( 2e-06 ) | -46.359757 ( 2.342032 ) | -2.340342 ( 2.184874 ) | 0.090083 ( 0.000263 ) | 1 ( 0 ) |
| 0.3 | ipw logit i | -0.011373 ( 0.000393 ) | 0.907 ( 0.009184 ) | 0.012432 ( 0.000278 ) | 0.014163 ( 3e-06 ) | -33.639486 ( 2.452102 ) | 13.922968 ( 2.548782 ) | 0.000284 ( 1.1e-05 ) | 1 ( 0 ) |
| 0.3 | ipw logit noi | 0.299517 ( 0.000438 ) | 0 ( 0 ) | 0.013848 ( 0.00031 ) | 0.013504 ( 2e-06 ) | -46.518796 ( 2.333195 ) | -2.484461 ( 2.18165 ) | 0.089902 ( 0.000263 ) | 1 ( 0 ) |

##### 2.2.2 Y is binary

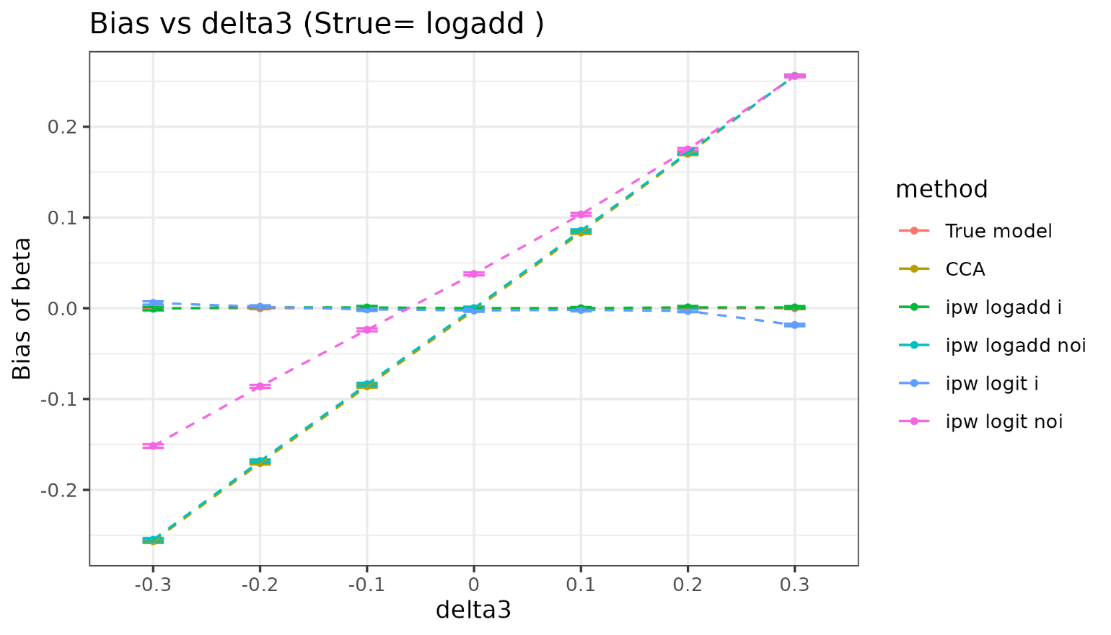

| delta3 | method | bias | coverage | EmpSE | ModSE | relative_precision | relative_error_ModSE | MSE | power |
| --- | --- | --- | --- | --- | --- | --- | --- | --- | --- |
| -0.3 | True model | 0 ( 0.000507 ) | 0.95 ( 0.006892 ) | 0.016037 ( 0.000359 ) | 0.015739 ( 2e-06 ) | 0 ( 0 ) | -1.858385 ( 2.195635 ) | 0.000257 ( 1.1e-05 ) | 1 ( 0 ) |
| -0.3 | CCA | -0.256635 ( 0.000934 ) | 0 ( 0 ) | 0.029548 ( 0.000661 ) | 0.028594 ( 7e-06 ) | -70.541106 ( 1.520769 ) | -3.228044 ( 2.165082 ) | 0.066734 ( 0.000483 ) | 1 ( 0 ) |
| -0.3 | ipw logadd i | -0.000479 ( 0.000949 ) | 0.948 ( 0.007021 ) | 0.030021 ( 0.000672 ) | 0.030384 ( 7e-06 ) | -71.46327 ( 1.456919 ) | 1.208002 ( 2.264347 ) | 0.000901 ( 3.8e-05 ) | 1 ( 0 ) |
| -0.3 | ipw logadd noi | -0.254977 ( 0.000949 ) | 0 ( 0 ) | 0.030015 ( 0.000671 ) | 0.02883 ( 7e-06 ) | -71.451872 ( 1.475228 ) | -3.948141 ( 2.14897 ) | 0.065913 ( 0.000487 ) | 1 ( 0 ) |
| -0.3 | ipw logit i | 0.005864 ( 0.000946 ) | 0.952 ( 0.00676 ) | 0.02991 ( 0.000669 ) | 0.030503 ( 8e-06 ) | -71.250907 ( 1.467214 ) | 1.98383 ( 2.281712 ) | 0.000928 ( 3.9e-05 ) | 1 ( 0 ) |
| -0.3 | ipw logit noi | -0.151697 ( 0.000967 ) | 0 ( 0 ) | 0.03058 ( 0.000684 ) | 0.029291 ( 7e-06 ) | -72.496404 ( 1.430223 ) | -4.214879 ( 2.143005 ) | 0.023946 ( 0.000298 ) | 1 ( 0 ) |
| -0.2 | True model | 0 ( 0.000489 ) | 0.948 ( 0.007021 ) | 0.015453 ( 0.000346 ) | 0.015739 ( 2e-06 ) | 0 ( 0 ) | 1.85193 ( 2.278641 ) | 0.000239 ( 1e-05 ) | 1 ( 0 ) |
| -0.2 | CCA | -0.170494 ( 0.000842 ) | 0 ( 0 ) | 0.02663 ( 0.000596 ) | 0.02736 ( 6e-06 ) | -66.329736 ( 1.783128 ) | 2.740538 ( 2.298599 ) | 0.029777 ( 0.000288 ) | 1 ( 0 ) |
| -0.2 | ipw logadd i | 0.001289 ( 0.000842 ) | 0.963 ( 0.005969 ) | 0.026625 ( 0.000596 ) | 0.028297 ( 6e-06 ) | -66.316646 ( 1.772783 ) | 6.278567 ( 2.377759 ) | 0.00071 ( 3.3e-05 ) | 1 ( 0 ) |
| -0.2 | ipw logadd noi | -0.168335 ( 0.000855 ) | 0 ( 0 ) | 0.027033 ( 0.000605 ) | 0.027558 ( 6e-06 ) | -67.324679 ( 1.738729 ) | 1.942311 ( 2.280739 ) | 0.029067 ( 0.000288 ) | 1 ( 0 ) |
| -0.2 | ipw logit i | 0.001675 ( 0.00084 ) | 0.965 ( 0.005812 ) | 0.026547 ( 0.000594 ) | 0.028314 ( 6e-06 ) | -66.11871 ( 1.783436 ) | 6.65306 ( 2.386137 ) | 0.000707 ( 3.3e-05 ) | 1 ( 0 ) |
| -0.2 | ipw logit noi | -0.086111 ( 0.000862 ) | 0.126 ( 0.010494 ) | 0.027266 ( 0.00061 ) | 0.027851 ( 6e-06 ) | -67.881944 ( 1.711341 ) | 2.145888 ( 2.285295 ) | 0.008158 ( 0.000151 ) | 1 ( 0 ) |
| -0.1 | True model | 0 ( 0.000497 ) | 0.944 ( 0.007271 ) | 0.01573 ( 0.000352 ) | 0.015743 ( 2e-06 ) | 0 ( 0 ) | 0.083381 ( 2.239077 ) | 0.000247 ( 1.1e-05 ) | 1 ( 0 ) |
| -0.1 | CCA | -0.085908 ( 0.000823 ) | 0.091 ( 0.009095 ) | 0.026035 ( 0.000582 ) | 0.026197 ( 5e-06 ) | -63.496796 ( 1.878804 ) | 0.62042 ( 2.251163 ) | 0.008057 ( 0.000144 ) | 1 ( 0 ) |
| -0.1 | ipw logadd i | 0.000959 ( 0.000783 ) | 0.965 ( 0.005812 ) | 0.024775 ( 0.000554 ) | 0.026593 ( 5e-06 ) | -59.68769 ( 2.028994 ) | 7.339274 ( 2.401477 ) | 0.000614 ( 2.8e-05 ) | 1 ( 0 ) |
| -0.1 | ipw logadd noi | -0.08368 ( 0.00083 ) | 0.11 ( 0.009894 ) | 0.026248 ( 0.000587 ) | 0.026351 ( 5e-06 ) | -64.084786 ( 1.855522 ) | 0.39271 ( 2.246068 ) | 0.007691 ( 0.000141 ) | 1 ( 0 ) |
| -0.1 | ipw logit i | -0.001353 ( 0.000781 ) | 0.966 ( 0.005731 ) | 0.024708 ( 0.000553 ) | 0.026589 ( 5e-06 ) | -59.468025 ( 2.039165 ) | 7.612804 ( 2.407596 ) | 0.000612 ( 2.8e-05 ) | 1 ( 0 ) |
| -0.1 | ipw logit noi | -0.023751 ( 0.000834 ) | 0.853 ( 0.011198 ) | 0.026367 ( 0.00059 ) | 0.026513 ( 5e-06 ) | -64.408671 ( 1.845738 ) | 0.555982 ( 2.249721 ) | 0.001259 ( 5e-05 ) | 1 ( 0 ) |
| 0 | True model | 0 ( 0.000493 ) | 0.945 ( 0.007209 ) | 0.015598 ( 0.000349 ) | 0.015743 ( 2e-06 ) | 0 ( 0 ) | 0.927701 ( 2.257967 ) | 0.000243 ( 1.1e-05 ) | 1 ( 0 ) |
| 0 | CCA | -0.002227 ( 0.000776 ) | 0.961 ( 0.006122 ) | 0.024551 ( 0.000549 ) | 0.025108 ( 5e-06 ) | -59.632529 ( 1.987904 ) | 2.269497 ( 2.28805 ) | 0.000607 ( 2.7e-05 ) | 1 ( 0 ) |
| 0 | ipw logadd i | 9.2e-05 ( 0.000754 ) | 0.971 ( 0.005307 ) | 0.02383 ( 0.000533 ) | 0.025215 ( 5e-06 ) | -57.155111 ( 2.04122 ) | 5.810332 ( 2.367266 ) | 0.000567 ( 2.4e-05 ) | 1 ( 0 ) |
| 0 | ipw logadd noi | 2.4e-05 ( 0.000782 ) | 0.953 ( 0.006693 ) | 0.02472 ( 0.000553 ) | 0.025214 ( 5e-06 ) | -60.183268 ( 1.959796 ) | 2.000098 ( 2.282022 ) | 0.00061 ( 2.7e-05 ) | 1 ( 0 ) |
| 0 | ipw logit i | -0.002427 ( 0.000753 ) | 0.971 ( 0.005307 ) | 0.023809 ( 0.000533 ) | 0.025214 ( 5e-06 ) | -57.077016 ( 2.044339 ) | 5.904191 ( 2.369366 ) | 0.000572 ( 2.4e-05 ) | 1 ( 0 ) |
| 0 | ipw logit noi | 0.0379 ( 0.000784 ) | 0.696 ( 0.014546 ) | 0.024793 ( 0.000555 ) | 0.025286 ( 5e-06 ) | -60.417227 ( 1.948927 ) | 1.989569 ( 2.281787 ) | 0.00205 ( 6.7e-05 ) | 1 ( 0 ) |
| 0.1 | True model | 0 ( 0.000504 ) | 0.947 ( 0.007085 ) | 0.015946 ( 0.000357 ) | 0.01574 ( 2e-06 ) | 0 ( 0 ) | -1.292767 ( 2.208289 ) | 0.000254 ( 1.1e-05 ) | 1 ( 0 ) |
| 0.1 | CCA | 0.083387 ( 0.00078 ) | 0.064 ( 0.00774 ) | 0.02468 ( 0.000552 ) | 0.024123 ( 5e-06 ) | -58.252933 ( 1.958776 ) | -2.257161 ( 2.186766 ) | 0.007562 ( 0.000135 ) | 1 ( 0 ) |
| 0.1 | ipw logadd i | 1.8e-05 ( 0.000745 ) | 0.961 ( 0.006122 ) | 0.023558 ( 0.000527 ) | 0.024143 ( 5e-06 ) | -54.183933 ( 2.09583 ) | 2.481473 ( 2.292782 ) | 0.000554 ( 2.5e-05 ) | 1 ( 0 ) |
| 0.1 | ipw logadd noi | 0.085355 ( 0.000783 ) | 0.055 ( 0.007209 ) | 0.024775 ( 0.000554 ) | 0.024182 ( 5e-06 ) | -58.57307 ( 1.952069 ) | -2.393042 ( 2.183726 ) | 0.007899 ( 0.000139 ) | 1 ( 0 ) |
| 0.1 | ipw logit i | -0.001859 ( 0.000745 ) | 0.958 ( 0.006343 ) | 0.02356 ( 0.000527 ) | 0.024145 ( 5e-06 ) | -54.190418 ( 2.095784 ) | 2.482113 ( 2.292796 ) | 0.000558 ( 2.5e-05 ) | 1 ( 0 ) |
| 0.1 | ipw logit noi | 0.103391 ( 0.000783 ) | 0.009 ( 0.002986 ) | 0.024774 ( 0.000554 ) | 0.024203 ( 5e-06 ) | -58.569467 ( 1.952973 ) | -2.305235 ( 2.185691 ) | 0.011303 ( 0.000167 ) | 1 ( 0 ) |
| 0.2 | True model | 0 ( 0.000485 ) | 0.95 ( 0.006892 ) | 0.015333 ( 0.000343 ) | 0.015743 ( 2e-06 ) | 0 ( 0 ) | 2.671548 ( 2.296979 ) | 0.000235 ( 1e-05 ) | 1 ( 0 ) |
| 0.2 | CCA | 0.170163 ( 0.000745 ) | 0 ( 0 ) | 0.023567 ( 0.000527 ) | 0.023286 ( 4e-06 ) | -57.669865 ( 1.994832 ) | -1.194036 ( 2.210543 ) | 0.02951 ( 0.000256 ) | 1 ( 0 ) |
| 0.2 | ipw logadd i | 0.00106 ( 0.000719 ) | 0.963 ( 0.005969 ) | 0.022725 ( 0.000508 ) | 0.023385 ( 4e-06 ) | -54.472921 ( 2.08272 ) | 2.904918 ( 2.30225 ) | 0.000517 ( 2.3e-05 ) | 1 ( 0 ) |
| 0.2 | ipw logadd noi | 0.171494 ( 0.000747 ) | 0 ( 0 ) | 0.023633 ( 0.000529 ) | 0.023305 ( 4e-06 ) | -57.905244 ( 1.982639 ) | -1.387079 ( 2.206224 ) | 0.029968 ( 0.000259 ) | 1 ( 0 ) |
| 0.2 | ipw logit i | -0.002958 ( 0.000718 ) | 0.956 ( 0.006486 ) | 0.022705 ( 0.000508 ) | 0.023395 ( 4e-06 ) | -54.396199 ( 2.087207 ) | 3.038031 ( 2.305229 ) | 0.000524 ( 2.3e-05 ) | 1 ( 0 ) |
| 0.2 | ipw logit noi | 0.17502 ( 0.000748 ) | 0 ( 0 ) | 0.02365 ( 0.000529 ) | 0.023307 ( 4e-06 ) | -57.967814 ( 1.980006 ) | -1.453071 ( 2.204747 ) | 0.031191 ( 0.000264 ) | 1 ( 0 ) |
| 0.3 | True model | 0 ( 0.000501 ) | 0.952 ( 0.00676 ) | 0.015855 ( 0.000355 ) | 0.015741 ( 2e-06 ) | 0 ( 0 ) | -0.72214 ( 2.221055 ) | 0.000251 ( 1.1e-05 ) | 1 ( 0 ) |
| 0.3 | CCA | 0.25598 ( 0.000687 ) | 0 ( 0 ) | 0.021734 ( 0.000486 ) | 0.022579 ( 4e-06 ) | -46.780521 ( 2.354971 ) | 3.888067 ( 2.324226 ) | 0.065998 ( 0.000354 ) | 1 ( 0 ) |
| 0.3 | ipw logadd i | 0.001002 ( 0.000677 ) | 0.96 ( 0.006197 ) | 0.021405 ( 0.000479 ) | 0.022892 ( 4e-06 ) | -45.131291 ( 2.381595 ) | 6.945039 ( 2.392624 ) | 0.000459 ( 2.1e-05 ) | 1 ( 0 ) |
| 0.3 | ipw logadd noi | 0.256061 ( 0.000687 ) | 0 ( 0 ) | 0.021734 ( 0.000486 ) | 0.02258 ( 4e-06 ) | -46.780323 ( 2.35437 ) | 3.889576 ( 2.32426 ) | 0.066039 ( 0.000354 ) | 1 ( 0 ) |
| 0.3 | ipw logit i | -0.018436 ( 0.000679 ) | 0.894 ( 0.009735 ) | 0.021474 ( 0.00048 ) | 0.023032 ( 4e-06 ) | -45.484398 ( 2.387137 ) | 7.251949 ( 2.399498 ) | 0.000801 ( 3.2e-05 ) | 1 ( 0 ) |
| 0.3 | ipw logit noi | 0.255785 ( 0.000687 ) | 0 ( 0 ) | 0.021727 ( 0.000486 ) | 0.02258 ( 4e-06 ) | -46.746851 ( 2.358056 ) | 3.922165 ( 2.324989 ) | 0.065898 ( 0.000354 ) | 1 ( 0 ) |

### 3 Reasons for ipw logadd i is biased for some values of $\boldsymbol{\delta}_{\boldsymbol{3}}$ and sel.prob=0.9 in DAG2

When varying $\delta_{3}$ and Strue=log-additive, P(S=1) could be over 1. We tried to find values of $\delta_{1}$ and $\delta_{2}$ to decrease the maximum % of P(S=1)>1 for each DAG. Figures 1, 2 and 3 below showing the maximum % of P(S=1)>1 for each DAG by each varied $\delta_{3}$. we can find sensible values of $\delta_{1}$ and $\delta_{2}$that make this % equal or close for DAGs 1 and 3, but not for DAG 2. Therefore ipw logadd i is unbiased for DAGs 1 and 3 when varying $\delta_{3}$ and Strue=log-additive. For DAG 2, this % is higher for$\delta_{3}=\pm0.3$, and this explains the reason that ipw logadd I is more biased for $\delta_{3}=\pm0.3$. This reason is similar when sel.prob=0.9, i.e., this % is not close to zero when sel.prob=0.9.

Figure 1. DAG 1.

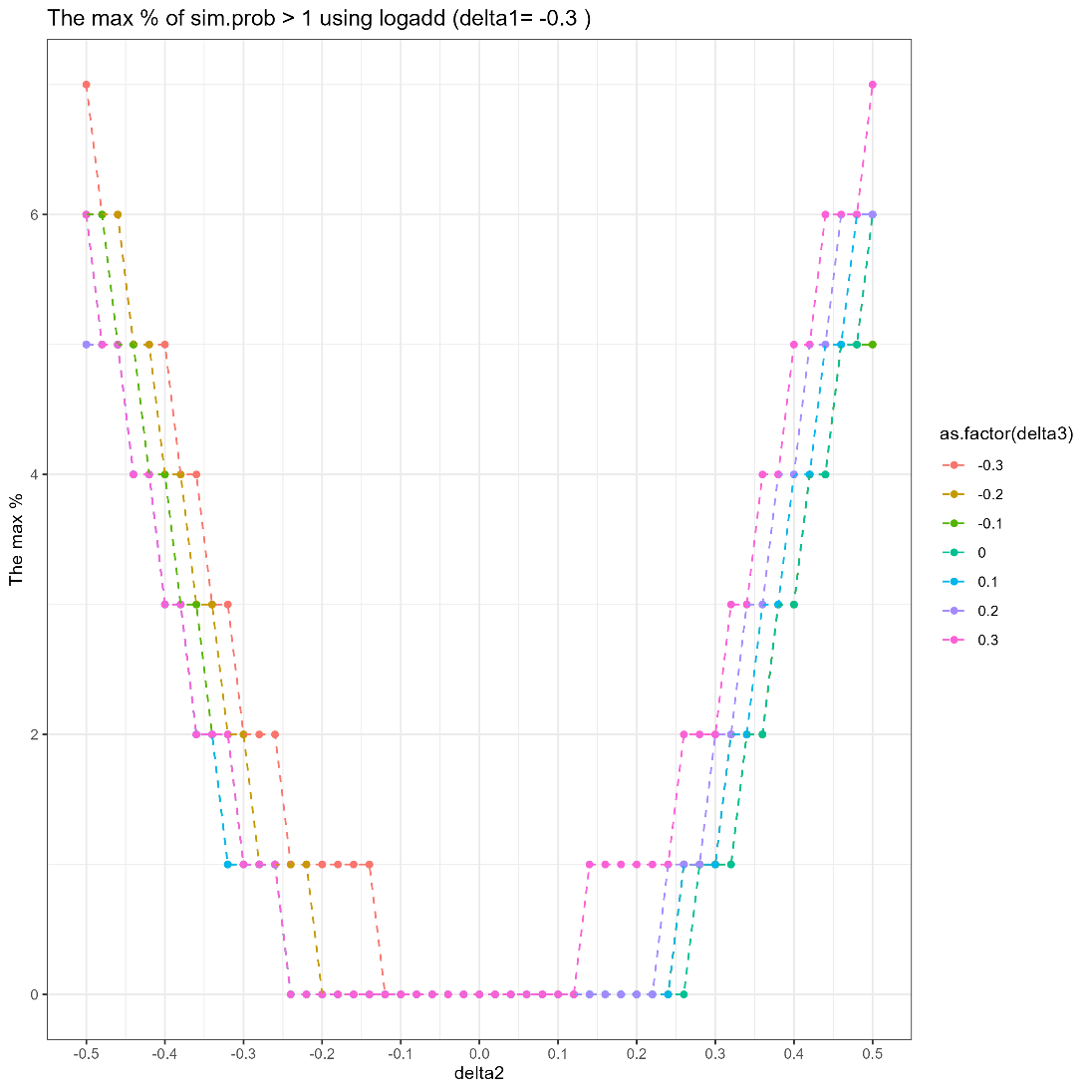

Figure 2. DAG 2.

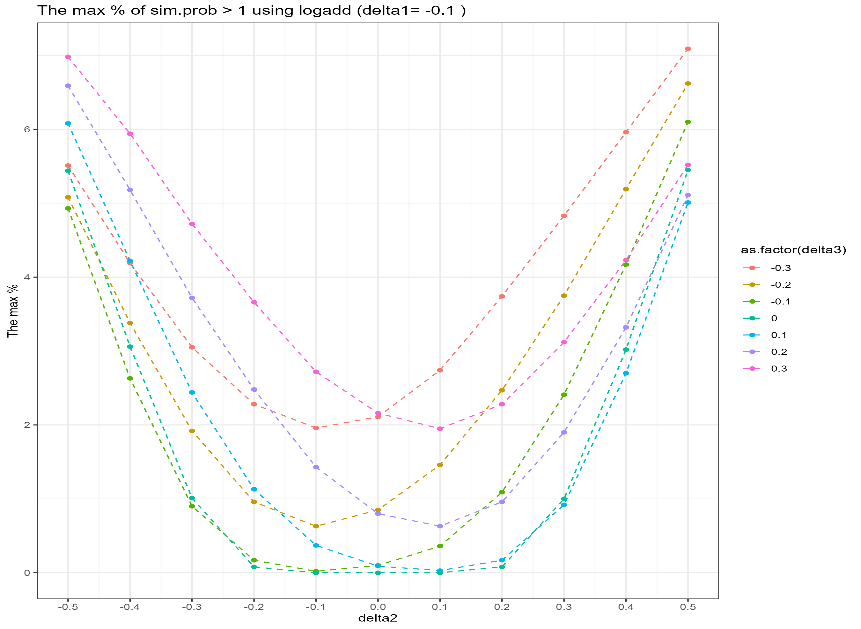

Figure 3. DAG 3.

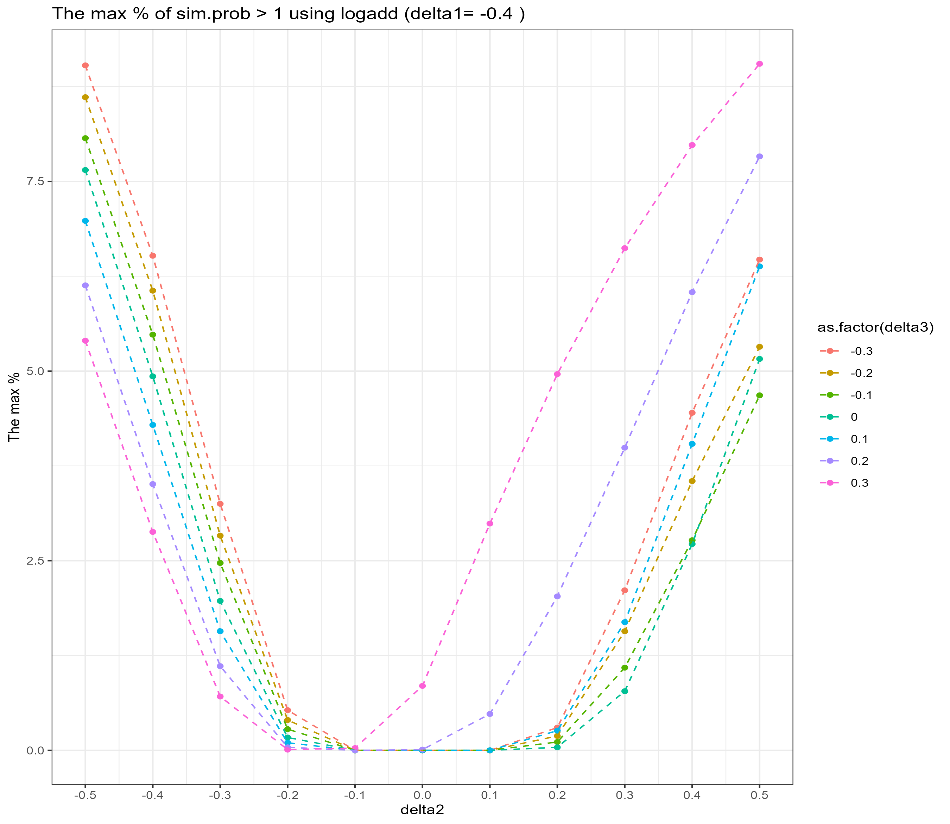
