## Supplementary material for "Selection bias due to omitting interactions from inverse probability weighting": Supplementary 2 for real-data application.docx

### 1 Descriptive statistics for variables used

|  | Mean (SD)/ Frequency (%) | |
| --- | --- | --- |
| Variables (number observed with %) | Full-data sample (n=32,117) | Complete cases (n=9,654) |
| Sleep duration **(n=10,682; 33%)** | 404 (78) | 405 (76) |
| Household participation weight | **0.797 (0.451)** | **0.879 (0.421)** |
| Individual participation weight | 1.265 (0.342) | 1.234 (0.268) |
| Unemployment status **(n=12,399; 39%)** |  |  |
| Employed | 11,260 (90.8%) | 8,862 (91.8%) |
| Umemployed | 1,139 (9.2%) | 792 (8.2%) |
| Age group | n=32,117 |  |
| 24-29 | 4,814 (15.0%) | 900 (9.3%) |
| 30-39 | 8,507 (26.5%) | 2,166 (22.4%) |
| 40-49 | 9,641 (30.0%) | 3,158 (32.7%) |
| 50-59 | 9,155 (28.5%) | 3,430 (35.5%) |
| Marital status **(n=14,623; 46%)** |  |  |
| Other | 4,415 (30.2%) | 2,694 (27.9%) |
| Married/civil partnership/living as couple | 10,208 (69.8%) | 6,960 (72.1%) |
| Educational level **(n=20,58; 64%)** |  |  |
| Degree/diploma | **13,144 (63.9%)** | **4,917 (50.9%)** |
| A level/equivalent | 1,571 (7.6%) | 1,042 (10.8%) |
| GCSE/equivalent | **4,249 (20.6%)** | **2,755 (28.5%)** |
| None | 1,617 (7.9%) | 940 (9.7%) |
| Whether anyone in household born in UK (n=32,116; 100%) |  |  |
| At least one UK born | 26,141 (81.4%) | 8,573 (88.8%) |
| All not UK born | 5,975 (18.6%) | 1,081 (11.2%) |
| Ethnic group (n=32,116; 100%) |  |  |
| White British | **20,467 (63.7%)** | **7,037 (72.9%)** |
| Other White | **1,916 (6.0%)** | **469 (4.9%)** |
| Asian | 5,499 (17.1%) | 1,310 (13.6%) |
| Black | 2,617 (8.1%) | 479 (5.0%) |
| Mixed/other | 1,617 (5.0%) | 359 (3.7%) |
| Sex (n=32,116; 100%) |  |  |
| Female | 18,156 (56.5%) | 5,735 (59.4%) |
| Male | 13,960 (43.5%) | 3,919 (40.6%) |

The table gives descriptive summary statistics for the full US sample and the subsample with complete data that was used for our analysis. Complete cases seem to be older, less educated and more likely to be native British.

### 2 Justification of the definition of individual response

Among recruited households, individuals can choose to be interviewed (“respond”) or not at the first wave in which they are an adult even if an individual does not respond at this wave, he/she might respond subsequently. Therefore, these individuals could have – in theory – been included in the model for estimating the complete-case weight. However, the majority (6169/9649=64%) of individuals who did not respond at their first wave of inclusion, did not respond at any further waves; these individuals could not be included in the model for estimating the complete case weight because they would not have any individual level data apart from basic demographic information. Thus, we decided to estimate the probability of being a complete case conditional on having responded as an individual at the first eligible wave and given that their household was recruited. As such individuals who did not respond at their initial wave are dropped.

### 3 Covariates for deriving individual participation weight.

The covariates used are recorded at recruitment (rather than wave 9): three individual characteristics are included in the model for individual initial response: age (grouped as 16-19, 20-24, 25-29, 30-39, 40-49, 50-60), sex, ethnicity (grouped as above); seven household characteristics were: net income in quintiles, whether or not there was at least one person in the household in paid employment, the highest household qualification (grouped as above), the number of bedrooms (1, 2, 3, 4 or more), housing tenure (owned/mortgaged, private rented, housing association/council rented/other), household size (1, 2, 3, 4 or more), and whether anyone in the household was born in the UK; three additional characteristics included government region (North East, North West, Yorkshire and the Humber, East Midlands, West Midlands, East of England, London, South East, South West, Wales, Scotland, Northern Ireland), sample origin (GPS GB, GPS NI, EMB, IEMB), and the wave at which an individual was first eligible for the adult interview. This model includes thirteen variables with all possible two-way interactions among these variables, some coefficients of which might be set to zero using lasso. If only one person is in a household, the probability of responding is 100%.
